# Multidisciplinary analysis of evolution based Abiraterone treatment for metastatic castrate resistant prostate cancer

**DOI:** 10.1101/2021.11.30.21267059

**Authors:** Jingsong Zhang, Jessica J. Cunningham, Joel S. Brown, Robert A. Gatenby

## Abstract

**Background:** We present a multidisciplinary approach to clinical trial design and analysis in a pilot study (NCT02415621) in which evolution-based mathematical models guide patient-specific dosing for Abiraterone treatment in men with castrate resistant metastatic prostate cancer.

**Methods:** Abiraterone plus prednisone were administered intermittently based on an evolutionary mathematical model. Outcomes are compared to historical controls and a matched contemporaneous cohort who met trial eligibility but received SOC dosing. Longitudinal cohort data allowed modification of pre-trial model parameter estimates. Model simulations of each patient using updated parameters critically evaluated trial design.

**Results:** Trial patients, on average, received no abiraterone during 59% of time on treatment. Median Time to Radiographic Progression (TTP) was 30.4 months compared to 14.3 months in the contemporaneous SOC group (p<0.001). All patients in the SOC group have progressed but 4 in the adaptive cohort remain on treatment at >1800 days. Longitudinal trial data found the competition coefficient ratio (*α*RS/*α*SR) of sensitive and resistant populations, a critical factor in intratumoral evolution, was 2 to 3-fold higher than pre-trial estimates. Computer simulations using the corrected parameter unexpectedly demonstrated optimal cycling can reduce the resistant cells. Longitudinal data from 4 trial patients who remain on treatment are consistent with model predictions. Modeling results predict protocol changes that will allow similar outcomes in most patients.

**Conclusions:** Administration of abiraterone using evolution-based mathematical models decreased drug dosing and increased radiographic TTP. Integration of mathematical models into trial design identifies novel insights into key treatment parameters and provides optimization strategies for follow-up investigations.

**Article Summary:** supplemental sections outline the methodology for parameter estimates from trial data, computer simulations, and comparison of simulation results and actual clinical data in every patient in both cohorts.

**Statement of Translational Relevance:** Integration of evolution-based mathematical models significantly increased TTP in abiraterone therapy for mCRPC. This multidisciplinary approach represents a novel clinical trial strategy in which the treatment protocol is framed mathematically, clinical data then refines model parameterization, and simulations using the updated model predict alternative strategies to improve outcomes. Here we demonstrate the mathematical models used to design the trial can also produce novel analytic approaches. By using longitudinal trial data, key model parameters can be refined. Simulations using the updated model can then be applied to every patient in the trial. Finally, additional simulations demonstrate alternative protocols that could improve results. These analyses demonstrate evolution-based approaches may allow consistent long-term control in patients with metastatic prostate cancer.

## Introduction

While often initially effective, nearly all cancer treatments ultimately fail due to evolution of resistance (1). Prior efforts to disrupt the molecular machinery of resistance (such as P-Glycoprotein during chemotherapy administration) has led to small or no improvement in outcomes (2, 3) indicating tumor cells generally have multiple available mechanisms of resistance. However, resistant tumors, regardless of the precise mechanism, require both deployment of molecular mechanisms of resistance and proliferation of surviving (i.e., resistant) populations to become clinically significant (4, 5). We hypothesize the former is an inevitable response to treatment, but the latter is governed by eco-evolutionary principles and potentially manageable by Darwinian controls (6). Evolution-based models show current treatment strategies, which apply therapy at maximum tolerated dose until progression, are often not evolutionarily optimal. While an initial response may be large, therapy fails because it strongly selects for resistance while eradicating all treatment sensitive cells. The resistant cells are now free from competition with sensitive cells – an evolutionary dynamic termed “competitive release” -which allows for rapid proliferation (7, 8).

One evolutionary strategy to delay population growth of the resistant phenotype, termed “adaptive therapy” (9), exploits the fitness costs incurred by production, maintenance, and operation of the molecular machinery required for treatment resistance (10). These resource demands are compensated for by increased fitness when treatment is applied. However, in the absence of treatment, the phenotypic costs reduce fitness compared to competing sensitive cells (10), particularly in resource limited tumor microenvironments. Thus, when multiple mechanisms of resistance are available, the precise cost will likely vary but seldom will there be *no* associated cost. In nature, similar cost/benefit trade-offs are seen in, for example, loss of eyes by cavefish as the resource costs of producing and maintaining them are balanced against their lack of utility in a continuously dark environment (11). Population dynamics dependent on the cost of resistance are now fundamental principles in pest management (12) and have been observed experimentally in cancer populations (13).

The phenotypic cost of resistance can be explicitly measured in, for example, membrane extrusion pumps (14). However, there are no data regarding the molecular dynamics leading to abiraterone resistance in metastatic castrate resistant prostate cancer (mCRPC). Furthermore, the precise mechanism of resistance may vary (15). When such measurements cannot be obtained, census methods developed in ecology (16) can estimate relative fitness based on population distributions. That is, if abiraterone sensitive cells are the dominant tumor subpopulation prior to treatment, they must be fitter than the resistant phenotypes in the absence of treatment even if the specific reason for this fitness difference is not known.

Adaptive therapy (9, 17) limits application of treatment to produce a moderate decrease in tumor volume while explicitly retaining a significant population of treatment sensitive cancer cells. Following an initial response, treatment is withdrawn allowing the cancer populations to proliferate. But, in the absence of selection pressures from treatment, the sensitive cells have a fitness advantage and will proliferate at the expense of the resistant cells. Thus, when the tumor returns to its pre-treatment volume, the subpopulation distribution is similar allowing the initial therapy to remain effective.

Because adaptive therapy starts and stops treatment, it is conceptually similar to “intermittent therapy” trials in which men with castration sensitive metastatic prostate cancer (mCSPC) were randomized into continuous or intermittent androgen deprivation therapy (ADT) treatment using LNRH analogues. Neither regimen proved superior (18). Although trial design has similarities to the adaptive therapy protocol, we note it included 8 months of “induction therapy” with ADT and that intermittent dosing instituted only if the PSA reached < 4ng/ml. With computer simulations (19, 20), we have demonstrated these two requirements consistently reduce the sensitive population to near-extinction levels; producing the observed outcomes from the intermittent arm that are indistinguishable from continuous therapy. Similar limitations apply to a prior study that used intermittent ADT with fixed 8-month intervals (21). Model simulations showed that the intervals were too long and promoted the dominance of resistant cancer cells.

These analyses illustrate the potential value of trial design based on evolutionary first principles framed by mathematical models. Here we present such a trial. Computer simulations of the model were used to predict optimal trial design. Later, the same models could be evaluated and parameterized to patient specific data allowing for novel approaches to trial analysis. Thus, longitudinal trial data and observed outcomes allow for refinement of pre-treatment parameter estimates. Simulations using the updated model can then be applied to each patient in the trial to analyze intratumoral evolutionary dynamics during treatment. Unlike conventional clinical trials, this approach allows both cohort and patient-specific analyses. Furthermore, the simulations critique trial design and performance thus providing guidance for alternative strategies and future investigations.

Thus, we hypothesize that formally integrating evolutionary dynamics into abiraterone treatment will delay proliferation of resistant cells prolonging TTP. Our trial objectives were twofold: 1. Test the underlying hypothesis in a small patient cohort. 2. Investigate a novel trial design in which protocol design is based on a mathematical model and analyzed through an iterative process in which trial data informs model parameter estimates and computer simulations from the updated model investigate intratumoral evolutionary dynamics in each trial patient.

Here we provide follow-up on an initial report submitted when the benefits of adaptive therapy compared to SOC treatment achieved statistical significance (17). However, at that time, we could only demonstrate the median time to progression was >27 months. We confirm the superiority of Adaptive Therapy over SOC. Additionally, we demonstrate how our multidisciplinary approach to treatment design and analysis provides novel patient-specific information that may reduce the need for large, expensive clinical trials.

## Methods

### Mathematical Models

#### Evolutionary subpopulations

The treatment protocol is based on a population model (17) that assumes three competing phenotypes: (i) TP cells have androgen receptors and upregulated CYP17A1 to produce testosterone – these are the primary target of abiraterone treatment; (ii) T+ cells requiring exogenous androgen – these are the primary target of androgen deprivation therapy (ADT) but will proliferate in the presence of TP cells as the produced androgen diffuses through the tumor microenvironment; and (iii) T-cells that are androgen-independent and resistant to ADT and abiraterone.

We used Lotka-Volterra competition equations to model the interactions between the three cell types based on parameters for intrinsic growth rates, *r*_*i*_, carrying capacities, *K*_*i*_, and the matrix of competition coefficients, *a*_*ij*_.

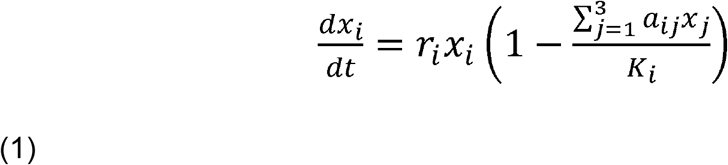

Since virtually all prostate cancers respond to ADT, we assumed T+ cells are the fittest population in the absence of any therapy. Fitness of TP and T-cells are diminished by the cost of resistance but cannot be directly measured. Since patients are included in the study only if their PSA declined by at least 50% after initial application of abiraterone, the TP cells are more prevalent than the T-cells. Applying the census methodology (16), we can estimate the TP cells are fitter than the T-cells prior to treatment. We assume that each phenotype produces roughly equal amounts of PSA, which may introduce error if this assumption is substantially violated.

Each competition coefficient (*a*_*ij*_) standardizes the competitive effect of an individual of type *j* on the per capita growth rate of type *i* in units of type *i*. In general, the value of the competition coefficient reflects the relative fitness of the populations. All *a*_*ii*_ = 1. If *a*_*ij*_ > 1, then inter-type competition is greater than intra-type; and vice-versa if *a*_*ij*_ < 1.

We model therapy by having abiraterone reduce the carrying capacities of the TP and T+ cells, with no effect on T-. We assume that TP cells are either killed or quiescent during abiraterone treatment. Since abiraterone inhibits the production of testosterone by the TP cells, the T+ “cheater” population will have no source of testosterone and they too decline with abiraterone.

### Pilot clinical trial

This is a single institution investigator-initiated pilot study (NCT02415621) at the Moffitt Cancer Center, Tampa, Florida. The protocol was approved by central IRB and monitored by Moffitt Cancer Center’s protocol monitoring committee. Details of the trial design have been previously published (17). Briefly, inclusion criteria were similar to phase III AA-302 trial (19) population except allowing ECOG 2 performance status (PFD), prior exposure to enzalutamide, Sipuleucel-T, and ketoconazole. Prior docetaxel was allowed if it was given during the castration sensitive phase. Patients on opioids for cancer-related pain were excluded. Patients could be enrolled in the study after achieving 50% or more decline of their pre-abiraterone PSA levels. Cohort size was designed to have sufficient statistical power to detect a 50% increase in TTP.

Each enrolled patient began on abiraterone (1000 mg by mouth daily) and prednisone (5 mg by mouth twice daily) until achieving a > 50% decline in their baseline levels of PSA pre-abiraterone. Upon achieving this decline, abiraterone therapy was suspended. Tumor regression or stability was confirmed by radiographic measurements.

Patients were monitored every 4-6 weeks with a lab (CBC, COMP, LDH and PSA) and clinic visit. Serum testosterone was not measured. Every 12 weeks, each patient received a bone scan, and a computed tomography (CT) of the abdomen and pelvis. Abiraterone plus prednisone were reinitiated when a patient’s PSA increased to or above the pre-abiraterone PSA baseline. Abiraterone therapy was stopped again after the patient’s PSA declined to > 50% of his baseline PSA. Each successive peak of PSA when abiraterone therapy was reinstated defined a complete cycle of adaptive therapy.

For patients who did not undergo surgical castration, GnRH analog treatment was continued to maintain castration levels of serum testosterone. Patients who did not achieve a 50% decline of their baseline PSA after restarting abiraterone remained on study until they developed radiographic progression while on abiraterone based on Prostate Cancer Working Group (PCWG) 2 criteria. Patients who developed radiographic progression while off abiraterone would restart abiraterone and remain on abiraterone until a partial response was noted in the measurable lesions and stable disease was noted in the non-measurable lesions in the repeat bone scan, and abdominal and pelvic CT. These subjects were then allowed to stop abiraterone and reenter the adaptive therapy cycles. Patients were followed until they developed radiographic progression or ECOG performance status deterioration while on abiraterone, whichever came first.

#### Standard of Care Cohort

Sixteen patients who were treated with continuous abiraterone as the standard of care and met the eligibility criteria for our adaptive therapy were identified through chart review of mCRPC patients treated at Moffitt Cancer Center during the time of the study enrollment. The results of the trial cohort were compared to these patients treated with standard of care abiraterone dosing during the time of the trial. All patients fulfilled trial eligibility requirements (including a > 50% drop in PSA) and chose SOC treatment.

#### Statistics

To compare patient characteristics between the trial and SOC cohorts, we used Kruskall-Wallis non-parametric one-way ANOVAs (done with SYSTAT13). To compare progression free survivorship between the two cohorts we used the Mantel logrank test (done with SYSTAT13).

#### Data Availability

The data generated in this study are available within the article and its supplementary data files. The code used for analysing, parameter fitting and simulations of the Lotka-Volterra model is available upon request from the corresponding author.

## Results

### Cohort Analysis

Twenty patients were accrued to the trial. One was lost to follow-up and three withdrew (one citing the psychological stress of changing treatments). In the 16 evaluable patients, the locations of metastases and initial Gleason Scores and pre-treatment PSA values are well matched between both trial and standard of care (SOC) cohorts (**Supplemental Table 1)**: Gleason scores: Kruskal-Wallis test statistic of 0.088 with p = 0.767 based on a Chi Square distribution, df = 1. Pre-treatment PSA levels: Kruskal-Wallis test statistic of 0.157 with p = 0.692 based on a Chi Square distribution, df = 1.

This study was conducted before abiraterone was approved in the castration sensitive setting. None of the patients enrolled in the adaptive therapy trial or included in the historical control had received new hormonal agent (abiraterone, enzalutamide or apalutamide) or docetaxel in the castration sensitive setting. Abiraterone was the frontline therapy for mCRPC for most patients in each group expect a few patients received sipuleucel-T prior to abirateone.

Given patients enrolled in the study had more frequent PSA checks than the historical control arm, more patients in the study cohort had > 50% PSA reduction within a month. The percentage of patient who had more than 50% PSA reduction within 2 months were similar 15/17 (88%) vs 12/16 (75%).

In a preliminary report, we found the median radiographic TTP could not be less than 27 months (22). Here, consistent with this and model predictions, evolution-based application of abiraterone significantly improved (Mantel logrank test, p<0.001) median radiographic TTP (**Figure 1**) in the adaptive therapy group (30.4 months) compared to the contemporaneous SOC group (14.3 months). All patients in the SOC group have progressed but 4 patients in the adaptive cohort remain on study with no evidence of imaging or symptomatic progression after >1800 days on trial and a median of 13 cycles per patient. Patients in the adaptive therapy group received an average abiraterone dosing rate (mg drug/patient/unit time) of 41% compared to SOC. That is, on average, the trial patients were not receiving abiraterone during 59% of their time on trial.

**Figure 1.**
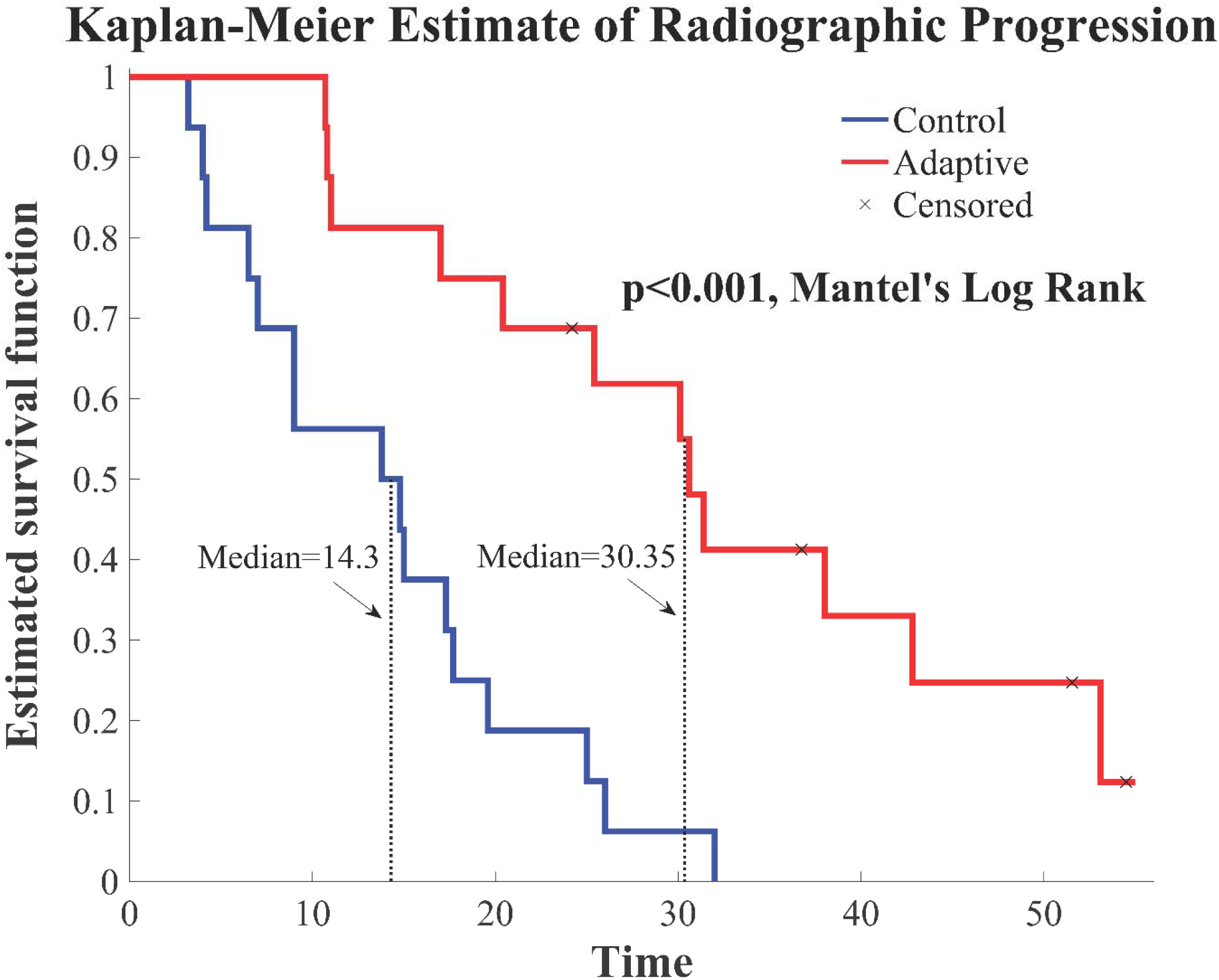
Survival curves. Kaplan-Meier curve for Time to Radiographic Progression in the adaptive cohort (n=16) compared to the SOC (n=16) cohort. Four (4) patients in the adaptive arm remain in the trial with no evidence of progression.

### Mathematical Analysis

Because ADT was continued during abiraterone therapy, we assumed that the T+ cell proliferation was linked exclusively to androgen production by the TP cells. That is, loss of TP cells would reduce intratumoral androgen concentrations necessitating a decline in the T+ population. Their linked fates allow us to reduce Equation 1 to a 2 species model with T-cells as the resistant population (*x*_*R*_) and the TP and T+ cells as a combined sensitive population (*x*_*S*_). Population sizes correspond to overall tumor burden. We arbitrarily set the carrying capacity to 10000 so that Equation 1 becomes:

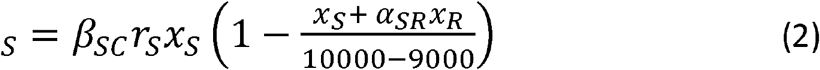

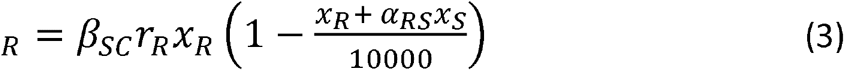

Where *r*_*S*_ and *r_r_* are the population growth rates (units of per day), *β*_*SC*_ is a scaling factor, and *⍰* (value of 1 during abiraterone treatment and 0 during drug holidays) is the effect of abiraterone on the carrying capacity of the sensitive cells.

The rates of PSA increase in the SOC cohort prior to abiraterone treatment (i.e., during progression while treated with ADT) and PSA increase during off treatment periods in adaptive therapy cycles were used to estimate the range of growth rates for abiraterone sensitive population. The rates of PSA increase in the SOC cohort during progression while treated with abiraterone were used to estimate the range of growth rates for abiraterone resistant population. Sensitive cells were estimated to have significantly higher mean growth rate (0.0156 per day [population increase of 1.56% per day]) than that for resistant cells (0.0091 per day) (p<0.05) (**Supplemental Section 1**) in the absence of treatment. Both values are within the range observed in clinical cancers and the difference is consistent with our theoretical premise that resistance incurs a cost which decreases fitness and proliferation when therapy is not applied.

Adaptive therapy relies on evolutionary dynamics in which the sensitive population, due to its fitness advantage in the absence of therapy, reduces proliferation of the resistant population. That is, for the adaptive strategy to be successful, the negative effects of the sensitive population on the resistant population (*α*_*RS*_) must be greater than effects of the resistant population on the sensitive one (*α*_*SR*_). To determine the value of *α*_*RS*_, we used constrained nonlinear multivariable optimization to estimate parameters by minimizing the least-squares difference between the output of the model and the actual patient data over the entire cohort (see **Supplemental Section 2**). As shown in **Figure 2**, the global minimum is found when *α*_*RS*_ ≈ 6 (with *α*_*SR*_ = 1). For perspective, the ratio of the competition coefficient of a dominant species over a non-dominant species in nature ranges from slightly above 1 to over 100 (23). Model simulations prior to the protocol estimated *α*_*RS*_/*α*_*SR*_ ratio to be less than 2. This change has considerable clinical significance. As shown in **Figure 3a**, if *α*_*RS*_/*α*_*SR*_ <1, the sensitive population has no fitness advantage and adaptive therapy will provide little benefit. If *α*_*RS*_/*α*_*SR*_ = 2, the increasing size the sensitive population (*x*_*S*_) following treatment cessation causes the resistant cells to (*x*_*R*_) stop proliferating so that the population remains constant (**Figure 3a)**. Thus, at each cycle, the simulations predict the resistant population will increase slightly (that is the resistant population grows during treatment when the sensitive population declines so that it no longer exerts a negative influence) with each cycle. This will inevitably result in a threshold effect in which the resistant population takes control. However, for *α*_*RS*_/*α*_*SR*_ = 6 the simulations (**Figure 3b**) show that the increasing sensitive population (*x*_*S*_) after treatment cessation causes a substantial *decrease* in the size of the resistant population (*x*_*R*_). In fact, *simulations using the α*_*RS*_*/α*_*SR*_ *estimated from patient-specific longitudinal data show how 3 to 4 consecutive optimal treatment cycles will cause the resistant population to approach 0*.

**Figure 2.**
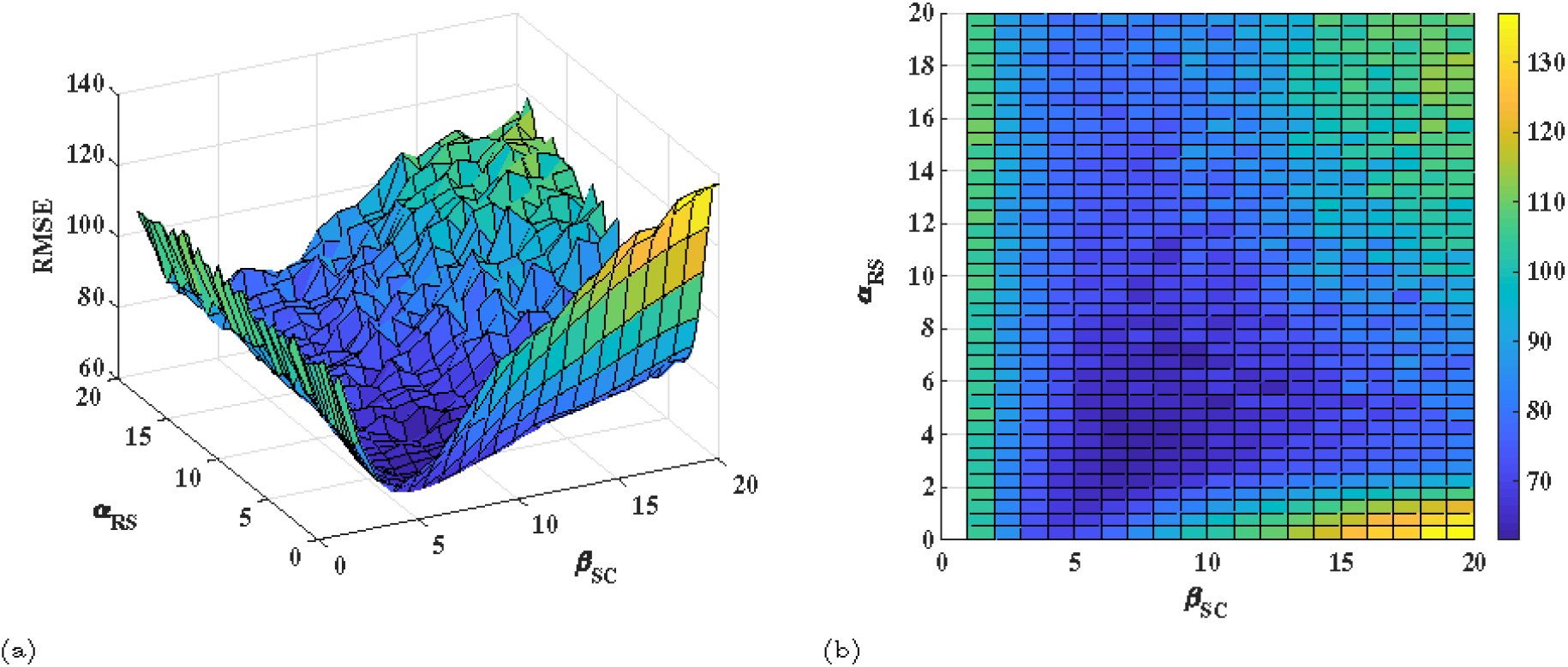
Key parameter estimates. To estimate *α*_*RS*_ and *β*_*SC*_, a constrained nonlinear multivariable optimization is performed to minimize the least-squares difference between the output of the model and the actual patient data over the entire cohort. The global minimum is found at *α*_*RS*_ = 6 and *β*_*SC*_ = 8.

**Figure 3.**
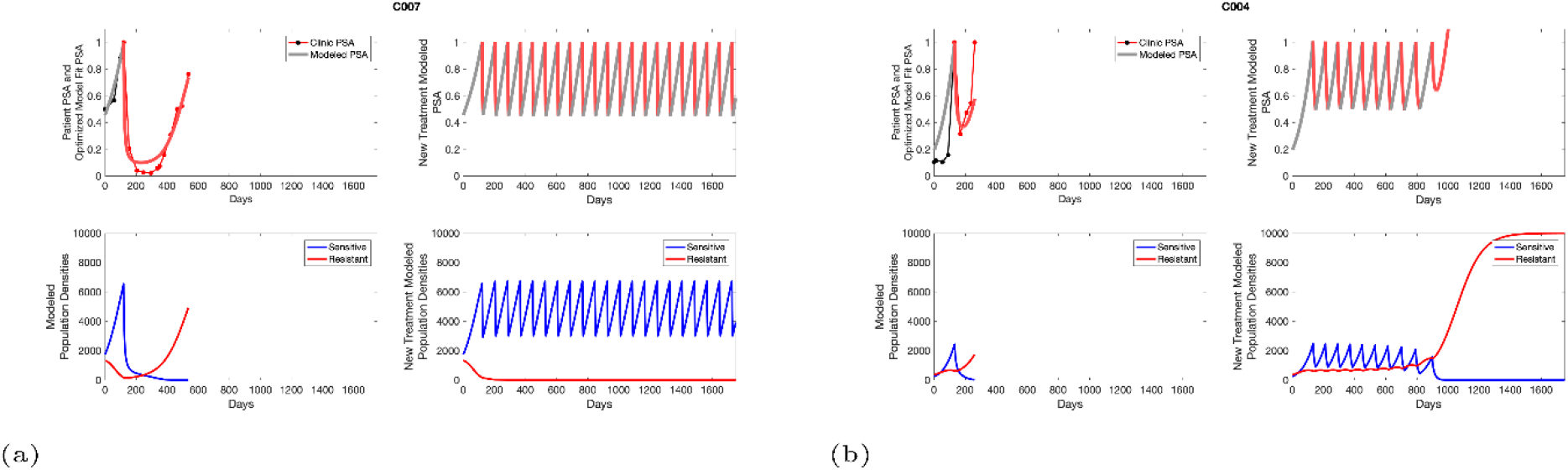
Patients with similar initial resistant fractions treated with SOC and adaptive protocols. <u>Left panel</u>, SOC patient C007 with pre-treatment resistant fraction of 0.03 with radiographic TTP of 526 days. <u>Right Panel</u>, SOC patient C004 with pretreatment resistant fraction of 0.25 and radiographic TTP of 128 days. In both patients, the adaptive therapy protocol would have permitted tumor control of >1000 days. Note the simulations predict that, in patient C007, the idealized treatment protocol would have caused the resistant population to monotonically decrease to 0. This dynamic reflects the unexpectedly large value of *α*_*RS*_ (**Figure 2**) as discussed in the text. Even with this large value of *α*_*RS*_, very large pre-treatment resistant fractions like that in patient C004 cannot be eliminated completely.

The relative fraction of the resistant population at the time of initial abiraterone therapy is also estimated using the population dynamics under the calculated growth rates and optimized values for *α*_*RS*_ and *β*_*SC*_ (**Supplemental Section 3**).

Comparing patient PSA to the best model fit in the 32 patients (**Supplemental Section 4**) allows several observations. First, there are just 6 parameters; two estimated from the rates of exponential increase during phases of therapy cycles and then set to the average values for all patients, two estimated from patient data and then fixed at the optimized values, and two allowed to vary to provide the best patient-specific fit. This prevents over-fitting the data and hence losing confidence in the statistical power of the fit.

Second, the fits are generally tight, but there are exceptions in the relatively poor fit to patients 1004, 1007, and C002. This suggests fixed model parameters (growth rates, scaling factor on growth rates, and competition coefficients) may vary in some patients. Alternatively, the serum PSA concentration may scale to population size differently in these patients. Relaxing any of these assumptions and refitting these three patients with patient-specific parameter changes does improve the model fit, but at the cost of having to do the same for all patients and risking over-fitting.

### Evolutionary dynamics in the adaptive and SOC subjects

As demonstrated in **Supplemental Figure 5**, mathematical estimates of pre-treatment fractions of sensitive and resistant cells correlated with subsequent radiographic TTP in both cohorts. We find radiographic TTP was greater in the adaptive cohort for every level of the pre-treatment resistant population. Consistent with this, model simulations using the derived parameter estimates, confirm that all patients in the SOC cohort would have had improved outcomes using the current adaptive therapy protocol (**Figure 4, Supplemental Section 5)**. Furthermore, no members of the adaptive cohort would have benefited from SOC dosing (**Supplemental Figure 5)**.

**Figure 4.**
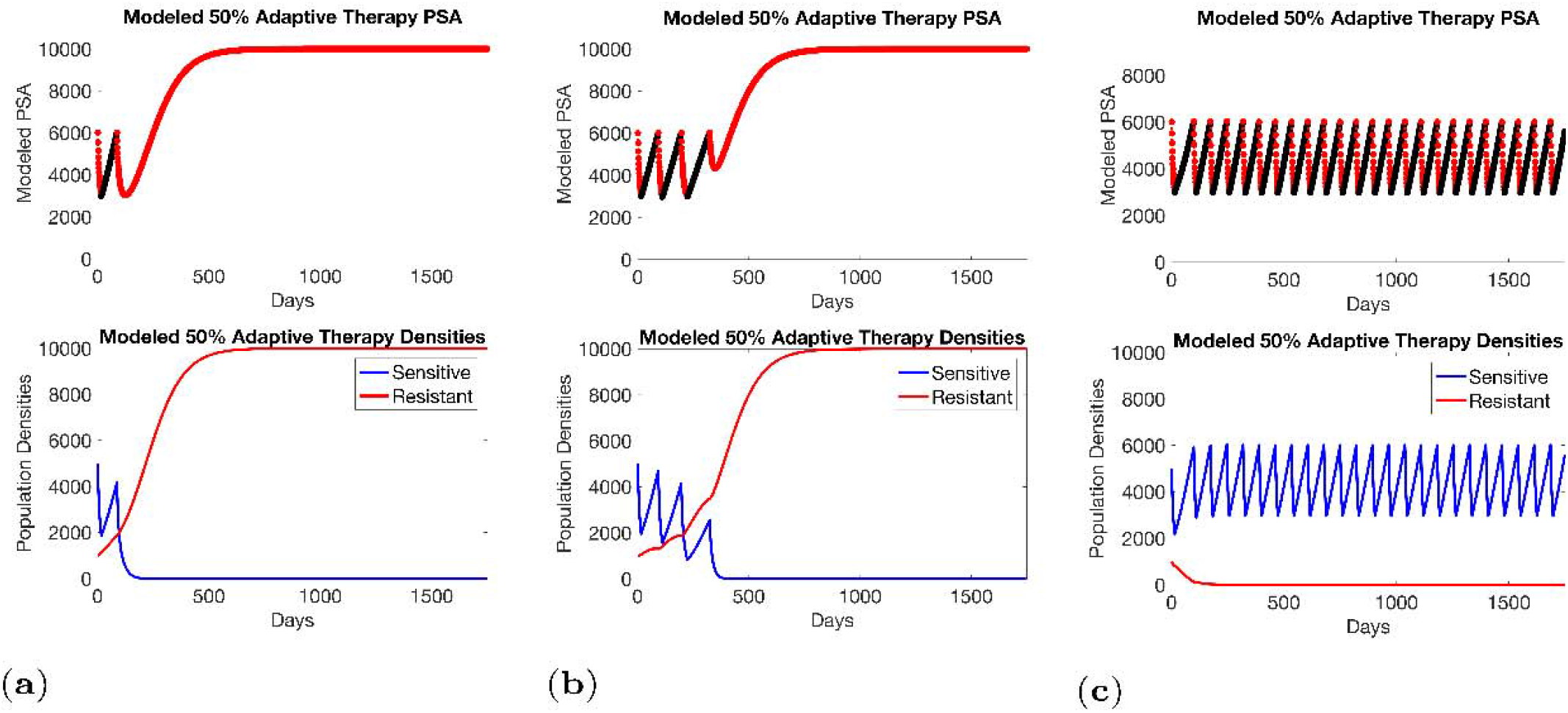
Sensitivity of resistant cell population to the value of the competition coefficient. Treatment dynamics are sensitive to the value of the competition coefficient (*α*_*RS*_), which is dependent on the fitness differences of the sensitive and resistant populations in the absence of treatment. In panel (a) we assume *α*_*RS*_ =0.8 and increase in *x*_*S*_ does not decrease the population *x*_*R*_ and adaptive therapy fails. In panel (b), *α*_*RS*_ = 2, the increase of *x*_*S*_ during treatment holidays slows the growth of x_R_ and delays treatment failure. In panel (c) the estimated *α*_*RS*_ = 6 results in a *negative* growth rate in *x*_*R*_ during proliferation of *x*_*S*_. Over 3 to 4 cycles, the *x*_*R*_ population approaches 0. This allows the cycling treatment to maintain tumor control indefinitely. Note, however, this represents an ideal setting and does not take into account other dynamics (see below) that may result in loss of control.

Once the model parameters were updated with longitudinal trial data, we conducted new simulations of every patient in both cohorts. In Figure 5, we demonstrate the key model prediction – 3 or 4 consecutive optimal cycles can reduce the population to near 0 – *is observed* in the 4 trial patients who remain in a stable cycling regime at >1800 days on treatment (**Figure 5**). In these patients, the PSA value of each cycle trough remains constant rather than steadily increasing due to step wise increases in the resistant population as predicted by the pre-trial simulations. *This discrepancy allows for a cautious conclusion that the resistant population has been greatly reduced and possibly eradicated*.

**Figure 5.**
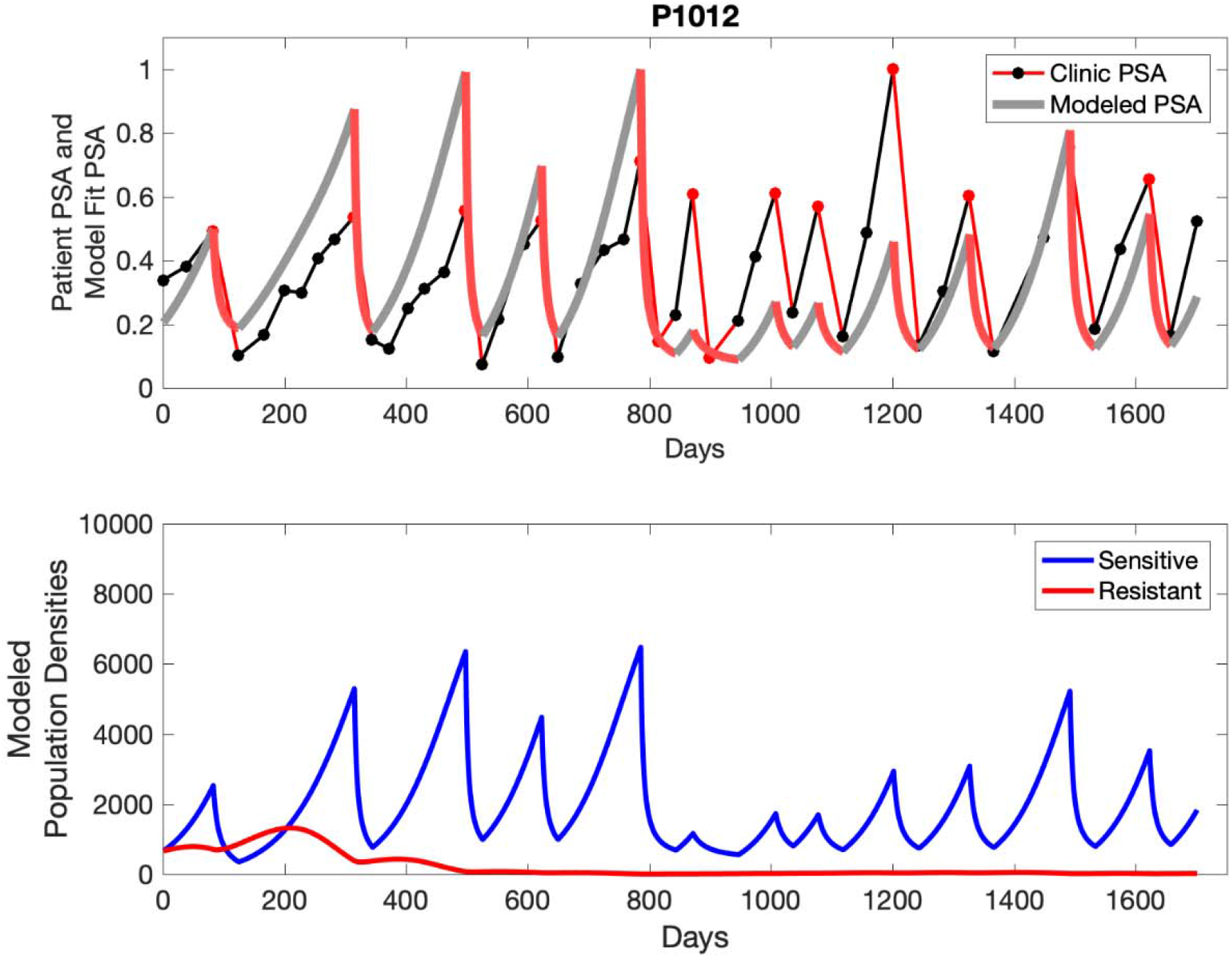
Simulations suggest optimal timing resulted in elimination of the resistant population in adaptive therapy patients with prolonged survival. Patient 1012 in the adaptive therapy cohort with enduring control (>1800 days). Model simulations suggest that the sequence of 2 to 4 treatment cycles caused the resistant population to reduce to near extinction permitting a stable cycling regime in which only abiraterone-sensitive cells are present.

Why was tumor progression observed in most members of the adaptive therapy cohort? Computer simulations suggest that they were over-treated. While protocol design required monthly PSA levels, radiographic studies were limited to 3-month intervals. As demonstrated in **Figure 6**, the protocol requirement for radiographic confirmation of response resulted in multiple weeks and even months in which patients remained on treatment even when the PSA was <10% of pretreatment values. Simulations suggest that this unintended, yet excessive, reduction of the sensitive population led to the proliferation and dominance of the resistant population.

**Figure 6.**
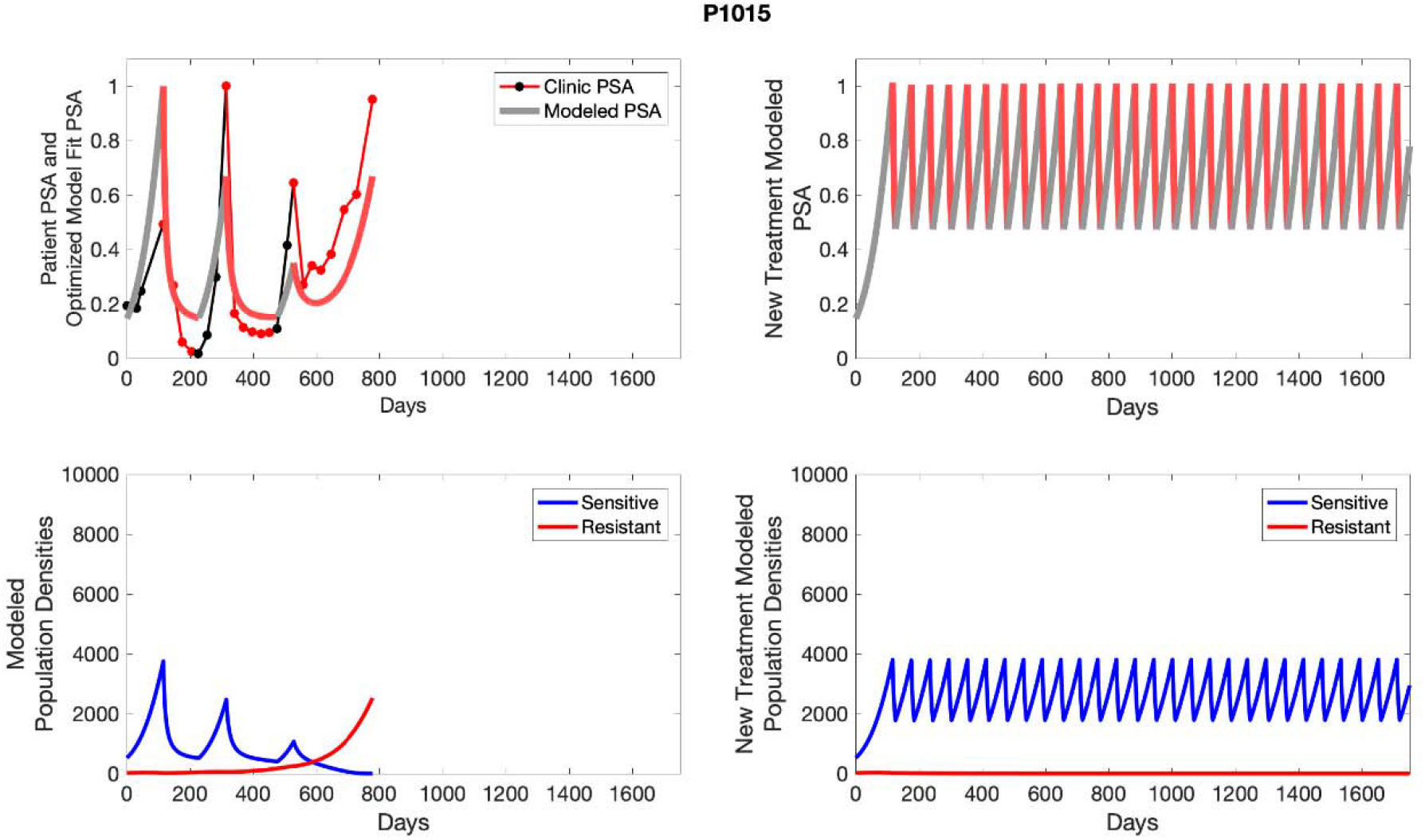
Patients who progressed in the adaptive panel are predicted to have benefited from reduced abiraterone administration. <u>Left panel</u>,Simulations suggest that unintended, yet excessive, reduction of the sensitive population led to the proliferation and dominance of the resistant population. <u>Right Panel</u>, Optimizing the timing of withdrawing therapy immediately upon reaching the 50% pretreatment PSA threshold, thereby preventing over treatment, allows maximal suppression of the resistant population and consistent long-term control in adaptive therapy patients.

## Discussion and Conclusion

Our results suggest cycling of sensitive cells, depending on key intra-tumoral evolutionary parameters, can maintain control of resistant cells often for prolonged time periods. More broadly, we present a conceptual model for trial design in which the treatment protocol is linked to predictions from a mathematical model. Here, analyses of trial results, in addition to traditional cohort outcomes, includes mathematical curve fitting of longitudinal data from individual patients to estimate key model parameters. Subsequently, computer simulations using the updated model can be applied to each patient to estimate the tumor evolutionary dynamics that led to the observed outcomes. Finally, computer simulations can examine alternative treatment strategies that would have produced better outcomes.

While this approach seemed successful in this pilot trial, it clearly must be validated in multiple other trials and include larger study cohorts. Furthermore, variable mechanisms of resistance can give rise to dynamics other than those observed in this relatively small cohort. In this study, for example, the patients received 3 different treatments (ADT, steroids, and abiraterone) but only abiraterone dosing was modulated. Thus, for example, intermittent dosing of steroids instead of or in addition to abiraterone is a potentially successful alternative strategy (24). We note these more complex strategies can and should be explored mathematically to identify optimal trial design. Lastly, adaptive dosing in prostate cancer treatment is enabled by a serum biomarker (PSA) that is a generally accurate metric of changing tumor burden within a patient (but not between patients). Other cancers that lack a serum biomarker will require clinical decisions to be made based on estimates of tumor volumes from imaging. This strategy has been used in animal experiments (13) but does add concerns regarding accuracy and cost in a clinical setting.

Nevertheless, acknowledging the above caveats, we demonstrate integration of evolution-based mathematical models into trial design significantly increased TTP in abiraterone therapy for mCRPC. Patients did not receive abiraterone, on average, during 59% of the trial period thus reducing potential toxicity and expense. While we did not use quality of life metrics to estimate these benefits, an economic analysis of the trial found an average cost reduction of $50,000 per patient per year (25). The cohort size in this pilot study is relatively small, but we note the increase in TTP was highly statistically significant (p<0.001) compared to a contemporaneous cohort and historical data (26-28). Furthermore, as noted above, the use of mathematical models in trial design and analysis expands information that can be obtained from even small cohorts.

The “After Action Analysis (29)” computer simulations, using the updated parameter estimates from the longitudinal trial data, predicted every SOC patient would have benefited from the adaptive application of abiraterone, and no members of the adaptive cohort would have benefited from SOC dosing. Computer simulations also identified important flaws in the trial protocol. Because PSA sampling occurred at monthly intervals and imaging at 3-month intervals, the decision to end or restart therapy often occurred weeks or months after the PSA value had crossed the necessary threshold. Somewhat counterintuitively, simulations demonstrated that delay in restarting abiraterone as the PSA increased had little clinical effect but delays in withdrawing treatment often resulted in excessive reduction of the sensitive tumor population which significant reduced TTP. That is, by waiting too long for therapy withdrawal, the sensitive population was reduced below levels that could effectively suppress proliferation of the resistant population. In fact, computer simulations demonstrate optimal timing of abiraterone withdrawal over 3 to 4 cycles could reduce the resistant population to near extinction thus permitted long-term control in all trial patients. This unexpected prediction is supported by longitudinal data in 4 members of the adaptive therapy cohort who remain on stably cycling on trial after >5 years. We note this also demonstrates a potential therapeutic strategy to attempt tumor eradication. Future plans for adaptive therapy trials in prostate cancer include more rapid withdrawal of therapy when PSA crosses the 50% threshold and more extensive monitoring of intratumoral evolution using serum biomarkers including testosterone as well as circulating DNA for AR amplification, AR mutations, and CYP17a expression. Finally, we note that the models fundamentally address prostate cancer interactions with testosterone and, thus, any therapy related to androgen receptors and androgen production can be modeled using this approach.

## Data Availability

All data produced in the present study are available upon reasonable request to the authors

## Acknowledgements

We thank Dr. Robert Gillies for his helpful comment and critiques, the patients who participated in the trial, and the trial coordinators and clinic staff who facilitated the complicated protocol.

## Notes

**<u>Funding</u>** This work was supported by the James S. McDonnell Foundation grant, “Cancer therapy: Perturbing a complex adaptive system,” grants from the Jacobson Foundation, NIH/National Cancer Institute (NCI) R01CA170595, Application of Evolutionary Principles to Maintain Cancer Control (PQ21), and NIH/NCI U54CA143970-05 (Physical Science Oncology Network (PSON)) “Cancer as a complex adaptive system”. This work has also been supported in part by the Clinical Trials Core Facility at the H. Lee Moffitt Cancer Center and Research Institute, an NCI-designated Comprehensive Cancer Center (P30-CA076292), and the European Union’s Horizon 2020 research and innovation program under the Marie Sklodowska-Curie grant agreement No 690817.

**<u>Conflict of Interests</u>** The authors declare no conflict of interests.

### Competing Interest Statement

The authors have declared no competing interest.

### Clinical Trial

NCT02415621

### Funding Statement

This work was supported by the James S. McDonnell Foundation grant Cancer therapy: Perturbing a complex adaptive system grants from the Jacobson Foundation NIH/National Cancer Institute (NCI) R01CA170595, Application of Evolutionary Principles to Maintain Cancer Control (PQ21), and NIH/NCI U54CA143970-05 (Physical Science Oncology Network (PSON)) Cancer as a complex adaptive system. This work has also been supported in part by the Clinical Trials Core Facility at the H. Lee Moffitt Cancer Center and Research Institute an NCI-designated Comprehensive Cancer Center (P30-CA076292) and the European Union Horizon 2020 research and innovation program under the Marie Sklodowska-Curie grant agreement No 690817.

### Author Declarations

IRB of Moffitt Cancer Center gave ethical approval for this work

