## Supplementary figures and text for "Multidisciplinary analysis of evolution based Abiraterone treatment for metastatic castrate resistant prostate cancer"

S0  Demographic Data From Trial Cohorts

| | **Control** | **Adaptive** |
|---|---|---|
| Mean Gleason | 7.85 | 7.94 |
| Range Gleason | [7, 10] | [6, 10] |
| Mean Pre Abi PSA | 36.52 | 29.7 |
| Range Pre Abi PSA | [2.71, 93.4] | [1.46, 109.4] |
| **AJCC 7th Ed. M stage** | | |
| M1a | 1 | 1 |
| M1b | 14 | 14 |
| M1c | 1 | 1 |

**Supplemental Table 1.** Gleason scores, pretreatment PSA values, and metastatic sites in the adaptive and SOC cohorts

#### S1 Estimating Growth Rates From Patient Data

We estimated the growth rates (units of per day) of the sensitive and resistant populations of cancer cells using the PSA measurements from the patients enrolled in the trial and the contemporaneous cohort. We used exponential fits during various treatment times to determine the growth rate of the populations. Note that P1002, while not used in the analysis of the trial due to noncompliance, is used for mathematical analysis as data for multiple adaptive cycles prior to noncompliance is available. This brings the total number of adaptive therapy patients used in parameter estimation to 17. Furthermore, historical PSA measurements are unavailable for one patient used in analysis of the trial, bringing the total number of contemporaneous patients used in parameter estimation to 15.

To estimate the resistant cell population growth rates (Table S1), we used time series of increasing PSA levels after progression from standard of care abiraterone. We did this for patients in the contemporaneous cohort that were receiving continuous abiraterone. At the point of disease progression, we assume that the populations of cancer cells are comprised of just abiraterone resistant cells. The patients of this cohort provided 11 good examples for estimating the resistant cell population growth rate). Figure S1 provides examples from two patients of these data and analyses.

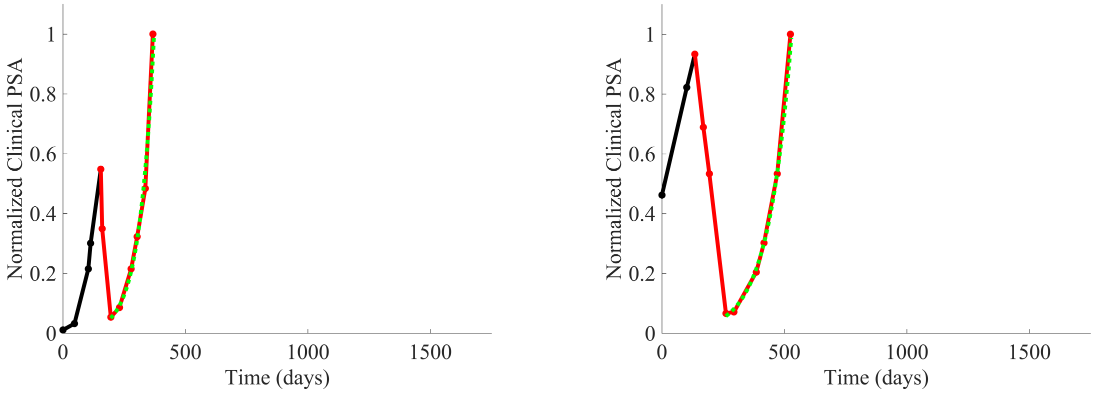


Figure S1: The PSA data for patients C005 (left) and C013 (right) are shown. Where the PSA dynamics are highlighted in red shows that abiraterone is being administered. The green dotted line shows the fit used to estimate the growth rate of the resistant cell populations after progression through abiraterone. As abiraterone is still being administered and cell populations are still growing, we assume that sensitive cells have been eliminated and only abiraterone resistant cells remain.

To estimate the population growth rates of sensitive cells (Table S1), we used time series of increasing PSA levels prior to any treatment with abiraterone. Prior to treatment, we expect that virtually all cancer cells are abiraterone sensitive cells. Ten patients from the contemporaneous cohort provided sufficient data. Figure S2 provides examples from two patients of these data and analyses.

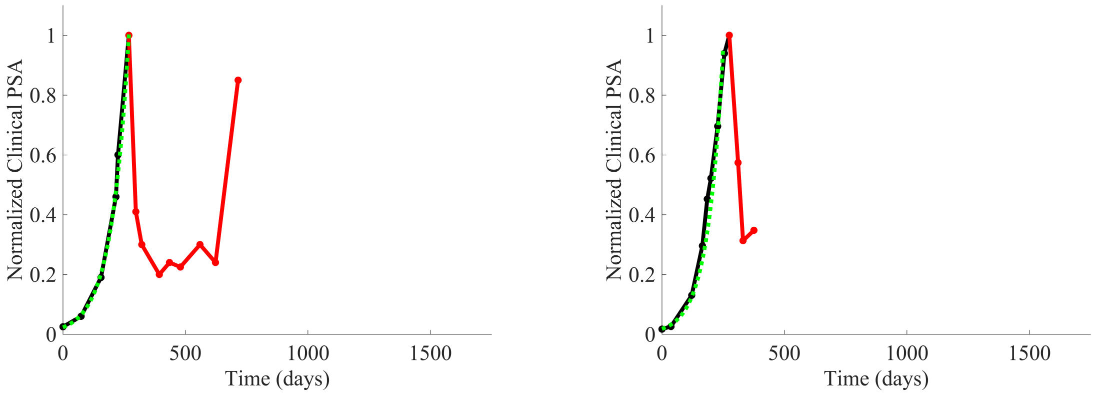


Figure S2: The PSA data for patients C014 (left) and C011 (right) are shown. Where the PSA dynamics are highlighted in red shows that abiraterone is being administered. The green dotted line shows the fit used to estimate the growth rate of the sensitive cell population before any treatment with abiraterone. As abiraterone has not yet been introduced, this growth is assumed to be completely from the abiraterone sensitive cells.

From patients on the adaptive therapy trial, we estimated the growth rates of sensitive cells using increases in PSA values during periods of the therapy cycle when patients were not receiving abiraterone. During such periods we assume that growth comes predominantly from the abiraterone sensitive population. Thirteen adaptive therapy patients were used to estimate the growth rates of sensitive cells. A given patient could provide from 1 to 6 estimates based on the number of therapy cycles. Figure S3 provides examples for two such patients.

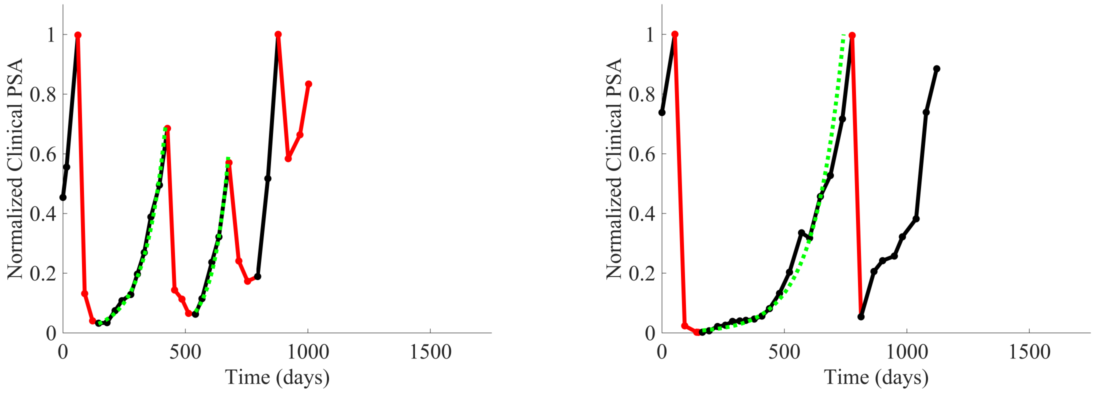


Figure S3: PSA data for patients P1016 (left) and P1017 (right). PSA dynamics highlighted in red and black show periods of abiraterone therapy on and off, respectively. Green dotted lines show exponential fits to periods when abiraterone was off. These provide estimates for the growth rates of the sensitive cell population during the off treatment period of an adaptive therapy cycle. We obtained two estimated values for P1016 and one for P1017. When off abiraterone, we assume that the growth in tumor burdens comes predominantly from the abiraterone sensitive cells.

| Sensitive Cells | | Resistant Cells | |
|---|---|---|---|
| Patient Identifier | Extracted Growth Rate | Patient Identifier | Extracted Growth Rate |
| C010 | 0.0071 | C013 | 0.0109 |
| C014 | 0.0142 | C012 | 0.0025 |
| C012 | 0.0075 | C010 | 0.0046 |
| C011 | 0.0123 | C005 | 0.0173 |
| C009 | 0.0107 | C008 | 0.0047 |
| C007 | 0.0062 | C007 | 0.0111 |
| C005 | 0.0214 | C006 | 0.0130 |
| C004 | 0.0196 | C005 | 0.0173 |
| C003 | 0.0100 | C004 | 0.0124 |
| C001 | 0.0062 | C002 | 0.0031 |
| P1018 | 0.0189 | C001 | 0.0022 |
| P1017 | 0.0068 | | |
| P1016 | 0.0106 | | |
| P1016 | 0.0146 | | |
| P1015 | 0.0419 | | |
| P1014 | 0.0109 | | |
| P1012 | 0.0059 | | |
| P1012 | 0.0076 | | |
| P1012 | 0.0091 | | |
| P1012 | 0.0160 | | |
| P1012 | 0.0100 | | |
| P1012 | 0.0170 | | |
| P1011 | 0.0191 | | |
| P1011 | 0.0191 | | |
| P1011 | 0.0304 | | |
| P1007 | 0.0118 | | |
| P1006 | 0.0216 | | |
| P1004 | 0.0071 | | |
| P1003 | 0.0124 | | |
| P1003 | 0.0116 | | |
| P1003 | 0.0105 | | |
| P1002 | 0.0245 | | |
| P1001 | 0.0446 | | |
| P1001 | 0.0317 | | |

Table S1: Estimates of sensitive and resistant growth rates from patient data in both the contemporaneous and adaptive therapy cohorts.

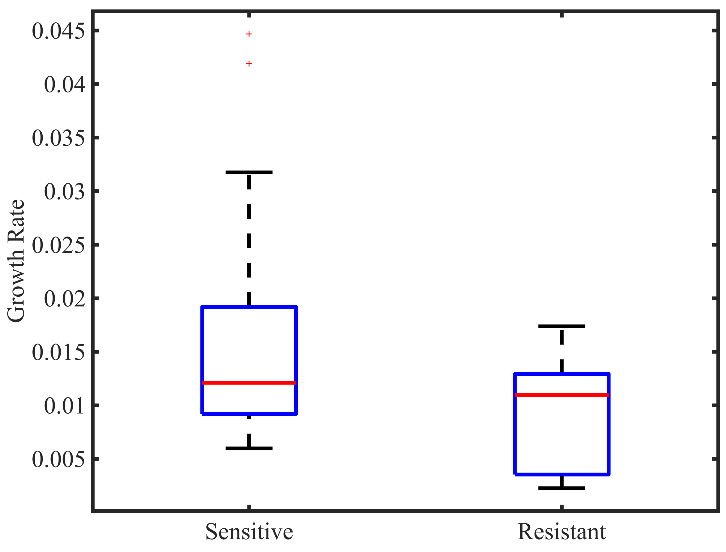


Figure S4: Box plot of sensitive and resistant growth rates.

Using estimates shown in Table S1, sensitive cells had a significantly higher mean growth rate (0.0156) than that for resistant cells (0.0091), $t_x = 2.11, p < 0.05$. For fitting patient data, we fixed the growth rates of sensitive and resistant cancer cells to $r_S = 0.0156$ and $r_R = 0.0091$. Because the actual PSA growth rates may be dampened by limits to growth, these estimates for the cancer cells' intrinsic growth rates may be underestimates. Higher intrinsic growth rates may better reflect the dynamics of the Lotka-Volterra competition model. Hence for fitting the L-V model to the patients' data we included a multiplier $\beta_{SC}$ as a scaling factor for the PSA dynamics.

### S2 Parameter Optimization

With the growth rates set to $r_S = 0.0156$ and $r_R = 0.0091$ for all patients, there remain just four parameters for fitting the patient data (Table S2). The parameter optimization seeks to estimate values for the ecological scaling of the growth rates $\beta_{SC}$, the competitive effect of sensitive cells on resistant cells $\alpha_{RS}$, and the initial tumor composition of sensitive $y_S(0)$ and resistant $y_R(0)$ cells. In what follows we set the competitive effect of resistant cells on sensitive cells to $\alpha_{SR} = 1$ which may be an overestimate if sensitive cells are weak competitors, but which provides another fixed parameter for all patients. Furthermore, like the growth rates, we shall make across patient estimates of $\beta_{SC}$ and $\alpha_{RS}$. In this way, we assume strong convergent and parallel evolution of prostate cancer between patients. We will let the initial population sizes of sensitive and resistant cancer cells be patient specific.

| **Parameter** | **Definition** |
|---|---|
| $y_S(0)$ | Abundance of sensitive cells at the time of initial treatment. |
| $y_R(0)$ | Abundance of resistant cells at the time of initial treatment. |
| $\alpha_{RS}$ | Competitive effect of sensitive cells on resistant cells. |
| $\beta_{SC}$ | Ecological scale factor. |

Table S2: Table of parameters and definitions to be optimized using constrained optimization.

We implemented constrained nonlinear multivariable optimization using the Matlab optimization toolbox (`fmincon`) in order to find the combination of variables presented in Table S2 that minimized the cumulative least squares difference between the output of the model and the actual patient data over the entire cohort. Patient PSA and model PSA are normalized to the maximum PSA value for that patient or simulation. In this way all PSA values of the patient data and the modeled data were $\in [0,1]$. The optimization minimizes the mean square error between the scaled PSA of the model and the scaled PSA of the patient data. Extra weight is given to the data points when a change in treatment occurs where using a weighting term of $w = 5$ at these points and $w = 1$ at all other points. Constraints on each of the four variables are presented in Table S3.

| **Parameter** | **Constraint** |
|---|---|
| $y_S(0)$ | $\in [1, 10000]$ |
| $y_R(0)$ | $\in [1, 10000]$ |
| $\alpha_{RS}$ | $\in [0, 20]$ |
| $\beta_{SC}$ | $\in [0, 20]$ |

Table S3: Table of constraints on parameters to be optimized using constrained optimization.

### S3 Optimized Parameters

The minimal mean square error was found with the optimized parameters as follows: $\alpha_{RS} = 6$ and $\beta_{SC} = 8$. The patient-specific estimates for the initial population sizes of sensitive and resistant cells with $\alpha_{RS} = 6$ and $\beta_{SC} = 8$ are shown in Table S4.

In the following analysis where we consider how each patient might have fared under different treatment protocols, we keep the patient-specific parameters as given in Table S4. This includes the patients that showed relatively poorer fits. We feel this maintains a conservative approach to the subsequent exploration of treatment options with the caveat that additional error propagation might occur insofar as other model parameters may indeed be patient specific.

| **Patient** | $y_S(0)$ | $y_R(0)$ | $\alpha_{RS}$ | $\beta_{SC}$ |
|---|---|---|---|---|
| P1001 | 86.74 | 1.00 | 6 | 8 |
| P1002 | 165.99 | 24.67 | 6 | 8 |
| P1003 | 10000.00 | 124.98 | 6 | 8 |
| P1004 | 19.90 | 400.57 | 6 | 8 |
| P1005 | 802.20 | 12.31 | 6 | 8 |
| P1006 | 102.28 | 174.59 | 6 | 8 |
| P1007 | 253.07 | 149.12 | 6 | 8 |
| P1009 | 20.41 | 23.61 | 6 | 8 |
| P1010 | 196.09 | 1001.19 | 6 | 8 |
| P1011 | 29.19 | 6.31 | 6 | 8 |
| P1012 | 567.60 | 663.13 | 6 | 8 |
| P1014 | 1640.76 | 273.61 | 6 | 8 |
| P1015 | 126.13 | 6.38 | 6 | 8 |
| P1016 | 4589.21 | 4078.11 | 6 | 8 |
| P1017 | 6035.16 | 4060.25 | 6 | 8 |
| P1018 | 1495.72 | 318.18 | 6 | 8 |
| P1020 | 2061.34 | 477.07 | 6 | 8 |
| C001 | 111.70 | 19.46 | 6 | 8 |
| C002 | 1867.68 | 1.00 | 6 | 8 |
| C003 | 61.96 | 5.81 | 6 | 8 |
| C004 | 110.57 | 156.36 | 6 | 8 |
| C005 | 66.07 | 127.48 | 6 | 8 |
| C006 | 1226.45 | 25.31 | 6 | 8 |
| C007 | 1740.08 | 1325.03 | 6 | 8 |
| C008 | 4362.58 | 24.08 | 6 | 8 |
| C009 | 148.30 | 936.54 | 6 | 8 |
| C010 | 273.95 | 798.61 | 6 | 8 |
| C011 | 19.85 | 129.22 | 6 | 8 |
| C012 | 2148.71 | 301.47 | 6 | 8 |
| C013 | 165.20 | 31.48 | 6 | 8 |
| C014 | 13.57 | 5.09 | 6 | 8 |
| C015 | 1211.43 | 483.69 | 6 | 8 |

Table S4: Optimized parameters for each patient resulting from nonlinear constrained optimization.

These initial conditions allowed for the extraction of the tumor composition at the time abiraterone was first administered clinically to each patient, which was not always at t=0. The relative fraction of the resistant to sensitive cells at the time of initial treatment is compared to the time to progression (months) reported for each patient (Table S5).

| **Patient** | $y_S(t_{ABI})$ | $y_R(t_{ABI})$ | $\frac{y_r(t_{ABI})}{y_S(t_{ABI})}\%$ | **TTP** |
|---|---|---|---|---|
| P1001 | 2158.32 | 52.35 | 2.43 | 30.6 |
| P1003 | 5782.98 | 105.32 | 1.82 | 53.1 |
| P1004 | 3475.70 | 1621.81 | 46.66 | 11.0 |
| P1005 | 4407.49 | 7.77 | 0.18 | 38.0 |
| P1006 | 2483.02 | 603.80 | 24.32 | 42.8 |
| P1007 | 3503.41 | 203.61 | 5.81 | 30.1 |
| P1009 | 2830.43 | 156.89 | 5.54 | 17.0 |
| P1010 | 2407.93 | 1765.24 | 73.31 | 10.7 |
| P1011 | 3285.31 | 26.37 | 0.80 | 25.4 |
| P1012 | 2530.65 | 706.23 | 27.91 | 54.0 |
| P1014 | 4708.02 | 127.91 | 2.72 | 50.0 |
| P1015 | 3757.24 | 23.55 | 0.63 | 20.4 |
| P1016 | 6418.04 | 678.74 | 10.58 | 31.4 |
| P1017 | 7268.07 | 557.30 | 7.67 | 35.5 |
| P1018 | 1887.88 | 315.85 | 16.73 | 10.8 |
| P1020 | 3534.42 | 346.12 | 9.79 | 23.0 |
| C001 | 2250.42 | 136.14 | 6.05 | 9.0 |
| C002 | 1867.68 | 1.00 | 0.05 | 26.0 |
| C003 | 1976.15 | 32.96 | 1.67 | 15.0 |
| C004 | 2435.97 | 617.06 | 25.33 | 4.2 |
| C005 | 2696.58 | 600.39 | 22.26 | 7.0 |
| C006 | 1998.86 | 25.70 | 1.29 | 17.7 |
| C007 | 6552.52 | 166.29 | 2.54 | 17.3 |
| C008 | 4362.58 | 24.08 | 0.55 | 19.6 |
| C009 | 4545.92 | 787.31 | 17.32 | 6.5 |
| C010 | 4951.79 | 472.52 | 9.54 | 9.0 |
| C011 | 2953.60 | 772.31 | 26.15 | 4.0 |
| C012 | 7754.85 | 15.28 | 0.20 | 25.0 |
| C013 | 2113.11 | 72.78 | 3.44 | 13.8 |
| C014 | 2645.67 | 44.98 | 1.70 | 14.8 |
| C015 | 2356.24 | 683.69 | 29.02 | 3.2 |

Table S5: Tumor composition of sensitive and resistant populations at the time of initial abiraterone therapy from the optimized model fits to patient data. The calculated percentage of resistant to sensitive cells alongside the clinical time to progression for each patient is also shown.

#### S4 Optimized Model Fits

For each patient, the optimized model fits are shown with patient-specific parameters from Table S4.

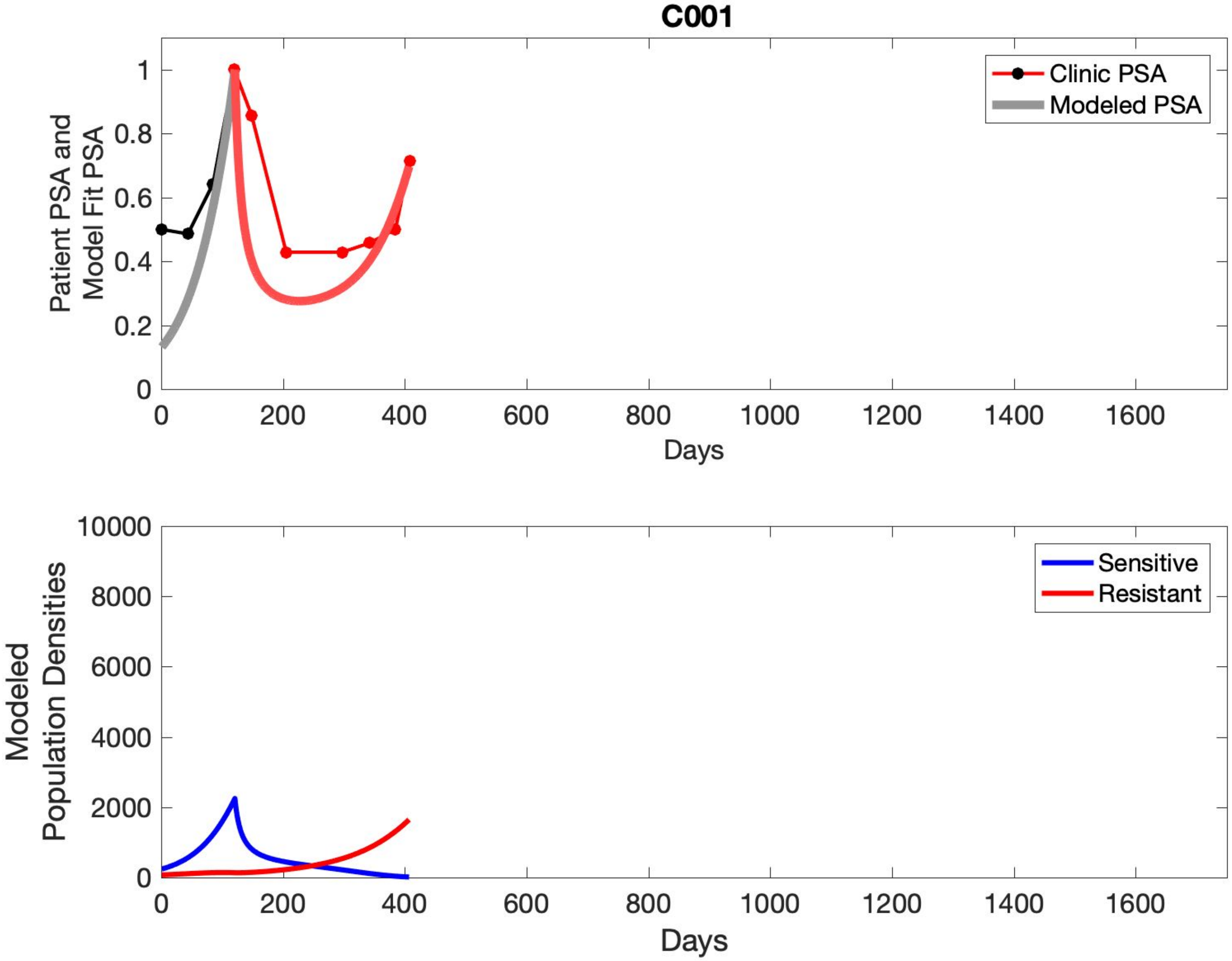
C001
Clinic PSA
Modeled PSA
Patient PSA and Model Fit PSA
0
0.2
0.4
0.6
0.8
1
0
200
400
600
800
1000
1200
1400
1600
Days
Sensitive
Resistant
Modeled Population Densities
0
2000
4000
6000
8000
10000
Days

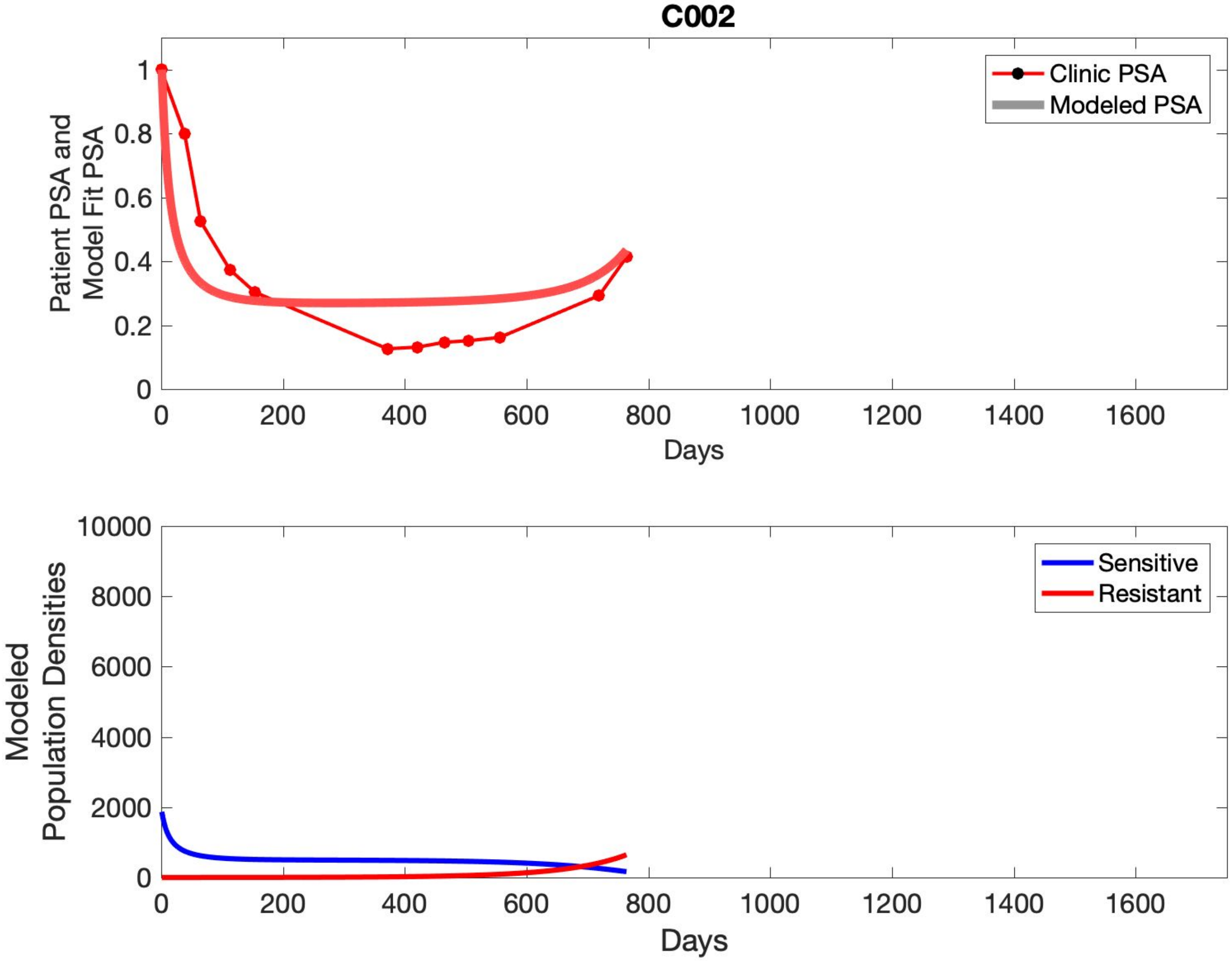

C002
Patient PSA and Model Fit PSA
Clinic PSA
Modeled PSA
Days
Modeled Population Densities
Sensitive
Resistant
Days

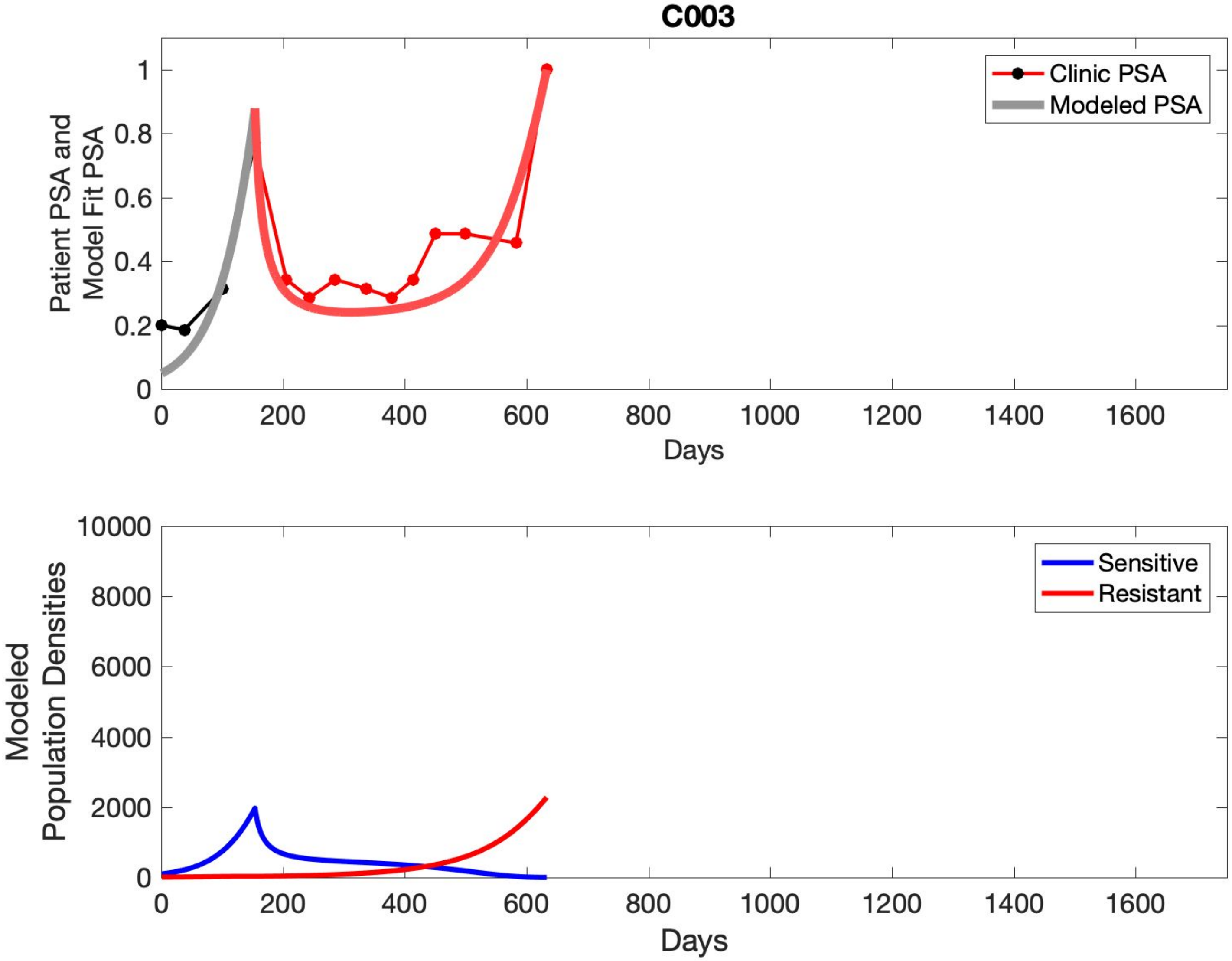

C003
Patient PSA and Model Fit PSA
Clinic PSA
Modeled PSA
Days
Modeled Population Densities
Sensitive
Resistant
Days

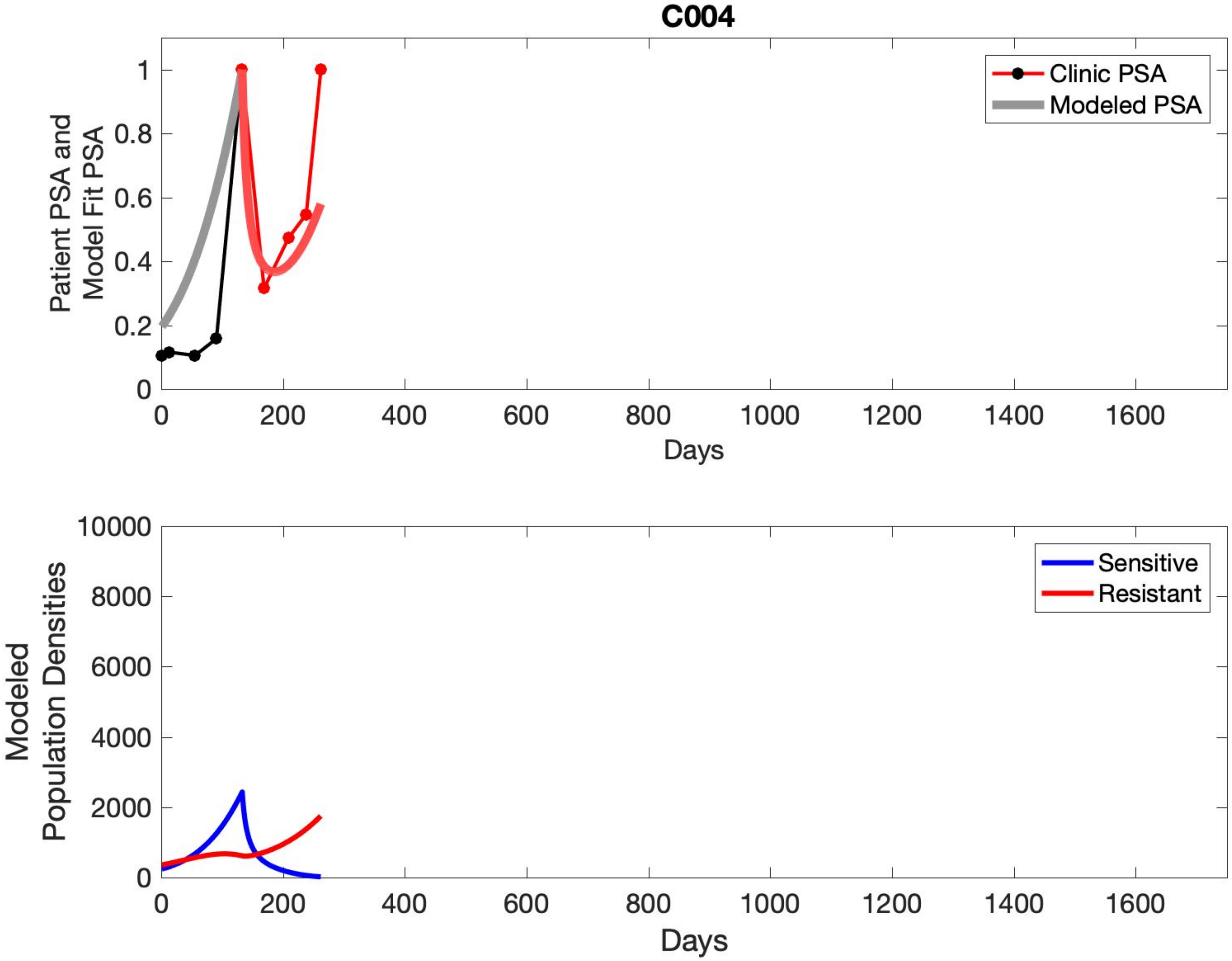

C004
Patient PSA and Model Fit PSA
Clinic PSA
Modeled PSA
0
0.2
0.4
0.6
0.8
1
0
200
400
600
800
1000
1200
1400
1600
Days
Modeled Population Densities
Sensitive
Resistant
0
2000
4000
6000
8000
10000
0
200
400
600
800
1000
1200
1400
1600
Days

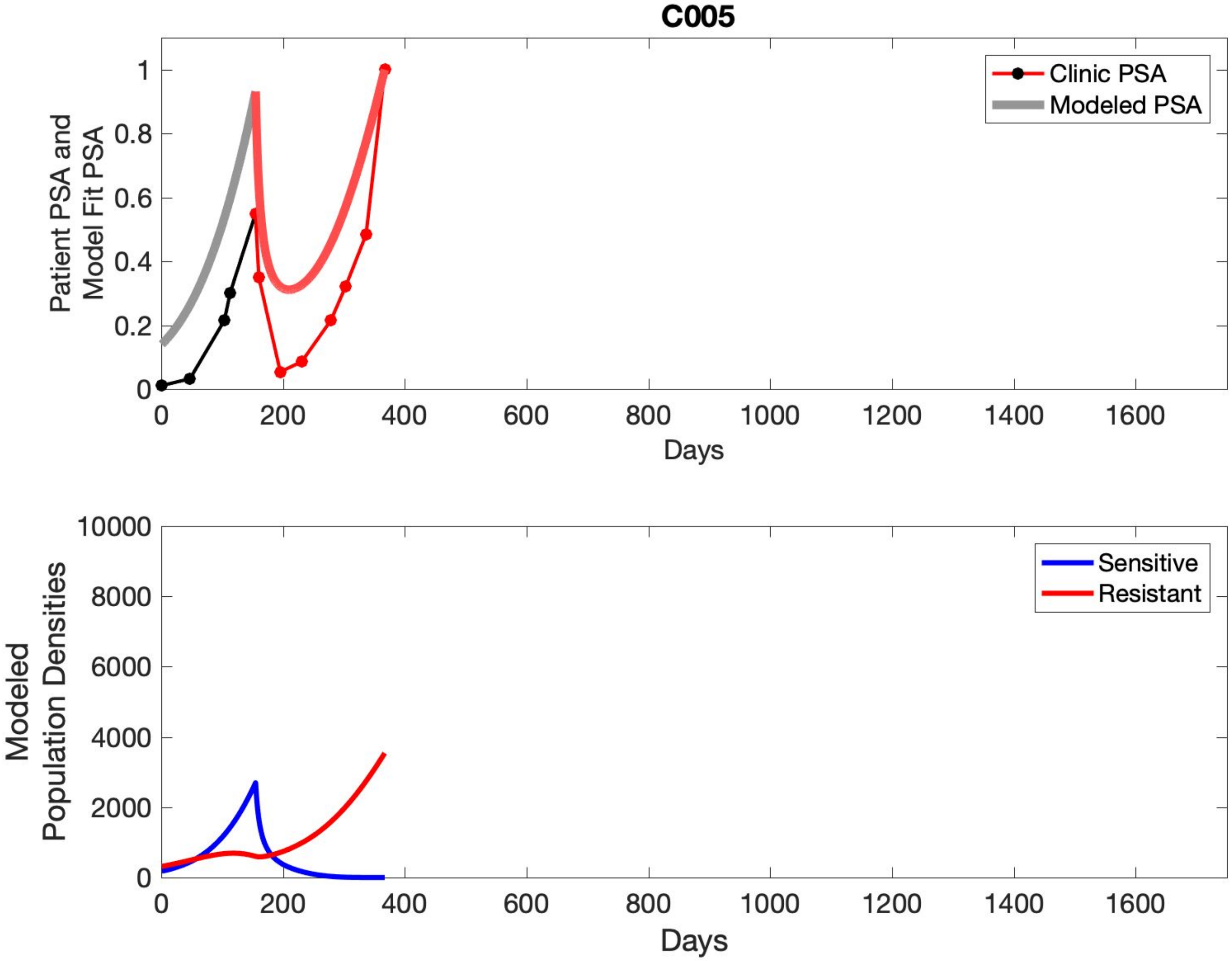
C005
Clinic PSA
Modeled PSA
Patient PSA and Model Fit PSA
0
0.2
0.4
0.6
0.8
1
0
200
400
600
800
1000
1200
1400
1600
Days
Sensitive
Resistant
Modeled Population Densities
0
2000
4000
6000
8000
10000
0
200
400
600
800
1000
1200
1400
1600
Days

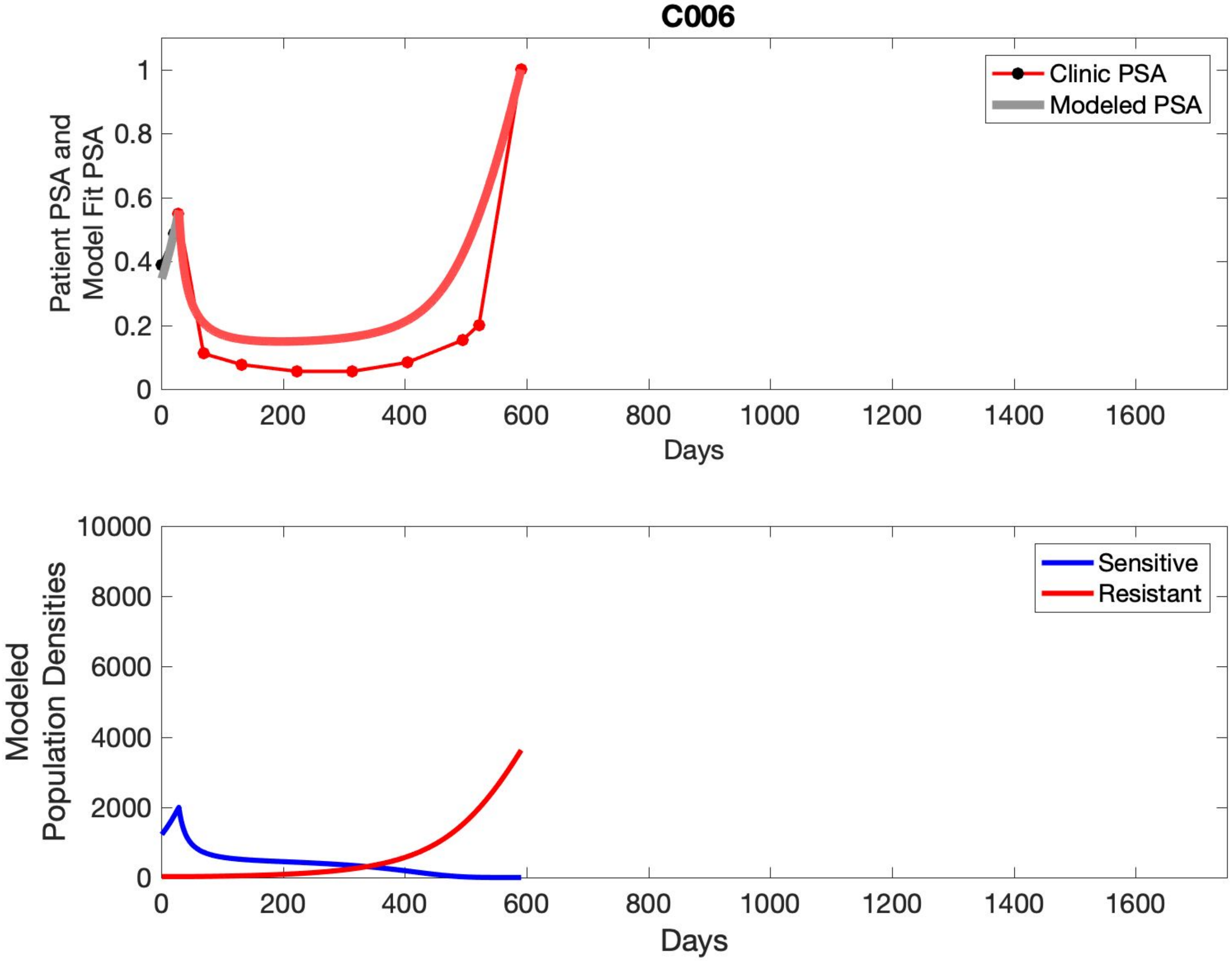

C006
Patient PSA and Model Fit PSA
Clinic PSA
Modeled PSA
Days
Modeled Population Densities
Sensitive
Resistant
Days

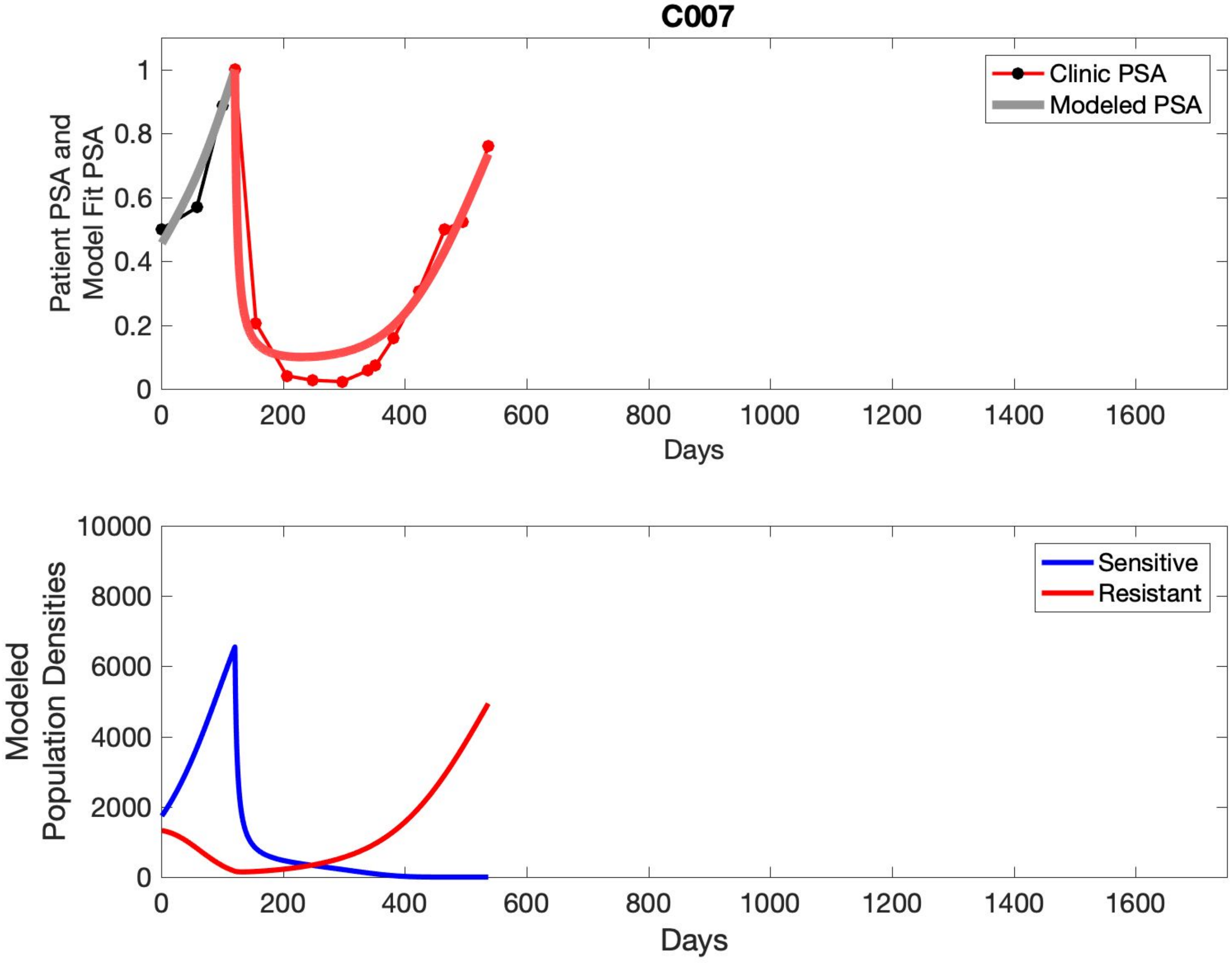

C007
Clinic PSA
Modeled PSA
Patient PSA and
Model Fit PSA
Days
Sensitive
Resistant
Modeled
Population Densities
Days

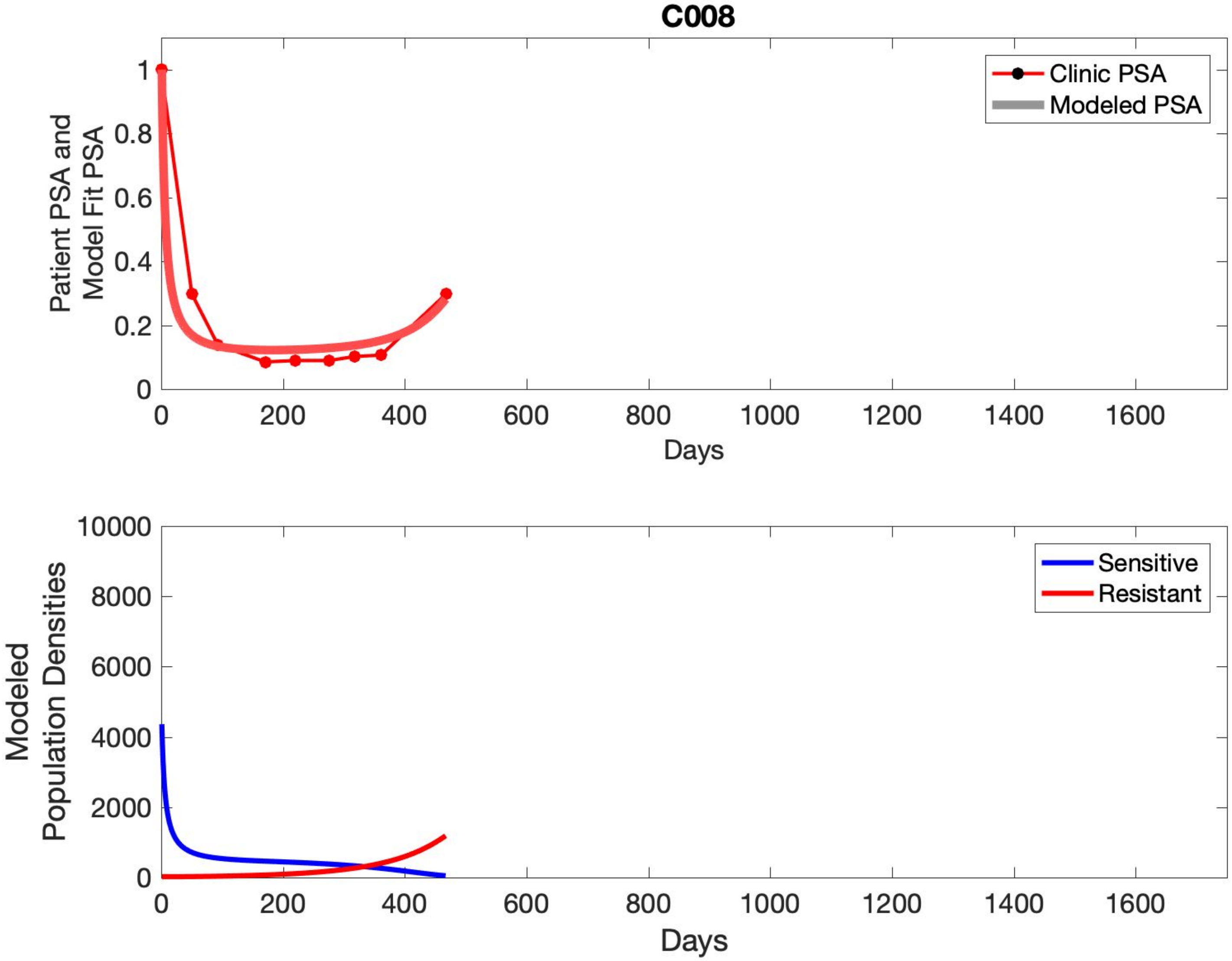
C008
Patient PSA and Model Fit PSA
Clinic PSA
Modeled PSA
Days
Modeled Population Densities
Sensitive
Resistant
Days

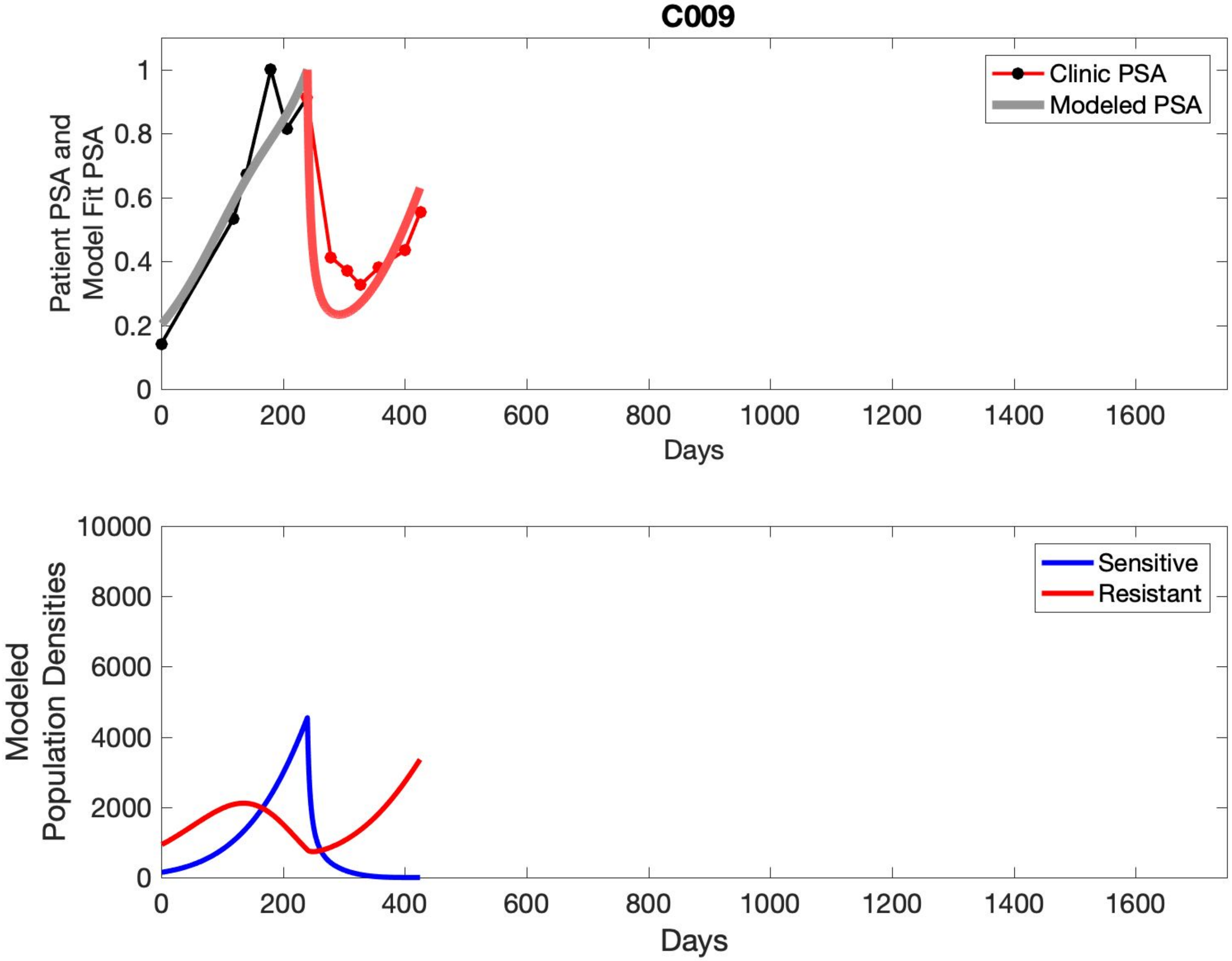
C009
Patient PSA and Model Fit PSA
Clinic PSA
Modeled PSA
0
0.2
0.4
0.6
0.8
1
0
200
400
600
800
1000
1200
1400
1600
Days
Modeled Population Densities
Sensitive
Resistant
0
2000
4000
6000
8000
10000
0
200
400
600
800
1000
1200
1400
1600
Days

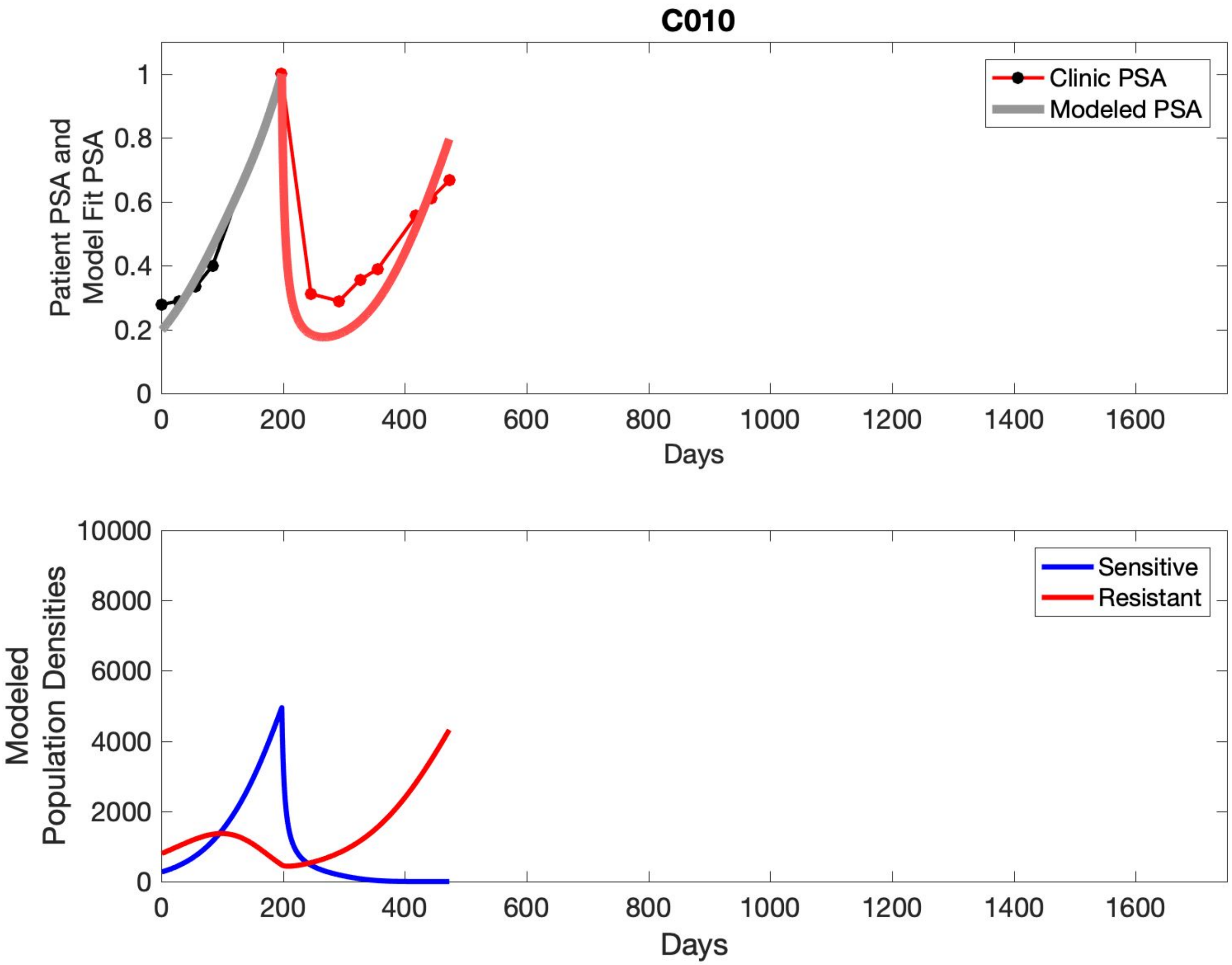

C010
Clinic PSA
Modeled PSA
Patient PSA and
Model Fit PSA
0
0.2
0.4
0.6
0.8
1
0
200
400
600
800
1000
1200
1400
1600
Days
Sensitive
Resistant
Modeled
Population Densities
0
2000
4000
6000
8000
10000
0
200
400
600
800
1000
1200
1400
1600
Days

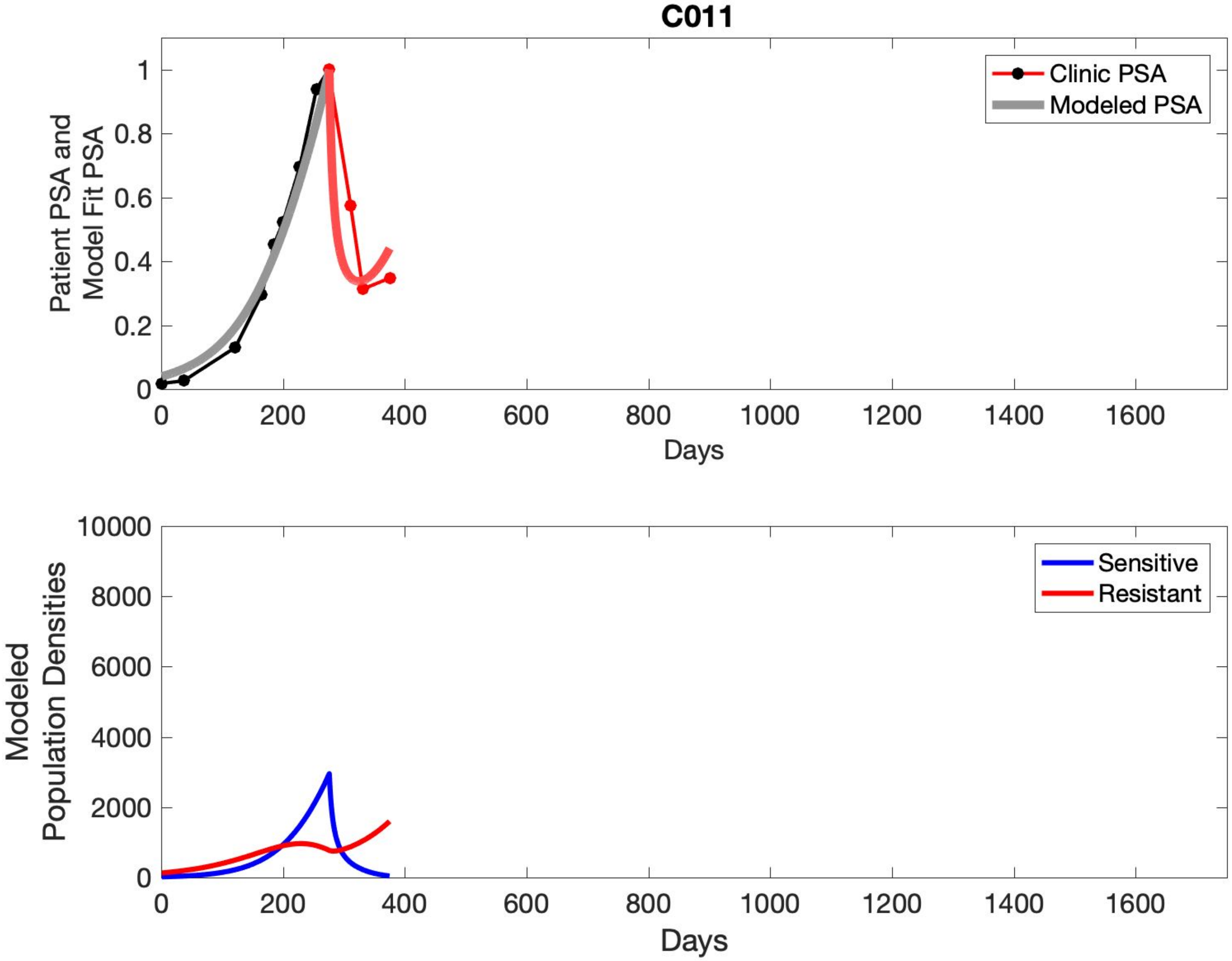

C011
Clinic PSA
Modeled PSA
Patient PSA and Model Fit PSA
0
0.2
0.4
0.6
0.8
1
0
200
400
600
800
1000
1200
1400
1600
Days
Sensitive
Resistant
Modeled Population Densities
0
2000
4000
6000
8000
10000
0
200
400
600
800
1000
1200
1400
1600
Days

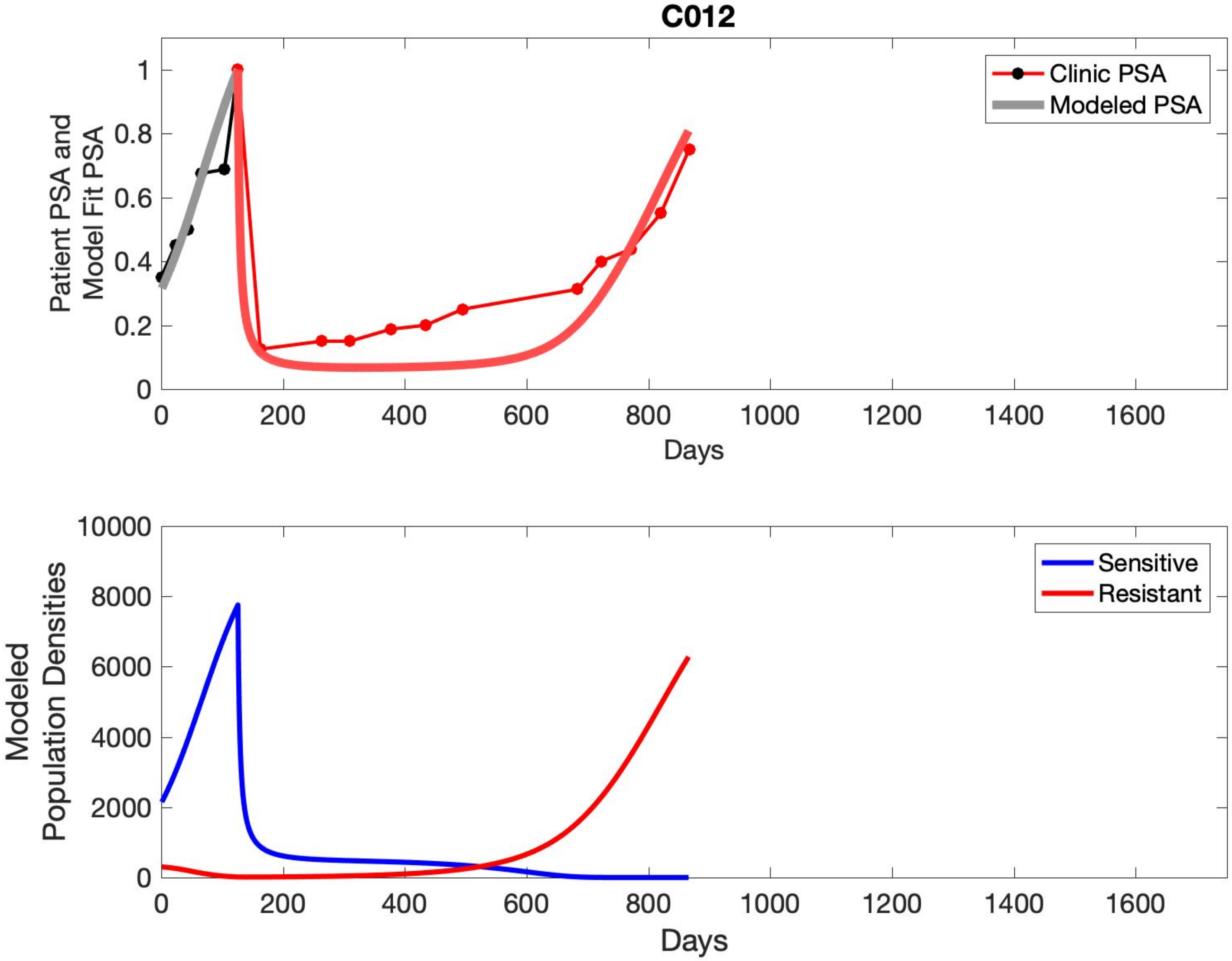
C012
Clinic PSA
Modeled PSA
Patient PSA and
Model Fit PSA
0
0.2
0.4
0.6
0.8
1
0
200
400
600
800
1000
1200
1400
1600
Days
Sensitive
Resistant
Modeled
Population Densities
0
2000
4000
6000
8000
10000
Days

## C013

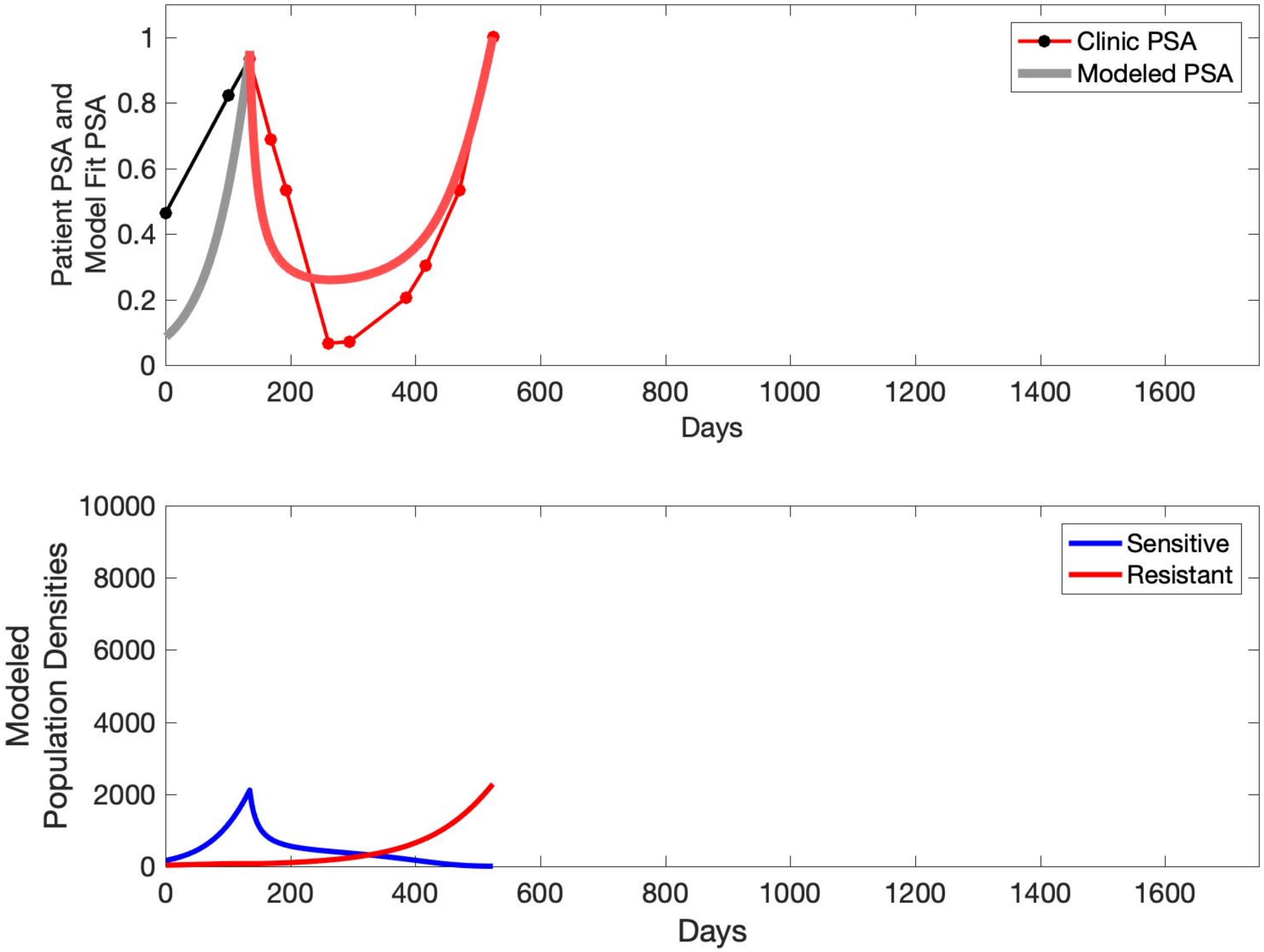
Clinic PSA
Modeled PSA
Patient PSA and Model Fit PSA
Days
Sensitive
Resistant
Modeled Population Densities
Days

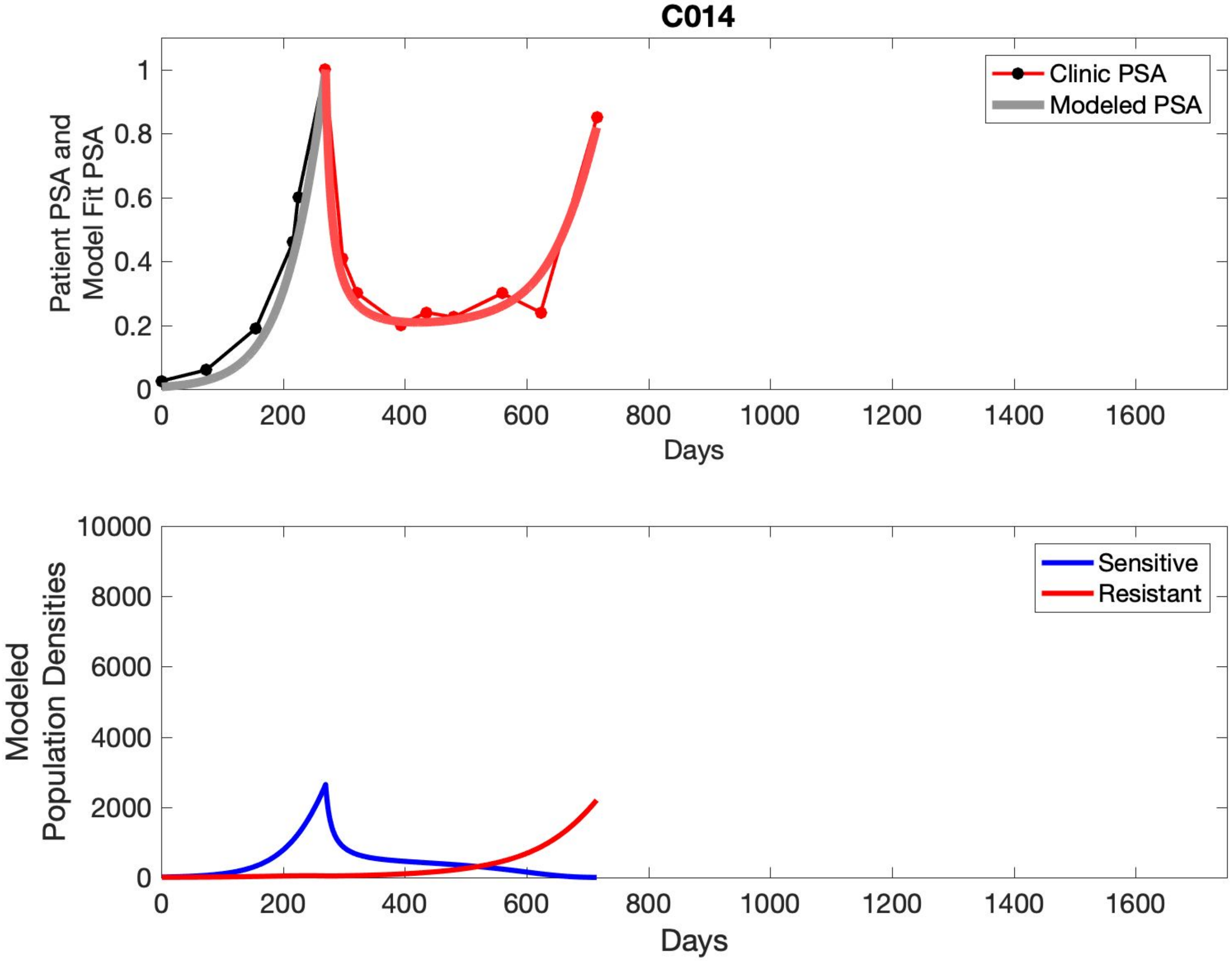

C014
Patient PSA and Model Fit PSA
Clinic PSA
Modeled PSA
0
0.2
0.4
0.6
0.8
1
0
200
400
600
800
1000
1200
1400
1600
Days
Modeled Population Densities
Sensitive
Resistant
0
2000
4000
6000
8000
10000
0
200
400
600
800
1000
1200
1400
1600
Days

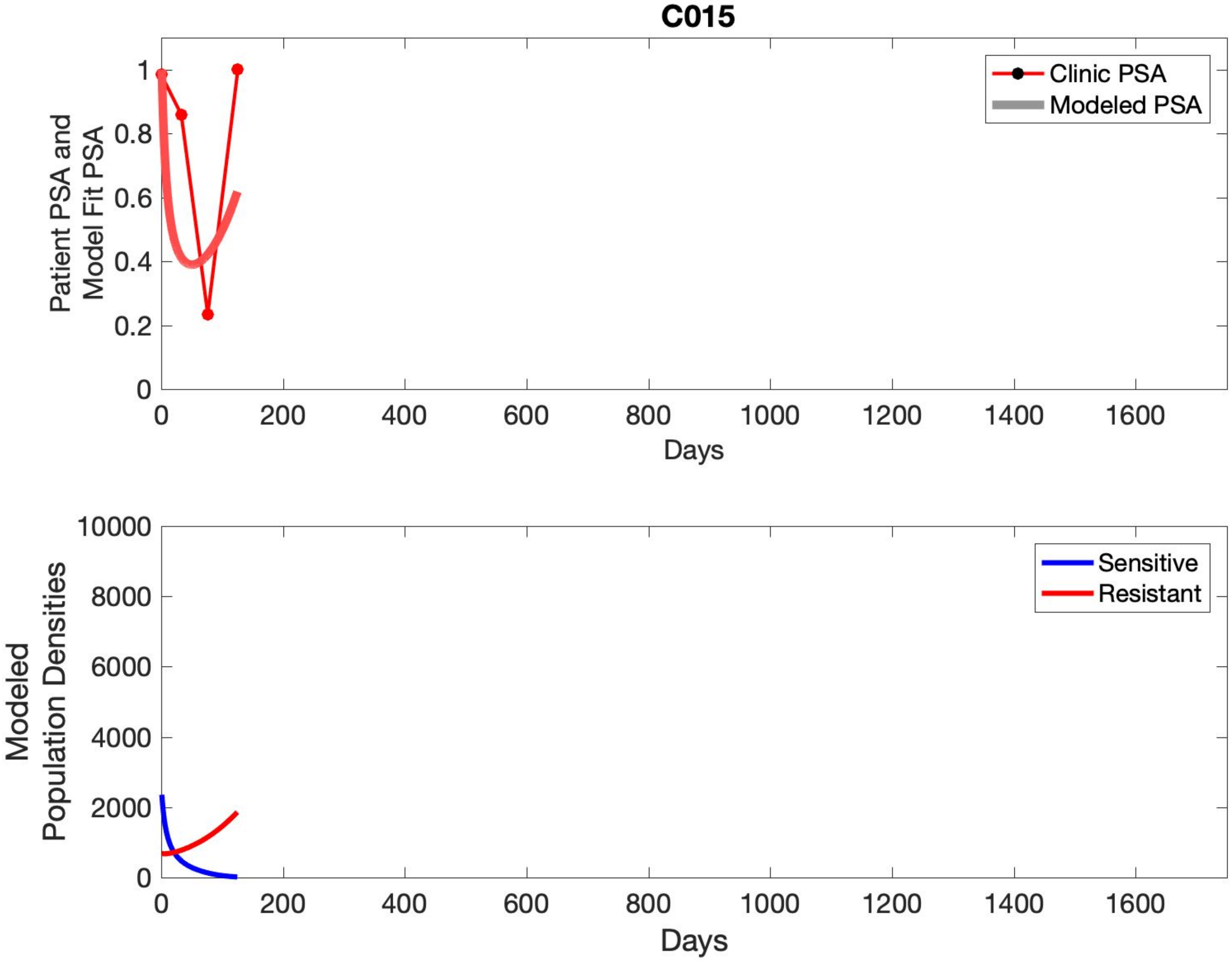
C015
Clinic PSA
Modeled PSA
Patient PSA and Model Fit PSA
Days
Sensitive
Resistant
Modeled Population Densities
Days

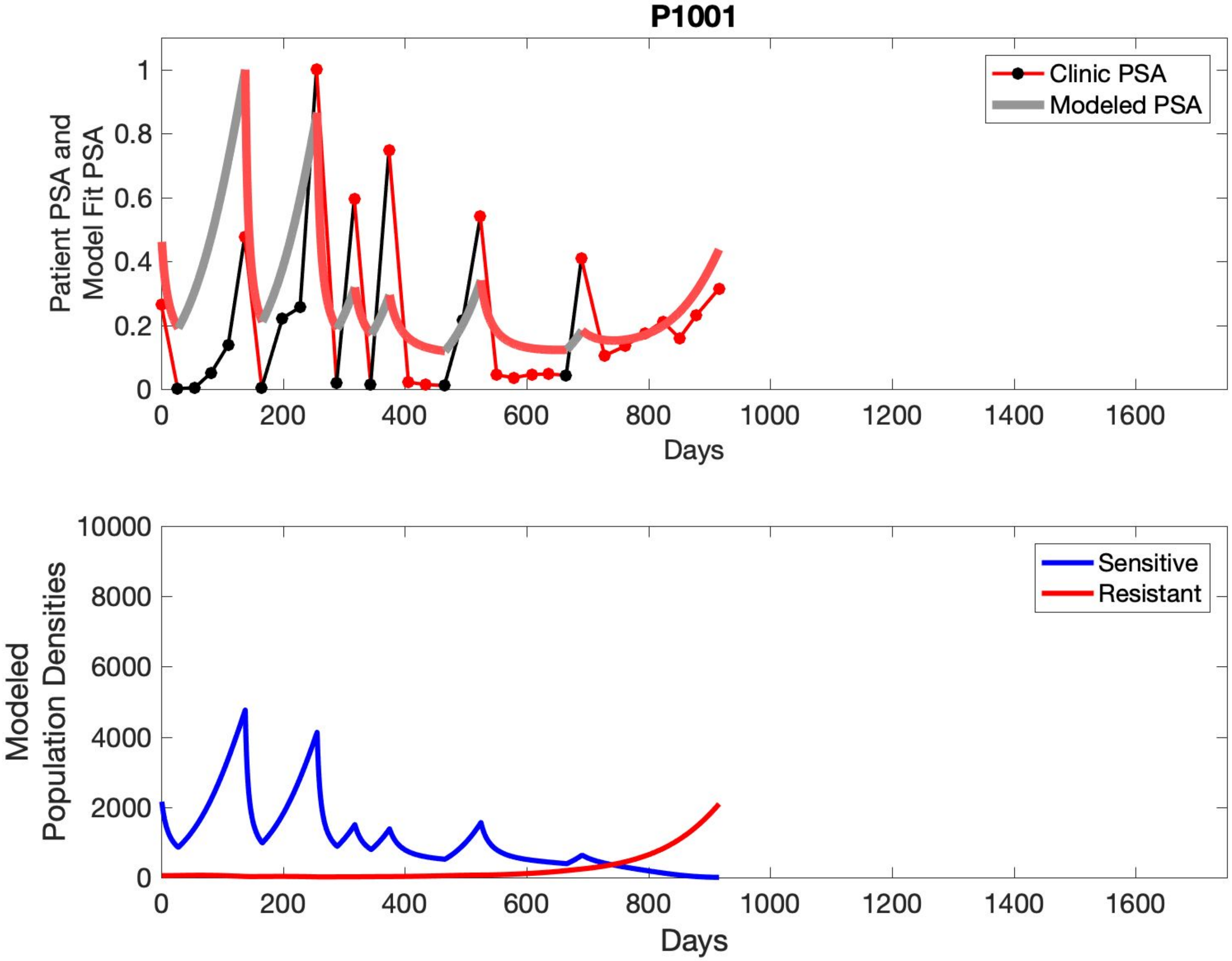

P1001
Patient PSA and Model Fit PSA
Clinic PSA
Modeled PSA
Days
Modeled Population Densities
Sensitive
Resistant
Days

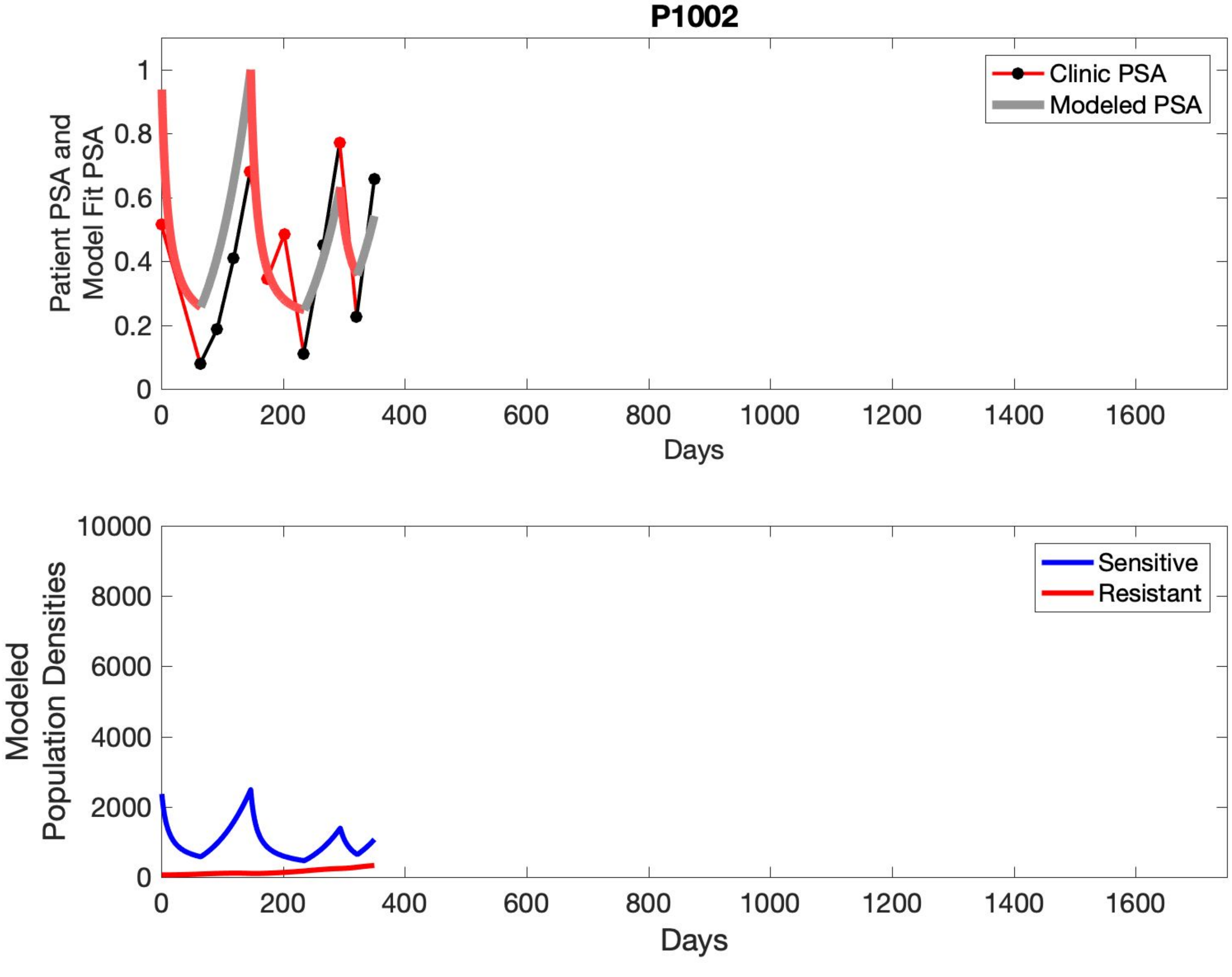
P1002
Patient PSA and Model Fit PSA
Clinic PSA
Modeled PSA
0
0.2
0.4
0.6
0.8
1
0
200
400
600
800
1000
1200
1400
1600
Days
Modeled Population Densities
Sensitive
Resistant
0
2000
4000
6000
8000
10000
0
200
400
600
800
1000
1200
1400
1600
Days

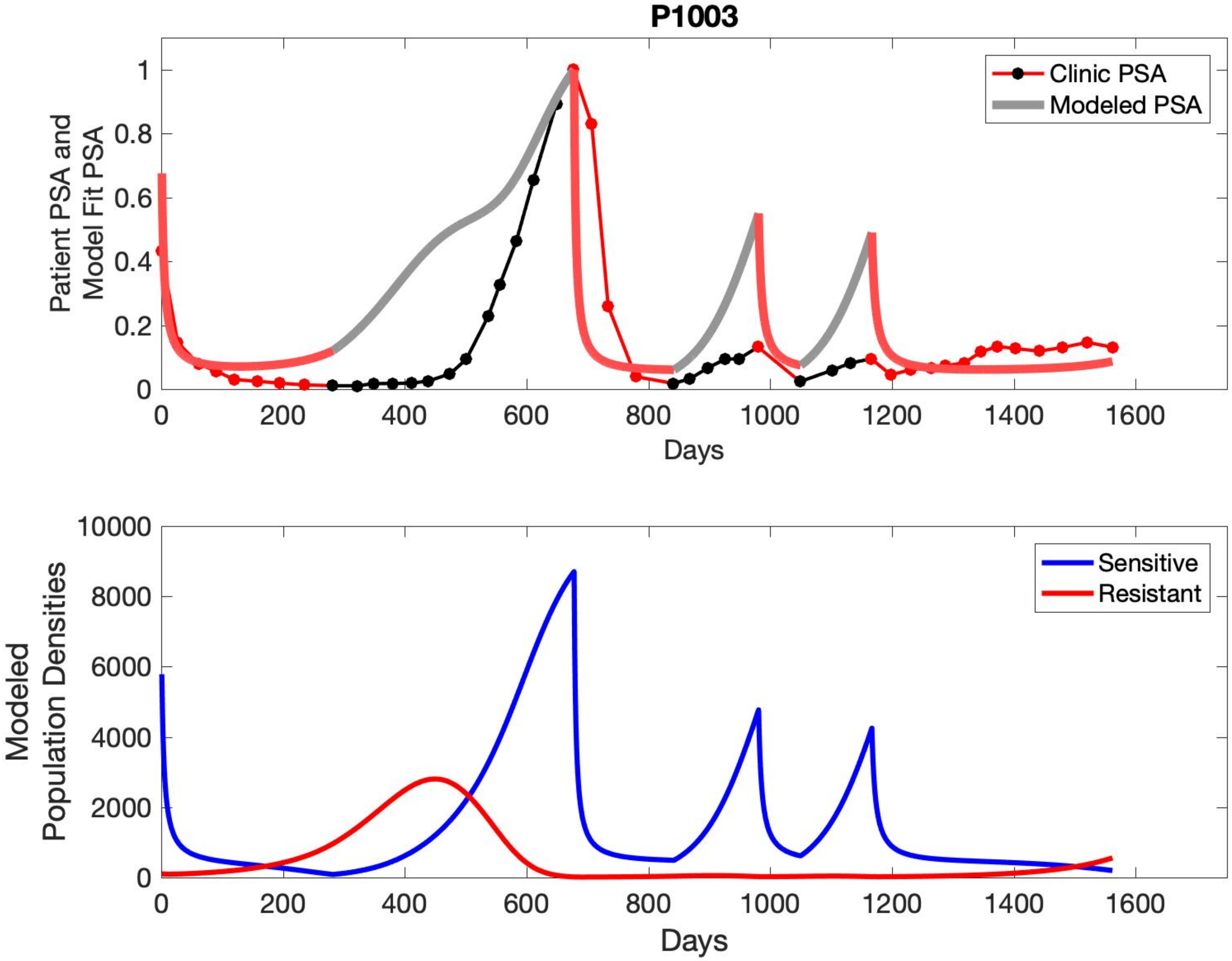
P1003
Patient PSA and Model Fit PSA
Days
Clinic PSA
Modeled PSA
Modeled Population Densities
Days
Sensitive
Resistant

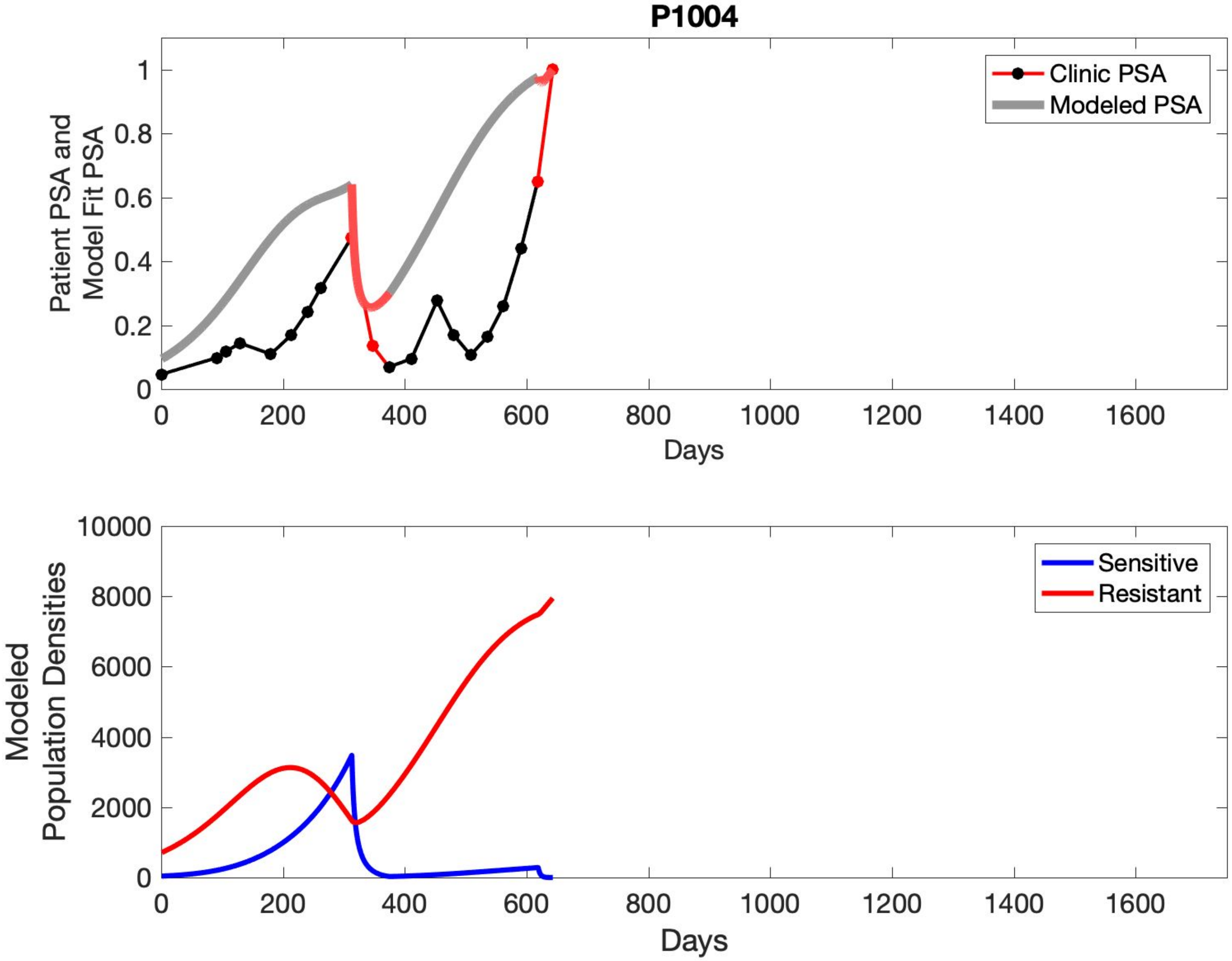

P1004
Patient PSA and Model Fit PSA
Clinic PSA
Modeled PSA
0
0.2
0.4
0.6
0.8
1
0
200
400
600
800
1000
1200
1400
1600
Days
Modeled Population Densities
Sensitive
Resistant
0
2000
4000
6000
8000
10000
0
200
400
600
800
1000
1200
1400
1600
Days

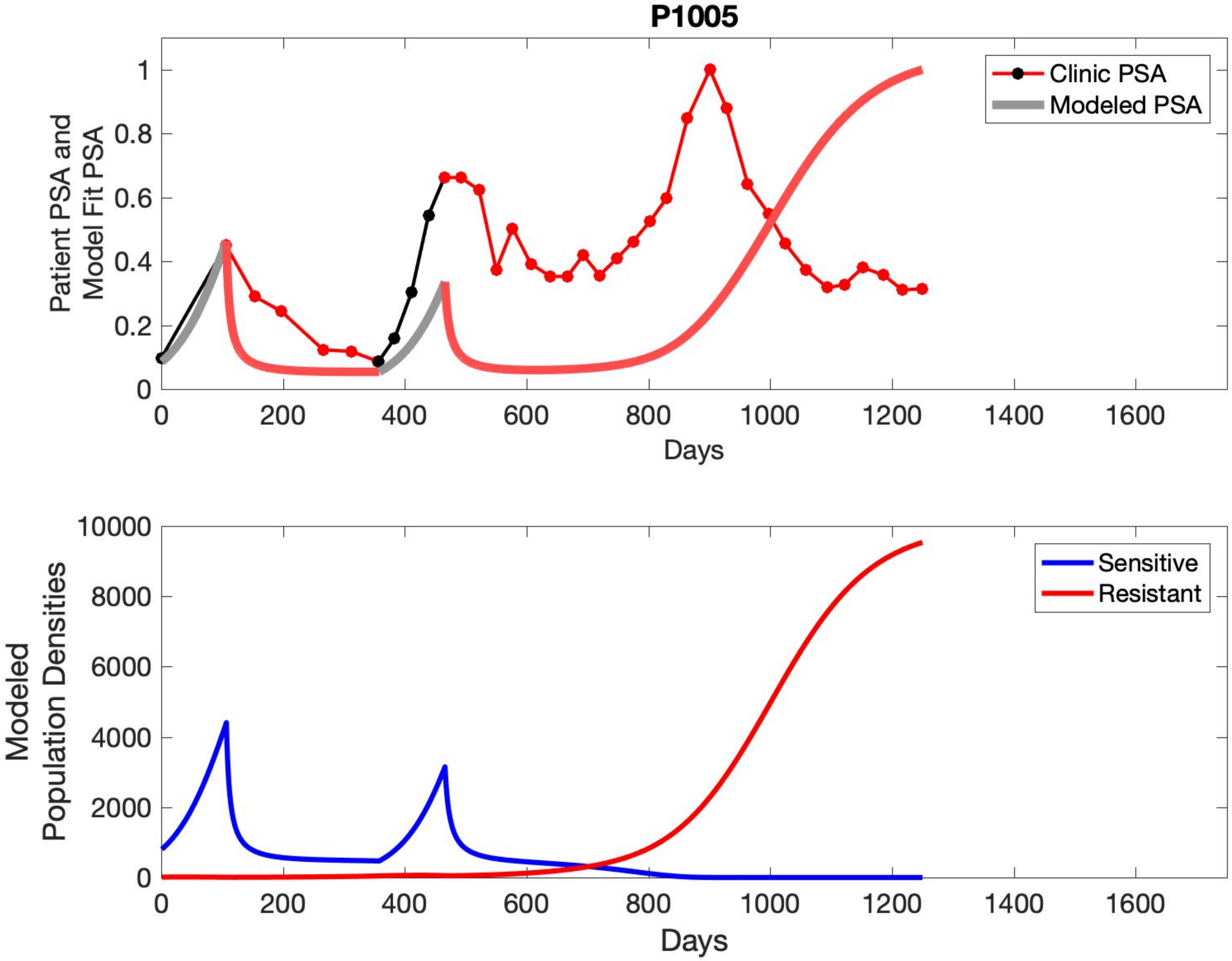

P1005
Patient PSA and Model Fit PSA
Clinic PSA
Modeled PSA
Days
Modeled Population Densities
Sensitive
Resistant
Days

## P1006

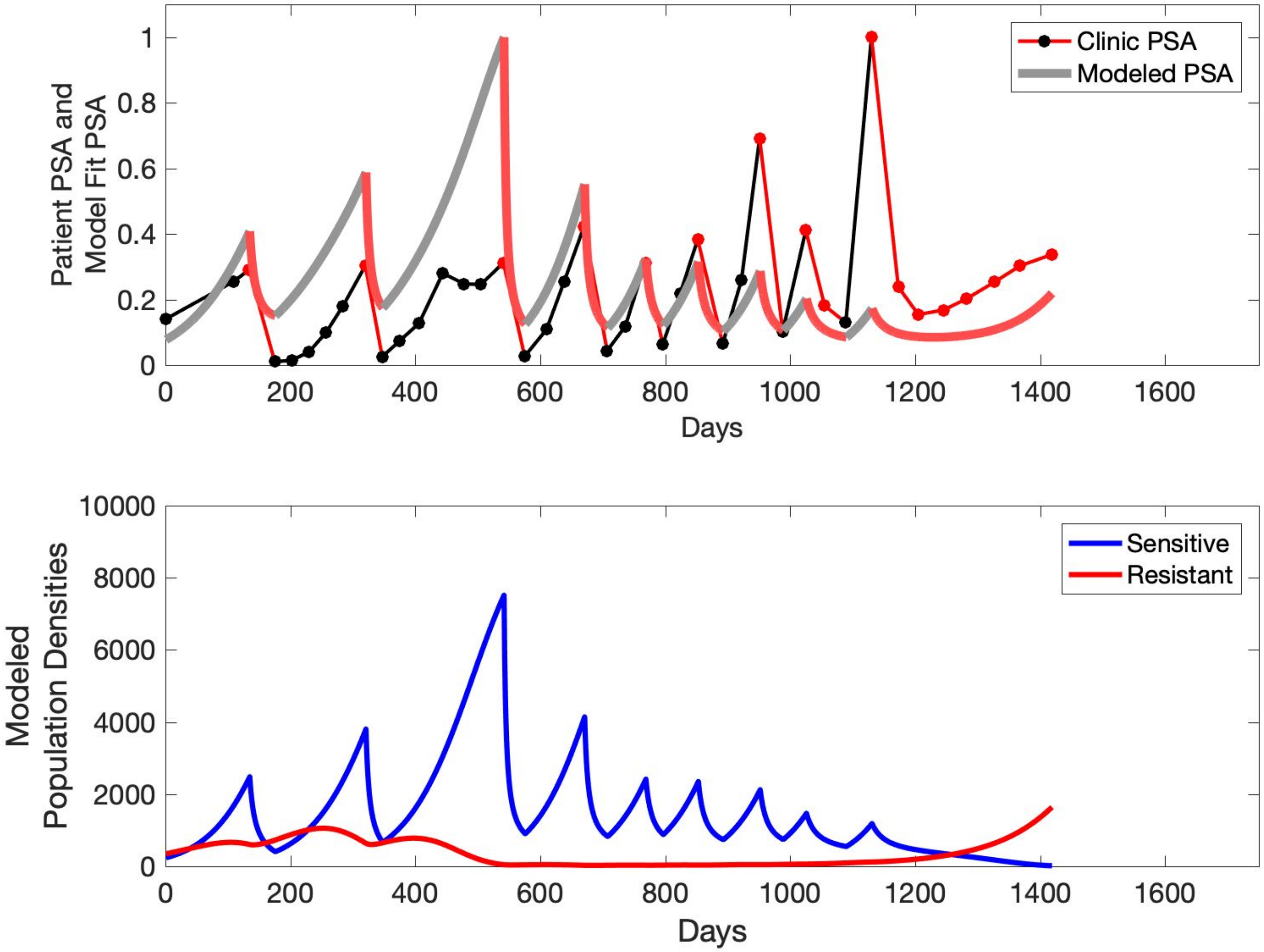

Patient PSA and
Model Fit PSA
Clinic PSA
Modeled PSA
Days
0
200
400
600
800
1000
1200
1400
1600
0
0.2
0.4
0.6
0.8
1
Modeled
Population Densities
Sensitive
Resistant
0
2000
4000
6000
8000
10000

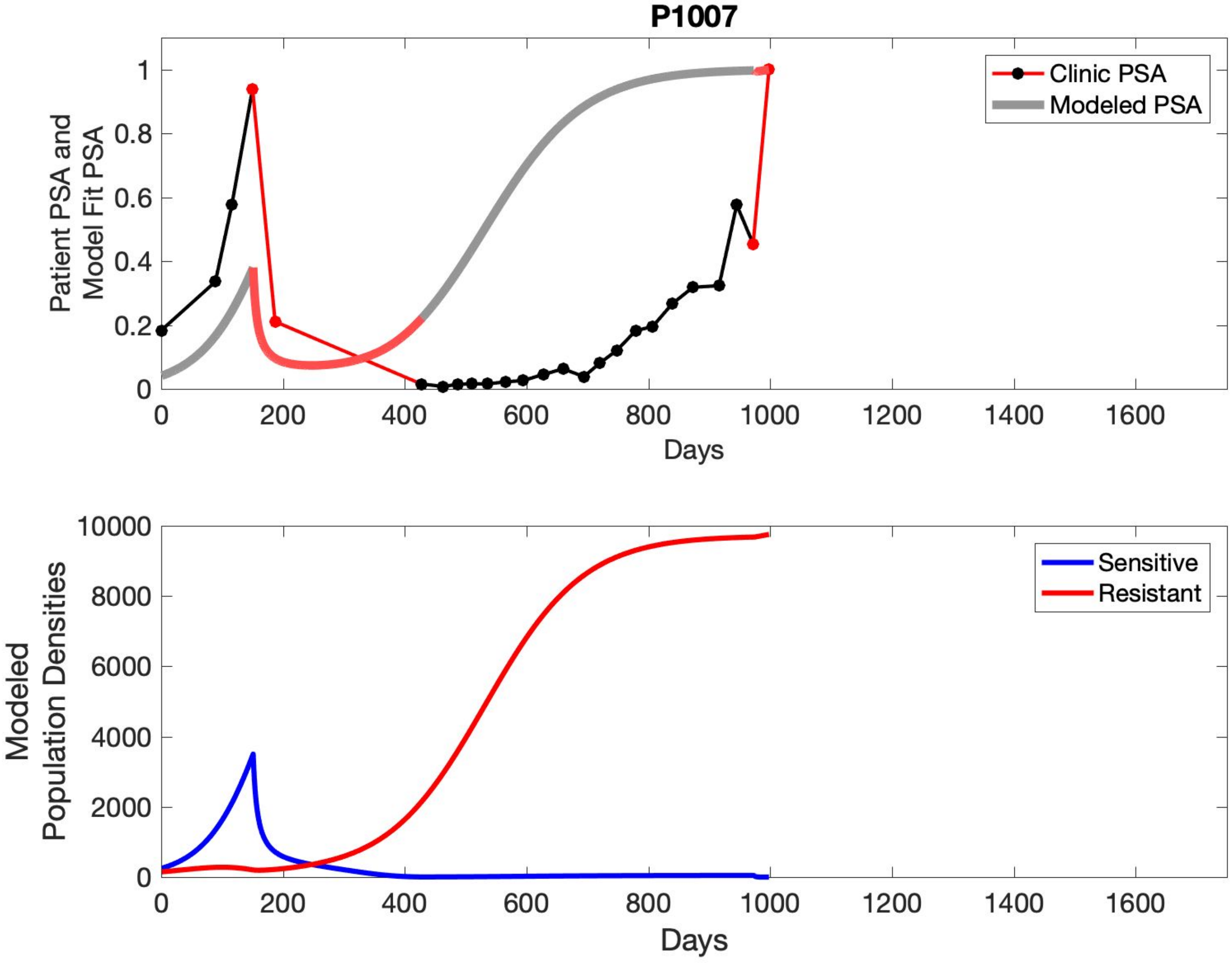
P1007
Patient PSA and Model Fit PSA
Days
Clinic PSA
Modeled PSA
0
0.2
0.4
0.6
0.8
1
200
400
600
800
1000
1200
1400
1600
Modeled Population Densities
Sensitive
Resistant
2000
4000
6000
8000
10000

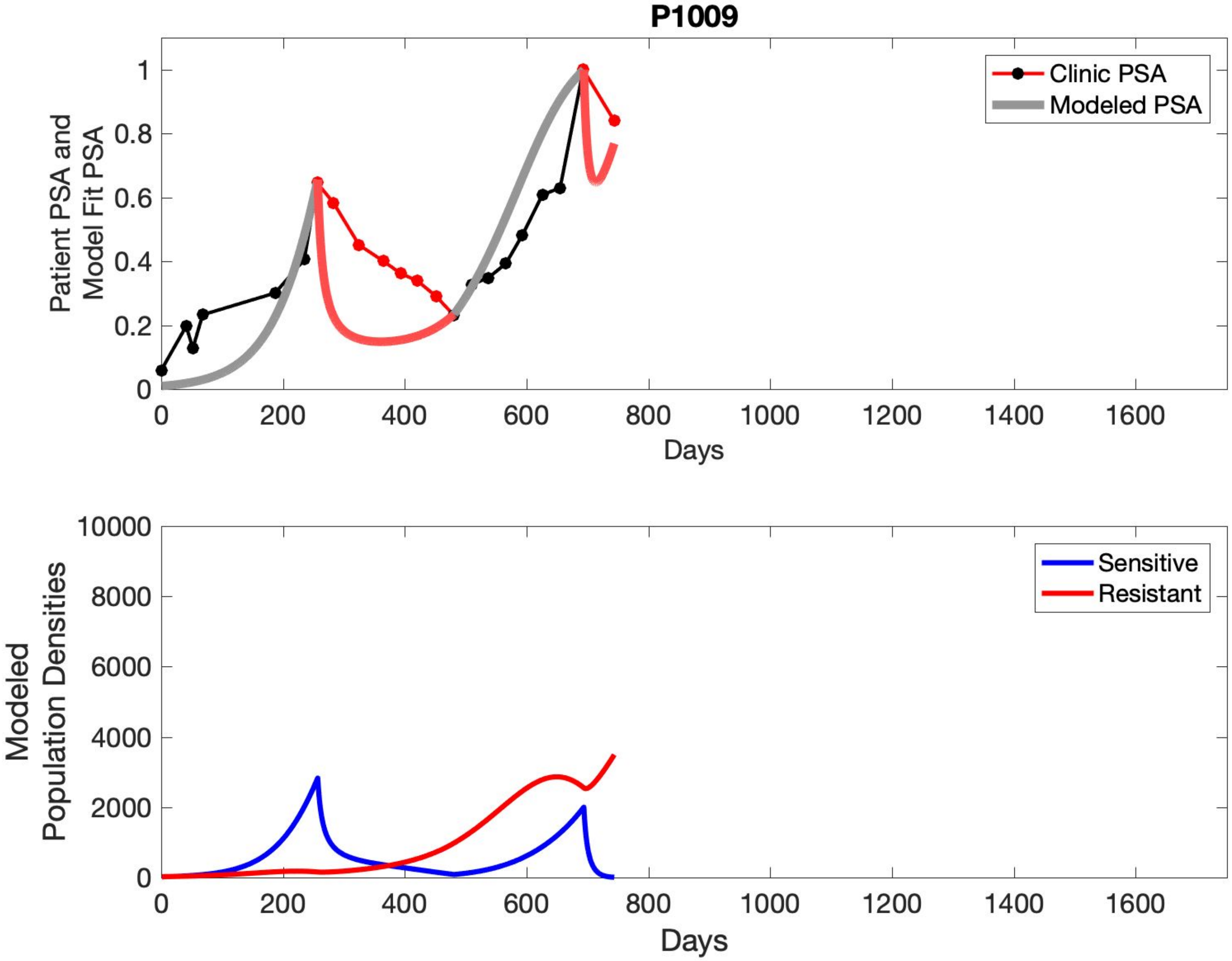

P1009
Patient PSA and Model Fit PSA
Days
Clinic PSA
Modeled PSA
0
0.2
0.4
0.6
0.8
1
200
400
600
800
1000
1200
1400
1600
Modeled Population Densities
Sensitive
Resistant
2000
4000
6000
8000
10000

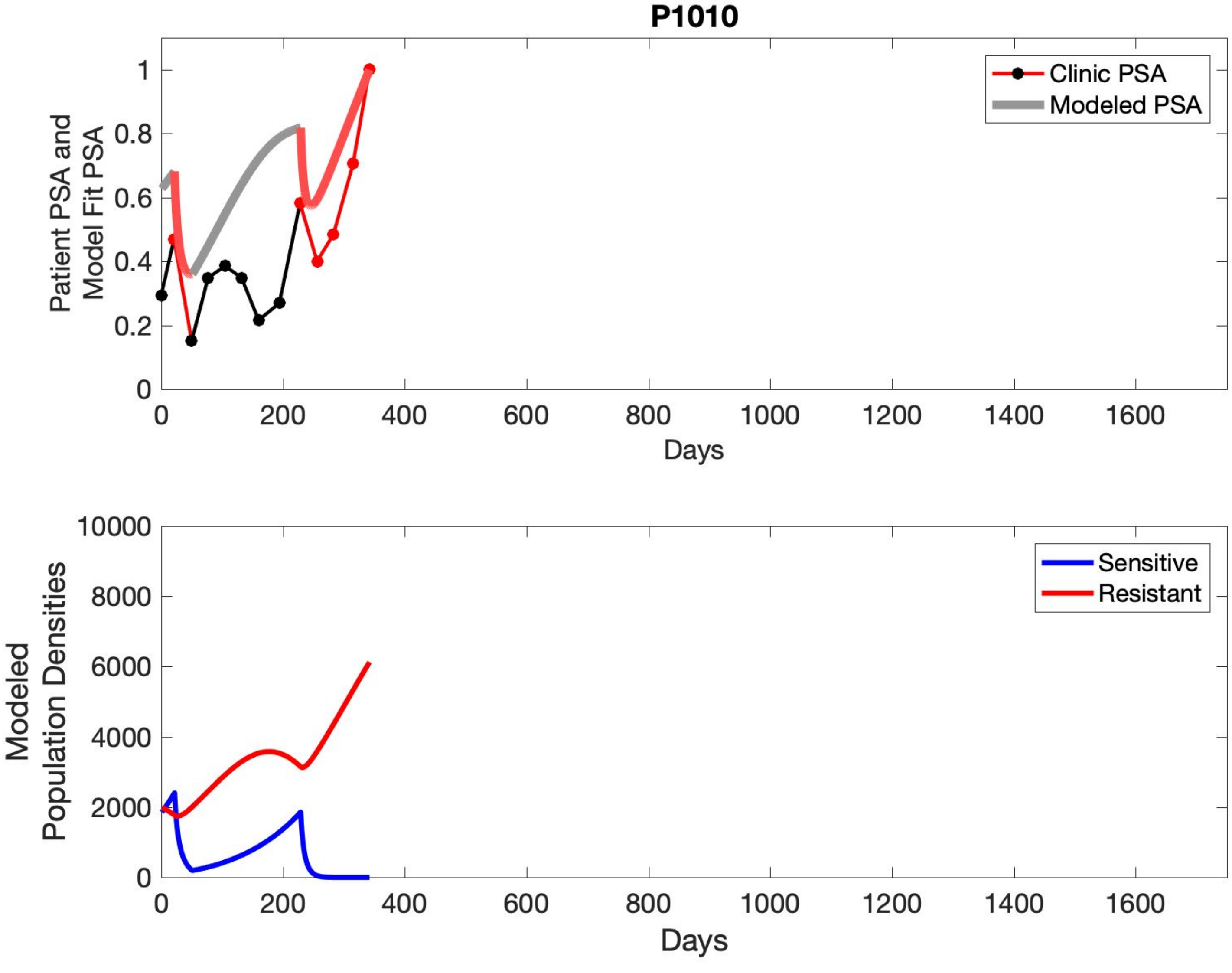

P1010
Patient PSA and Model Fit PSA
Clinic PSA
Modeled PSA
Days
Modeled Population Densities
Sensitive
Resistant
Days

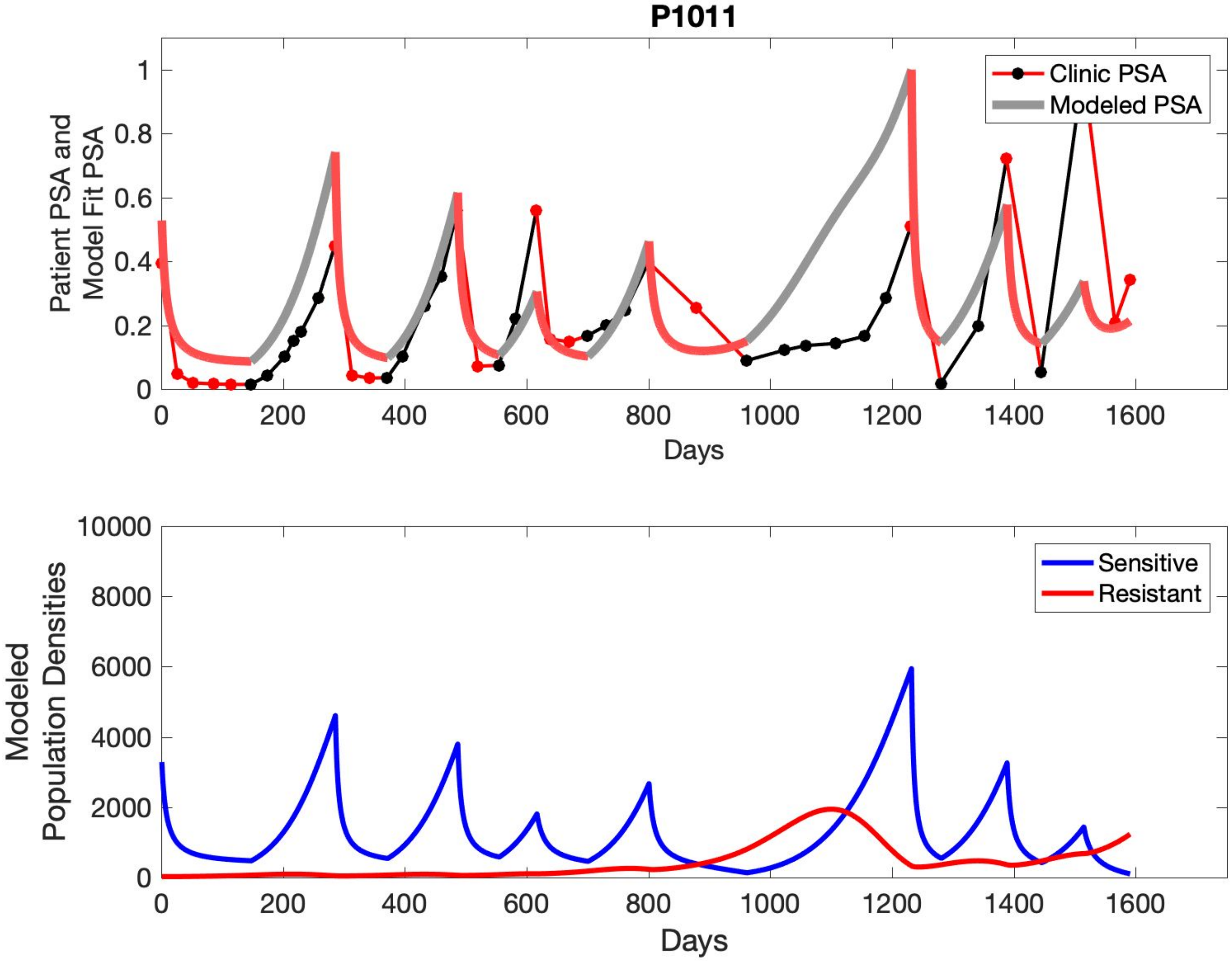

P1011
Clinic PSA
Modeled PSA
Patient PSA and Model Fit PSA
Days
Sensitive
Resistant
Modeled Population Densities
Days

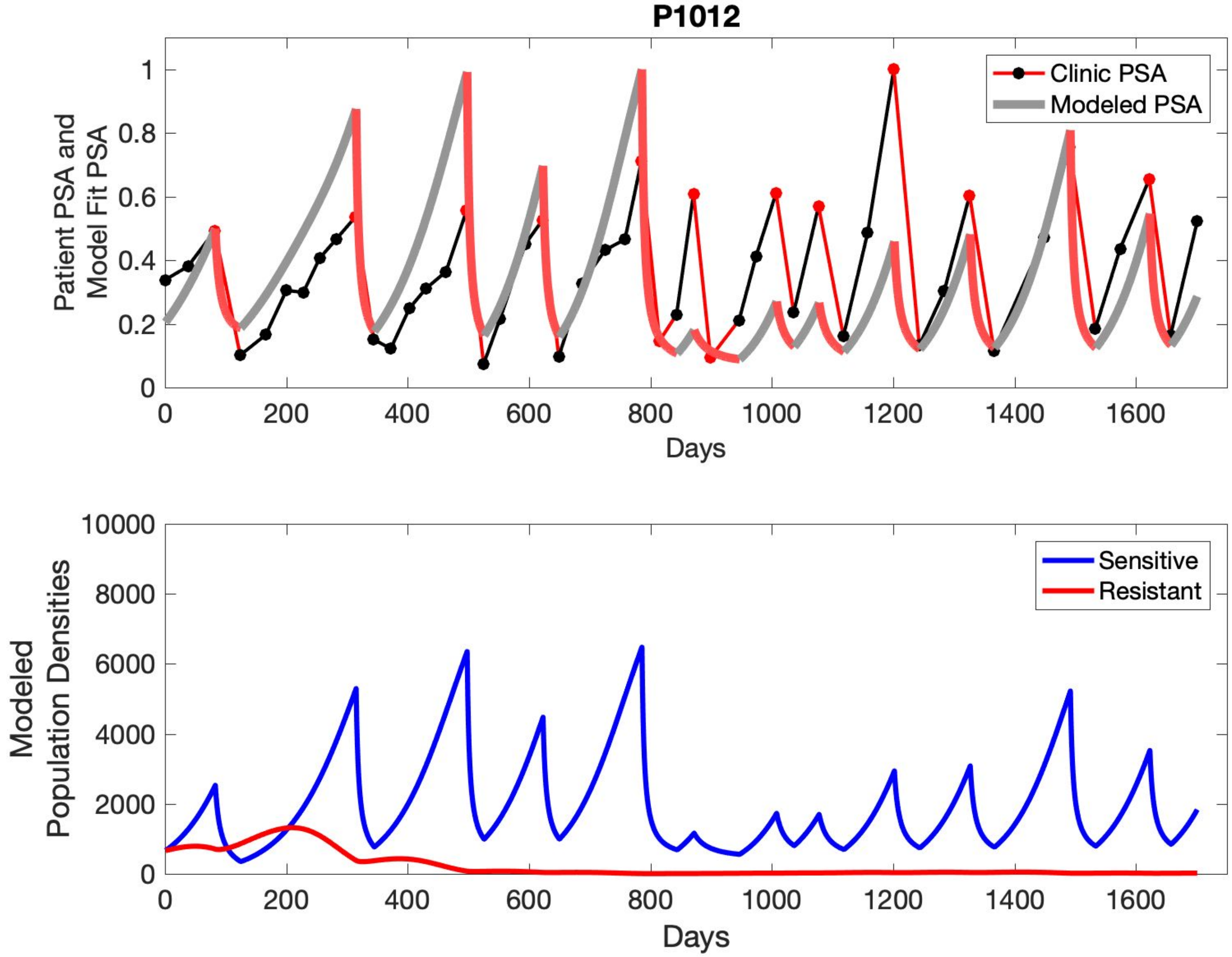

P1012
Patient PSA and Model Fit PSA
Clinic PSA
Modeled PSA
0
0.2
0.4
0.6
0.8
1
0
200
400
600
800
1000
1200
1400
1600
Days
Modeled Population Densities
Sensitive
Resistant
0
2000
4000
6000
8000
10000
0
200
400
600
800
1000
1200
1400
1600
Days

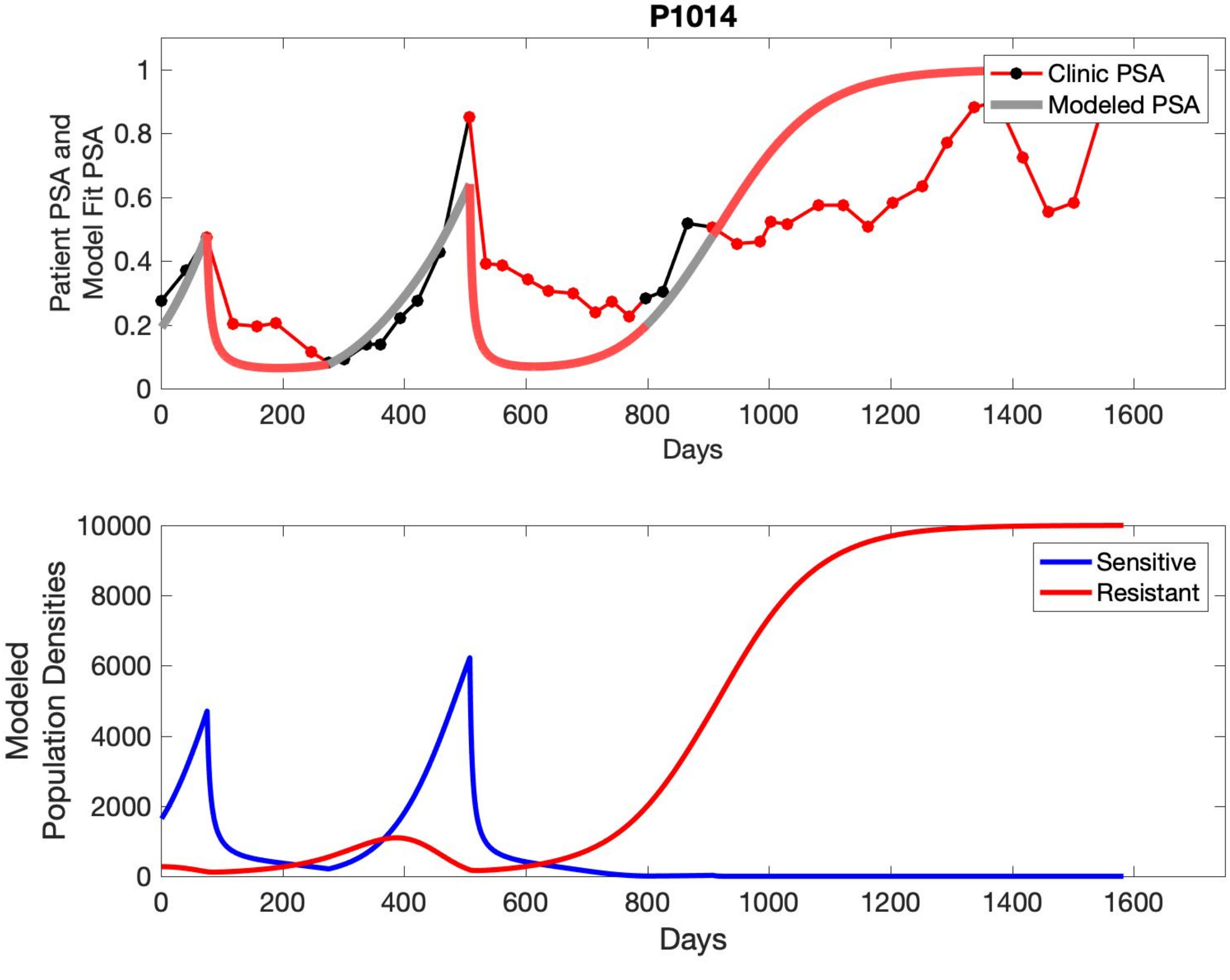

P1014
Patient PSA and Model Fit PSA
Clinic PSA
Modeled PSA
Days
Modeled Population Densities
Sensitive
Resistant
Days

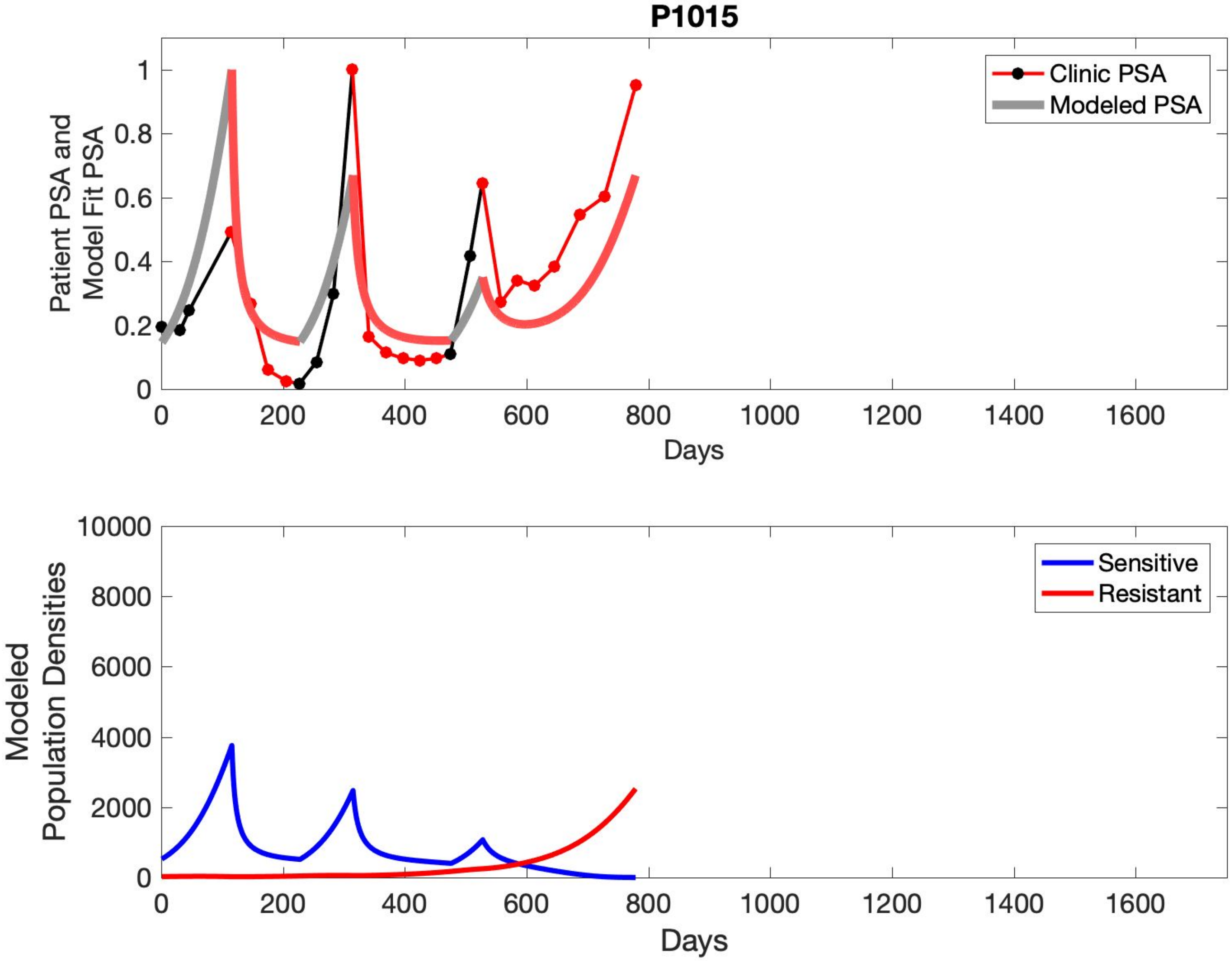
P1015
Clinic PSA
Modeled PSA
Patient PSA and Model Fit PSA
Days
Sensitive
Resistant
Modeled Population Densities
Days

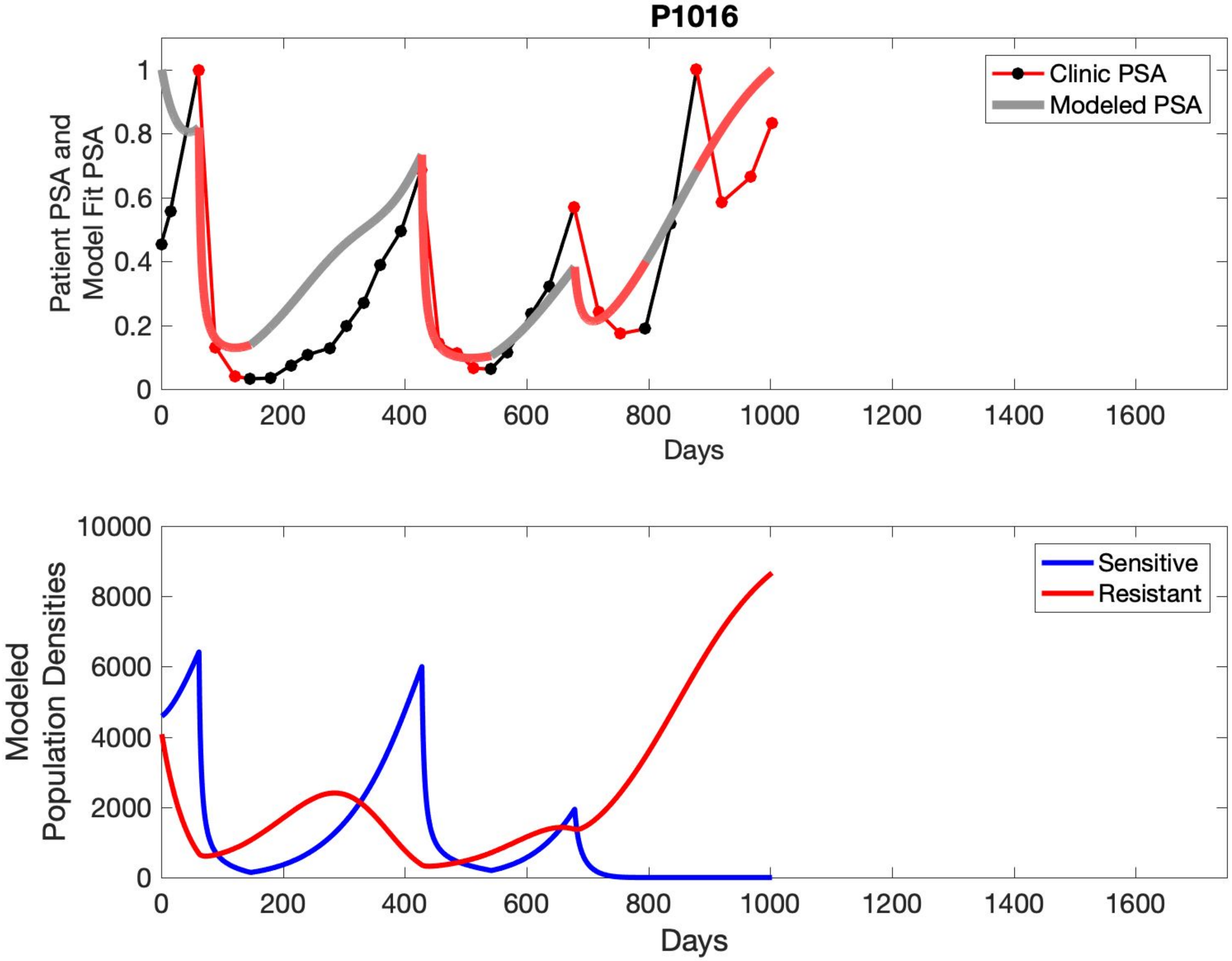
P1016
Patient PSA and
Model Fit PSA
Clinic PSA
Modeled PSA
Days
Modeled
Population Densities
Sensitive
Resistant
Days

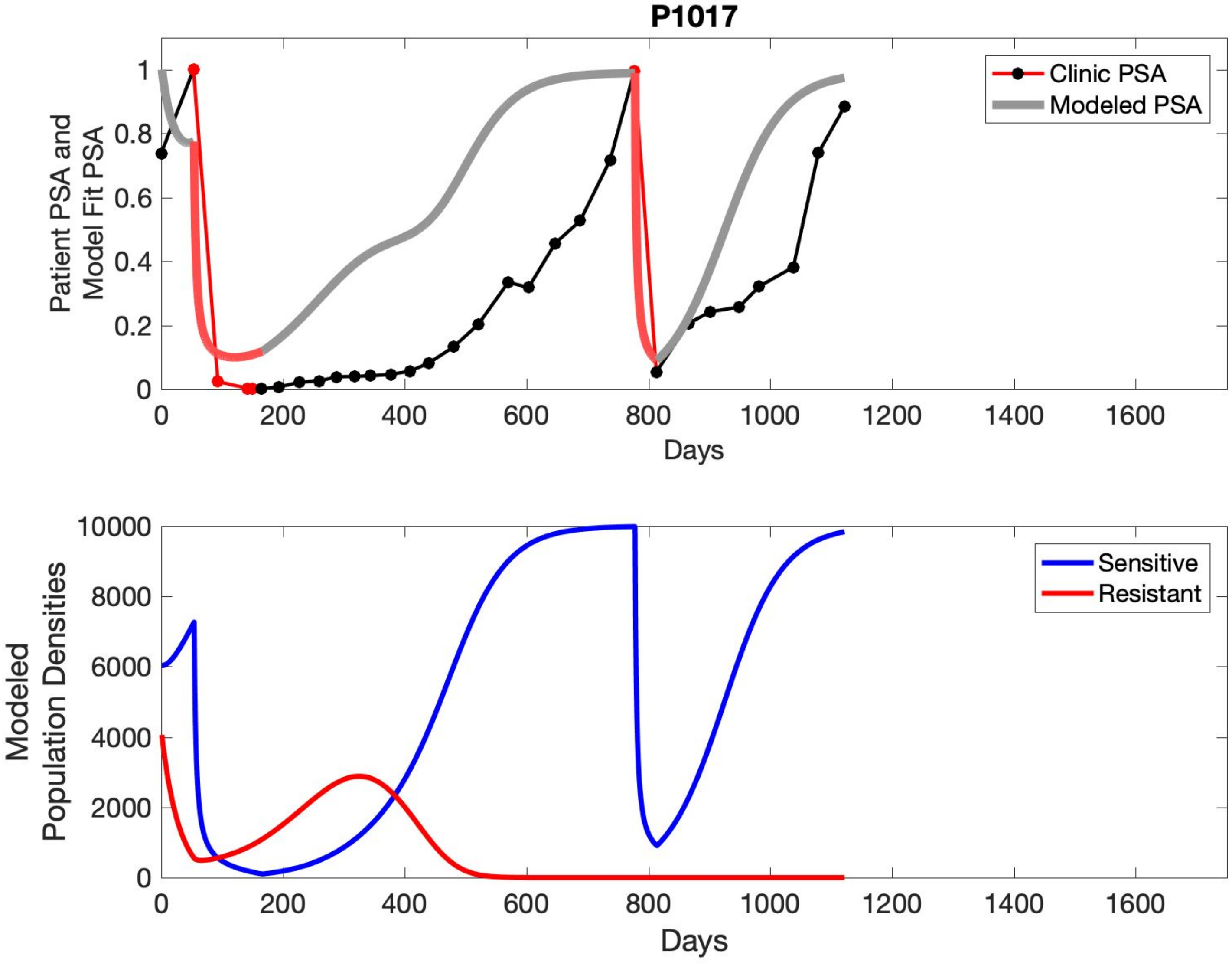
P1017
Clinic PSA
Modeled PSA
Patient PSA and Model Fit PSA
Days
Sensitive
Resistant
Modeled Population Densities
Days

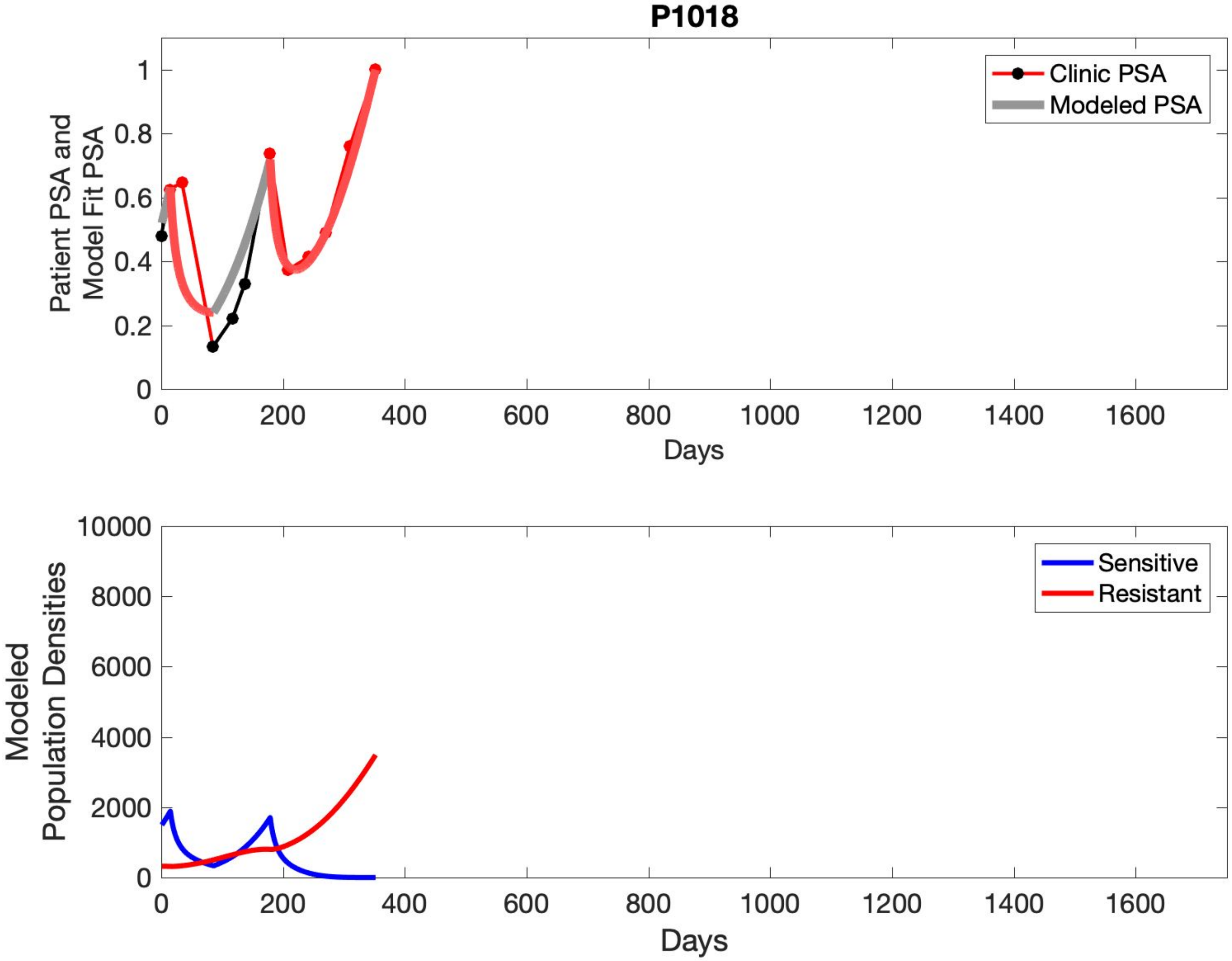
P1018
Patient PSA and Model Fit PSA
Clinic PSA
Modeled PSA
Days
Modeled Population Densities
Sensitive
Resistant
Days

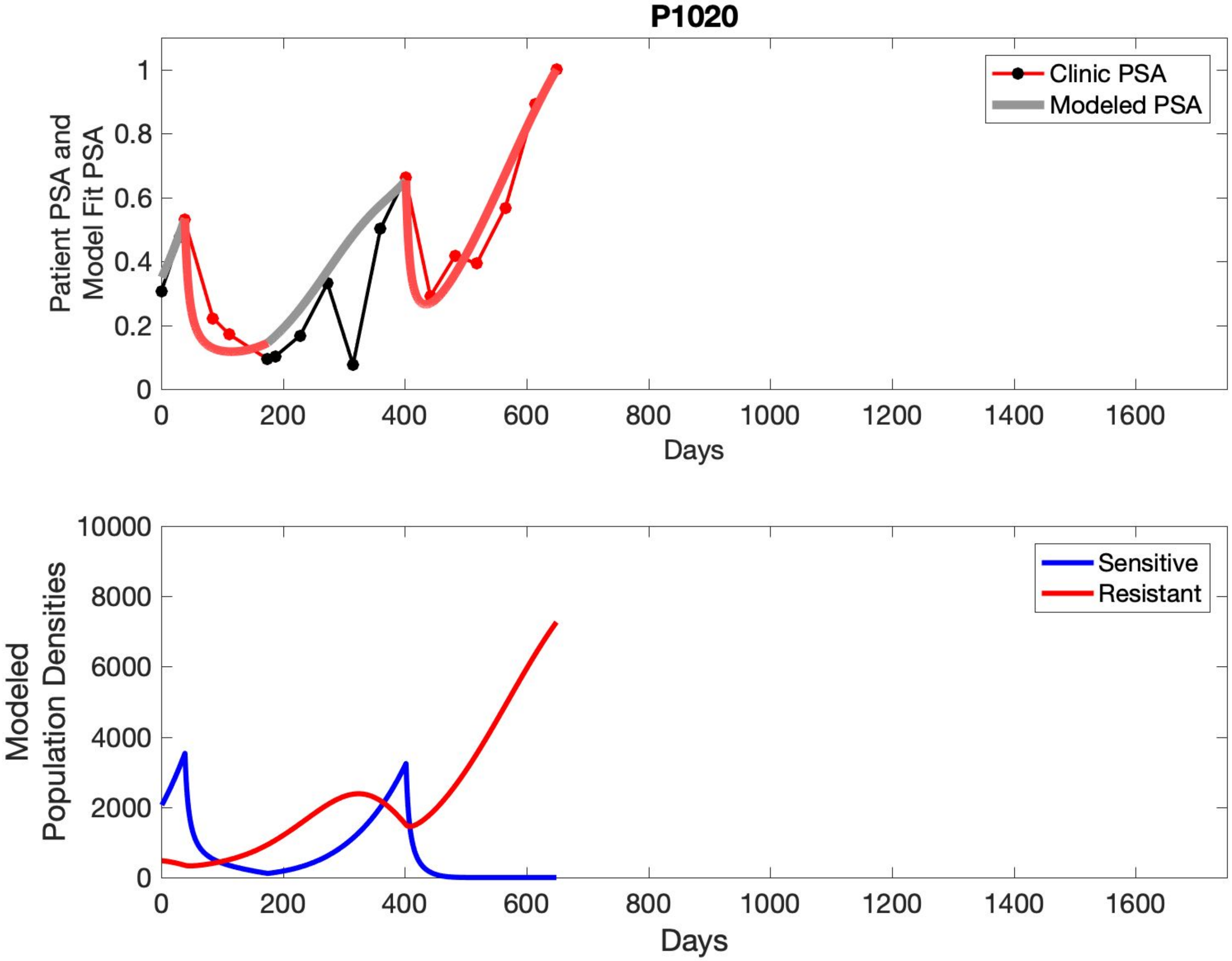

P1020
Clinic PSA
Modeled PSA
Patient PSA and Model Fit PSA
Days
Sensitive
Resistant
Modeled Population Densities
Days

#### S5 Simulated Idealized Adaptive Therapy

To evaluate an ideal 50% adaptive therapy protocol, we ran a simulation for each patient (both cohorts). In line with the trial, abiraterone was administered until there was a 50% drop in the patient's PSA at which point therapy ceased until PSA levels returned to the patient's initial value at which point therapy was resumed, and so on. If the PSA ceased to decline to 50% of the initial PSA, than abiraterone was continued indefinitely. For patients in the contemporaneous cohort we can predict how they might have fared under adaptive therapy. For the patients that clinically received adaptive therapy, we can see how they would have fared if the therapy had been idealized to start and stop at the exact switch points. In reality, the start and stop to abiraterone often occurred at PSA levels higher than the initial level and lower than the 50% threshold, respectively. This treatment protocol was run for each patient using their patient-specific parameters from Table S4.

**C001**

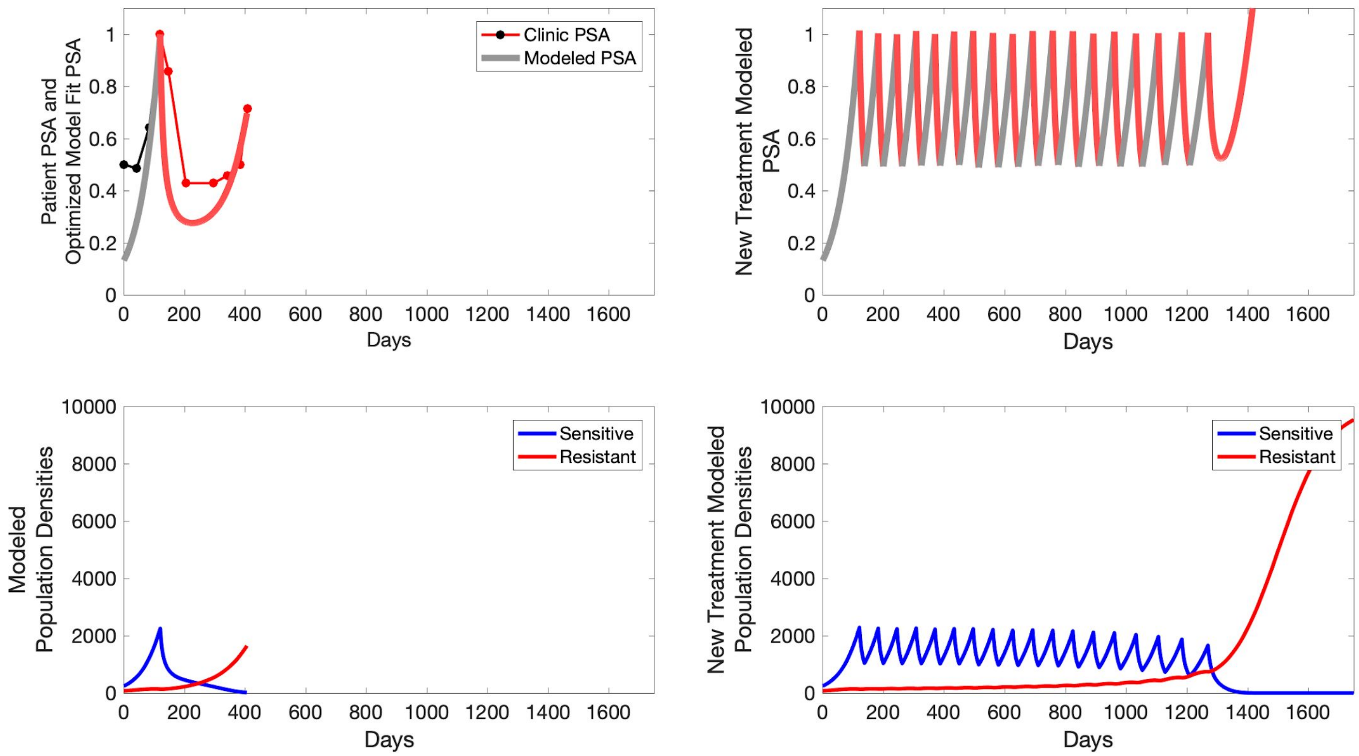

Patient PSA and Optimized Model Fit PSA
Clinic PSA
Modeled PSA
Days
New Treatment Modeled PSA
Modeled Population Densities
Sensitive
Resistant
New Treatment Modeled Population Densities

**C002**

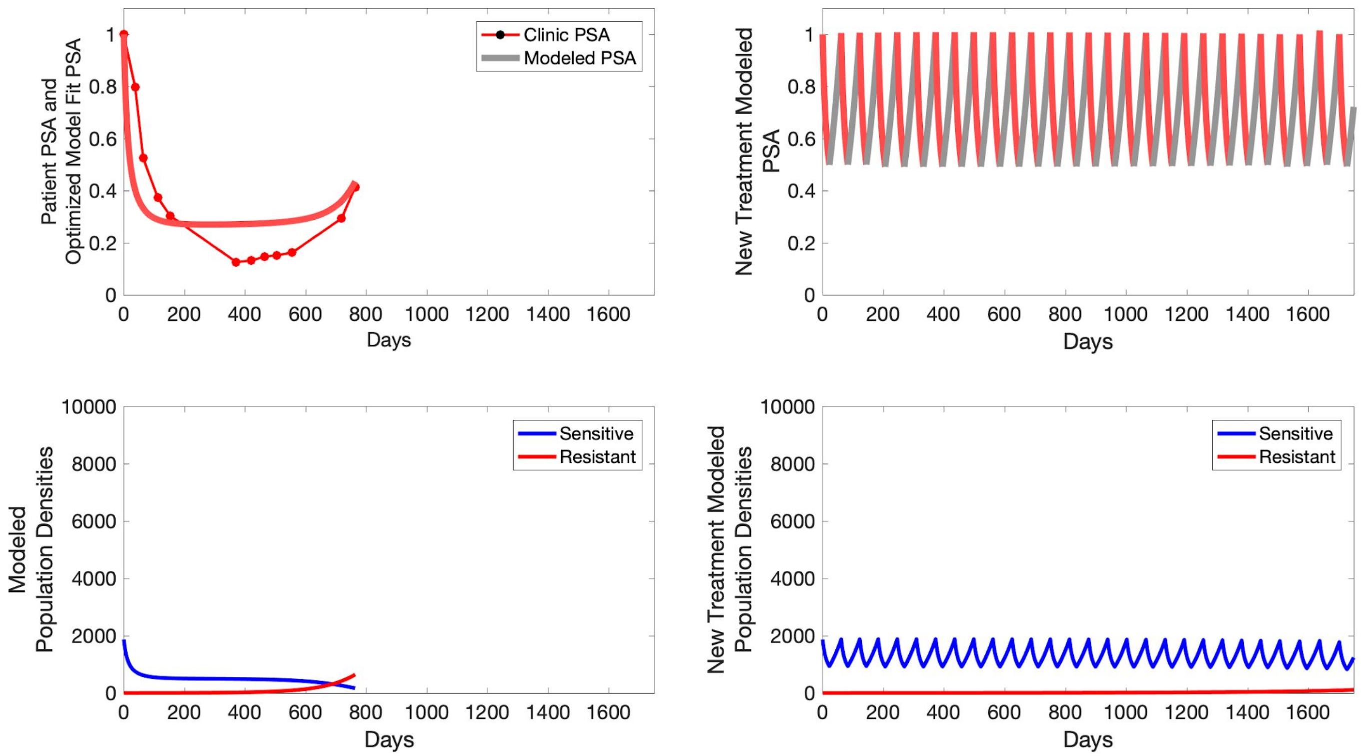

Clinic PSA
Modeled PSA
Patient PSA and Optimized Model Fit PSA
Days
New Treatment Modeled PSA
Sensitive
Resistant
Modeled Population Densities
New Treatment Modeled Population Densities

**C003**

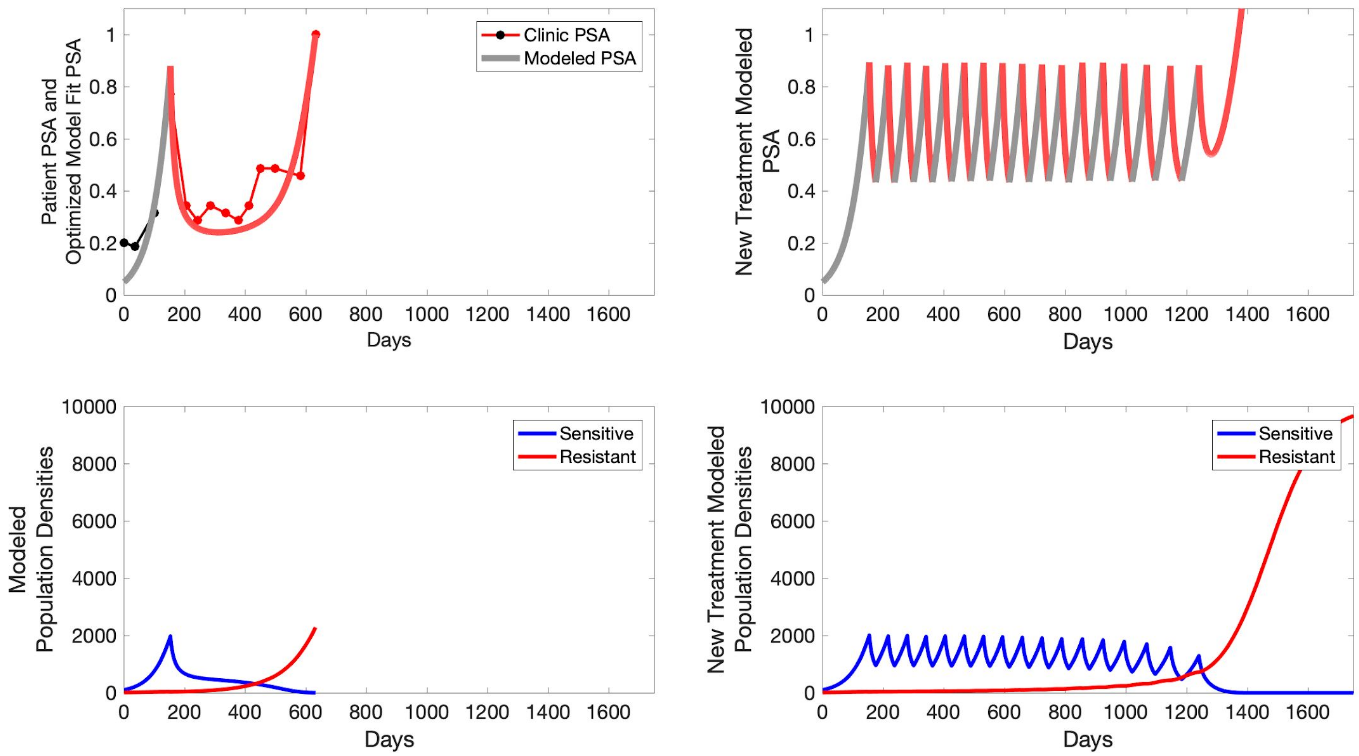

Patient PSA and Optimized Model Fit PSA
Clinic PSA
Modeled PSA
Days
New Treatment Modeled PSA
Days
Modeled Population Densities
Sensitive
Resistant
Days
New Treatment Modeled Population Densities
Sensitive
Resistant
Days

**C004**

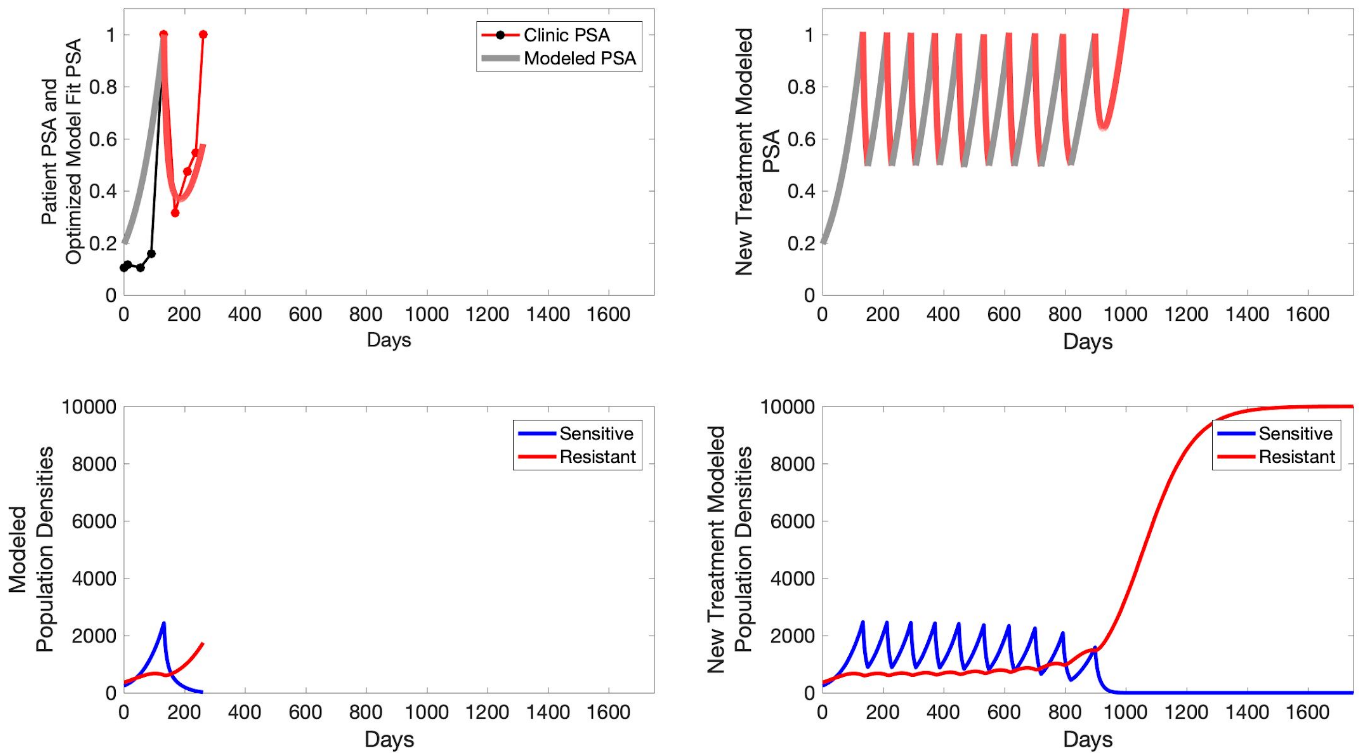

Patient PSA and Optimized Model Fit PSA
Clinic PSA
Modeled PSA
Days
New Treatment Modeled PSA
Days
Modeled Population Densities
Sensitive
Resistant
Days
New Treatment Modeled Population Densities
Sensitive
Resistant
Days

**C005**

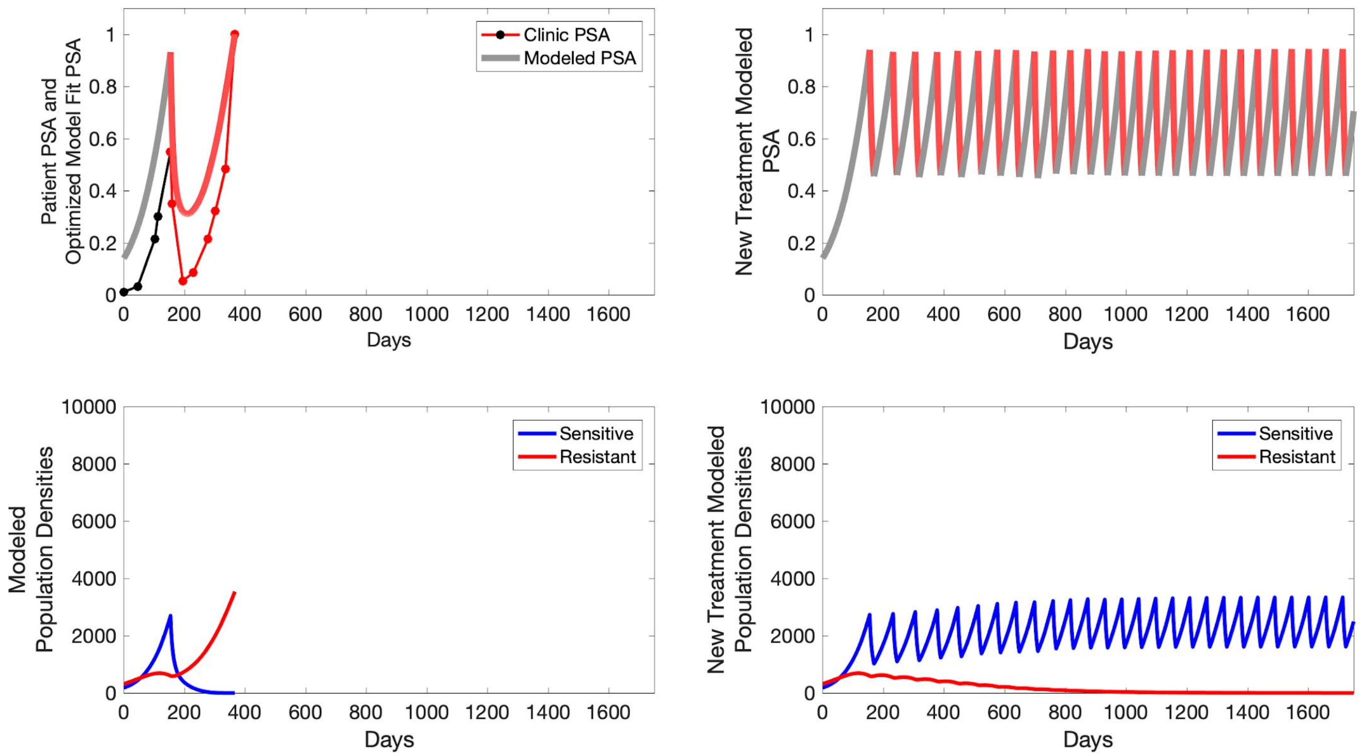

Clinic PSA
Modeled PSA
Patient PSA and Optimized Model Fit PSA
New Treatment Modeled PSA
Sensitive
Resistant
Modeled Population Densities
New Treatment Modeled Population Densities
Days

**C006**

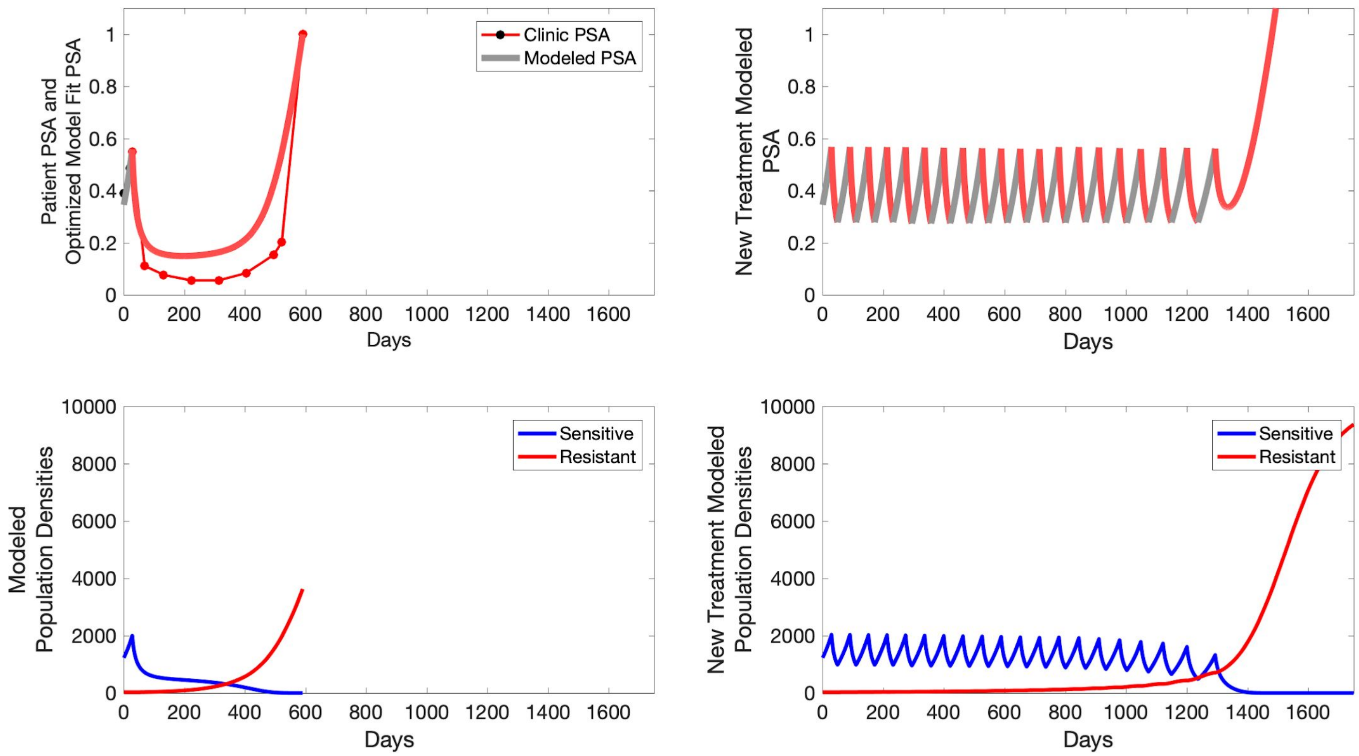

Patient PSA and Optimized Model Fit PSA
Clinic PSA
Modeled PSA
Days
New Treatment Modeled PSA
Days
Modeled Population Densities
Sensitive
Resistant
Days
New Treatment Modeled Population Densities
Sensitive
Resistant
Days

**C007**

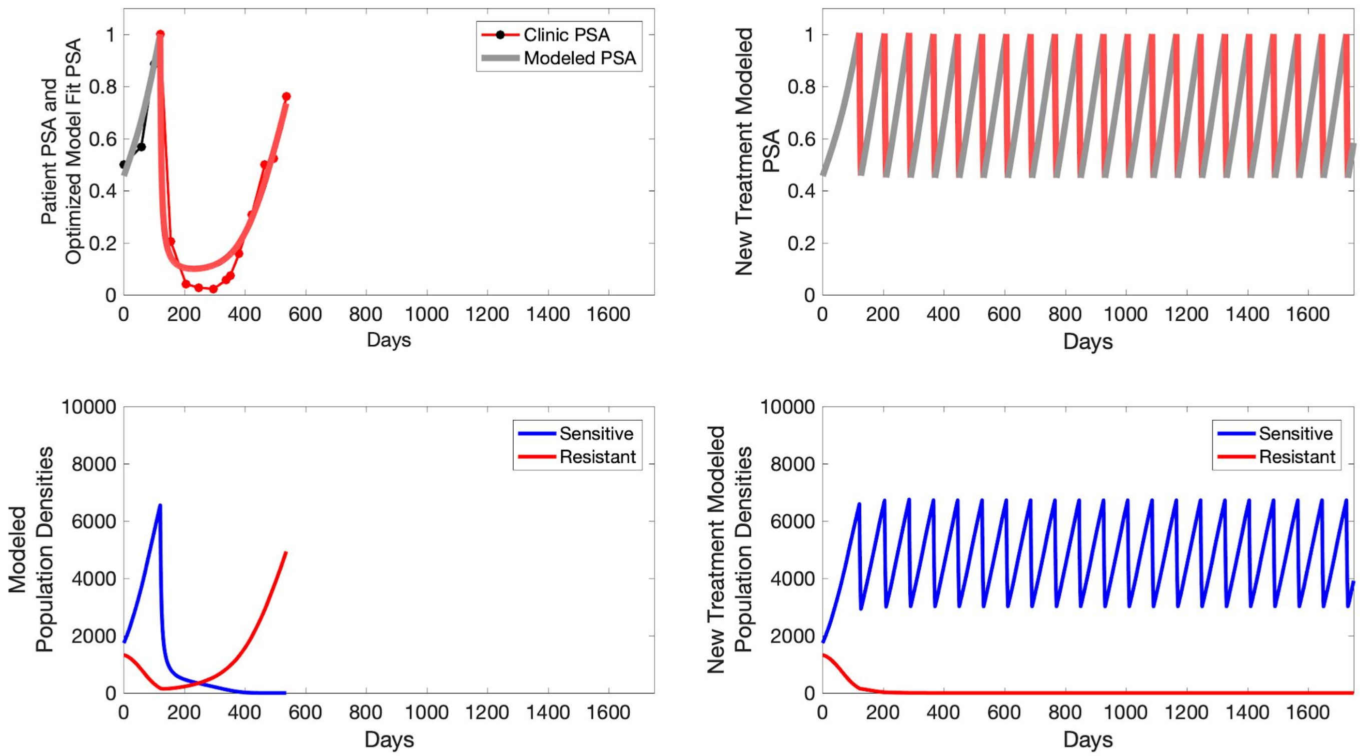

Clinic PSA
Modeled PSA
Patient PSA and Optimized Model Fit PSA
New Treatment Modeled PSA
Days
Sensitive
Resistant
Modeled Population Densities
New Treatment Modeled Population Densities

**C008**

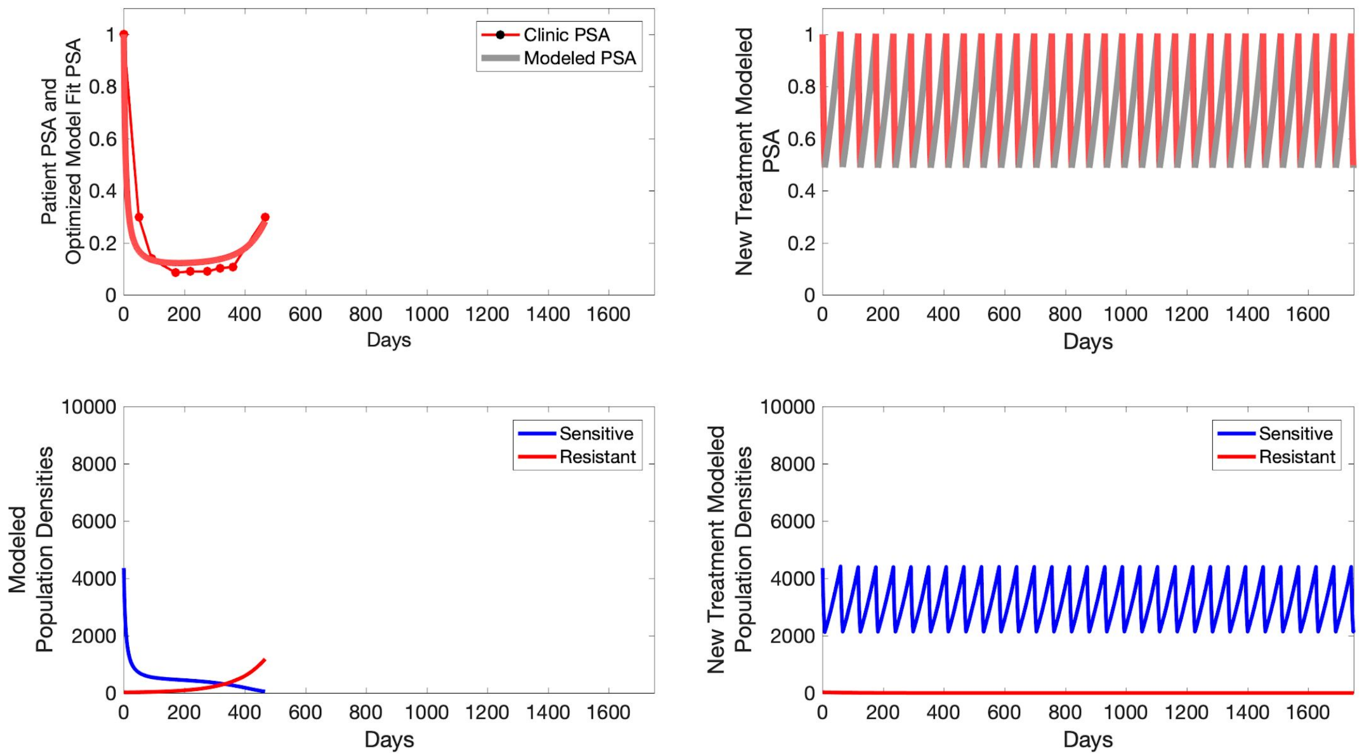
Patient PSA and Optimized Model Fit PSA
Clinic PSA
Modeled PSA
Days
New Treatment Modeled PSA
Days
Modeled Population Densities
Sensitive
Resistant
Days
New Treatment Modeled Population Densities
Sensitive
Resistant
Days

**C009**

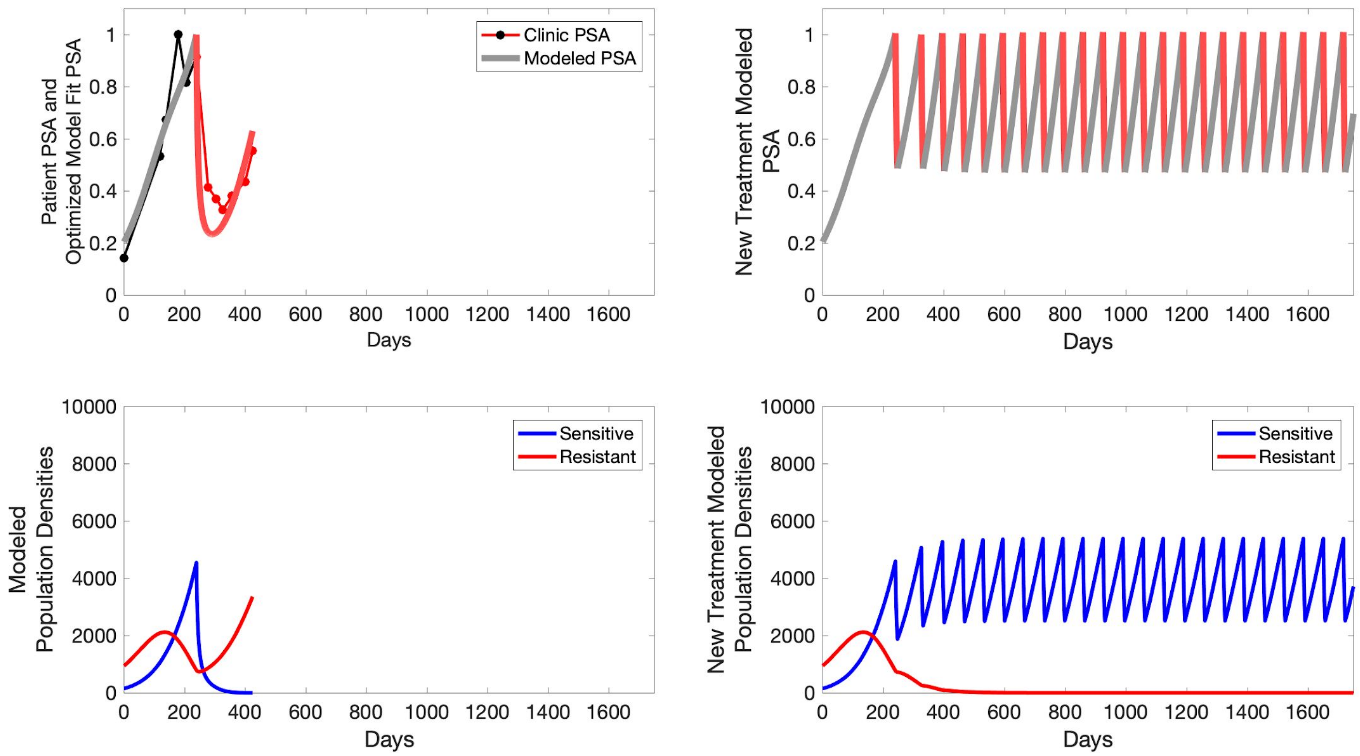

Patient PSA and Optimized Model Fit PSA
Clinic PSA
Modeled PSA
Days
New Treatment Modeled PSA
Days
Modeled Population Densities
Sensitive
Resistant
Days
New Treatment Modeled Population Densities
Sensitive
Resistant
Days

**C010**

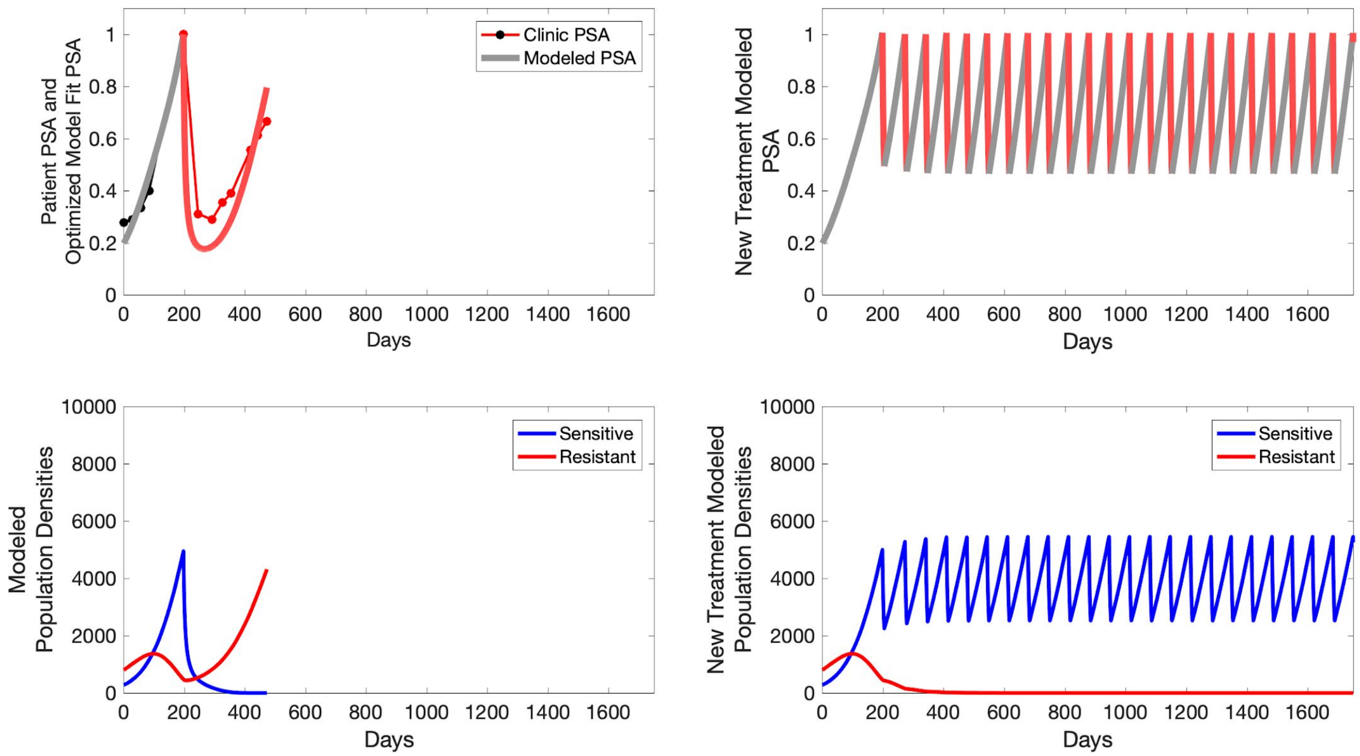

Patient PSA and
Optimized Model Fit PSA
Clinic PSA
Modeled PSA
Days
New Treatment Modeled
PSA
Days
Modeled
Population Densities
Sensitive
Resistant
Days
New Treatment Modeled
Population Densities
Sensitive
Resistant
Days

**C011**

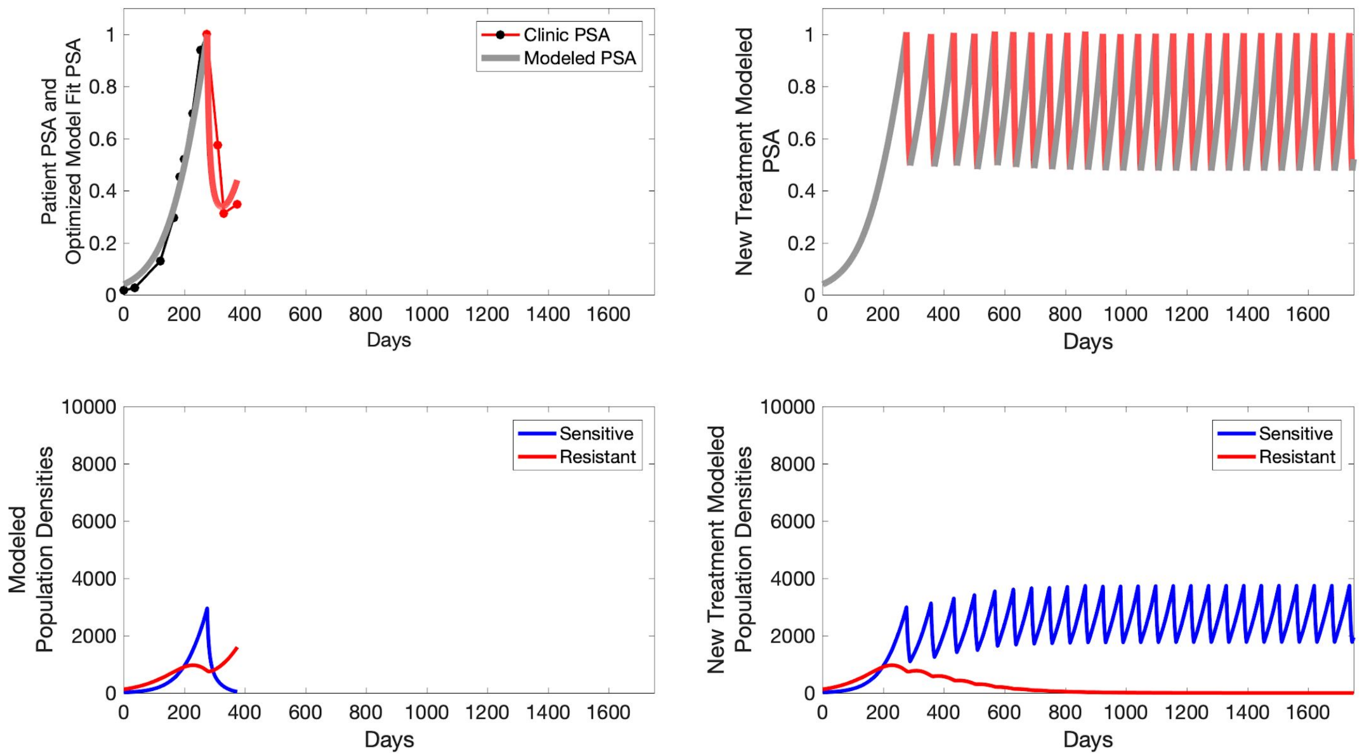

Clinic PSA
Modeled PSA
Patient PSA and
Optimized Model Fit PSA
Days
New Treatment Modeled
PSA
Days
Sensitive
Resistant
Modeled
Population Densities
Days
Sensitive
Resistant
New Treatment Modeled
Population Densities
Days

**C012**

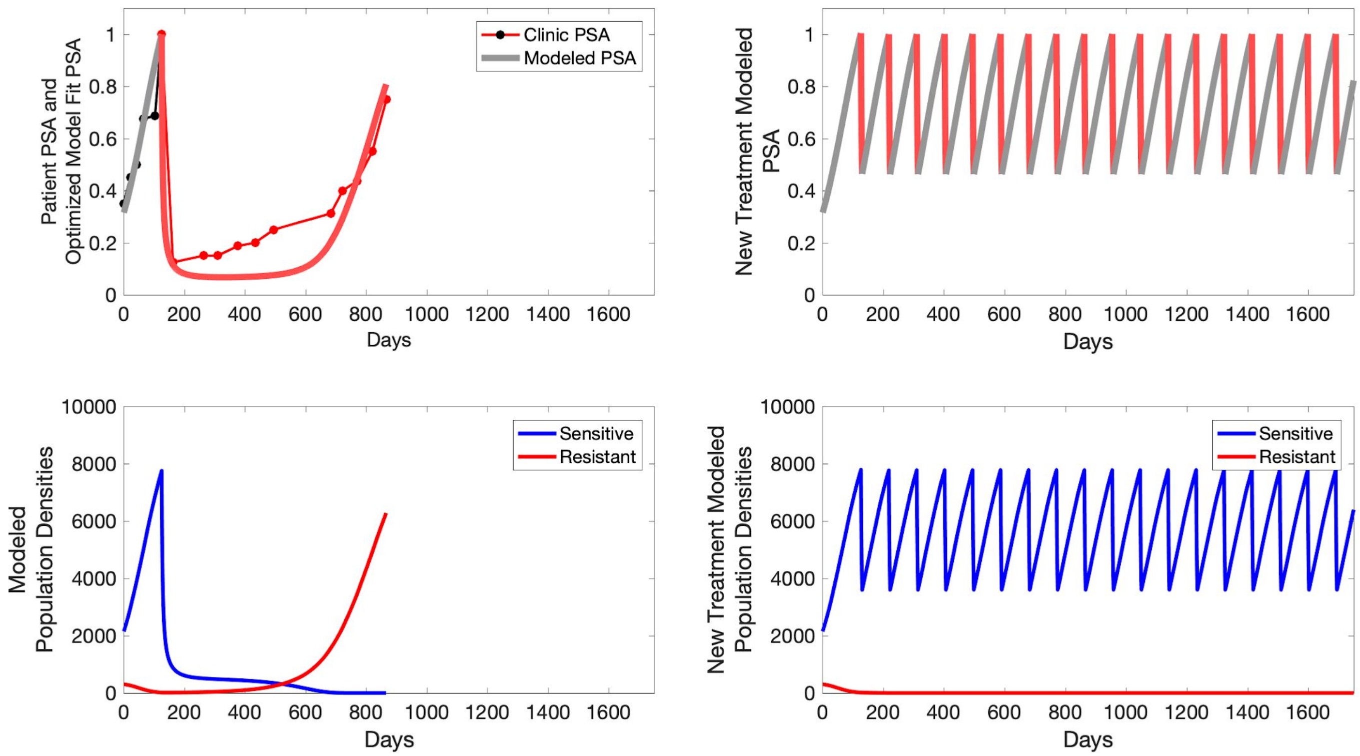

Clinic PSA
Modeled PSA
Patient PSA and
Optimized Model Fit PSA
Days
New Treatment Modeled
PSA
Days
Sensitive
Resistant
Modeled
Population Densities
Days
Sensitive
Resistant
New Treatment Modeled
Population Densities
Days

**C013**

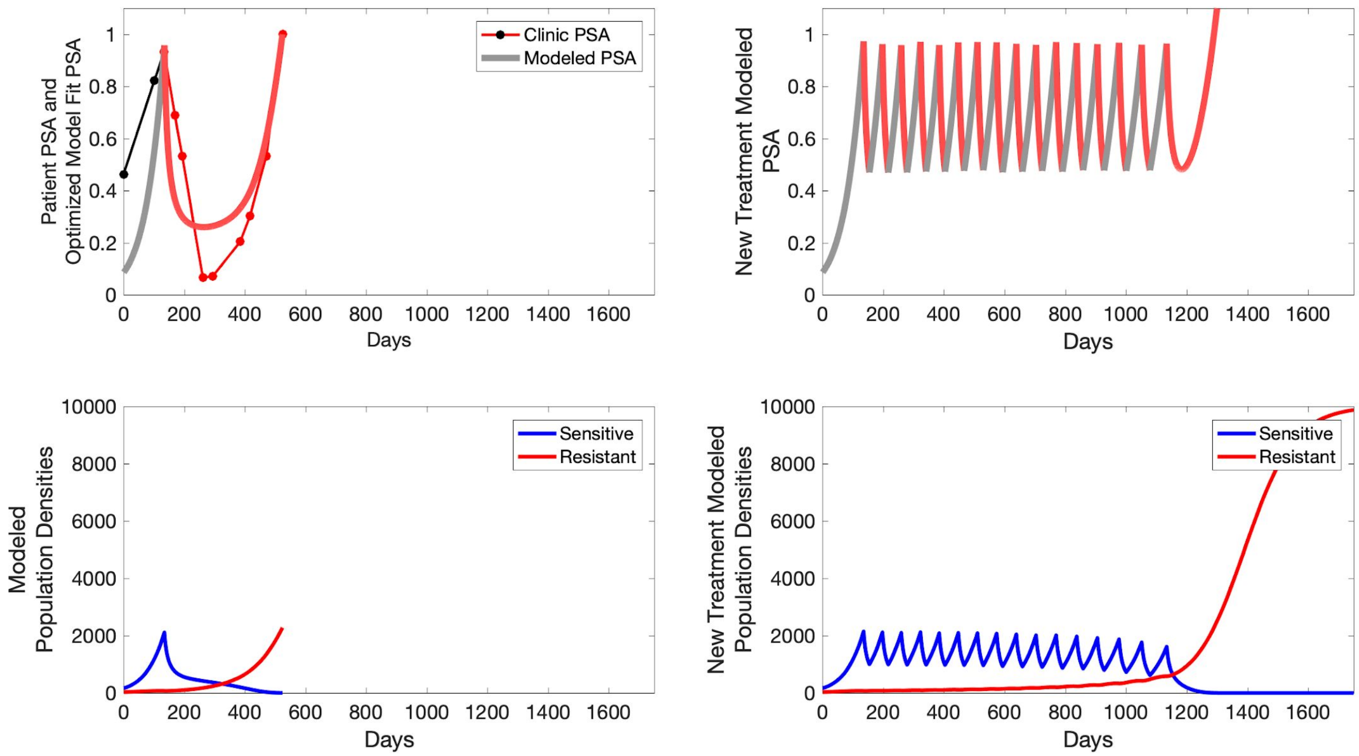
Clinic PSA
Modeled PSA
Patient PSA and
Optimized Model Fit PSA
Days
New Treatment Modeled
PSA
Days
Sensitive
Resistant
Modeled
Population Densities
Days
Sensitive
Resistant
New Treatment Modeled
Population Densities
Days

**C014**

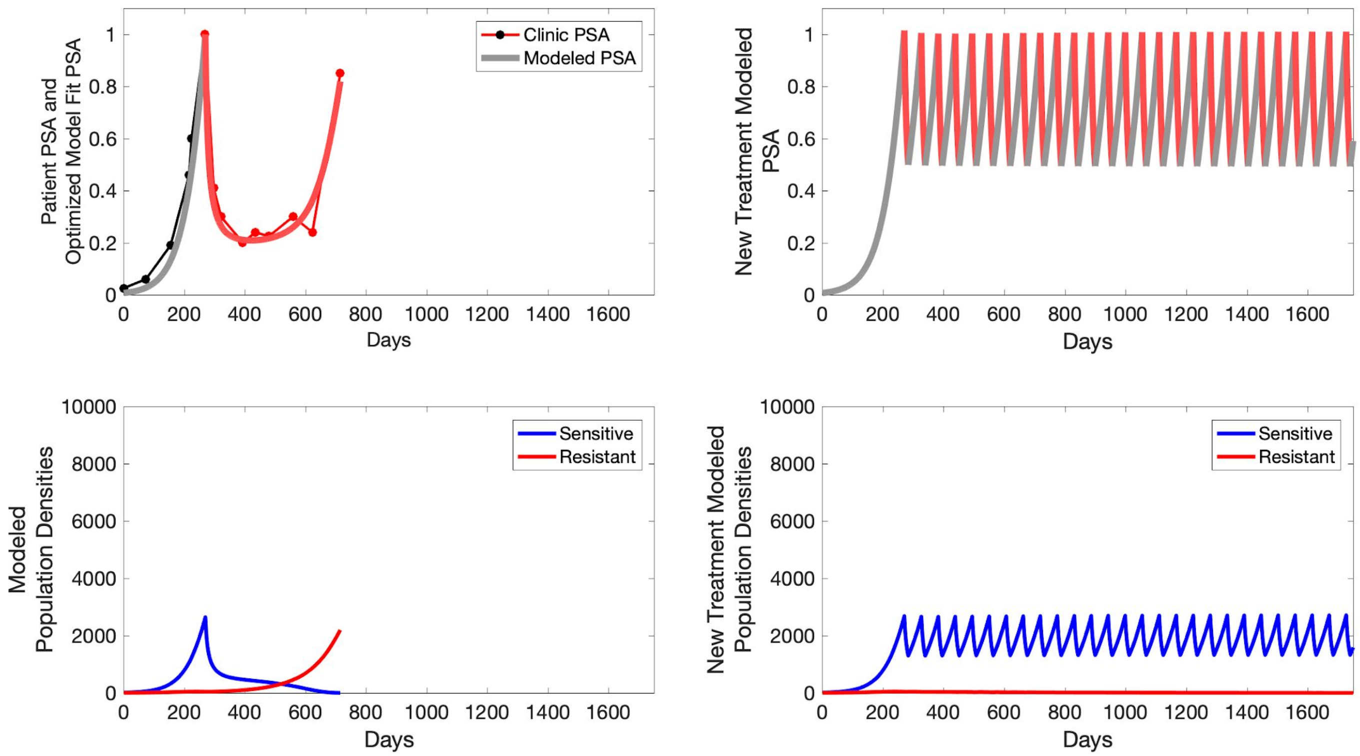

Patient PSA and Optimized Model Fit PSA
Clinic PSA
Modeled PSA
Days
New Treatment Modeled PSA
Days
Modeled Population Densities
Sensitive
Resistant
Days
New Treatment Modeled Population Densities
Sensitive
Resistant
Days

**C015**

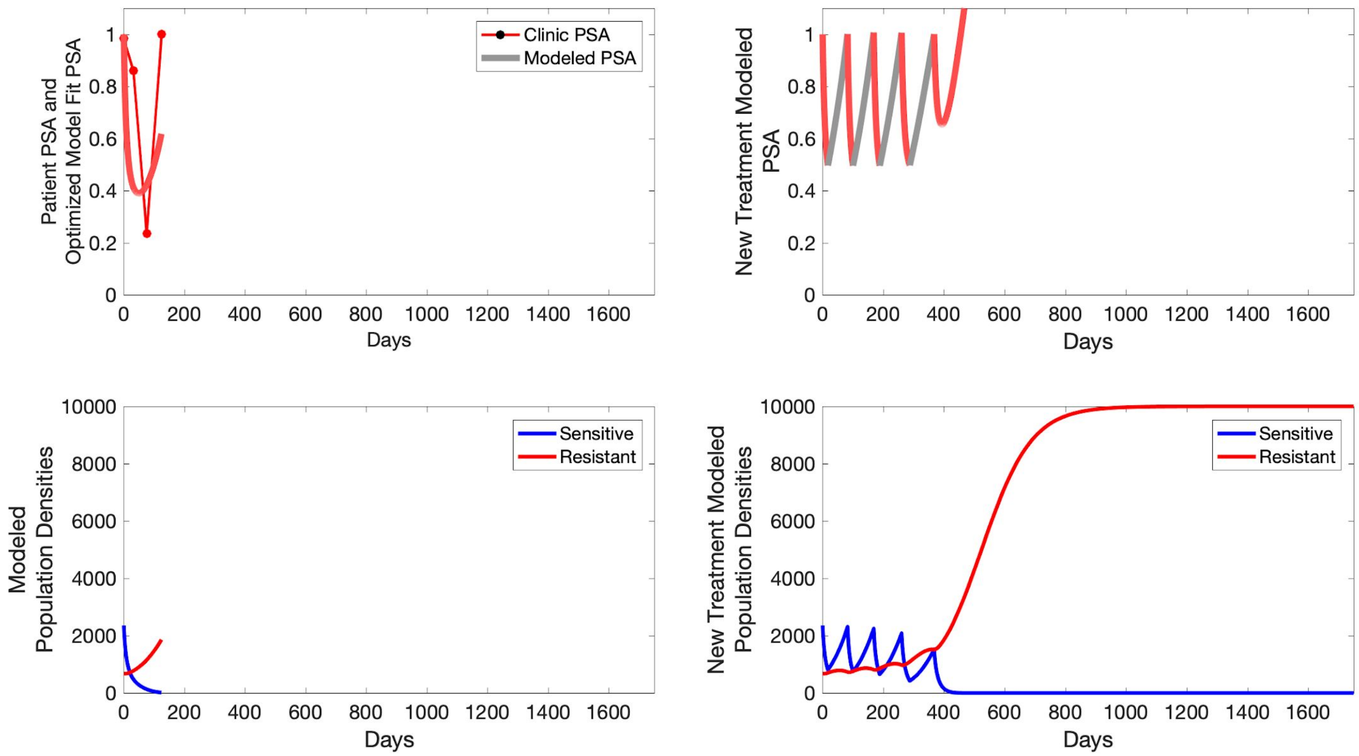

Clinic PSA
Modeled PSA
Patient PSA and Optimized Model Fit PSA
Days
New Treatment Modeled PSA
Days
Sensitive
Resistant
Modeled Population Densities
Days
Sensitive
Resistant
New Treatment Modeled Population Densities
Days

**P1001**

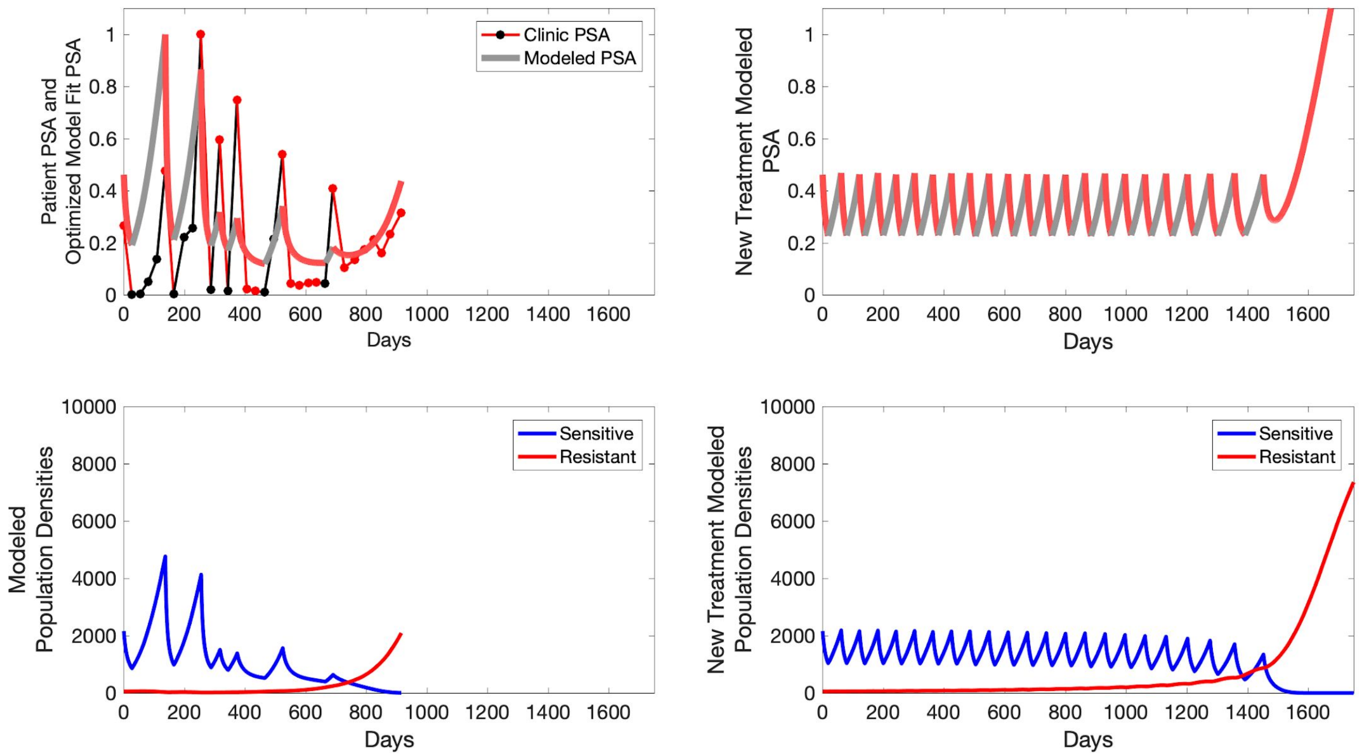

Patient PSA and Optimized Model Fit PSA
Clinic PSA
Modeled PSA
Days
New Treatment Modeled PSA
Days
Modeled Population Densities
Sensitive
Resistant
Days
New Treatment Modeled Population Densities
Sensitive
Resistant
Days

**P1002**

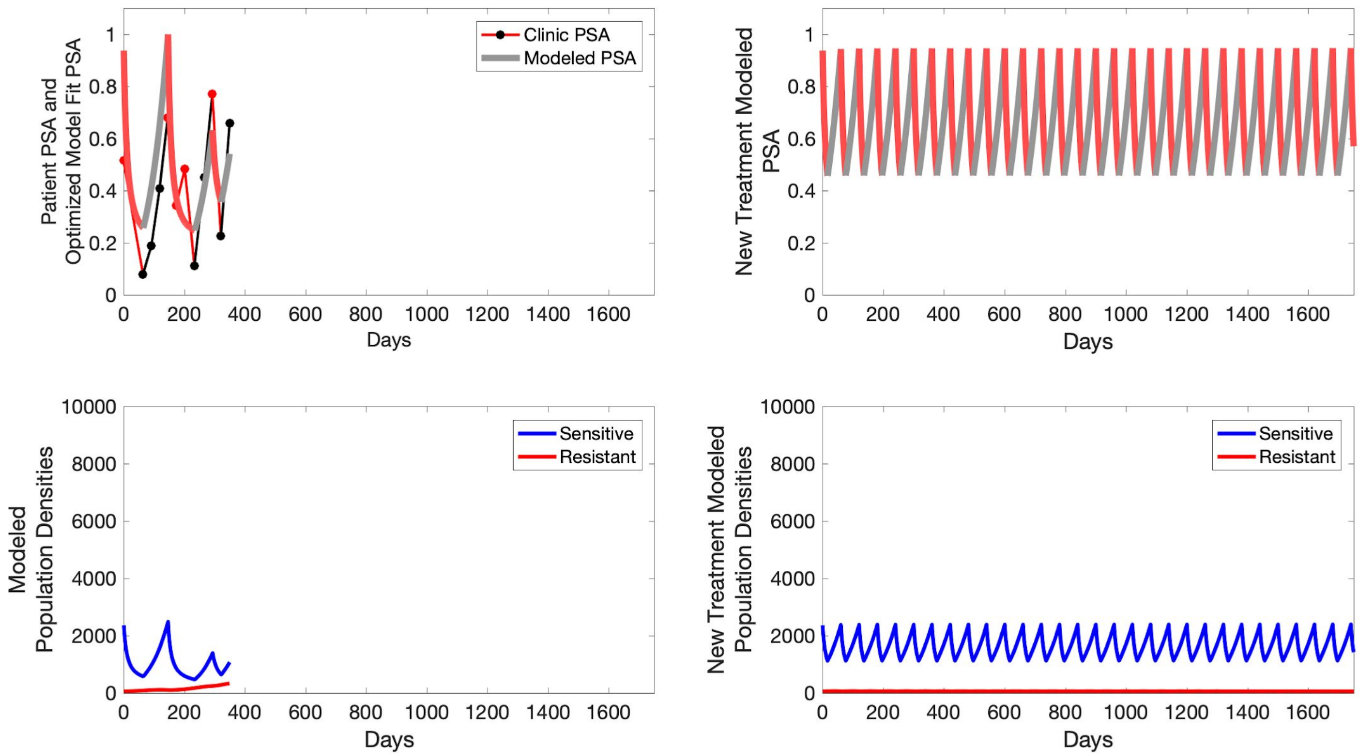

Clinic PSA
Modeled PSA
Patient PSA and
Optimized Model Fit PSA
Days
New Treatment Modeled
PSA
Sensitive
Resistant
Modeled
Population Densities
New Treatment Modeled
Population Densities

**P1003**

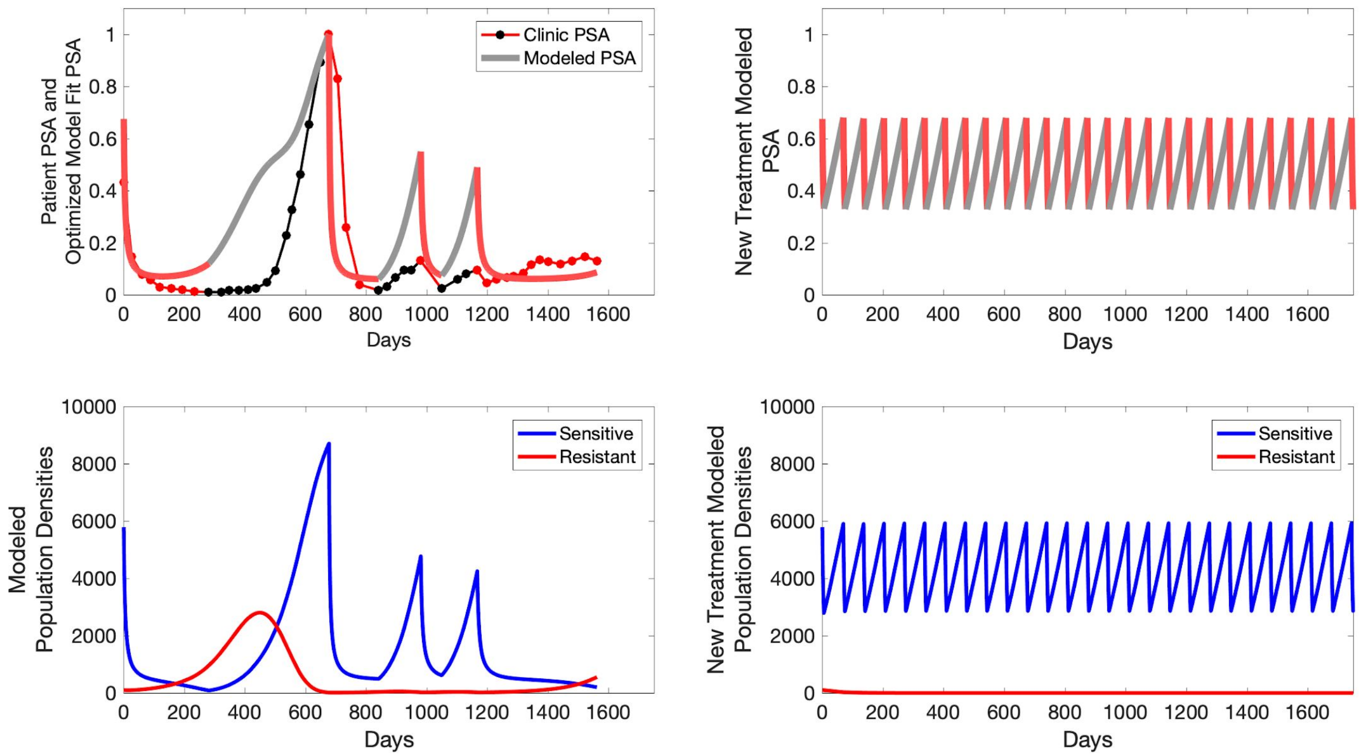

Clinic PSA
Modeled PSA
Patient PSA and Optimized Model Fit PSA
Days
New Treatment Modeled PSA
Days
Sensitive
Resistant
Modeled Population Densities
Days
Sensitive
Resistant
New Treatment Modeled Population Densities
Days

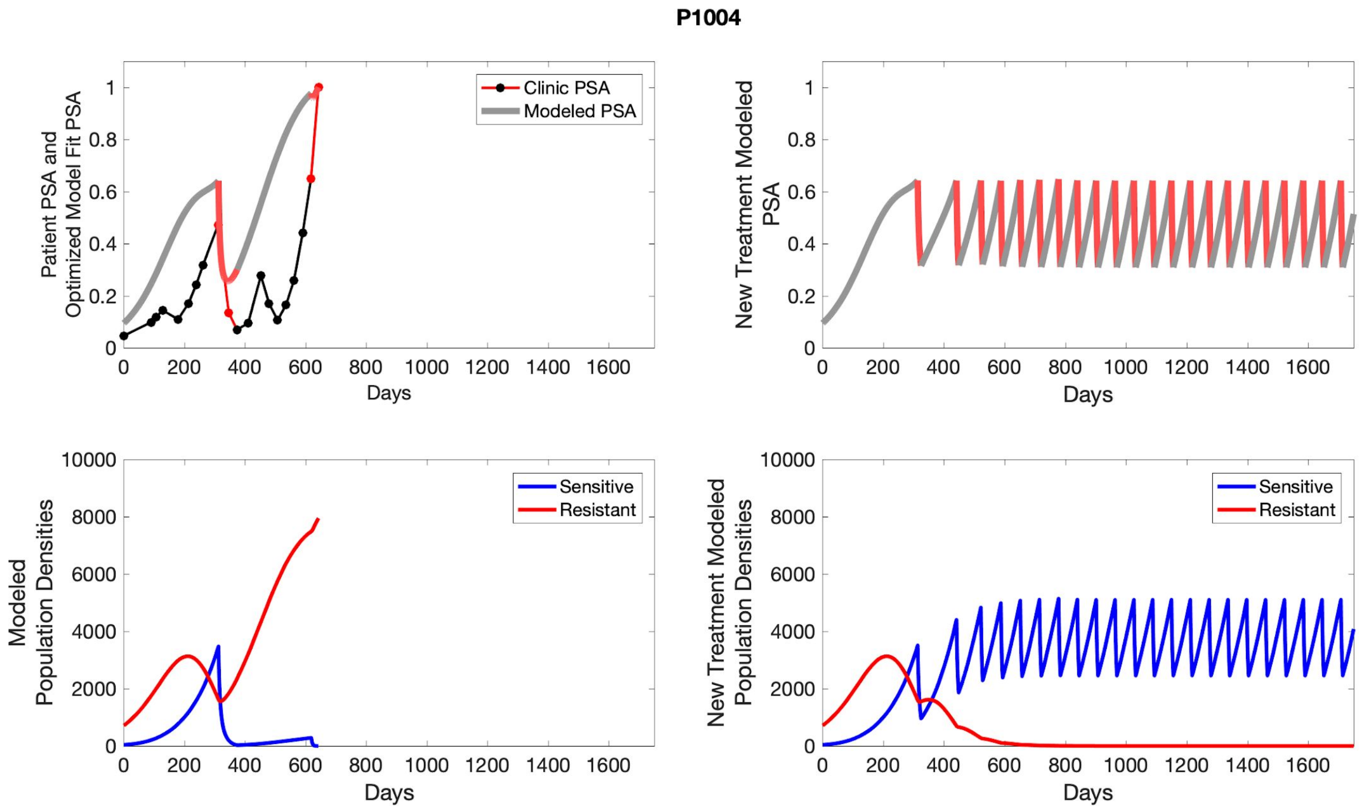
P1004
Patient PSA and Optimized Model Fit PSA
Clinic PSA
Modeled PSA
Days
New Treatment Modeled PSA
Days
Modeled Population Densities
Sensitive
Resistant
Days
New Treatment Modeled Population Densities
Sensitive
Resistant
Days

**P1005**

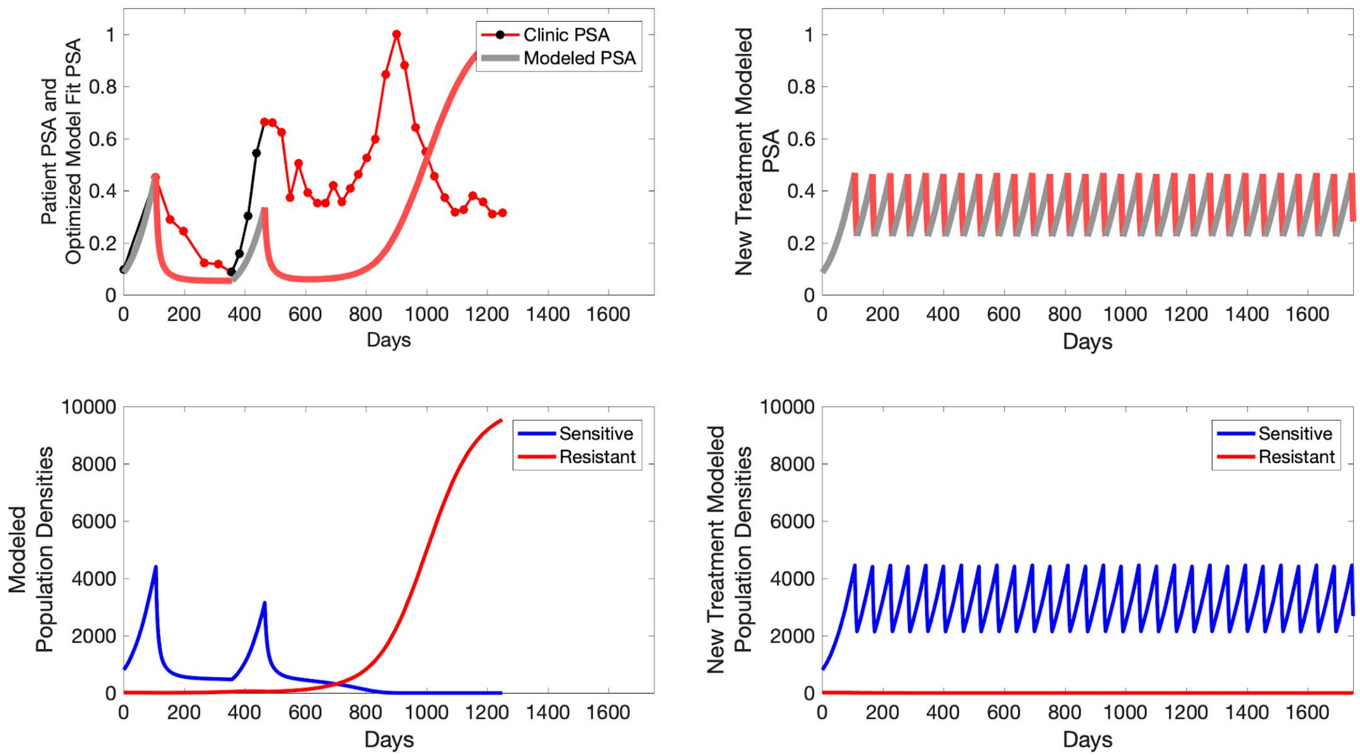

Clinic PSA
Modeled PSA
Patient PSA and Optimized Model Fit PSA
Days
New Treatment Modeled PSA
Days
Sensitive
Resistant
Modeled Population Densities
Days
Sensitive
Resistant
New Treatment Modeled Population Densities
Days

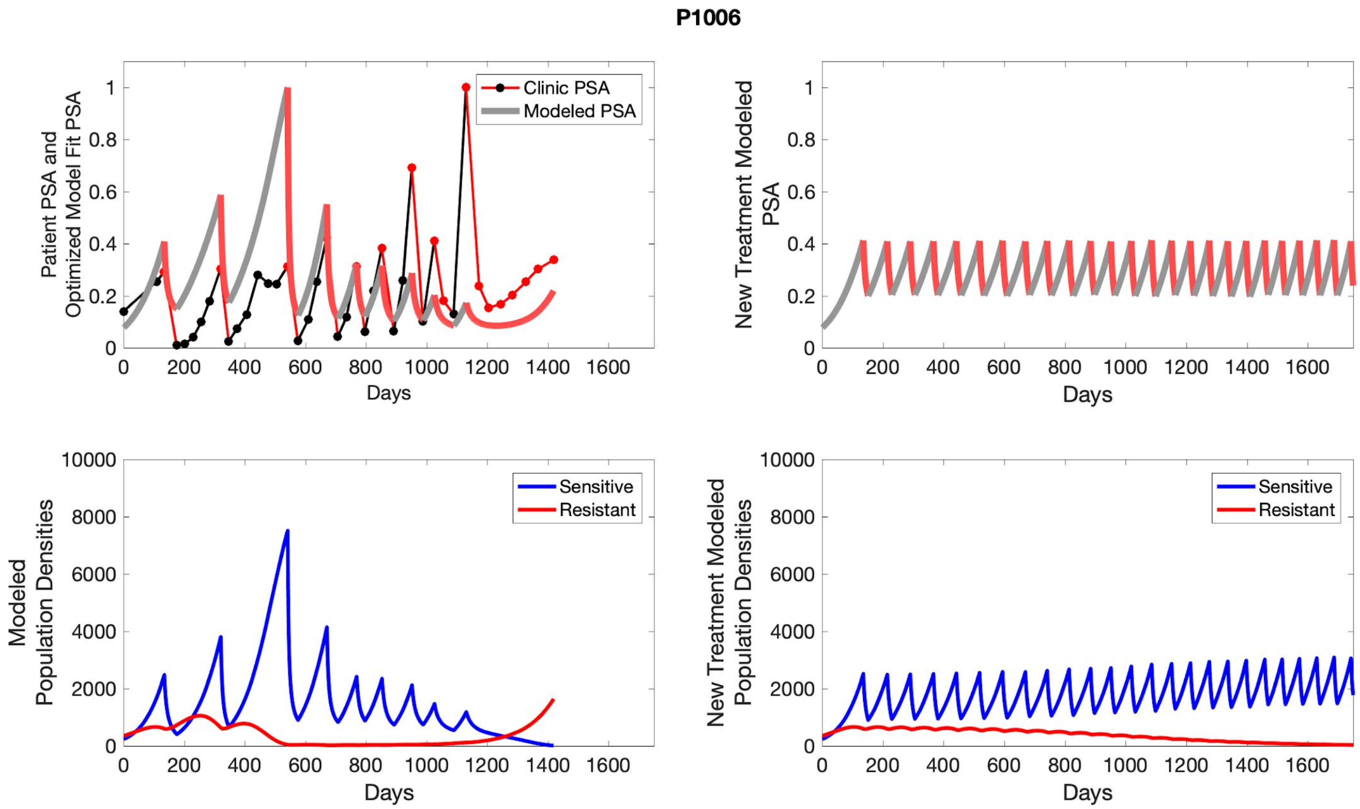
P1006
Patient PSA and Optimized Model Fit PSA
Clinic PSA
Modeled PSA
Days
New Treatment Modeled PSA
Days
Modeled Population Densities
Sensitive
Resistant
Days
New Treatment Modeled Population Densities
Sensitive
Resistant
Days

**P1007**

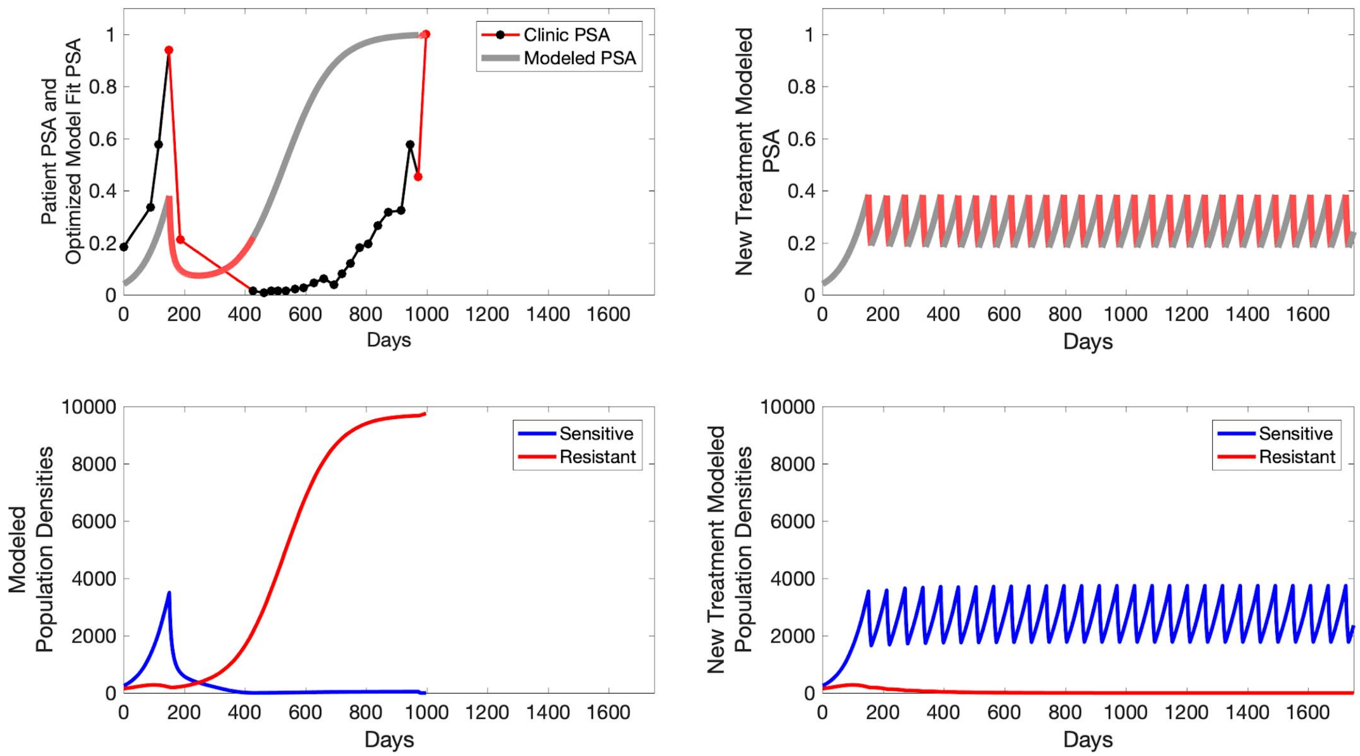

Patient PSA and Optimized Model Fit PSA
Clinic PSA
Modeled PSA
Days
New Treatment Modeled PSA
Days
Modeled Population Densities
Sensitive
Resistant
Days
New Treatment Modeled Population Densities
Sensitive
Resistant
Days

**P1009**

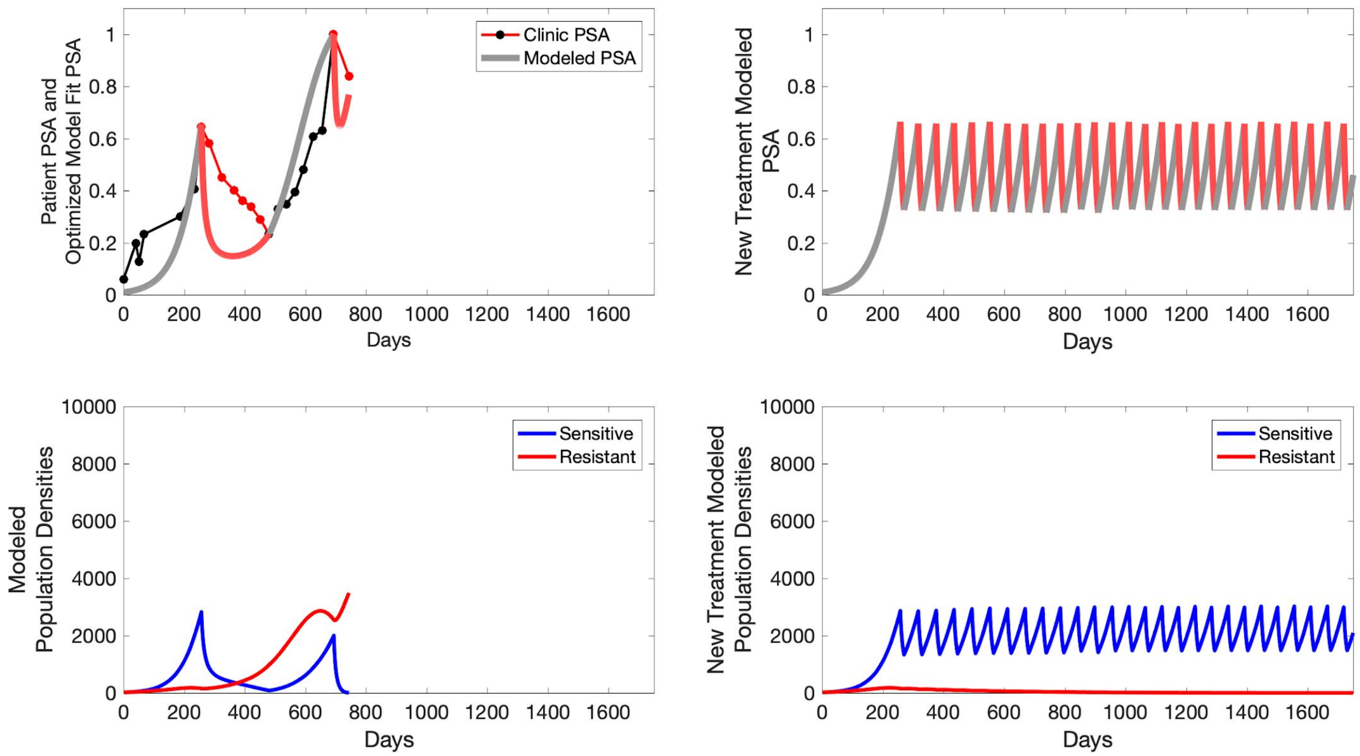

Clinic PSA
Modeled PSA
Patient PSA and Optimized Model Fit PSA
New Treatment Modeled PSA
Days
Sensitive
Resistant
Modeled Population Densities
New Treatment Modeled Population Densities

**P1010**

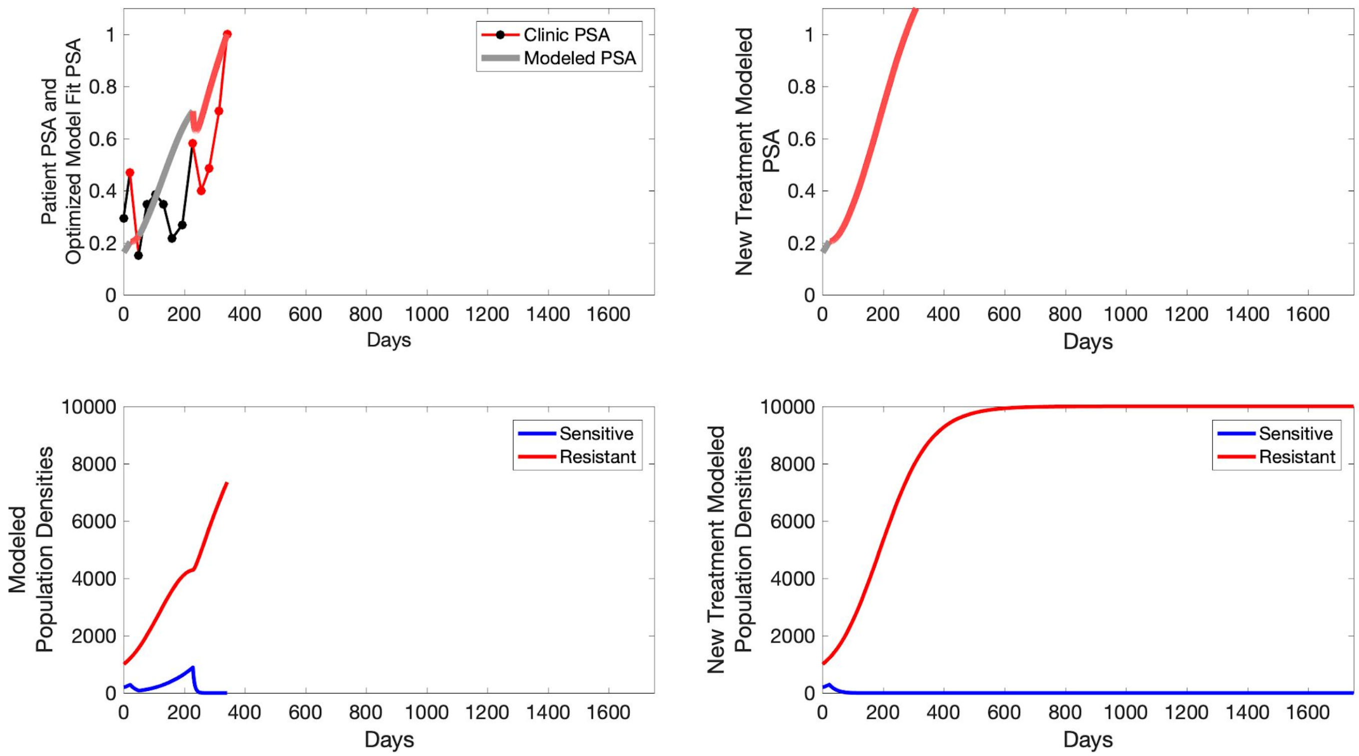

Patient PSA and Optimized Model Fit PSA
Clinic PSA
Modeled PSA
Days
New Treatment Modeled PSA
Days
Modeled Population Densities
Sensitive
Resistant
Days
New Treatment Modeled Population Densities
Sensitive
Resistant
Days

**P1011**

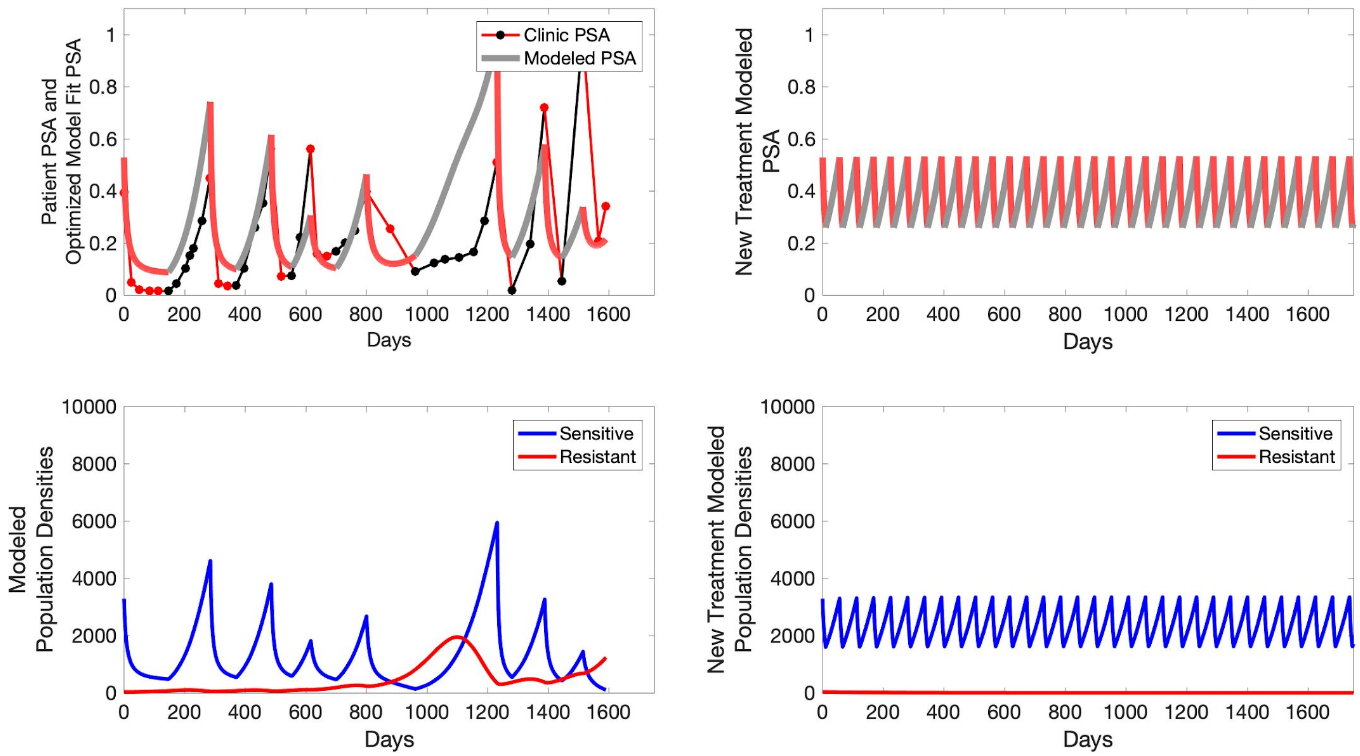

Clinic PSA
Modeled PSA
Patient PSA and Optimized Model Fit PSA
Days
New Treatment Modeled PSA
Days
Sensitive
Resistant
Modeled Population Densities
Days
Sensitive
Resistant
New Treatment Modeled Population Densities
Days

**P1012**

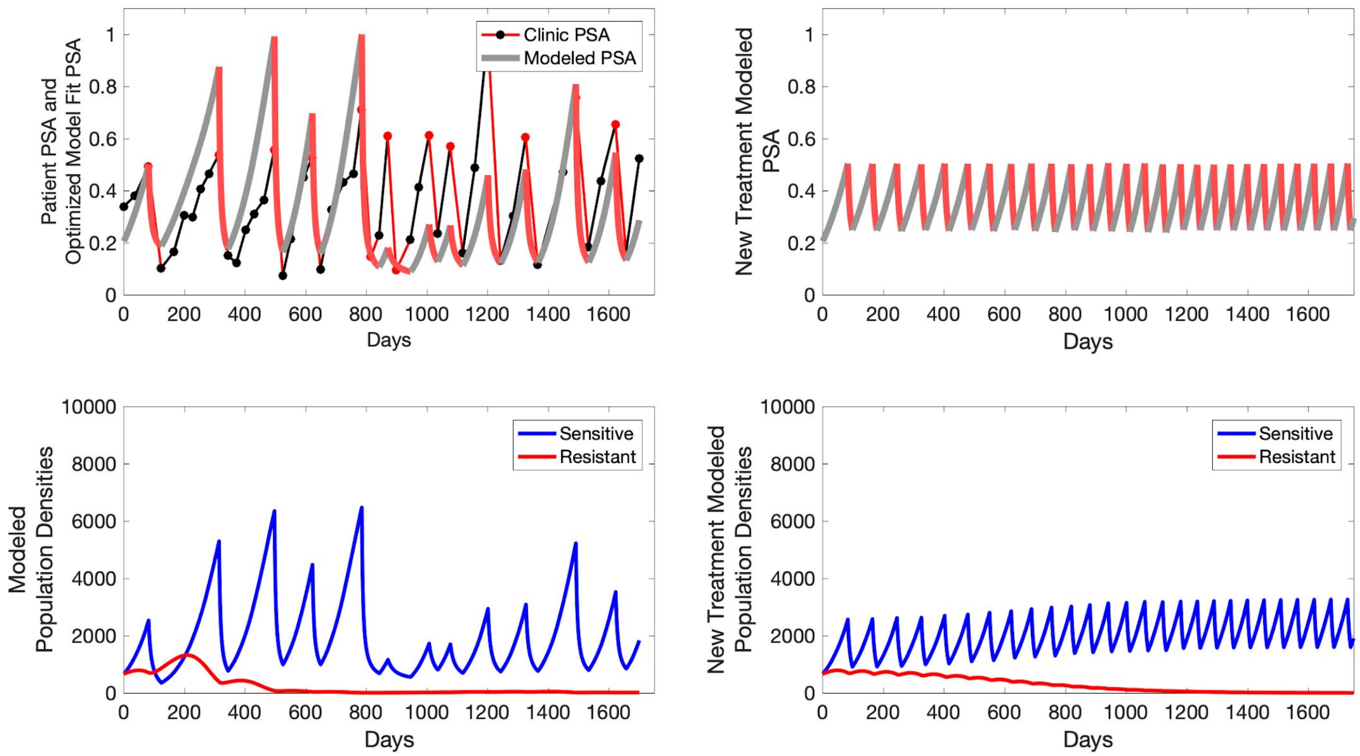

Patient PSA and Optimized Model Fit PSA
Clinic PSA
Modeled PSA
Days
New Treatment Modeled PSA
Days
Modeled Population Densities
Sensitive
Resistant
Days
New Treatment Modeled Population Densities
Sensitive
Resistant
Days

**P1014**

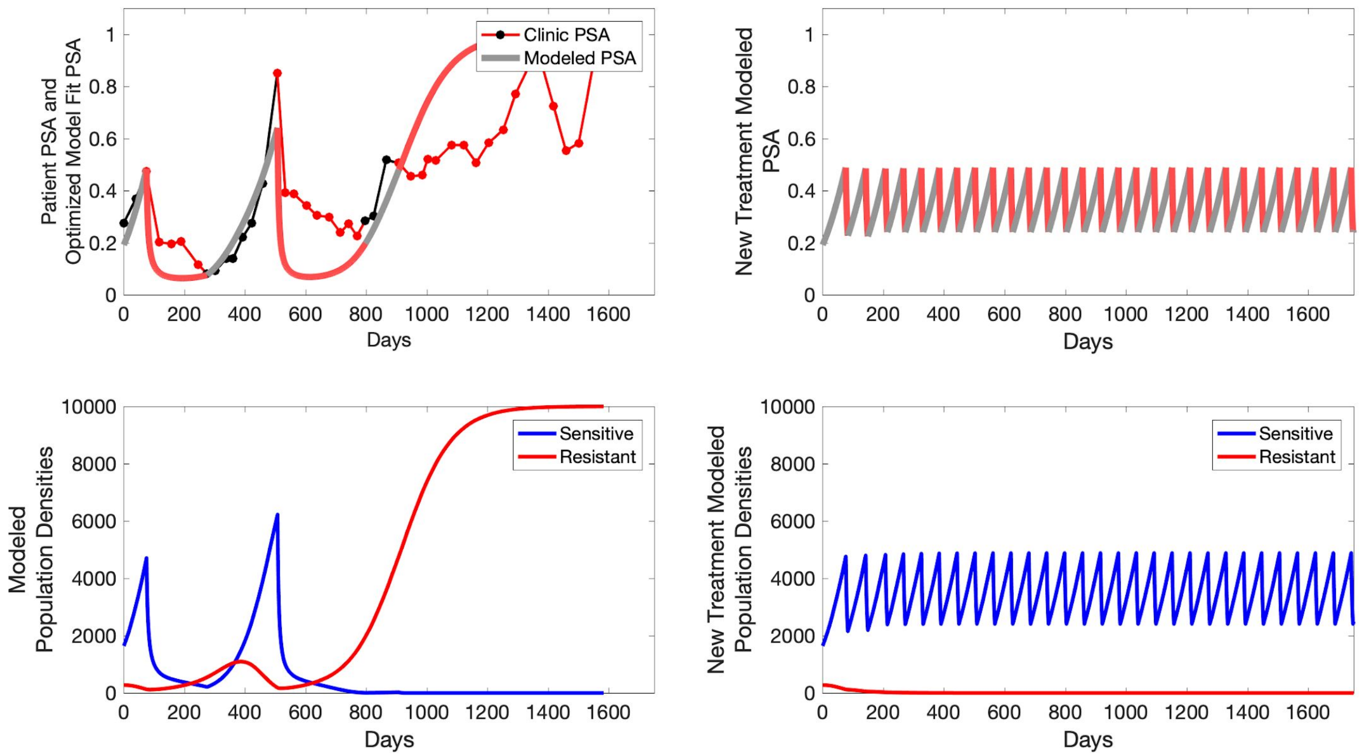

Patient PSA and Optimized Model Fit PSA
Clinic PSA
Modeled PSA
Days
New Treatment Modeled PSA
Days
Modeled Population Densities
Sensitive
Resistant
Days
New Treatment Modeled Population Densities
Sensitive
Resistant
Days

P1015

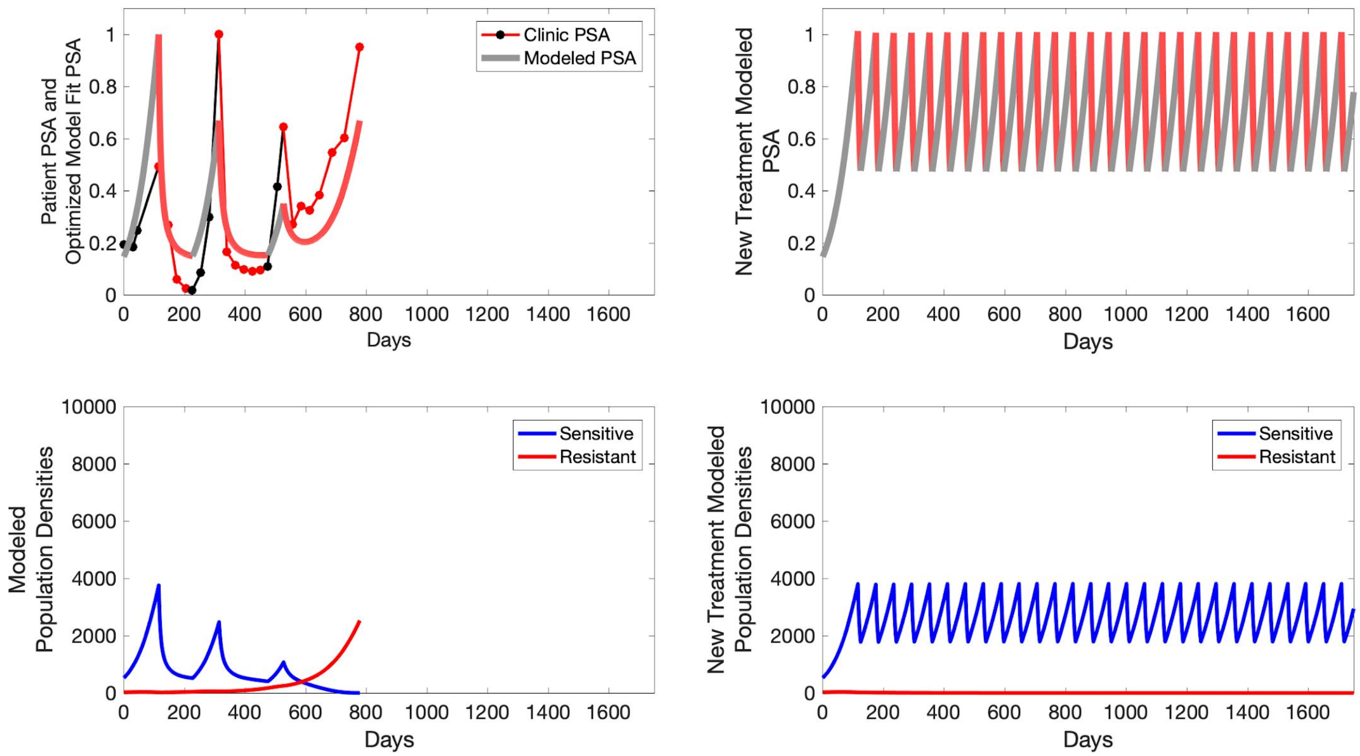

Patient PSA and Optimized Model Fit PSA
Clinic PSA
Modeled PSA
Days
New Treatment Modeled PSA
Days
Modeled Population Densities
Sensitive
Resistant
Days
New Treatment Modeled Population Densities
Sensitive
Resistant
Days

**P1016**

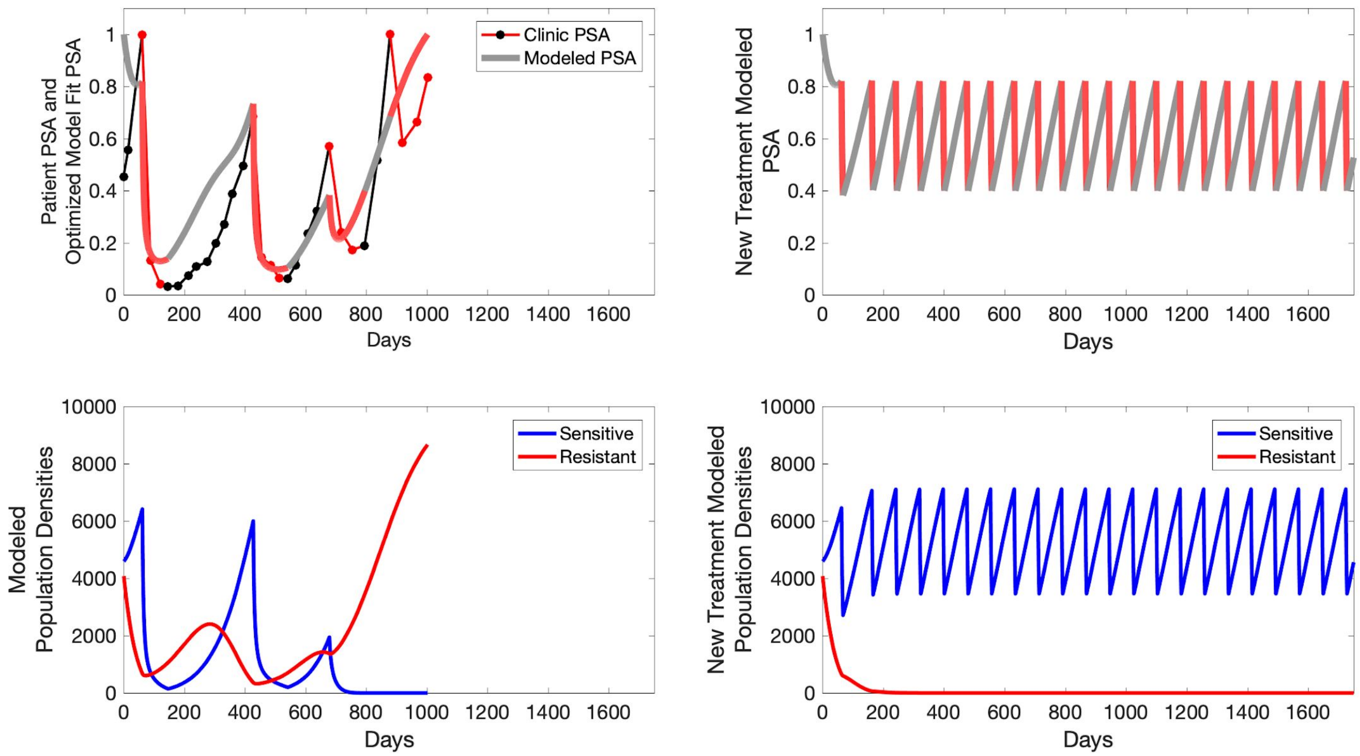

Patient PSA and Optimized Model Fit PSA
Clinic PSA
Modeled PSA
Days
New Treatment Modeled PSA
Days
Modeled Population Densities
Sensitive
Resistant
Days
New Treatment Modeled Population Densities
Sensitive
Resistant
Days

**P1017**

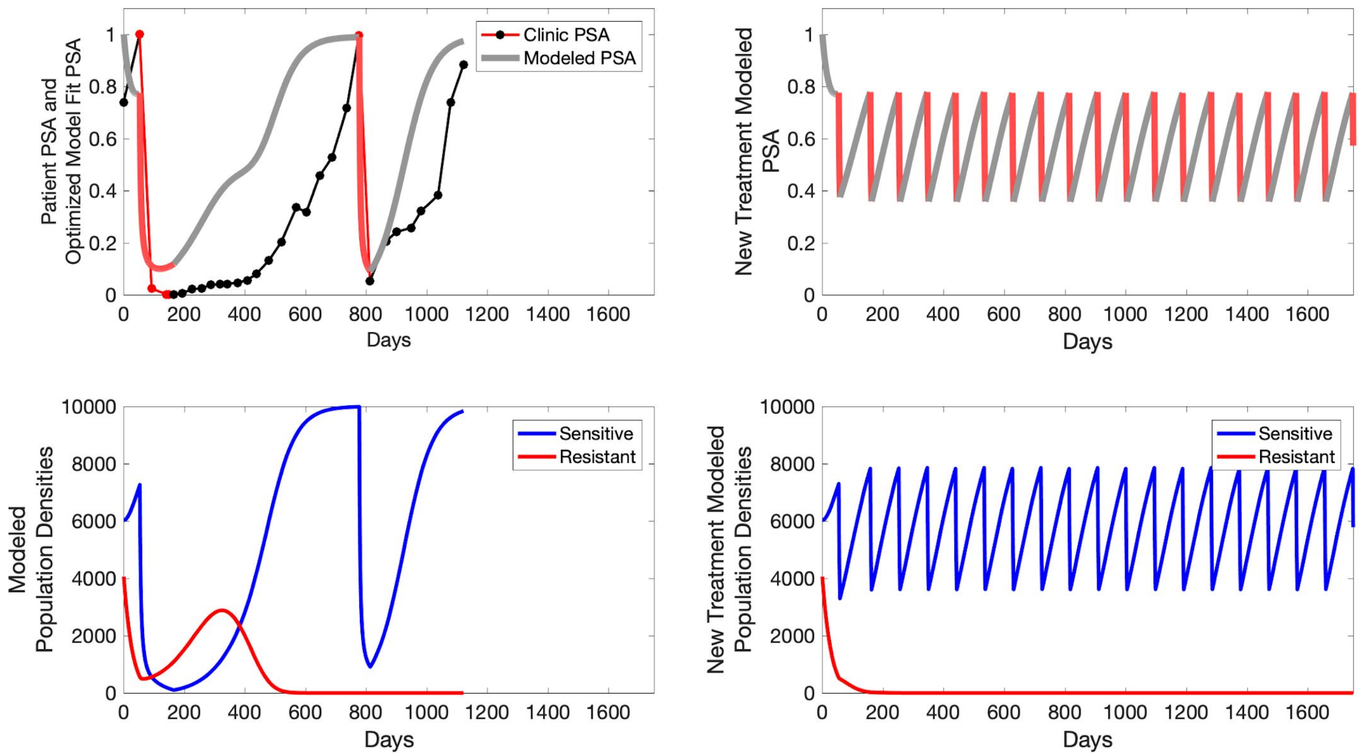

Clinic PSA
Modeled PSA
Patient PSA and Optimized Model Fit PSA
Days
New Treatment Modeled PSA
Days
Sensitive
Resistant
Modeled Population Densities
Days
Sensitive
Resistant
New Treatment Modeled Population Densities
Days

**P1018**

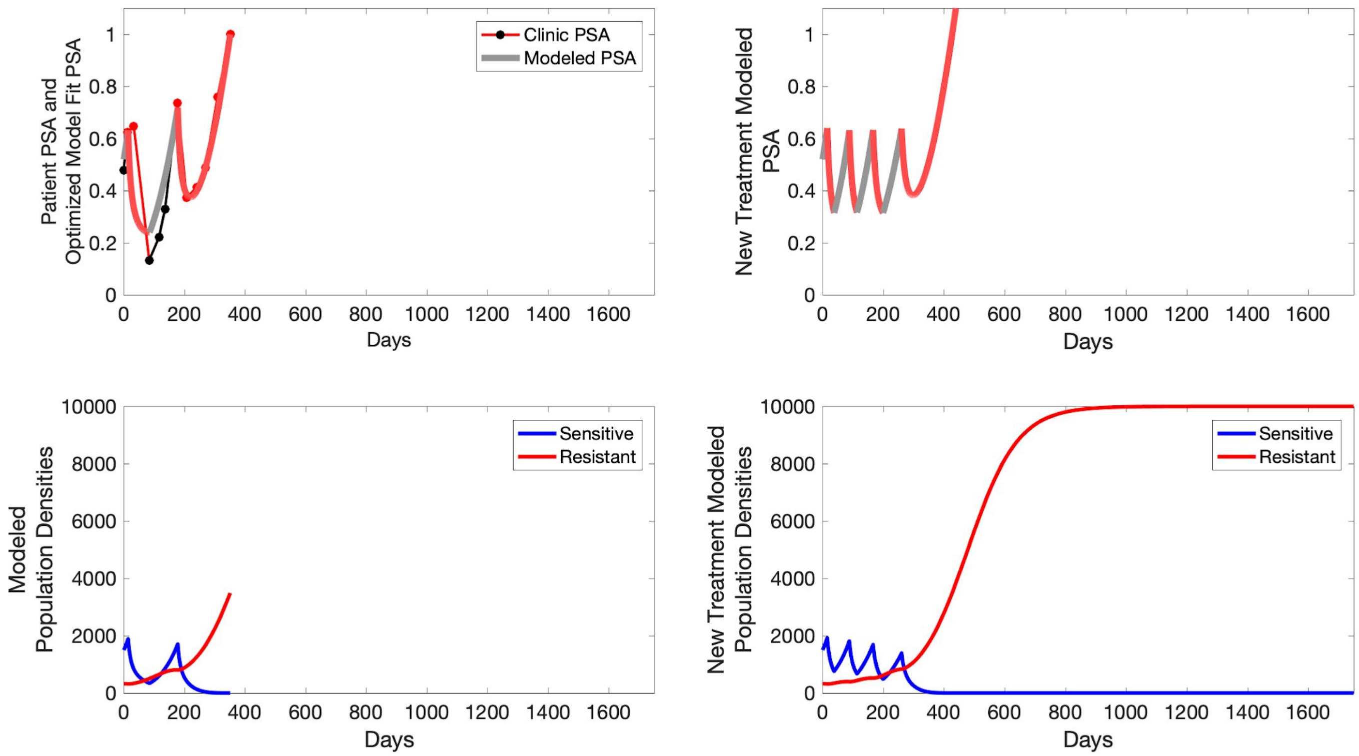

Clinic PSA
Modeled PSA
Patient PSA and Optimized Model Fit PSA
New Treatment Modeled PSA
Days
Sensitive
Resistant
Modeled Population Densities
New Treatment Modeled Population Densities

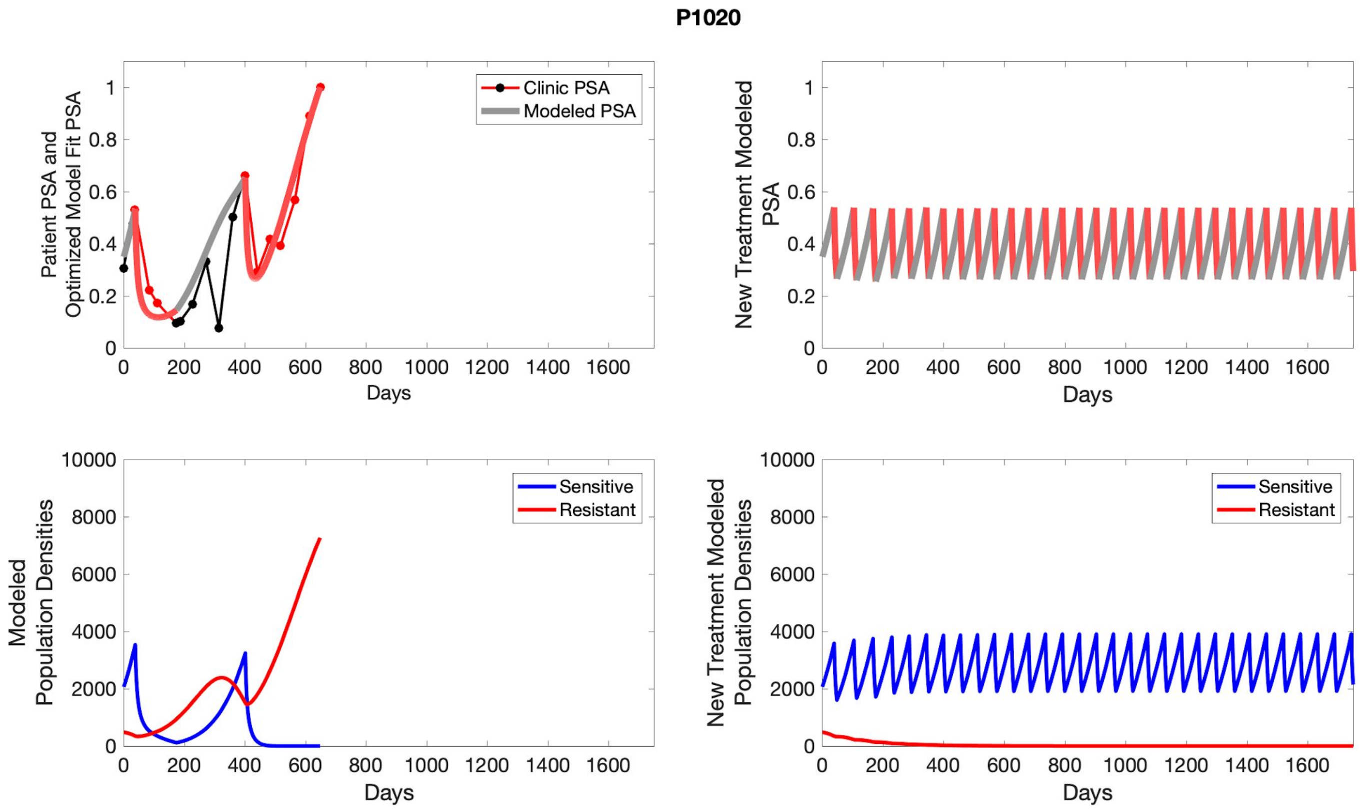
P1020
Clinic PSA
Modeled PSA
Patient PSA and Optimized Model Fit PSA
Days
New Treatment Modeled PSA
Sensitive
Resistant
Modeled Population Densities
New Treatment Modeled Population Densities

#### S6 Simulated Standard Of Care

To evaluate standard of care protocol, we ran a simulation for each patient (both cohorts). For each patient, we initiated model runs using the patient specific parameters from Table S4. We considered continuous abiraterone therapy for both cohorts. Standard of care was initiated at the time abiraterone was first administered clinically to each patient, which was not always at t=0. For the contemporaneous cohort that received standard of care clinically, we ran the dynamics of standard of care beyond clinical measurements to predict how the disease would have progressed in the absence of any other treatments. For the adaptive therapy patients, we could predict how each patient would have fared under standard of care abiraterone.

C001

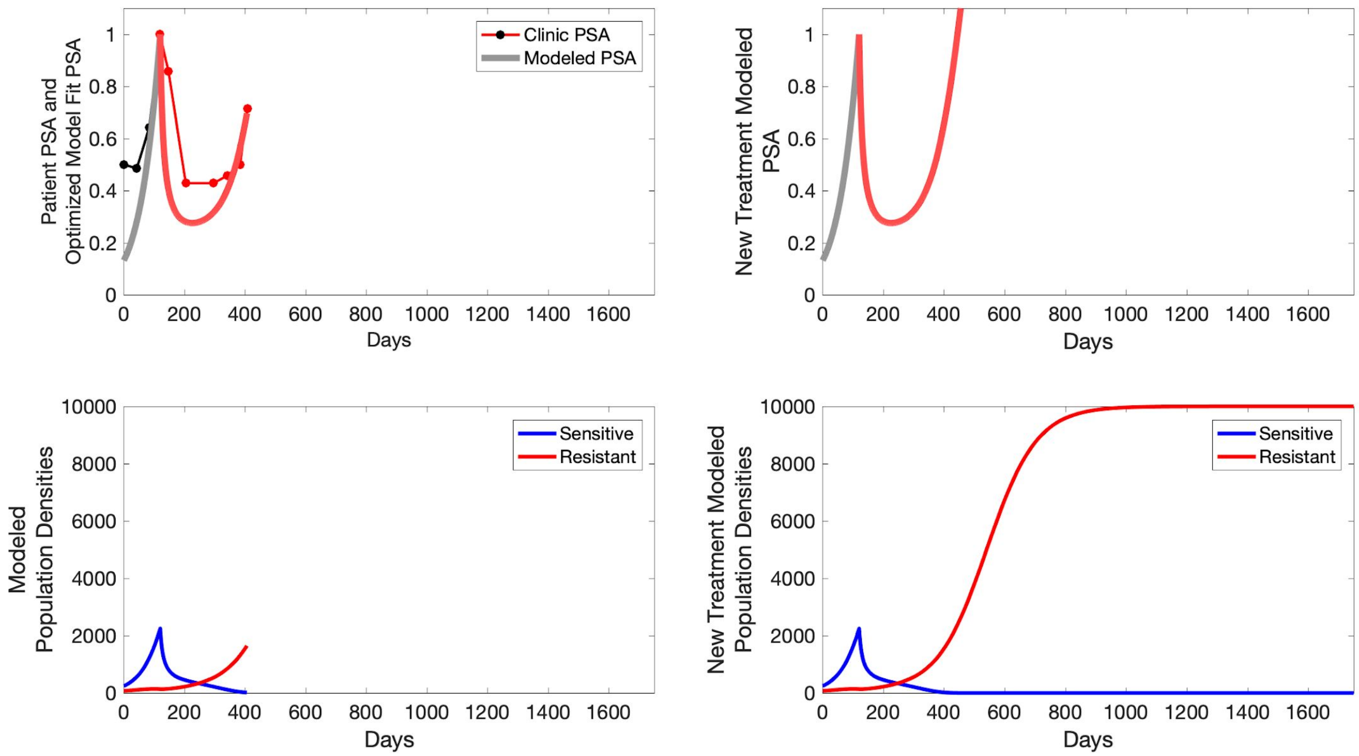

Patient PSA and Optimized Model Fit PSA
Clinic PSA
Modeled PSA
Days
New Treatment Modeled PSA
Days
Modeled Population Densities
Sensitive
Resistant
Days
New Treatment Modeled Population Densities
Sensitive
Resistant
Days

**C002**

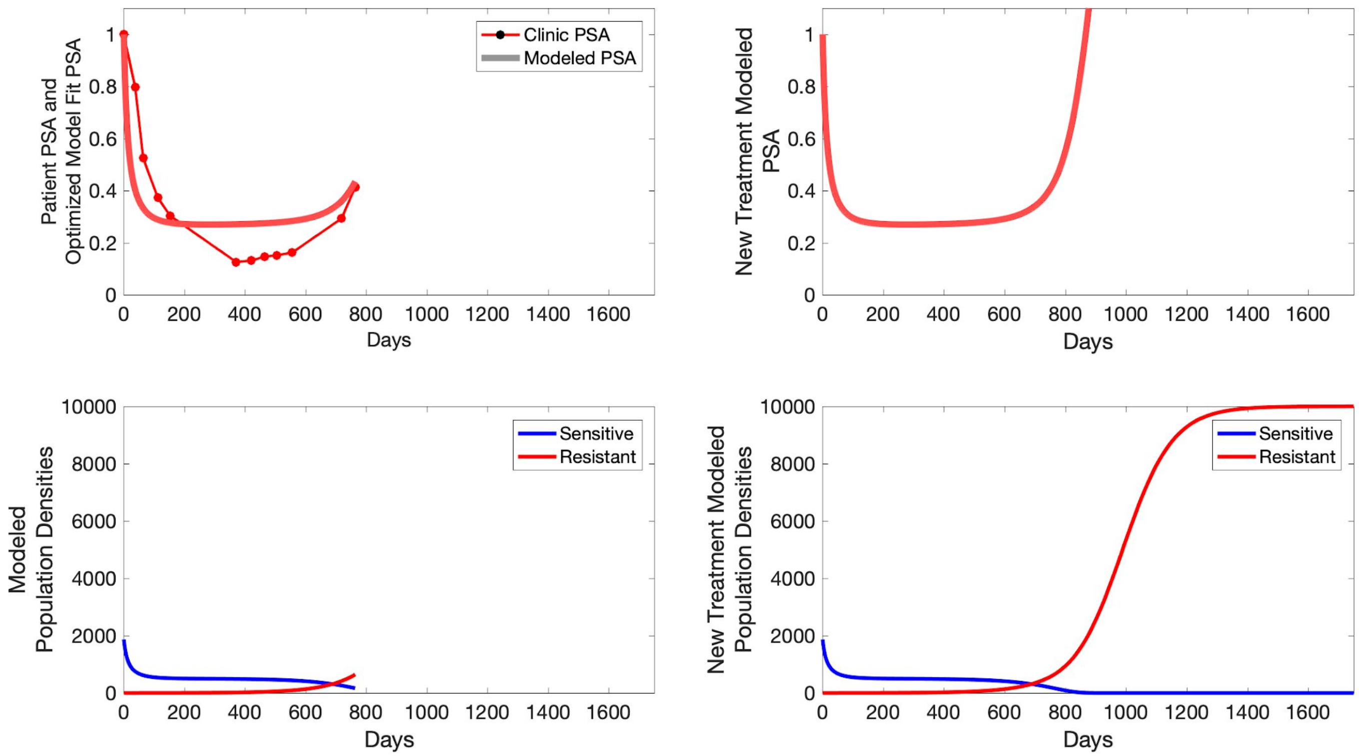

Patient PSA and Optimized Model Fit PSA
Clinic PSA
Modeled PSA
Days
New Treatment Modeled PSA
Days
Modeled Population Densities
Sensitive
Resistant
Days
New Treatment Modeled Population Densities
Sensitive
Resistant
Days

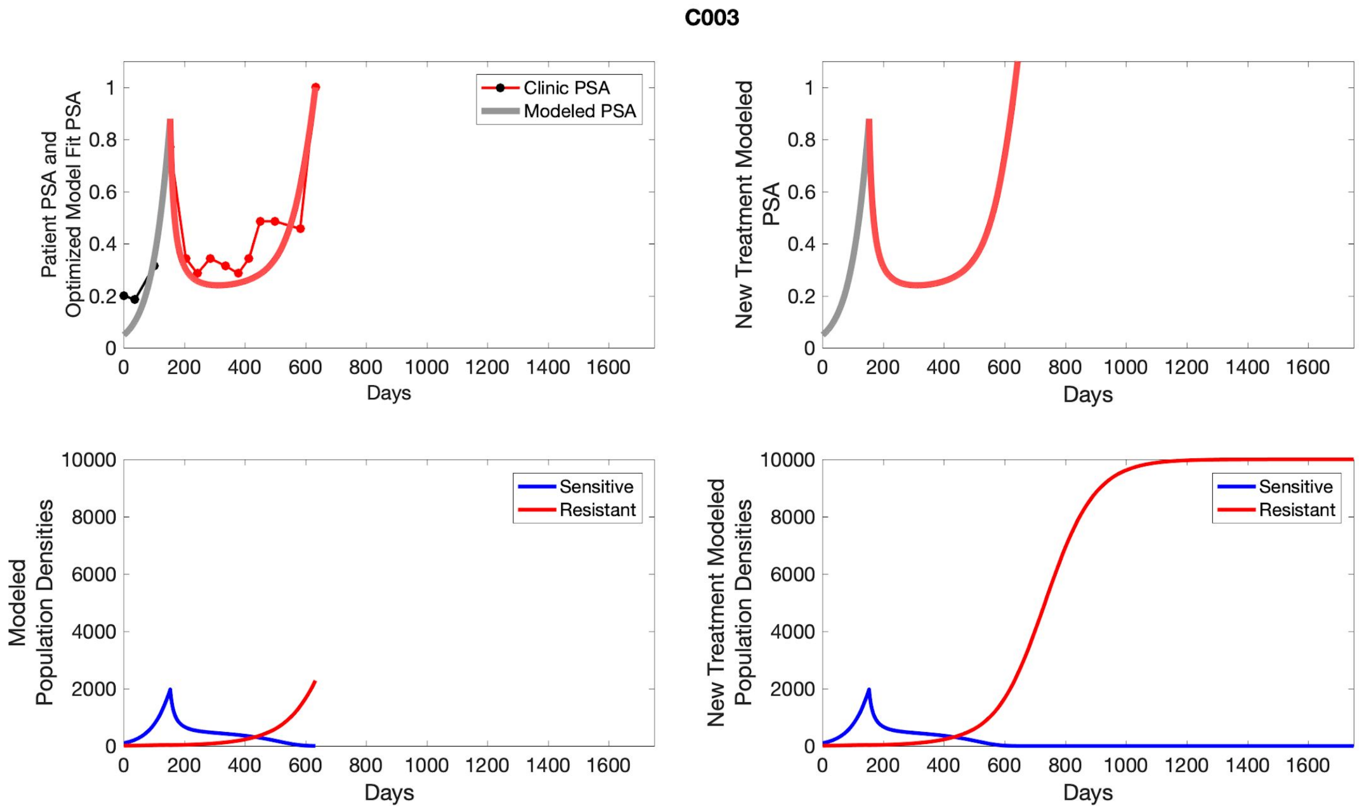
C003
Patient PSA and Optimized Model Fit PSA
Clinic PSA
Modeled PSA
Days
New Treatment Modeled PSA
Modeled Population Densities
Sensitive
Resistant
New Treatment Modeled Population Densities

**C004**

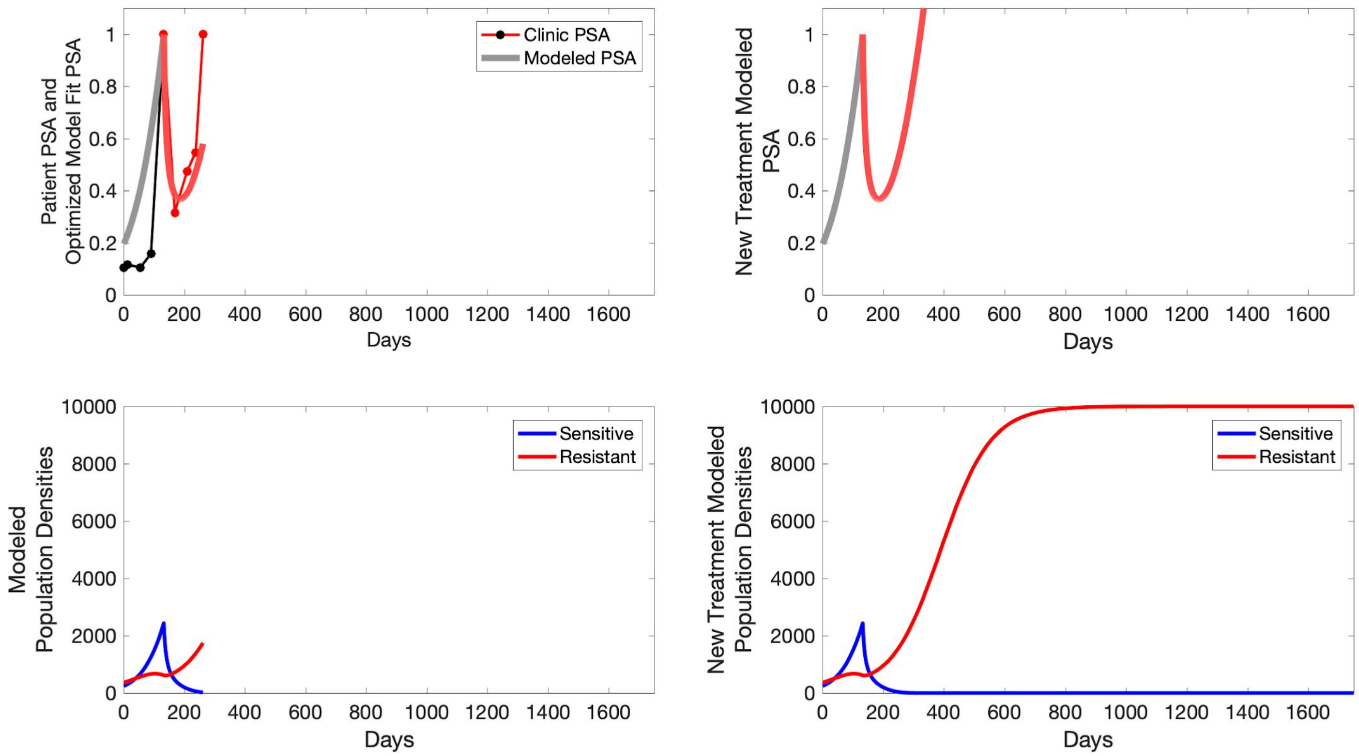

Patient PSA and Optimized Model Fit PSA
Clinic PSA
Modeled PSA
Days
New Treatment Modeled PSA
Days
Modeled Population Densities
Sensitive
Resistant
Days
New Treatment Modeled Population Densities
Sensitive
Resistant
Days

**C005**

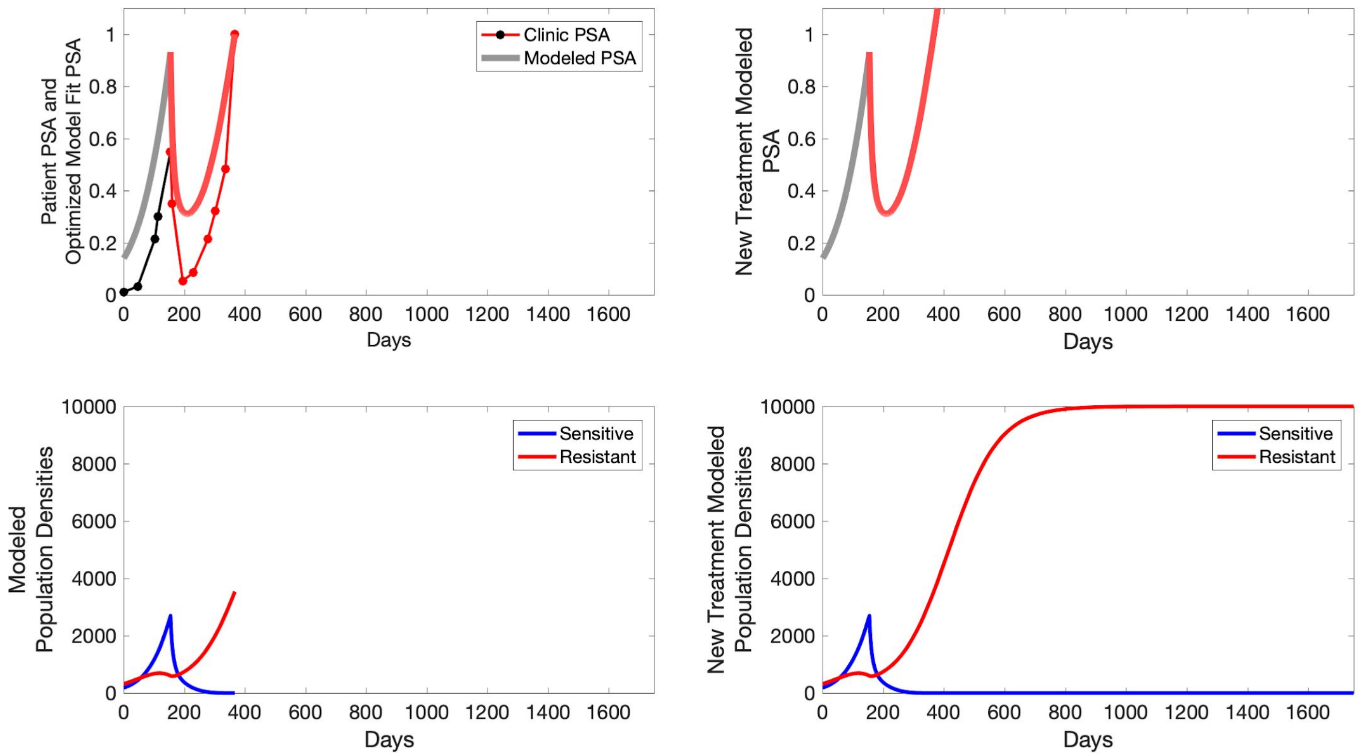

Clinic PSA
Modeled PSA
Patient PSA and Optimized Model Fit PSA
New Treatment Modeled PSA
Days
Sensitive
Resistant
Modeled Population Densities
New Treatment Modeled Population Densities

**C006**

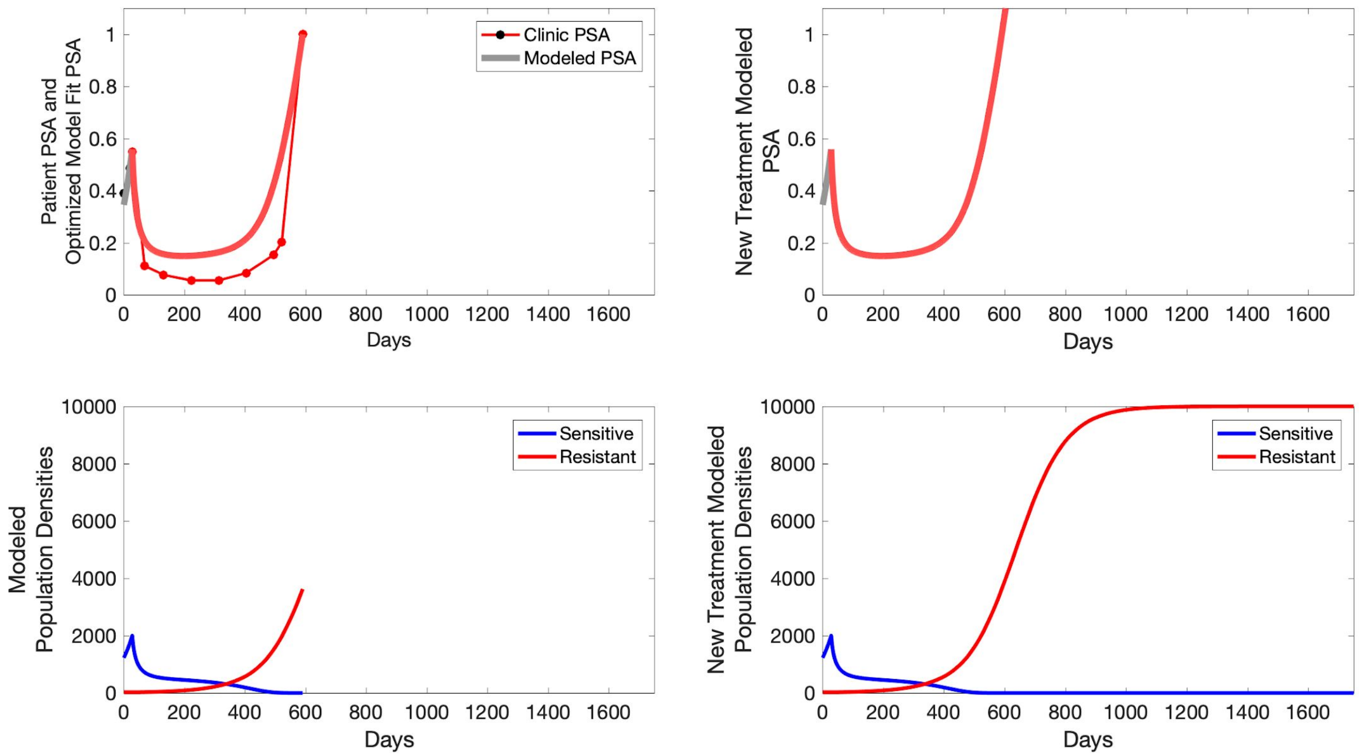

Patient PSA and Optimized Model Fit PSA
Clinic PSA
Modeled PSA
Days
New Treatment Modeled PSA
Days
Modeled Population Densities
Sensitive
Resistant
Days
New Treatment Modeled Population Densities
Sensitive
Resistant
Days

**C007**

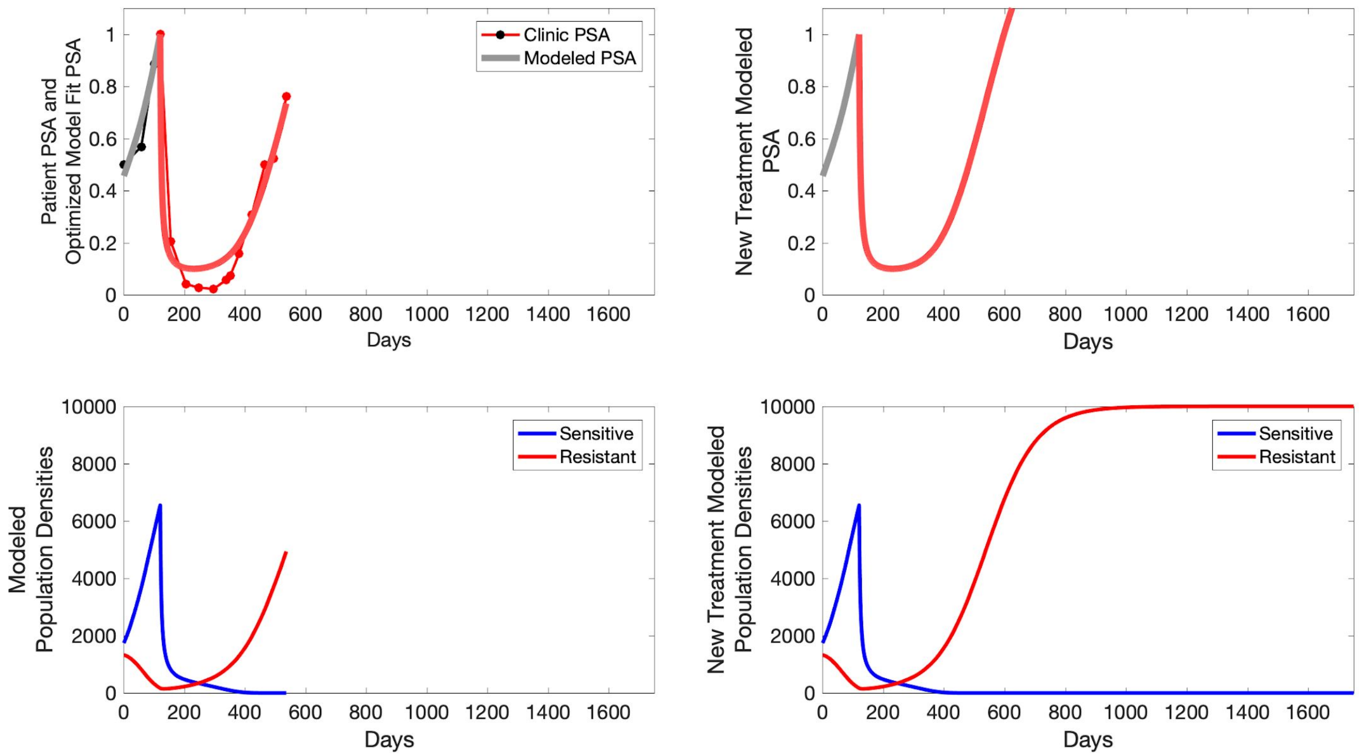

Patient PSA and Optimized Model Fit PSA
Clinic PSA
Modeled PSA
Days
New Treatment Modeled PSA
Days
Modeled Population Densities
Sensitive
Resistant
Days
New Treatment Modeled Population Densities
Sensitive
Resistant
Days

**C008**

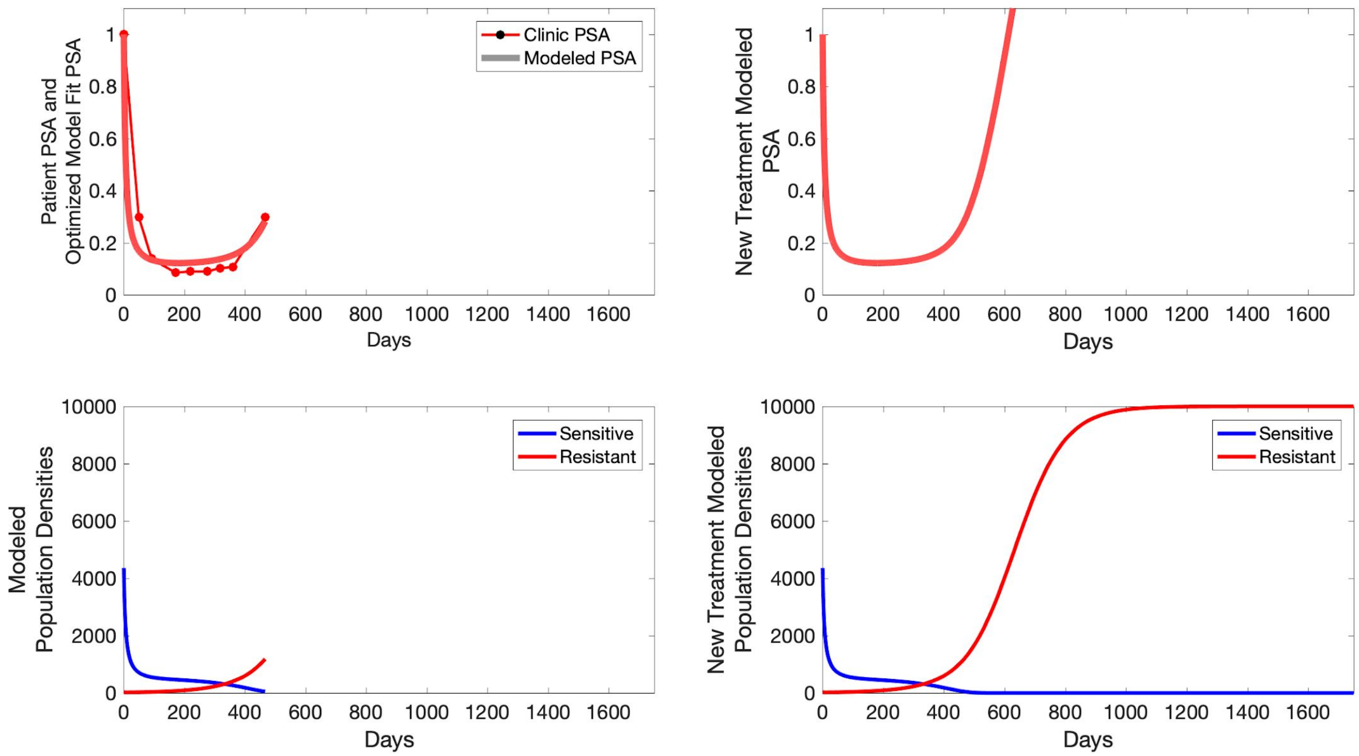

Patient PSA and Optimized Model Fit PSA
Clinic PSA
Modeled PSA
Days
New Treatment Modeled PSA
Days
Modeled Population Densities
Sensitive
Resistant
Days
New Treatment Modeled Population Densities
Sensitive
Resistant
Days

**C009**

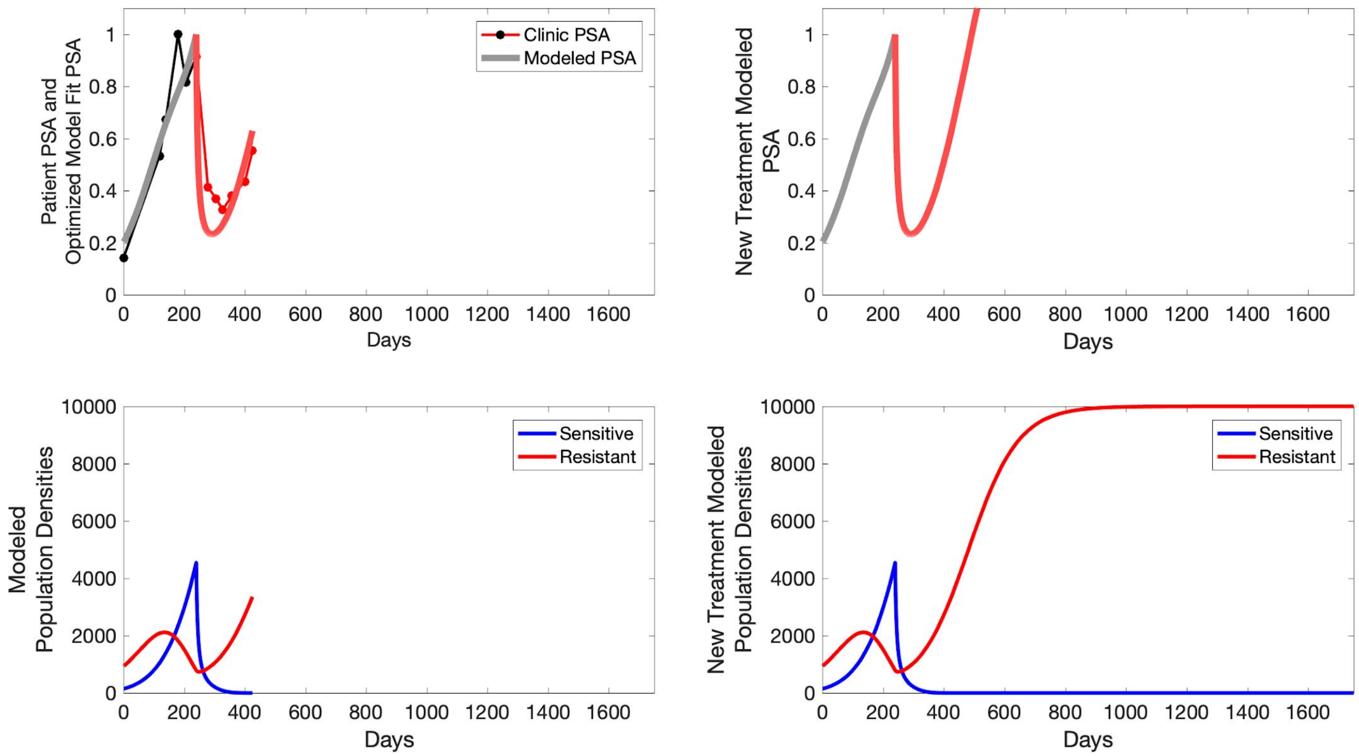

Patient PSA and Optimized Model Fit PSA
Clinic PSA
Modeled PSA
Days
New Treatment Modeled PSA
Days
Modeled Population Densities
Sensitive
Resistant
Days
New Treatment Modeled Population Densities
Sensitive
Resistant
Days

**C010**

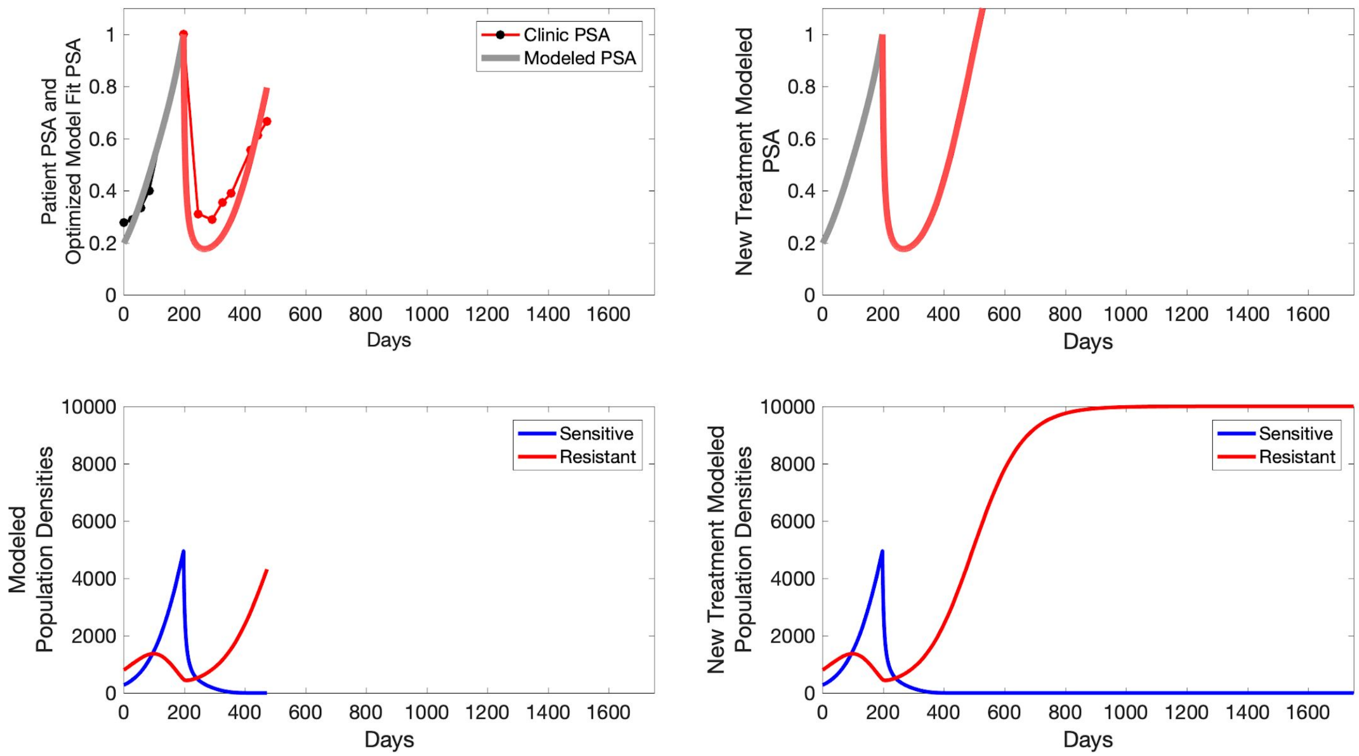

Patient PSA and
Optimized Model Fit PSA
Clinic PSA
Modeled PSA
Days
New Treatment Modeled
PSA
Days
Modeled
Population Densities
Sensitive
Resistant
Days
New Treatment Modeled
Population Densities
Sensitive
Resistant
Days

**C011**

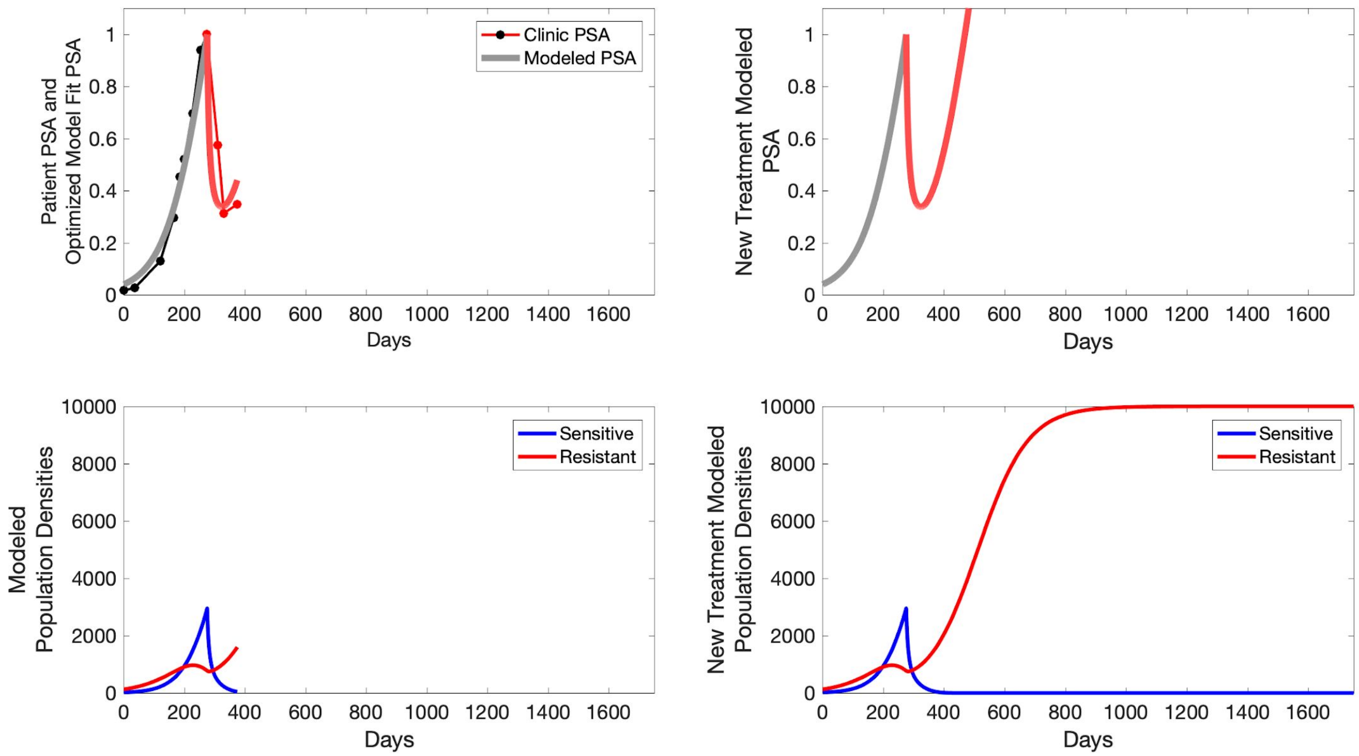

Patient PSA and Optimized Model Fit PSA
Clinic PSA
Modeled PSA
Days
New Treatment Modeled PSA
Days
Modeled Population Densities
Sensitive
Resistant
Days
New Treatment Modeled Population Densities
Sensitive
Resistant
Days

**C012**

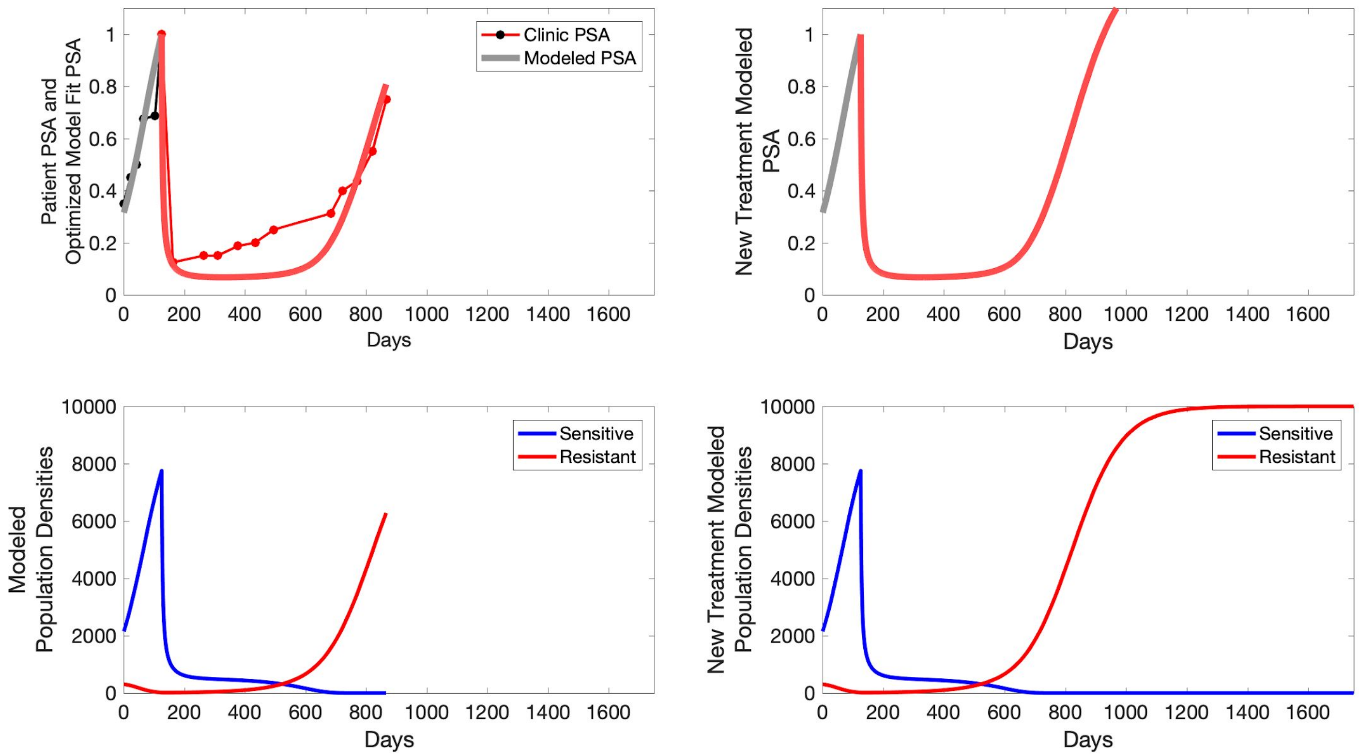
Patient PSA and Optimized Model Fit PSA
Clinic PSA
Modeled PSA
Days
New Treatment Modeled PSA
Days
Modeled Population Densities
Sensitive
Resistant
Days
New Treatment Modeled Population Densities
Sensitive
Resistant
Days

**C013**

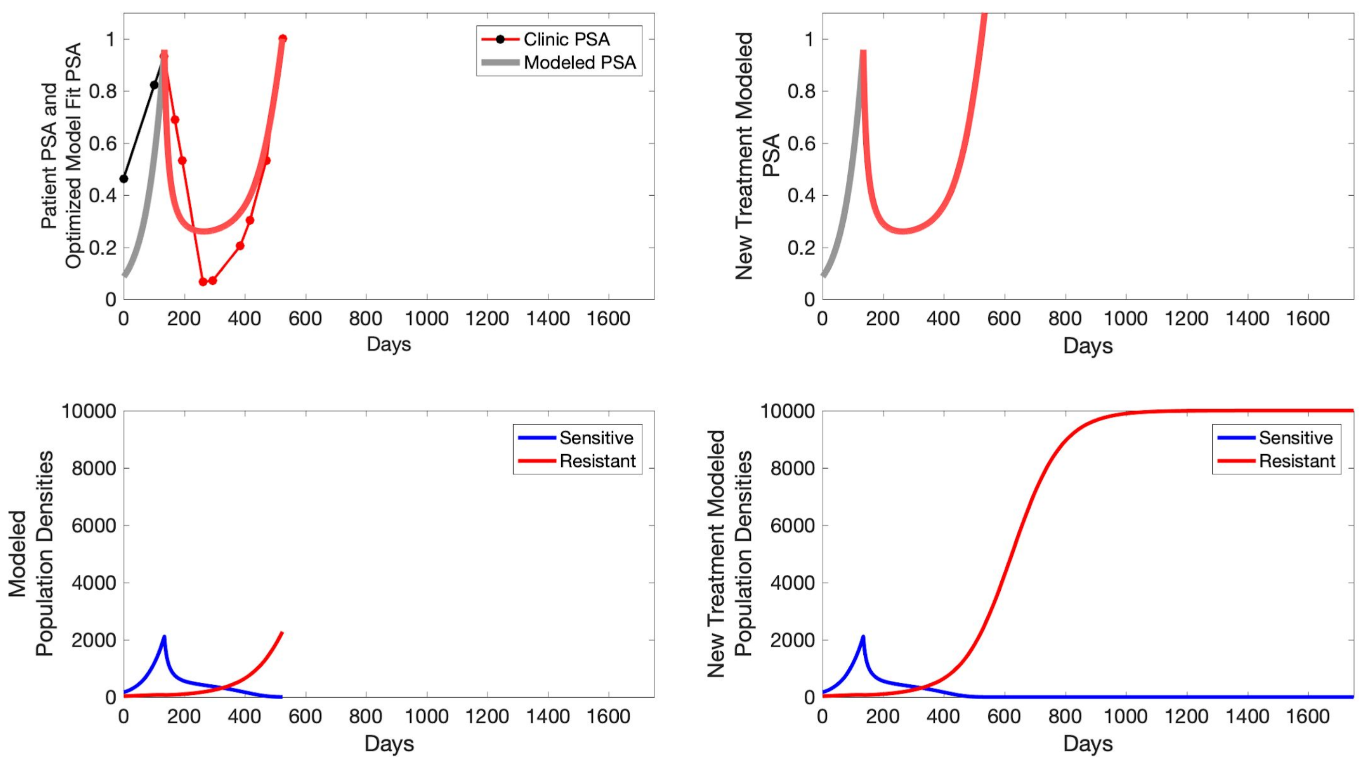

Patient PSA and Optimized Model Fit PSA
Clinic PSA
Modeled PSA
Days
New Treatment Modeled PSA
Days
Modeled Population Densities
Sensitive
Resistant
Days
New Treatment Modeled Population Densities
Sensitive
Resistant
Days

**C014**

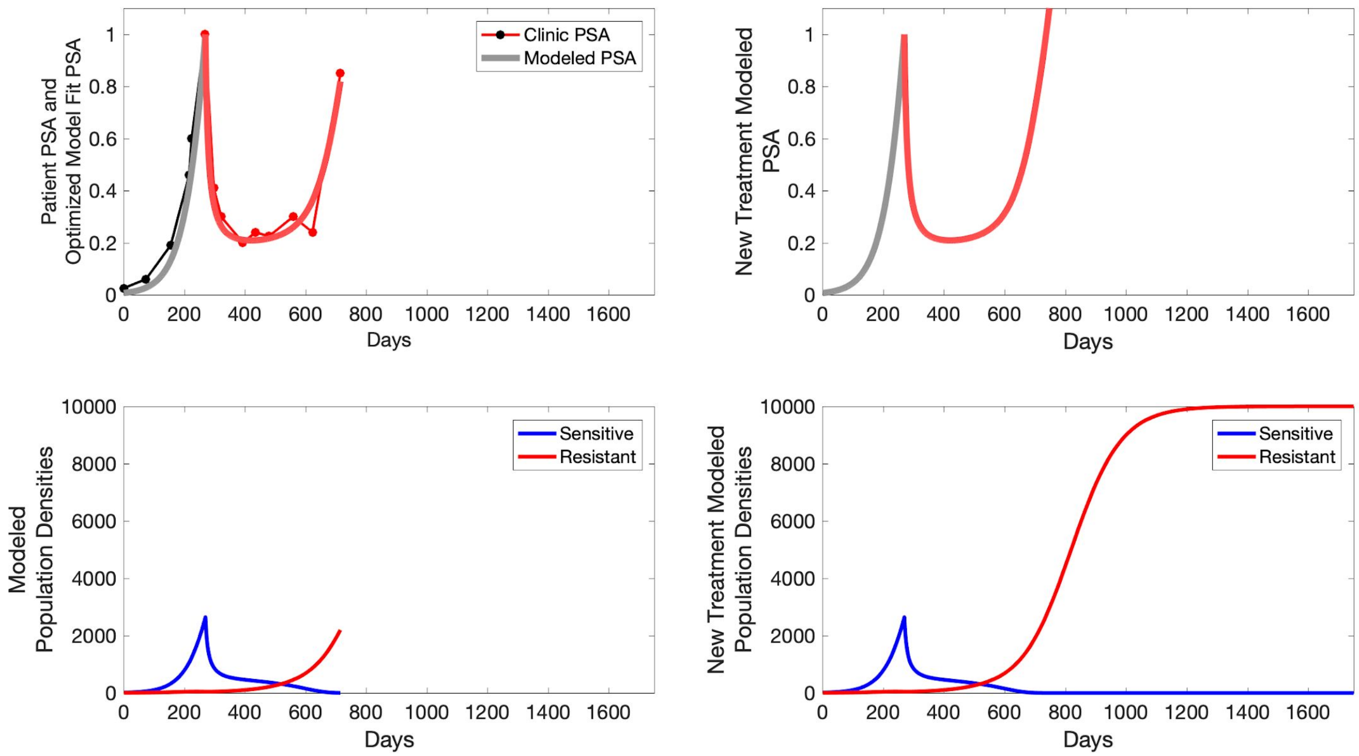

Patient PSA and Optimized Model Fit PSA
Clinic PSA
Modeled PSA
Days
New Treatment Modeled PSA
Days
Modeled Population Densities
Sensitive
Resistant
Days
New Treatment Modeled Population Densities
Sensitive
Resistant
Days

**C015**

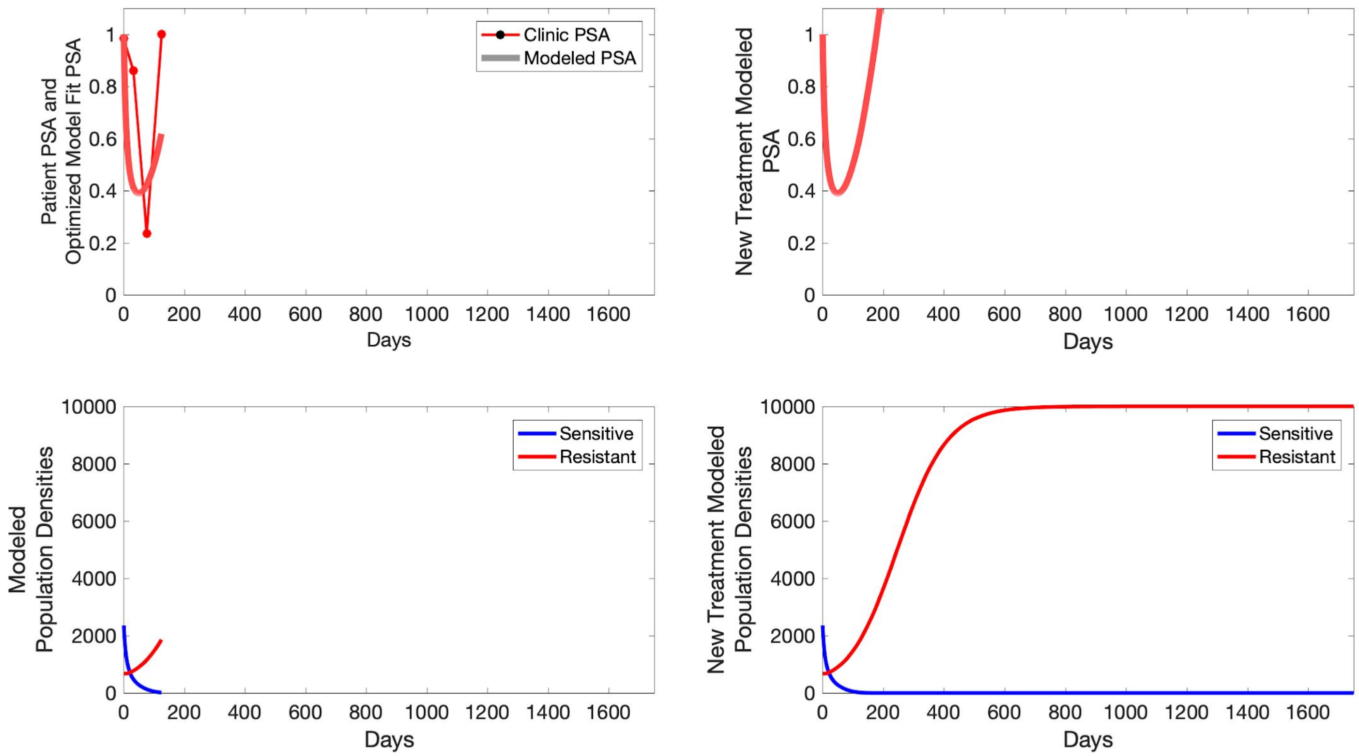

Clinic PSA
Modeled PSA
Patient PSA and Optimized Model Fit PSA
New Treatment Modeled PSA
Days
Sensitive
Resistant
Modeled Population Densities
New Treatment Modeled Population Densities
0
0.2
0.4
0.6
0.8
1
200
400
600
800
1000
1200
1400
1600
2000
4000
6000
8000
10000

**P1001**

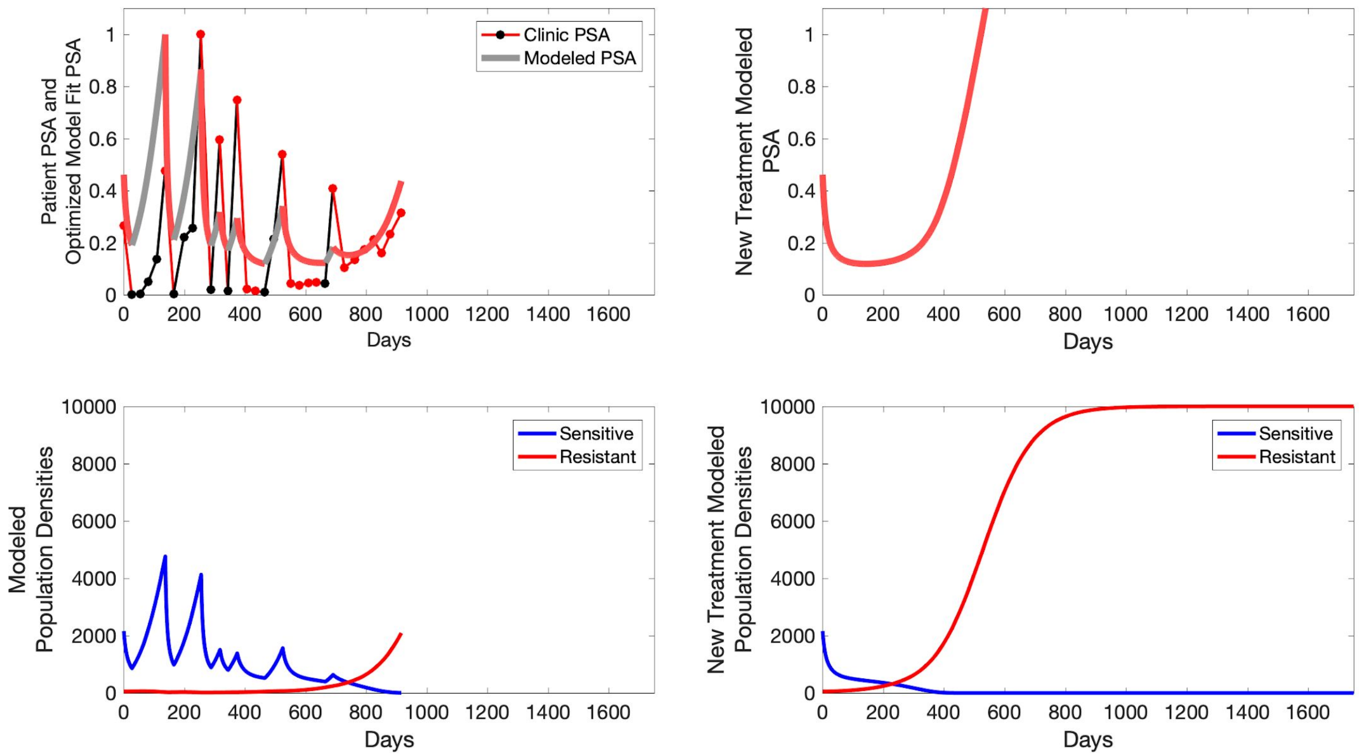

Patient PSA and Optimized Model Fit PSA
Clinic PSA
Modeled PSA
Days
New Treatment Modeled PSA
Days
Modeled Population Densities
Sensitive
Resistant
Days
New Treatment Modeled Population Densities
Sensitive
Resistant
Days

**P1002**

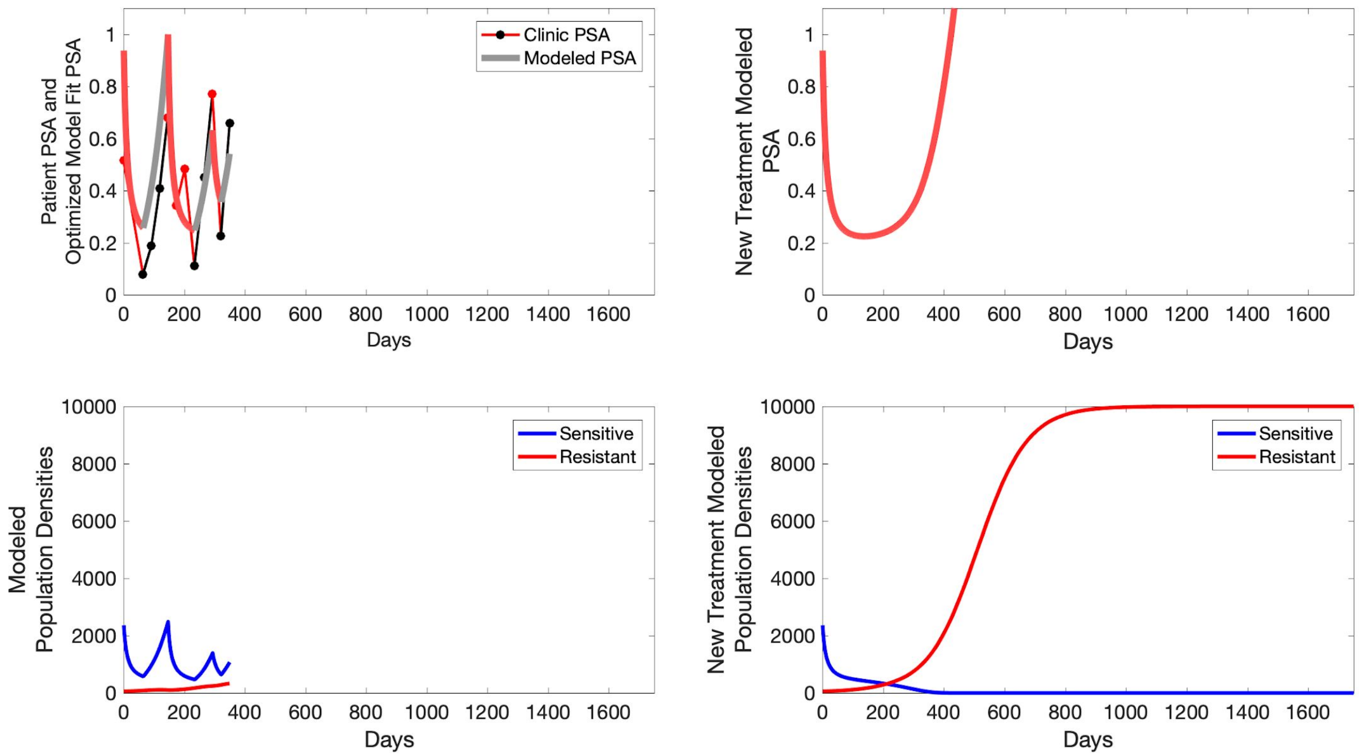
Patient PSA and Optimized Model Fit PSA
Clinic PSA
Modeled PSA
Days
New Treatment Modeled PSA
Days
Modeled Population Densities
Sensitive
Resistant
Days
New Treatment Modeled Population Densities
Sensitive
Resistant
Days

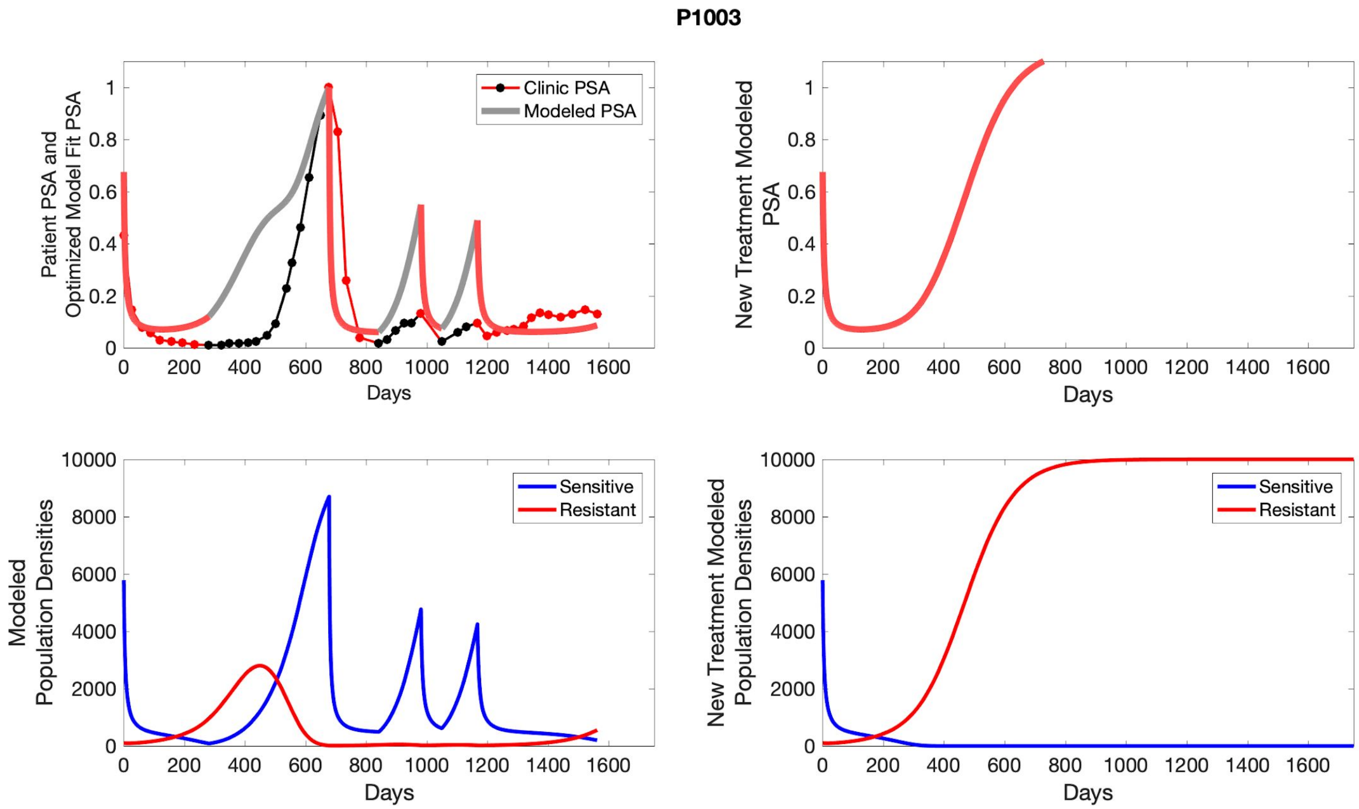
P1003
Clinic PSA
Modeled PSA
Patient PSA and Optimized Model Fit PSA
Days
New Treatment Modeled PSA
Days
Sensitive
Resistant
Modeled Population Densities
Days
Sensitive
Resistant
New Treatment Modeled Population Densities
Days

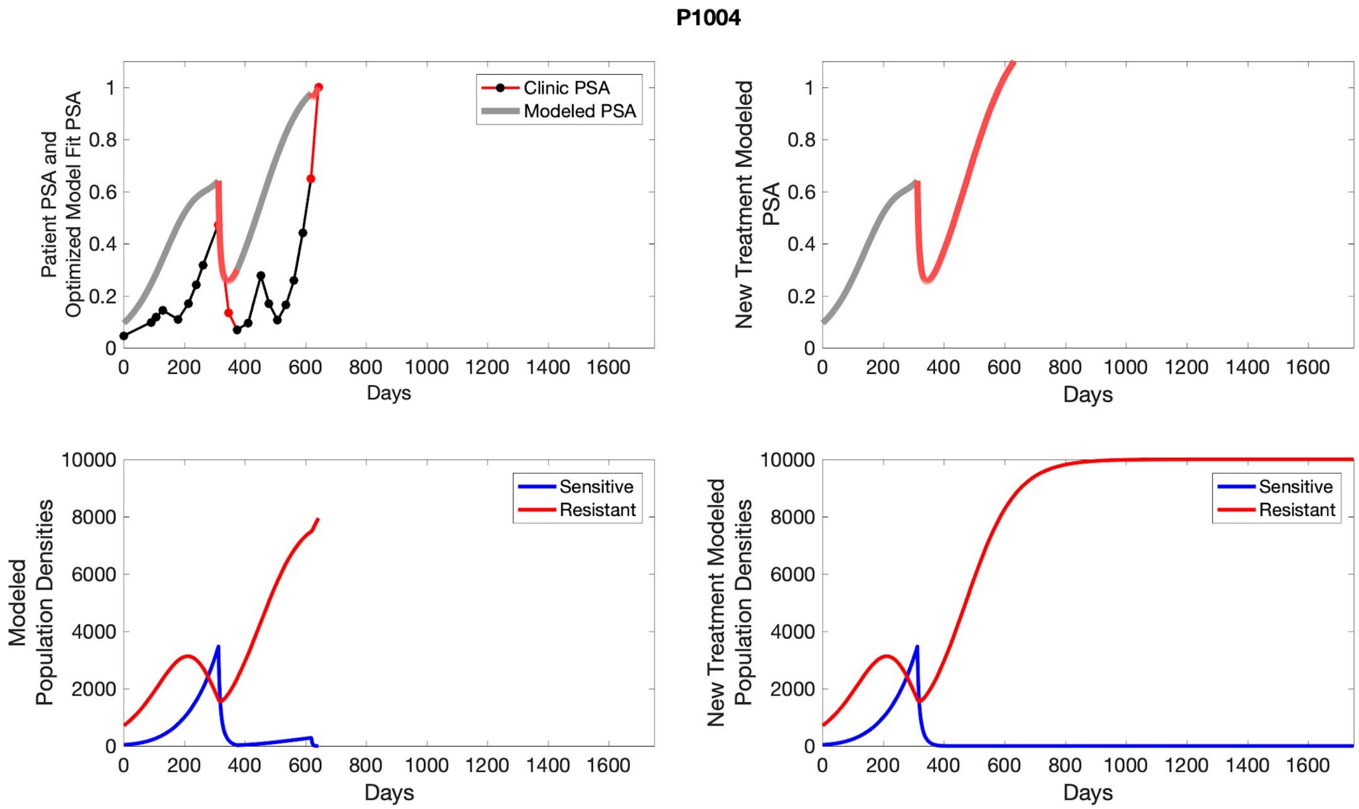

P1004
Clinic PSA
Modeled PSA
Patient PSA and Optimized Model Fit PSA
New Treatment Modeled PSA
Days
Sensitive
Resistant
Modeled Population Densities
New Treatment Modeled Population Densities

**P1005**

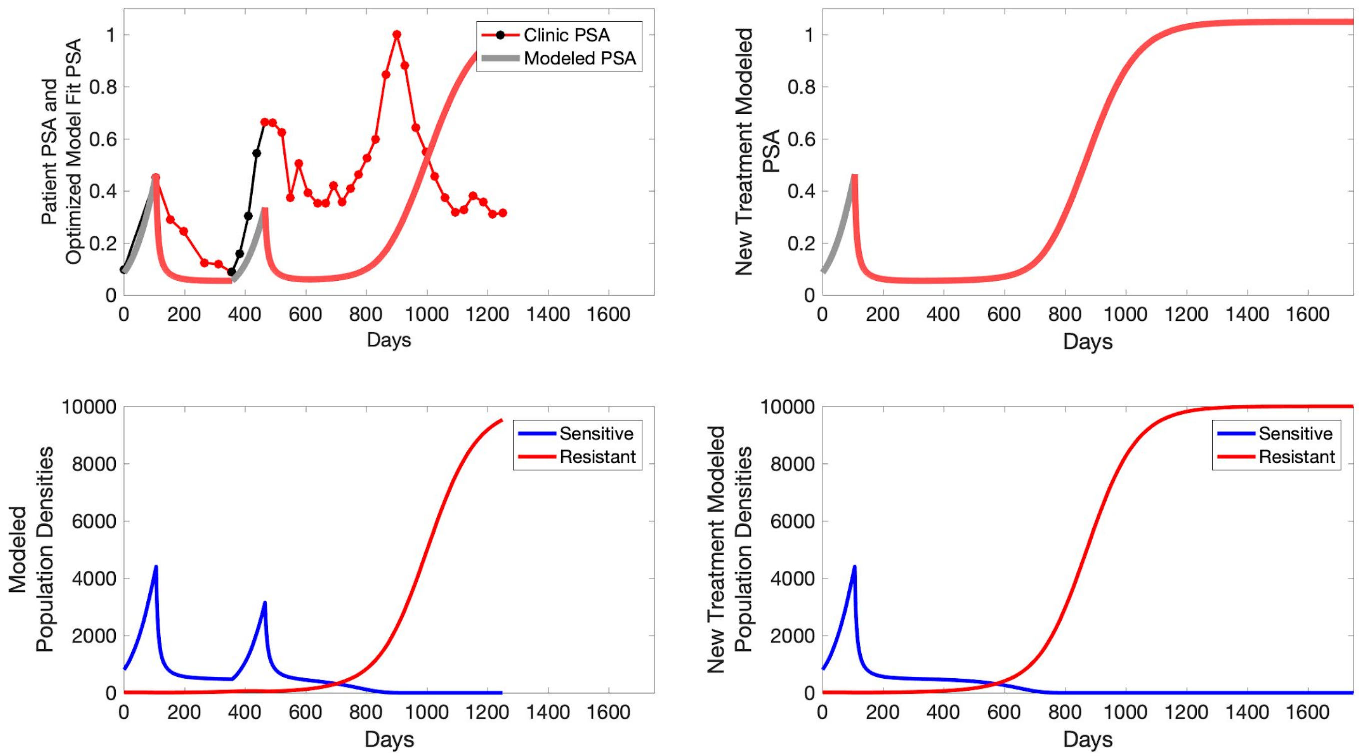

Patient PSA and Optimized Model Fit PSA
Clinic PSA
Modeled PSA
Days
New Treatment Modeled PSA
Days
Modeled Population Densities
Sensitive
Resistant
Days
New Treatment Modeled Population Densities
Sensitive
Resistant
Days

**P1006**

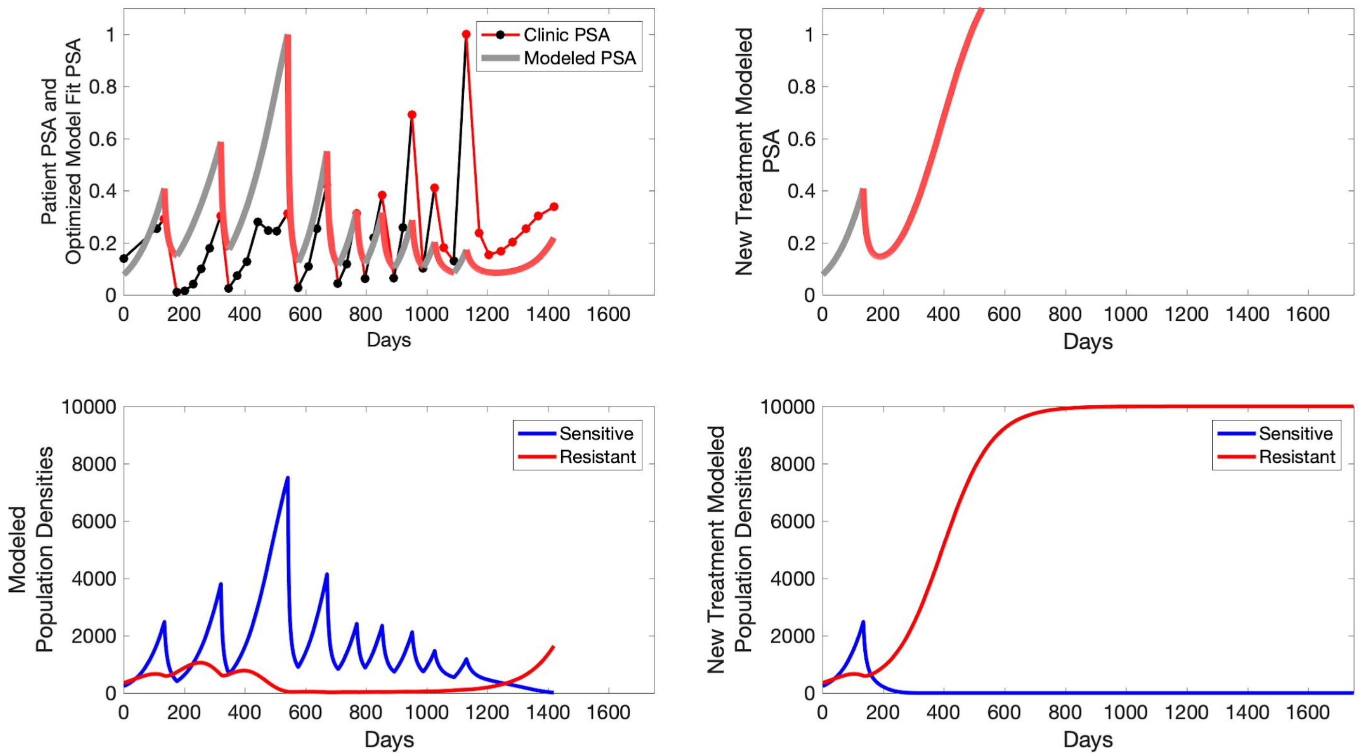

Patient PSA and Optimized Model Fit PSA
Clinic PSA
Modeled PSA
Days
New Treatment Modeled PSA
Days
Modeled Population Densities
Sensitive
Resistant
Days
New Treatment Modeled Population Densities
Sensitive
Resistant
Days

**P1007**

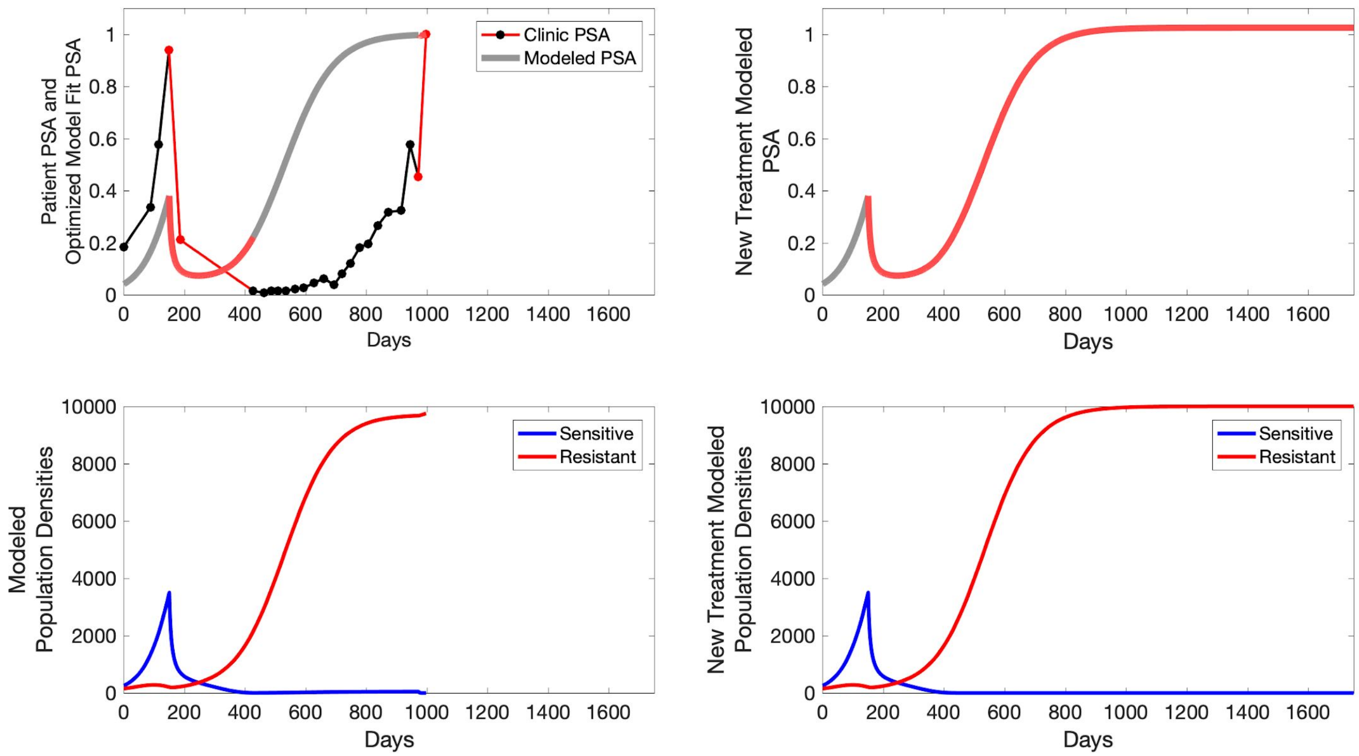

Patient PSA and Optimized Model Fit PSA
Clinic PSA
Modeled PSA
Days
New Treatment Modeled PSA
Days
Modeled Population Densities
Sensitive
Resistant
Days
New Treatment Modeled Population Densities
Sensitive
Resistant
Days

P1009

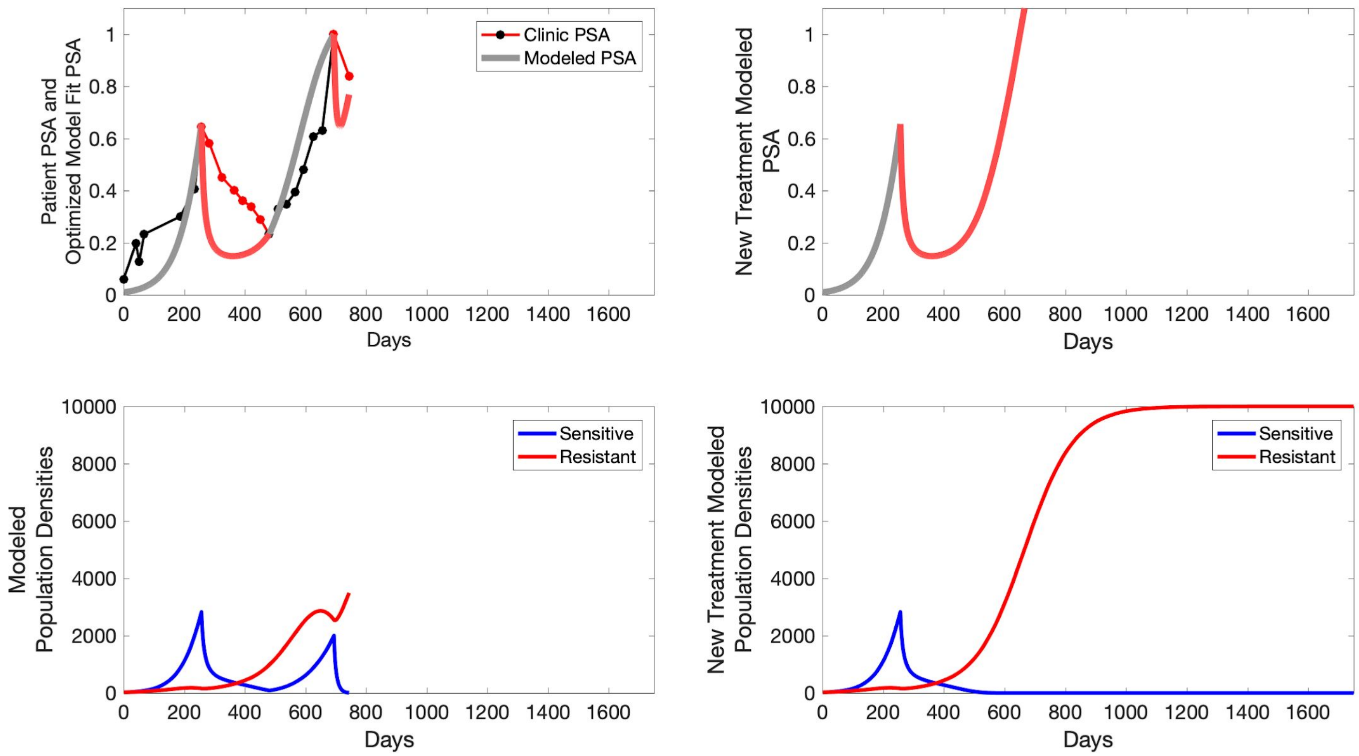
Patient PSA and Optimized Model Fit PSA
Clinic PSA
Modeled PSA
Days
New Treatment Modeled PSA
Days
Modeled Population Densities
Sensitive
Resistant
Days
New Treatment Modeled Population Densities
Sensitive
Resistant
Days

**P1010**

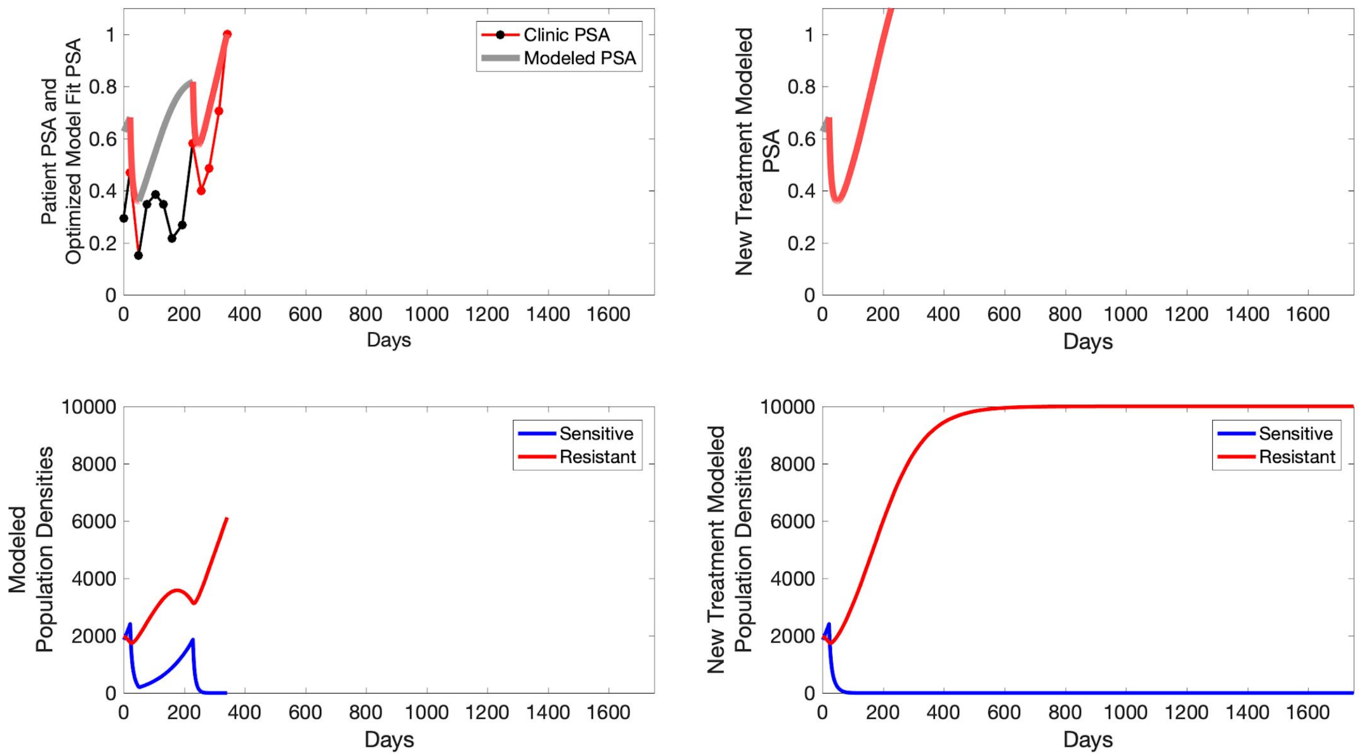

Patient PSA and Optimized Model Fit PSA
Clinic PSA
Modeled PSA
Days
New Treatment Modeled PSA
Days
Modeled Population Densities
Sensitive
Resistant
Days
New Treatment Modeled Population Densities
Sensitive
Resistant
Days

**P1011**

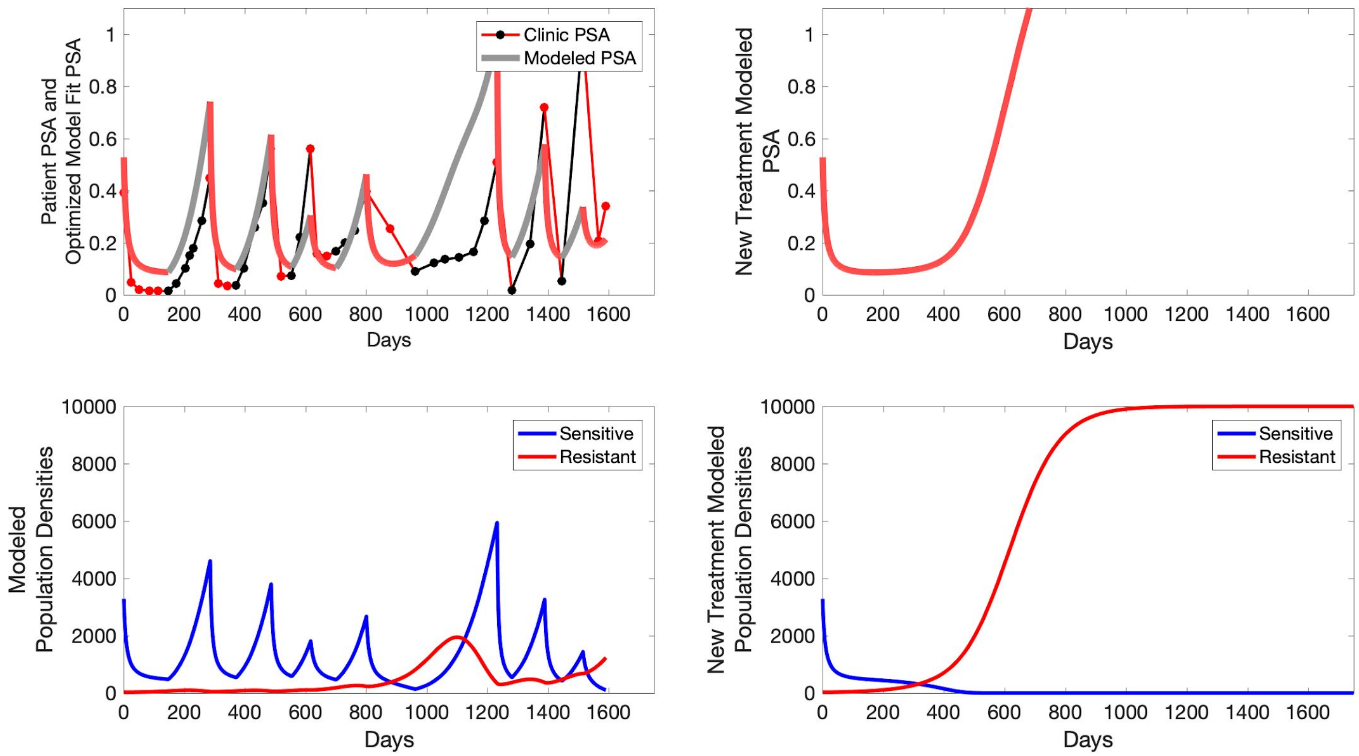

Patient PSA and Optimized Model Fit PSA
Clinic PSA
Modeled PSA
Days
New Treatment Modeled PSA
Days
Modeled Population Densities
Sensitive
Resistant
Days
New Treatment Modeled Population Densities
Sensitive
Resistant
Days

**P1012**

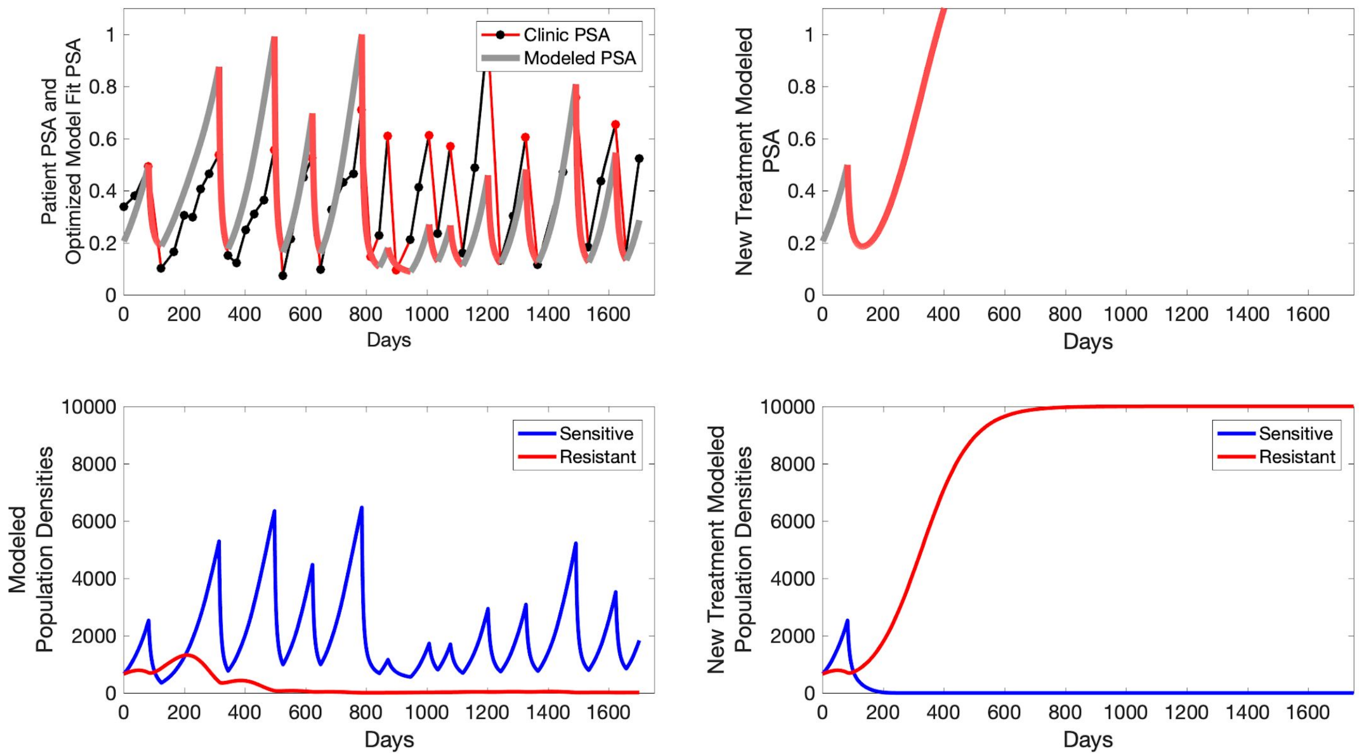
Clinic PSA
Modeled PSA
Patient PSA and Optimized Model Fit PSA
Days
New Treatment Modeled PSA
Days
Sensitive
Resistant
Modeled Population Densities
Days
Sensitive
Resistant
New Treatment Modeled Population Densities
Days

**P1014**

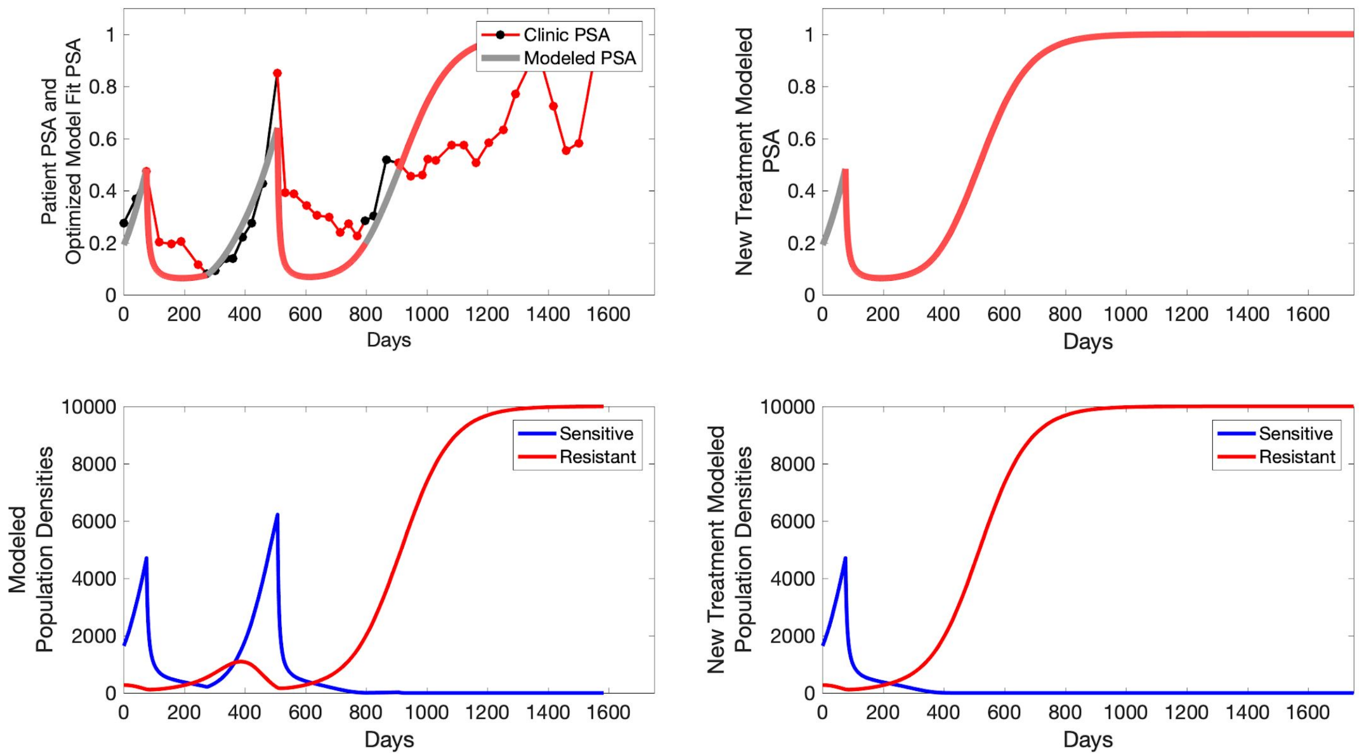

Patient PSA and Optimized Model Fit PSA
Clinic PSA
Modeled PSA
Days
New Treatment Modeled PSA
Days
Modeled Population Densities
Sensitive
Resistant
Days
New Treatment Modeled Population Densities
Sensitive
Resistant
Days

**P1015**

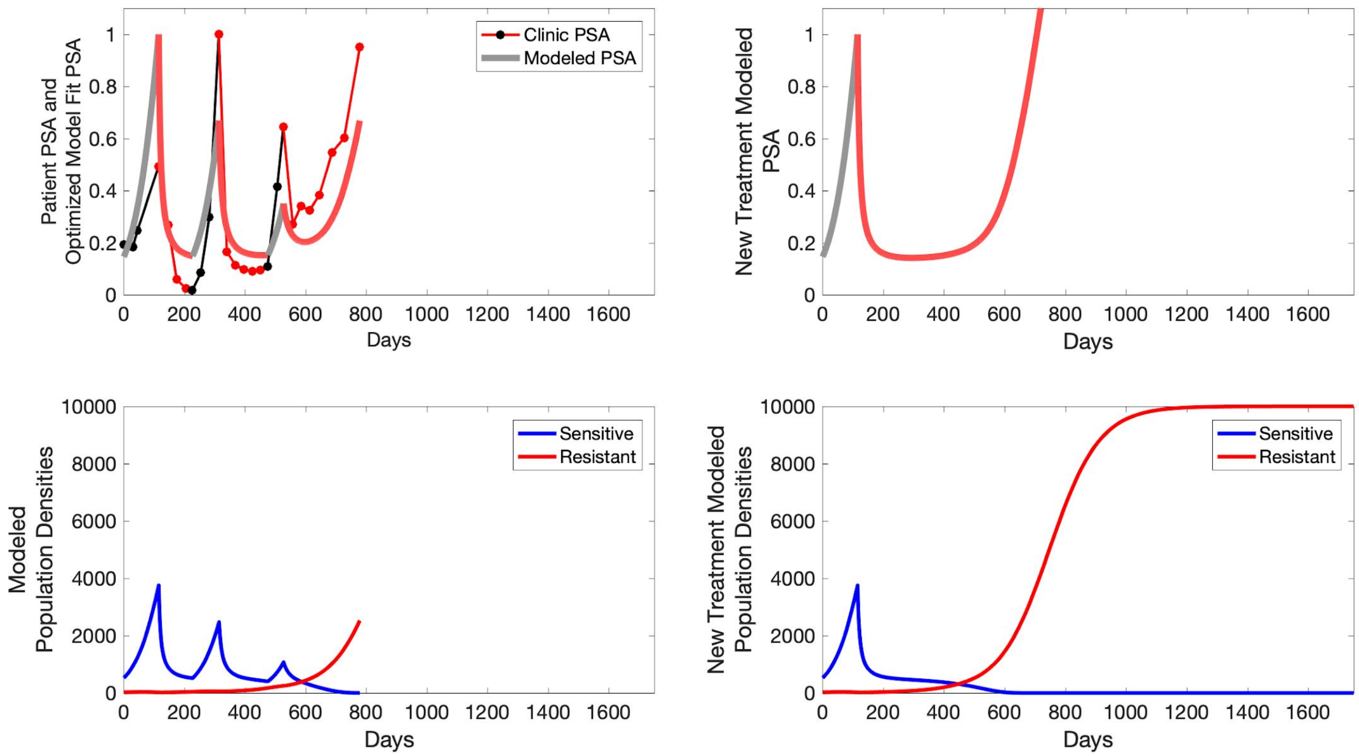

Clinic PSA
Modeled PSA
Patient PSA and Optimized Model Fit PSA
Days
New Treatment Modeled PSA
Sensitive
Resistant
Modeled Population Densities
New Treatment Modeled Population Densities

**P1016**

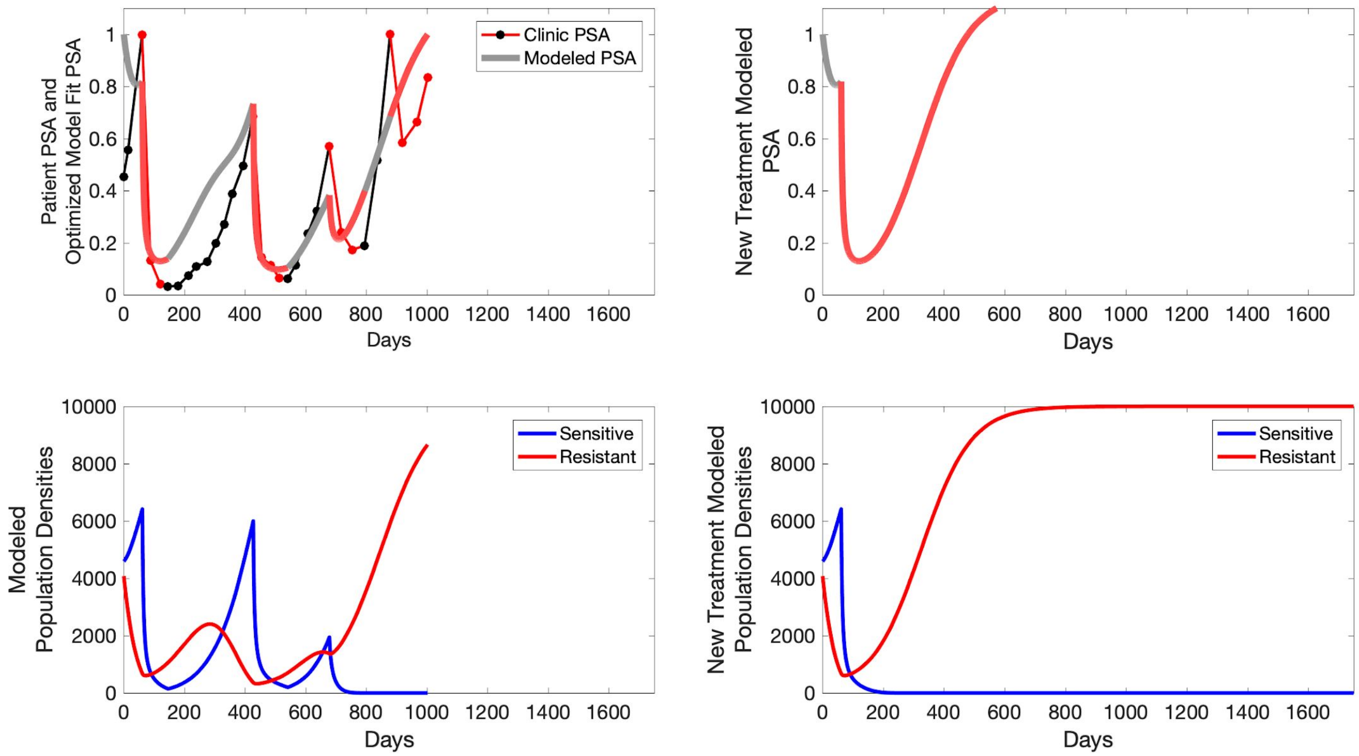

Clinic PSA
Modeled PSA
Patient PSA and Optimized Model Fit PSA
New Treatment Modeled PSA
Days
Sensitive
Resistant
Modeled Population Densities
New Treatment Modeled Population Densities

**P1017**

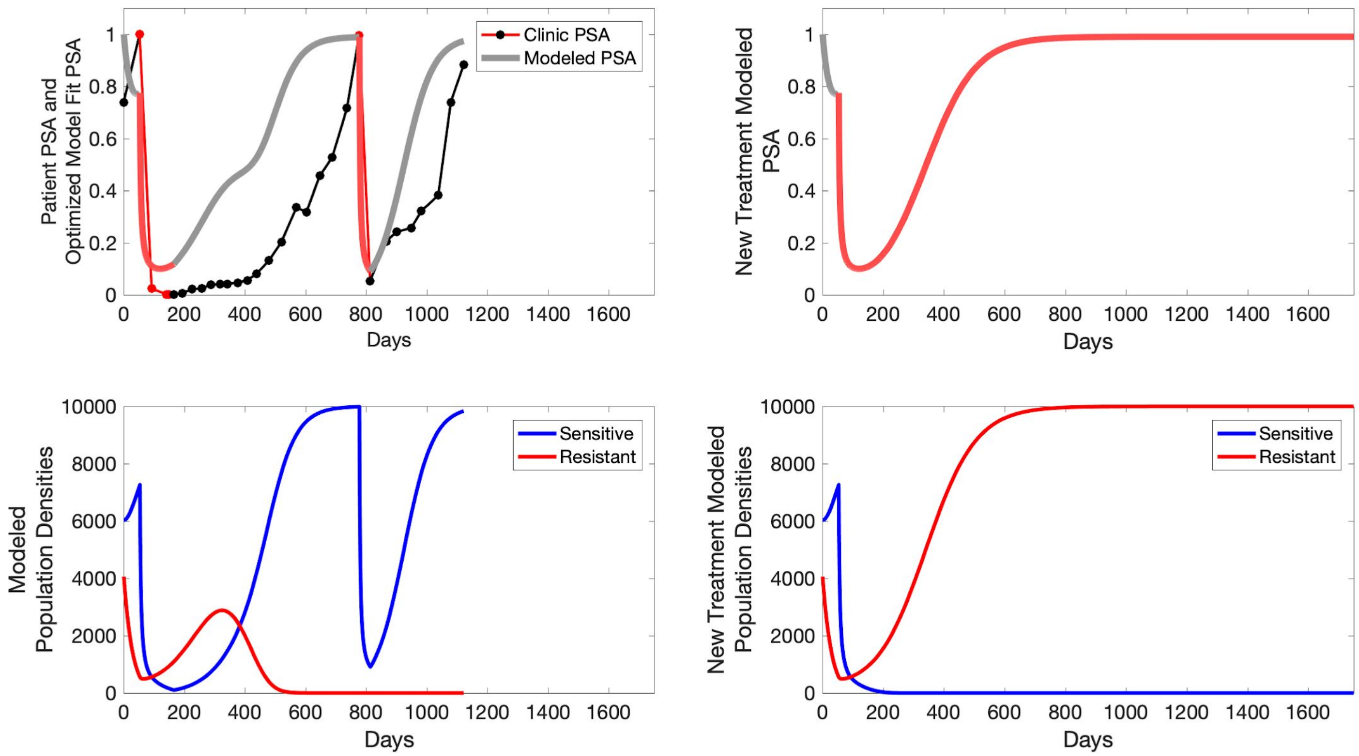

Patient PSA and Optimized Model Fit PSA
Clinic PSA
Modeled PSA
Days
New Treatment Modeled PSA
Days
Modeled Population Densities
Sensitive
Resistant
Days
New Treatment Modeled Population Densities
Sensitive
Resistant
Days

**P1018**

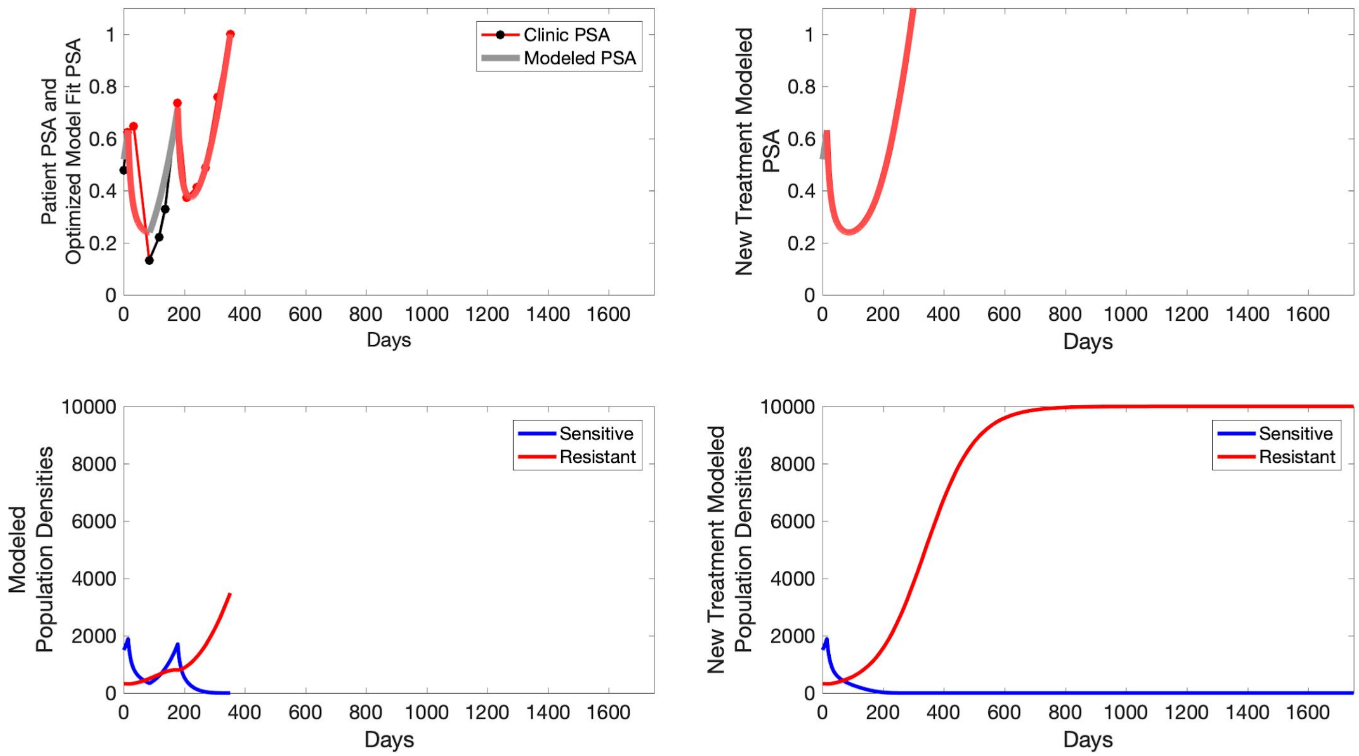
Clinic PSA
Modeled PSA
Patient PSA and Optimized Model Fit PSA
New Treatment Modeled PSA
Days
Sensitive
Resistant
Modeled Population Densities
New Treatment Modeled Population Densities

**P1020**

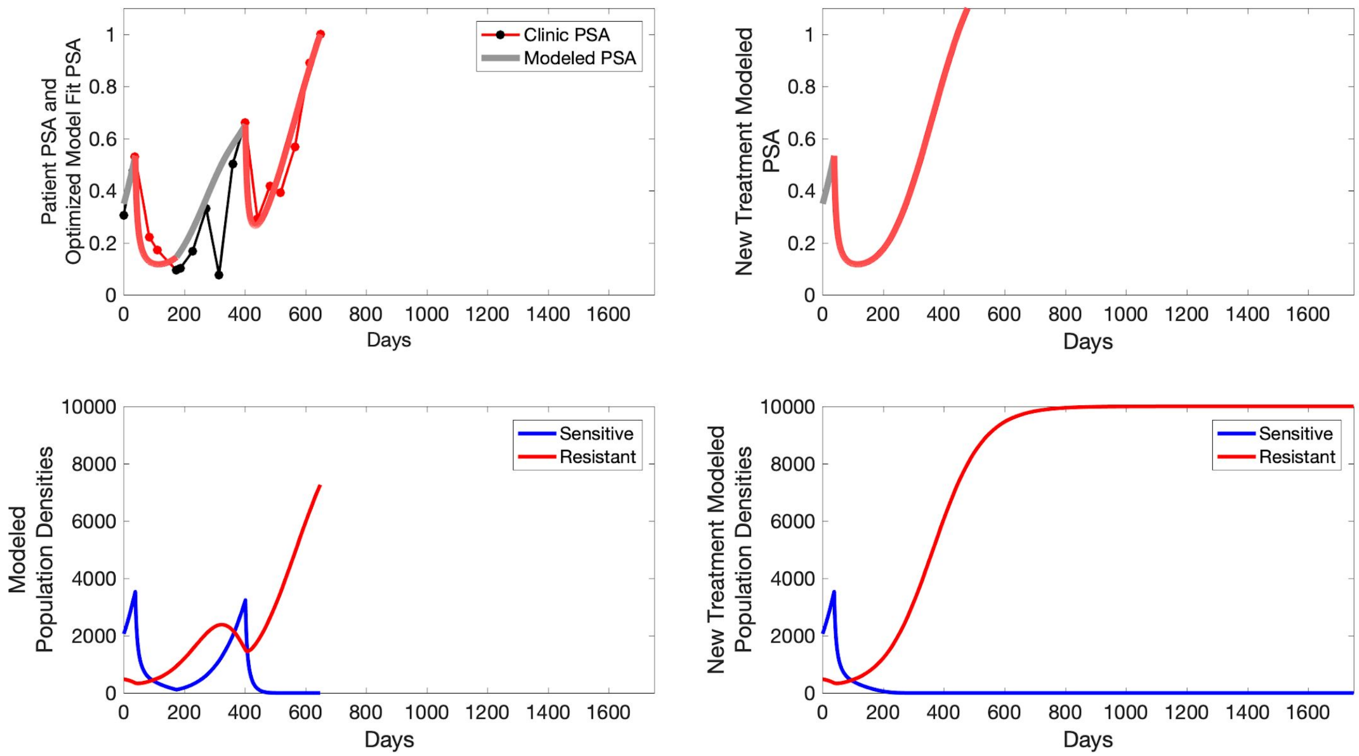

Patient PSA and Optimized Model Fit PSA
Clinic PSA
Modeled PSA
Days
New Treatment Modeled PSA
Days
Modeled Population Densities
Sensitive
Resistant
Days
New Treatment Modeled Population Densities
Sensitive
Resistant
Days

#### S7 Simulated Intermittent Therapy

To evaluate an intermittent therapy (one that had not been used as a trial arm), we ran a simulation for each patient (both cohorts) with a treatment protocol that started with an 8 month (243 day) induction period beginning at the time abiraterone was first administered clinically to each patient, which was not always at t=0. Following the induction period, abiraterone was then discontinued until either another 243 days had passed, or until the patient's PSA returned to the patient's initial baseline PSA level. If abiraterone is reinstated, it remains for 243 days. For each patient, we initiated model runs using the patient specific parameters from Table S4.

**C001**

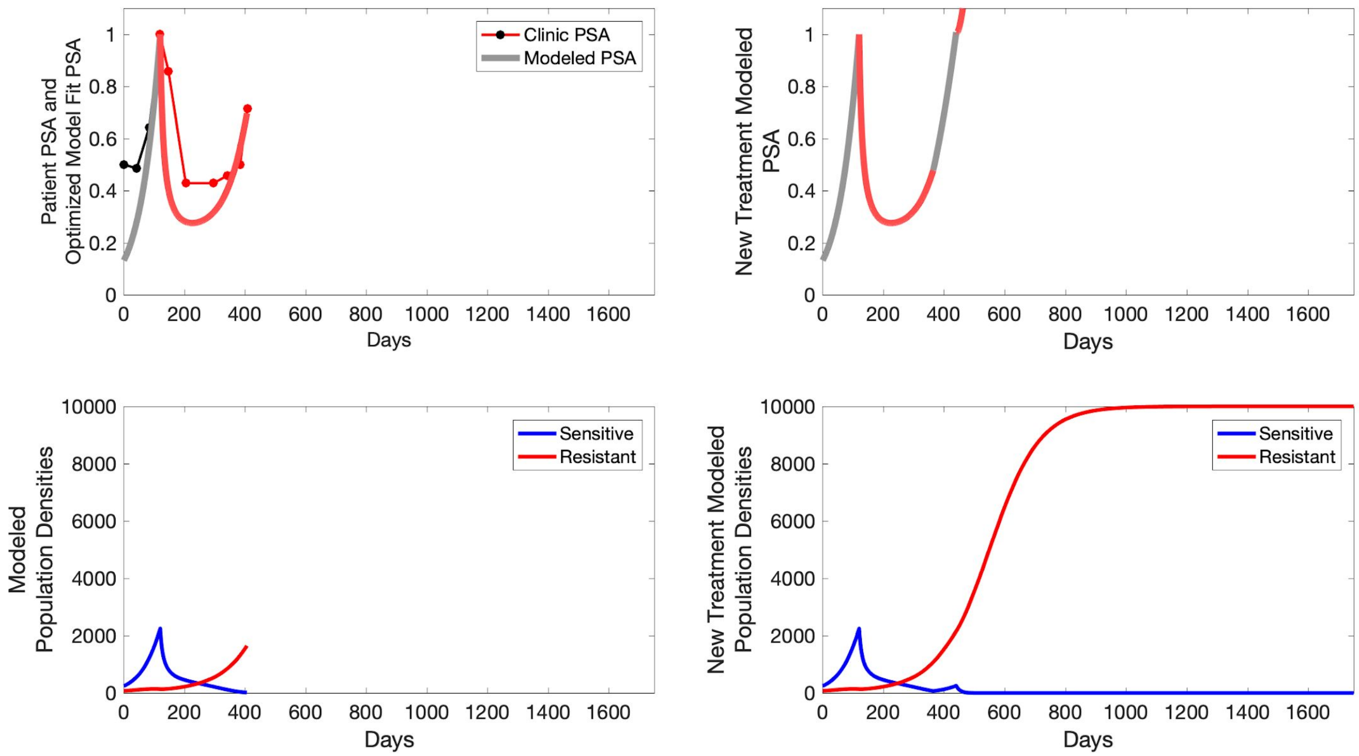

Clinic PSA
Modeled PSA
Patient PSA and Optimized Model Fit PSA
New Treatment Modeled PSA
Days
Sensitive
Resistant
Modeled Population Densities
New Treatment Modeled Population Densities

**C002**

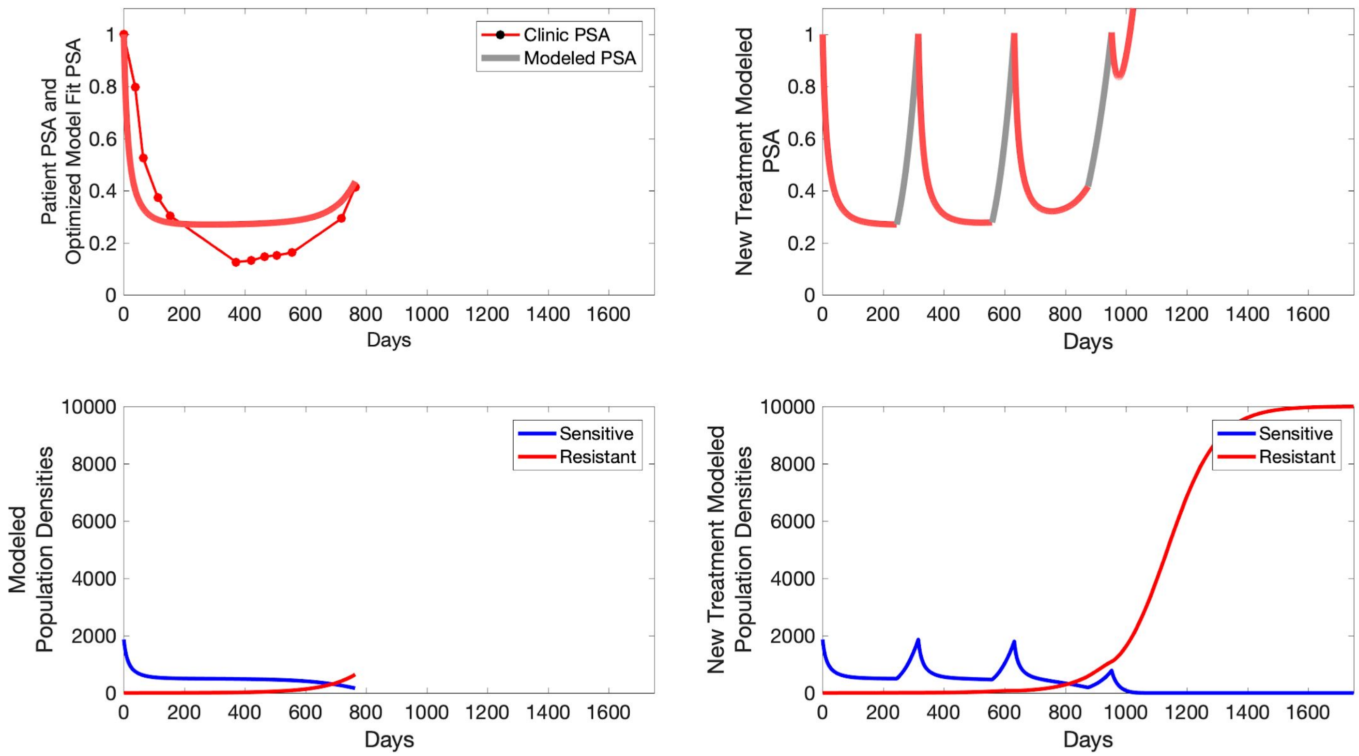
Patient PSA and
Optimized Model Fit PSA
Clinic PSA
Modeled PSA
Days
New Treatment Modeled
PSA
Days
Modeled
Population Densities
Sensitive
Resistant
Days
New Treatment Modeled
Population Densities
Sensitive
Resistant
Days

**C003**

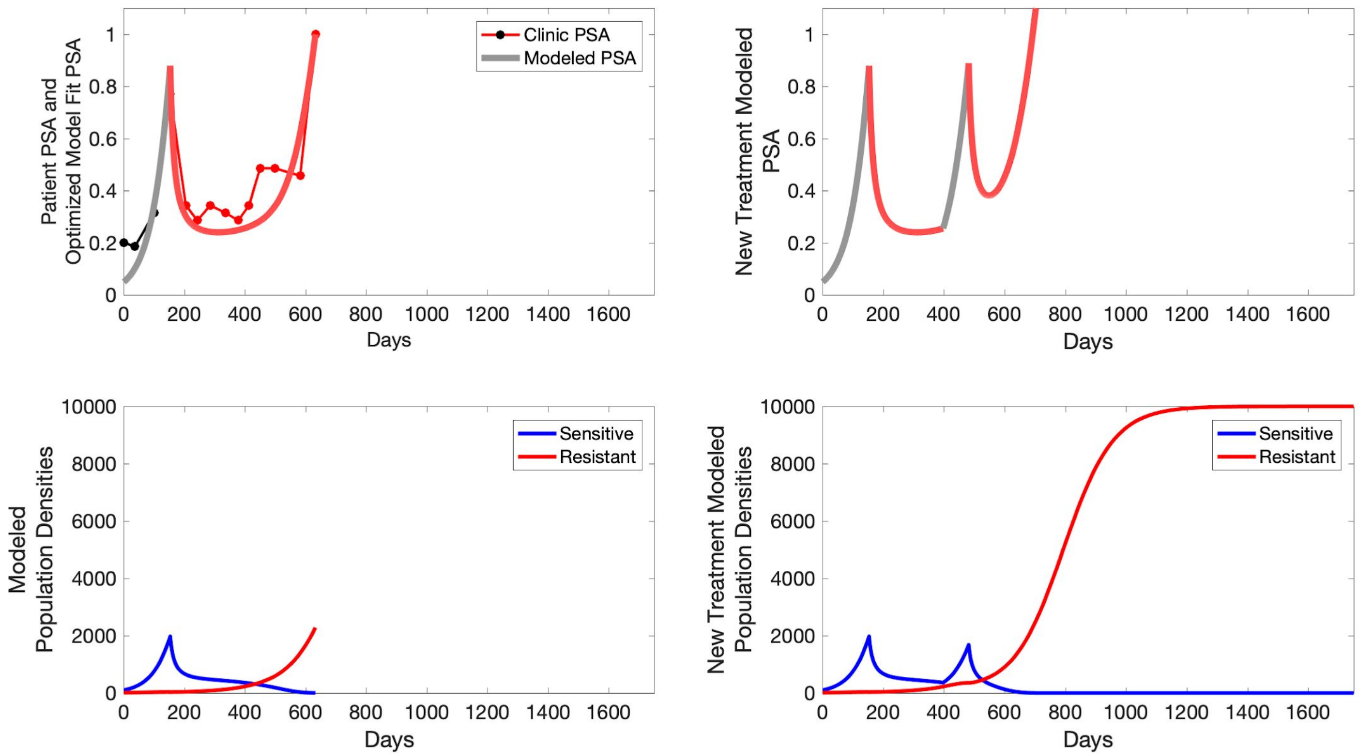

Patient PSA and Optimized Model Fit PSA
Clinic PSA
Modeled PSA
Days
New Treatment Modeled PSA
Days
Modeled Population Densities
Sensitive
Resistant
Days
New Treatment Modeled Population Densities
Sensitive
Resistant
Days

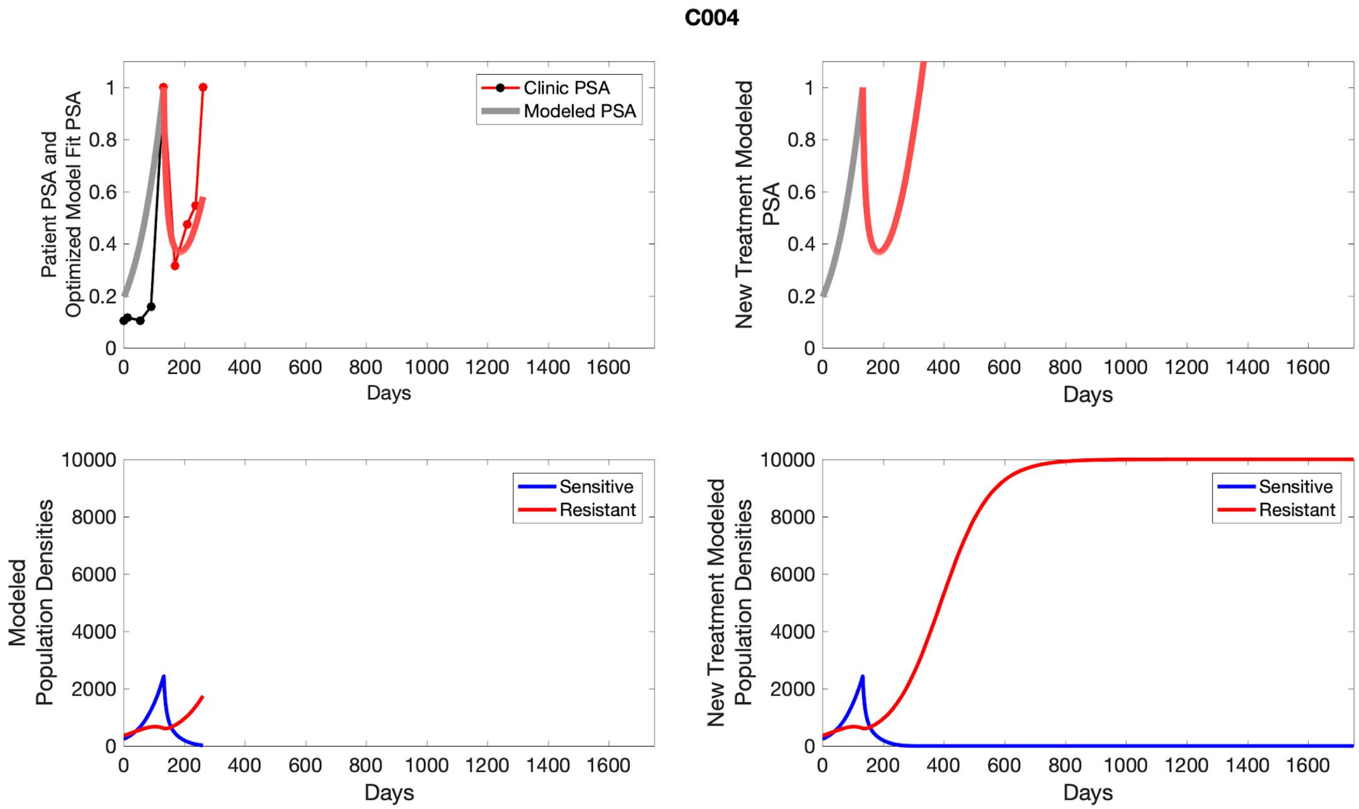
C004
Patient PSA and Optimized Model Fit PSA
Clinic PSA
Modeled PSA
Days
New Treatment Modeled PSA
Modeled Population Densities
Sensitive
Resistant
New Treatment Modeled Population Densities

**C005**

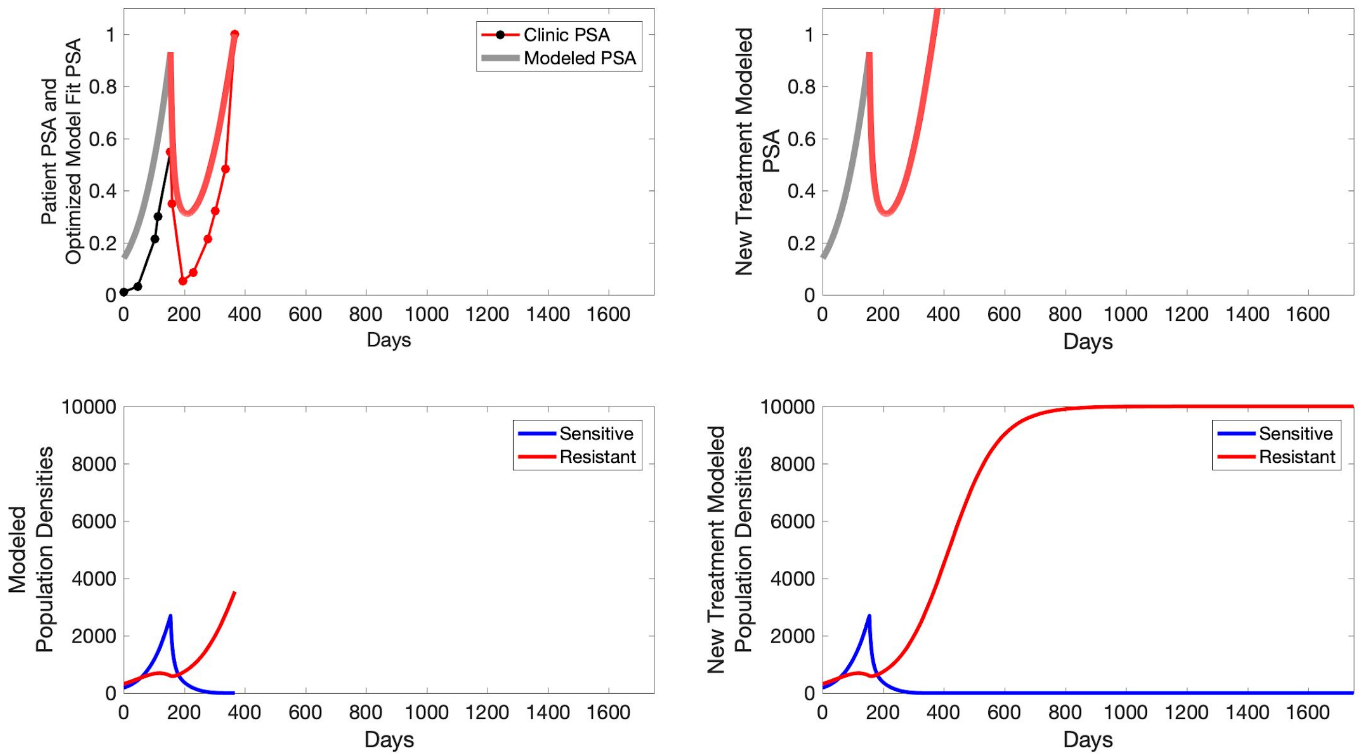

Clinic PSA
Modeled PSA
Patient PSA and Optimized Model Fit PSA
Days
New Treatment Modeled PSA
Days
Sensitive
Resistant
Modeled Population Densities
Days
New Treatment Modeled Population Densities
Days

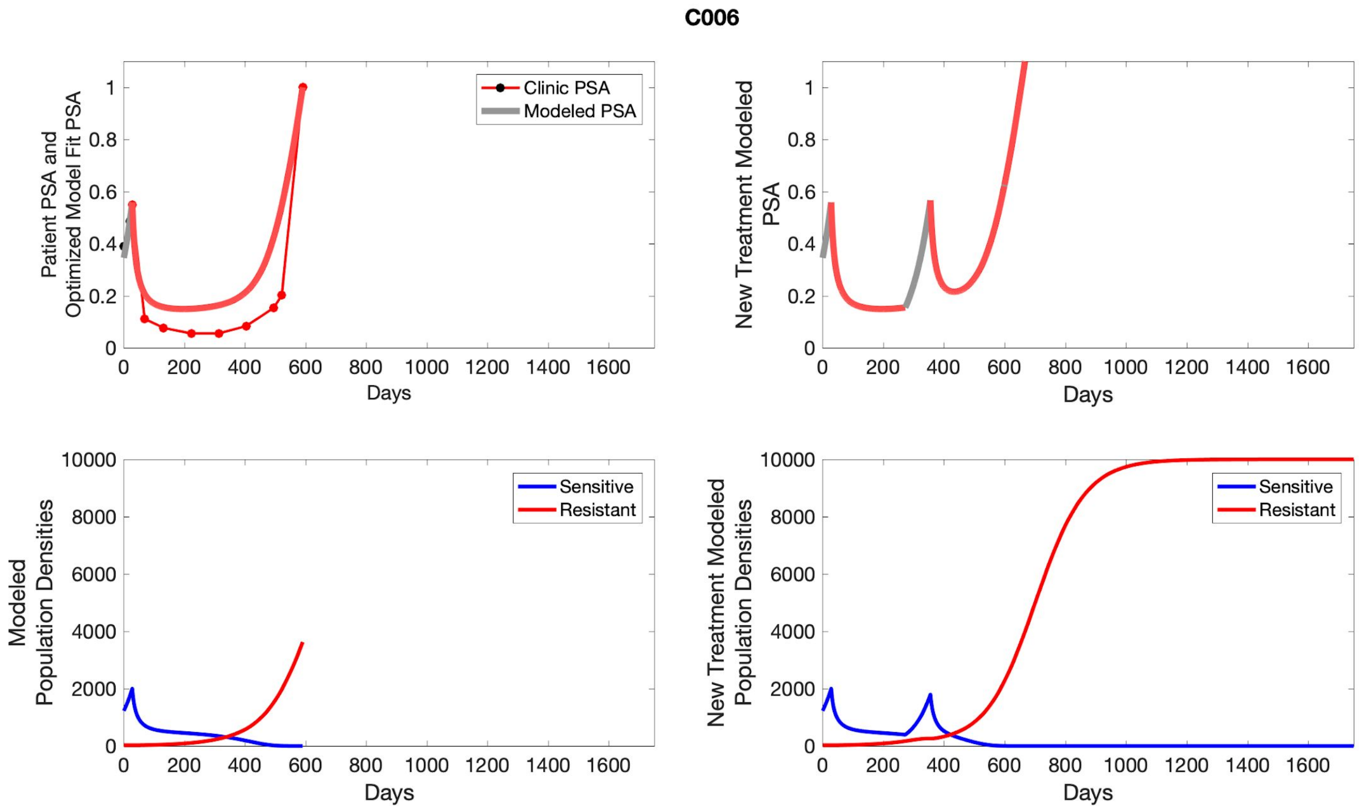
C006
Clinic PSA
Modeled PSA
Patient PSA and Optimized Model Fit PSA
New Treatment Modeled PSA
Days
Sensitive
Resistant
Modeled Population Densities
New Treatment Modeled Population Densities

## C007

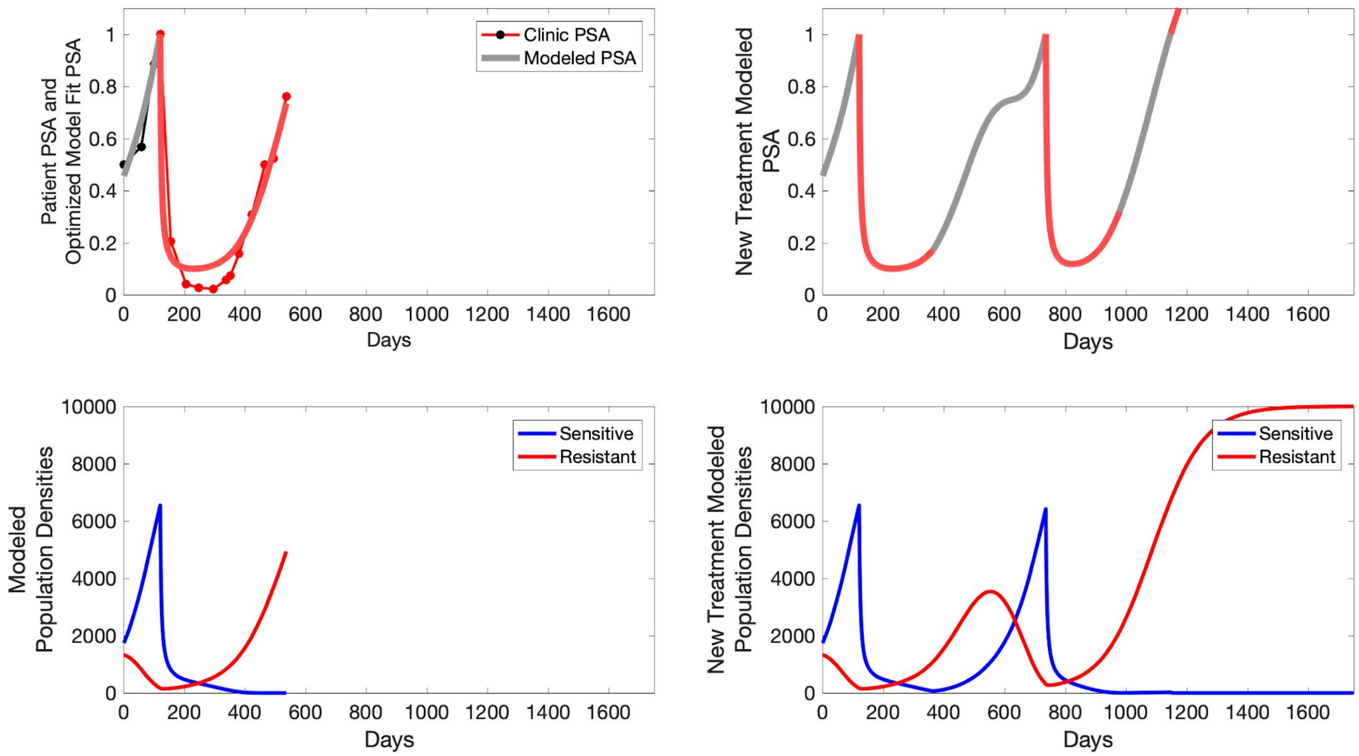
Patient PSA and Optimized Model Fit PSA
Clinic PSA
Modeled PSA
Days
New Treatment Modeled PSA
Days
Modeled Population Densities
Sensitive
Resistant
Days
New Treatment Modeled Population Densities
Sensitive
Resistant
Days

**C008**

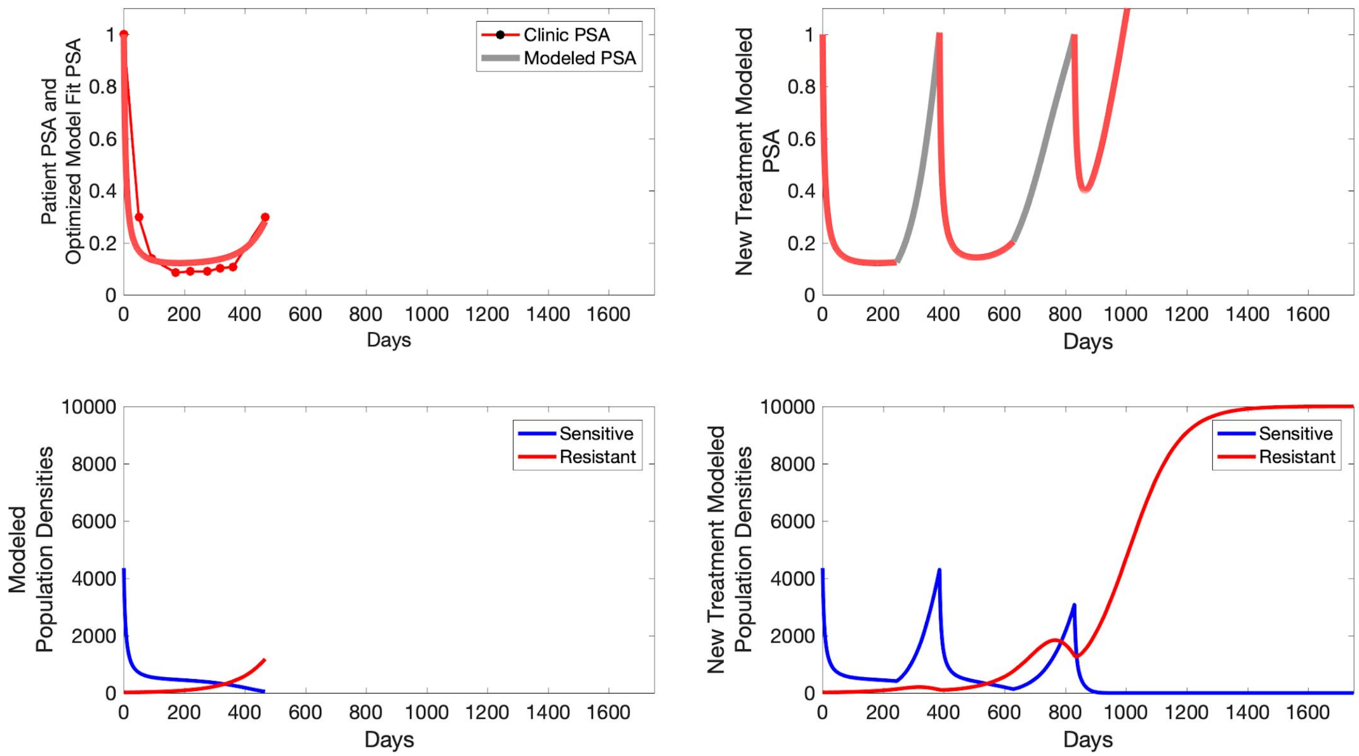
Patient PSA and Optimized Model Fit PSA
Clinic PSA
Modeled PSA
Days
New Treatment Modeled PSA
Days
Modeled Population Densities
Sensitive
Resistant
Days
New Treatment Modeled Population Densities
Sensitive
Resistant
Days

**C009**

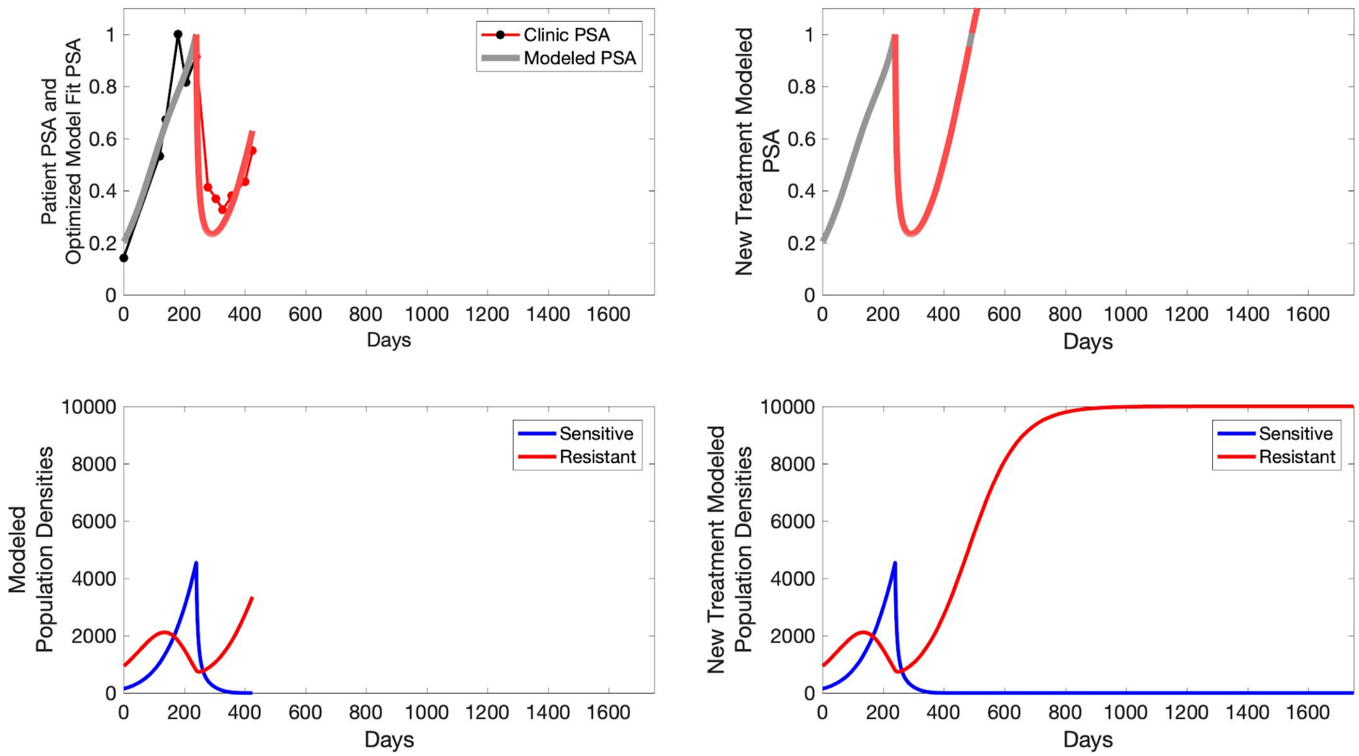

Patient PSA and Optimized Model Fit PSA
Clinic PSA
Modeled PSA
Days
New Treatment Modeled PSA
Days
Modeled Population Densities
Sensitive
Resistant
Days
New Treatment Modeled Population Densities
Sensitive
Resistant
Days

**C010**

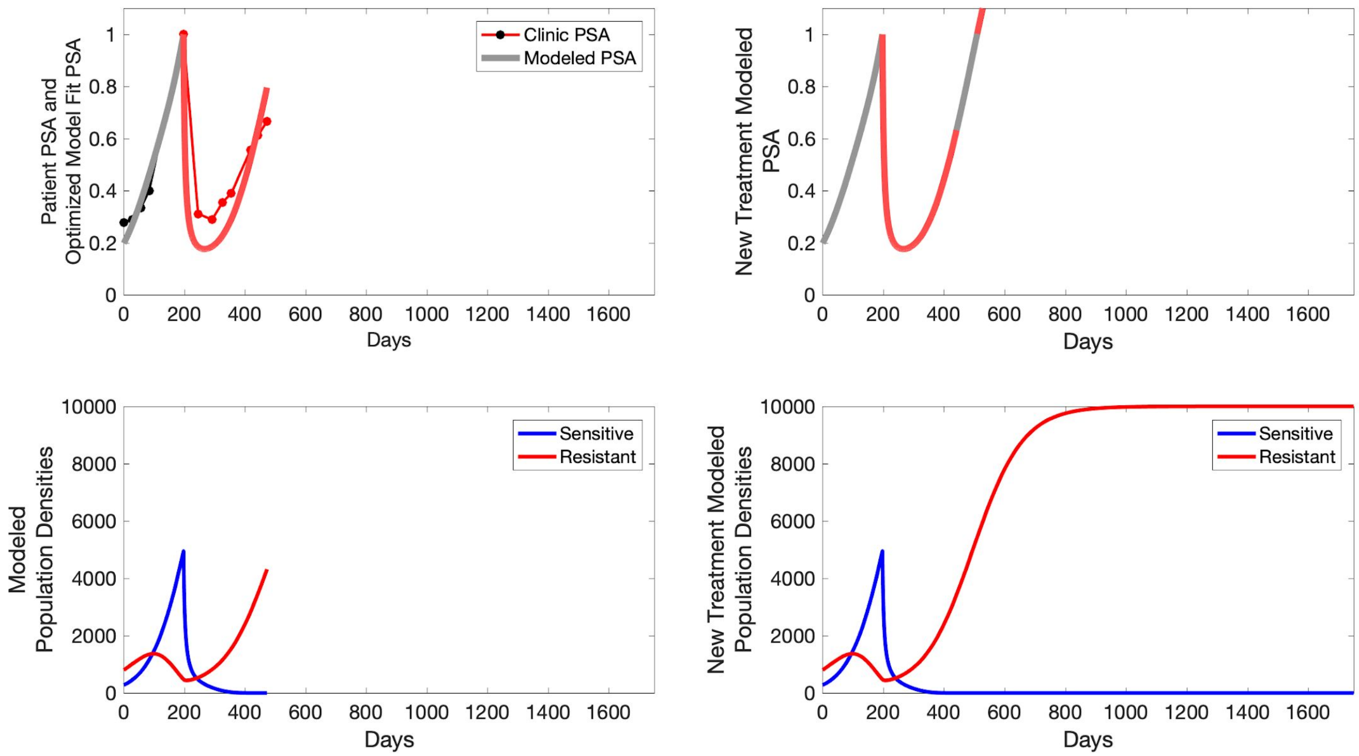

Clinic PSA
Modeled PSA
Patient PSA and Optimized Model Fit PSA
New Treatment Modeled PSA
Days
Sensitive
Resistant
Modeled Population Densities
New Treatment Modeled Population Densities

**C011**

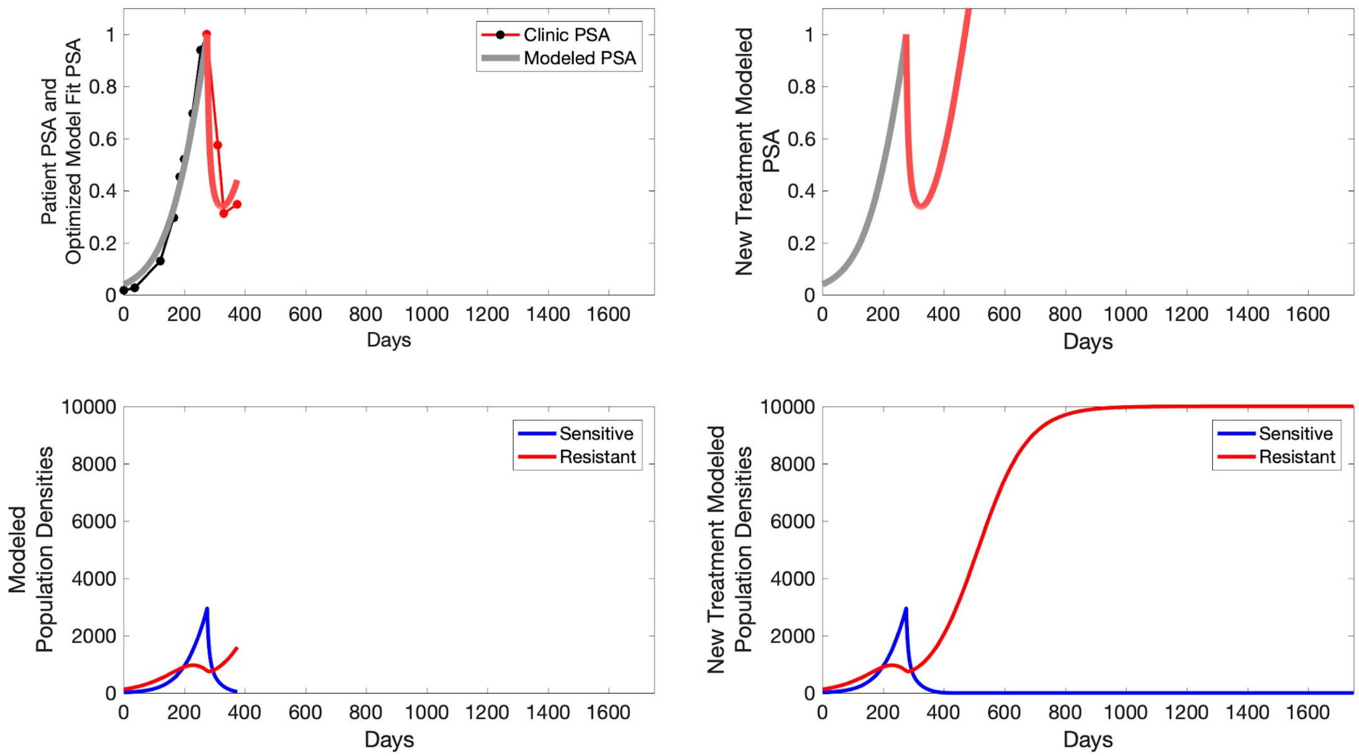

Patient PSA and Optimized Model Fit PSA
Clinic PSA
Modeled PSA
Days
New Treatment Modeled PSA
Days
Modeled Population Densities
Sensitive
Resistant
Days
New Treatment Modeled Population Densities
Sensitive
Resistant
Days

**C012**

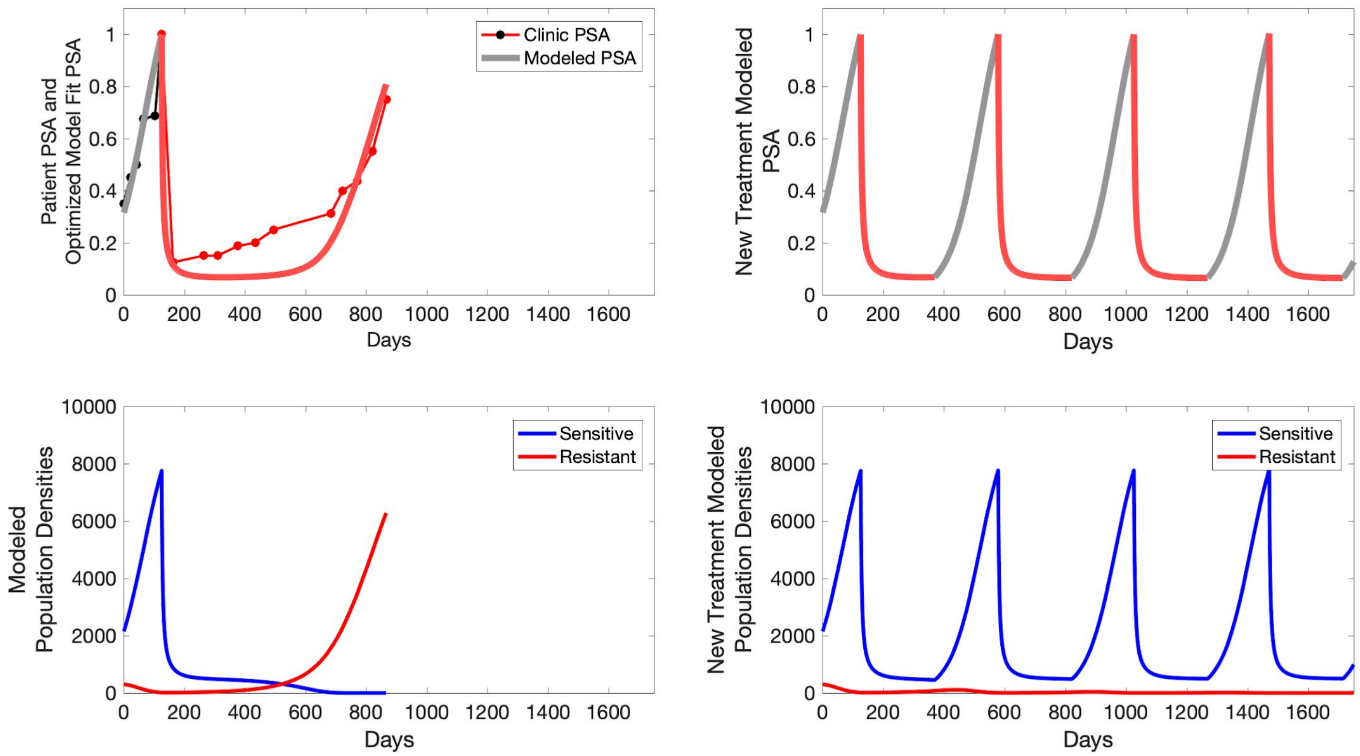

Patient PSA and Optimized Model Fit PSA
Clinic PSA
Modeled PSA
Days
New Treatment Modeled PSA
Days
Modeled Population Densities
Sensitive
Resistant
Days
New Treatment Modeled Population Densities
Sensitive
Resistant
Days

**C013**

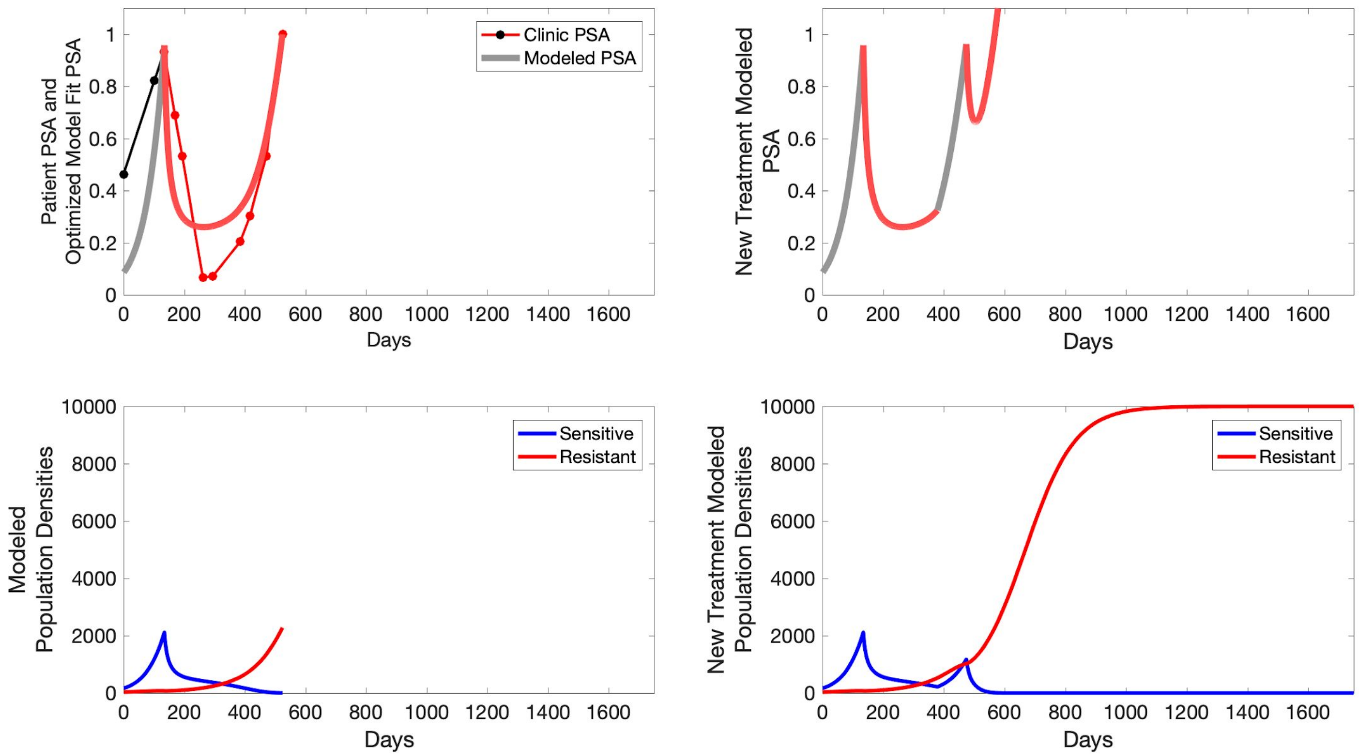
Clinic PSA
Modeled PSA
Patient PSA and Optimized Model Fit PSA
New Treatment Modeled PSA
Days
Sensitive
Resistant
Modeled Population Densities
New Treatment Modeled Population Densities

**C014**

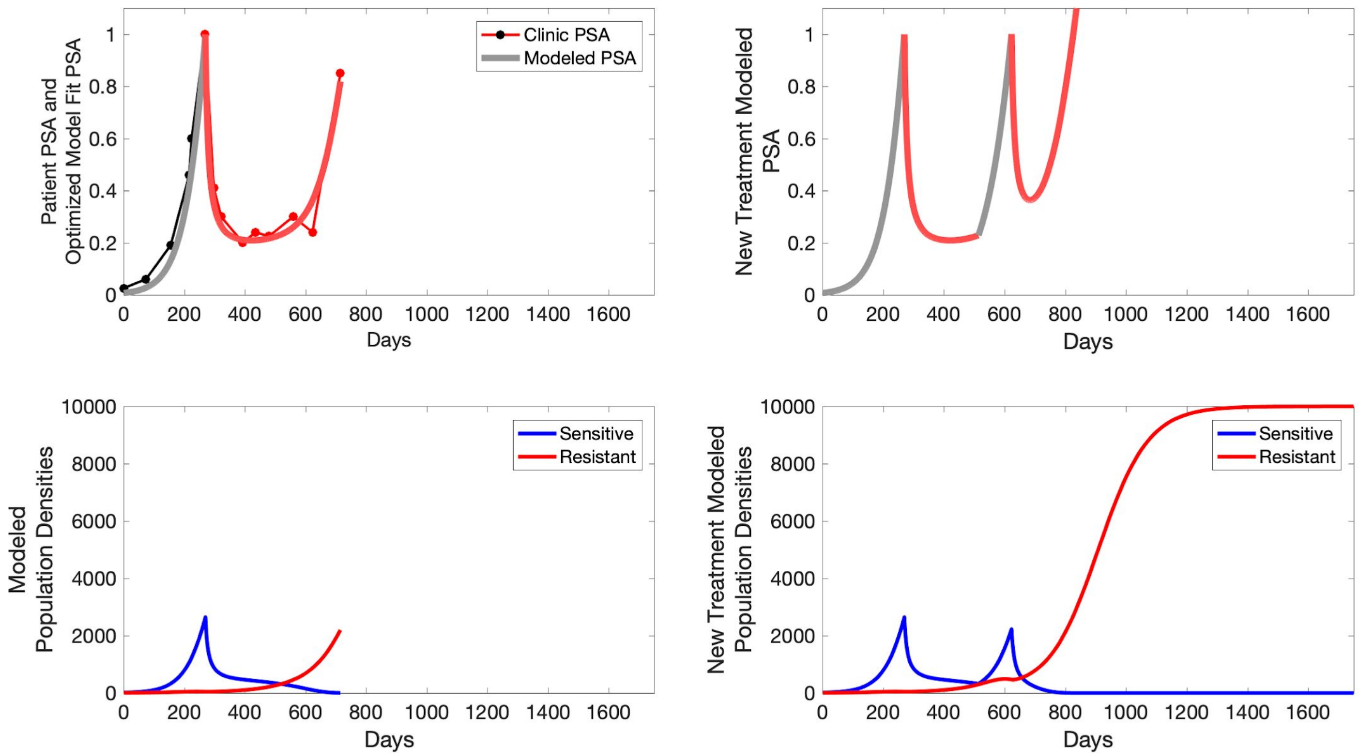

Patient PSA and Optimized Model Fit PSA
Clinic PSA
Modeled PSA
Days
New Treatment Modeled PSA
Days
Modeled Population Densities
Sensitive
Resistant
Days
New Treatment Modeled Population Densities
Sensitive
Resistant
Days

**C015**

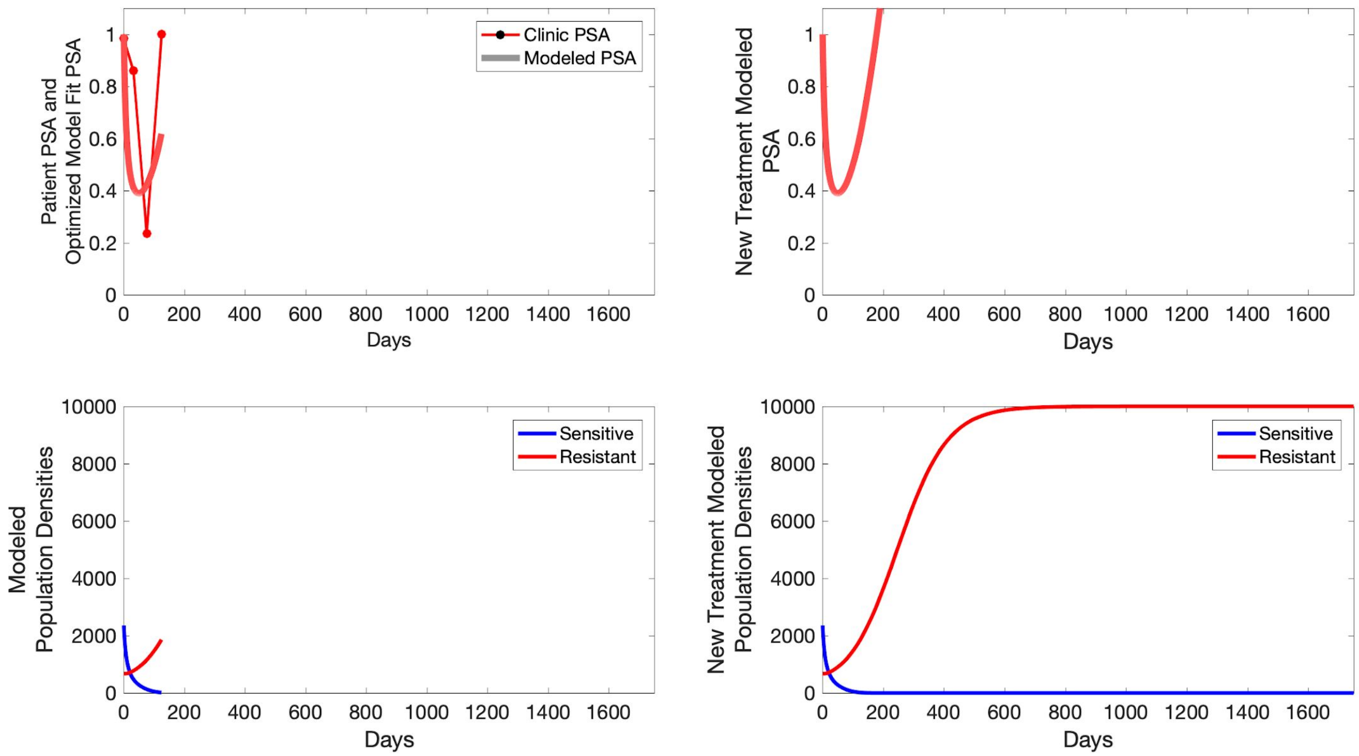
Patient PSA and Optimized Model Fit PSA
Clinic PSA
Modeled PSA
Days
New Treatment Modeled PSA
Days
Modeled Population Densities
Sensitive
Resistant
Days
New Treatment Modeled Population Densities
Sensitive
Resistant
Days

**P1001**

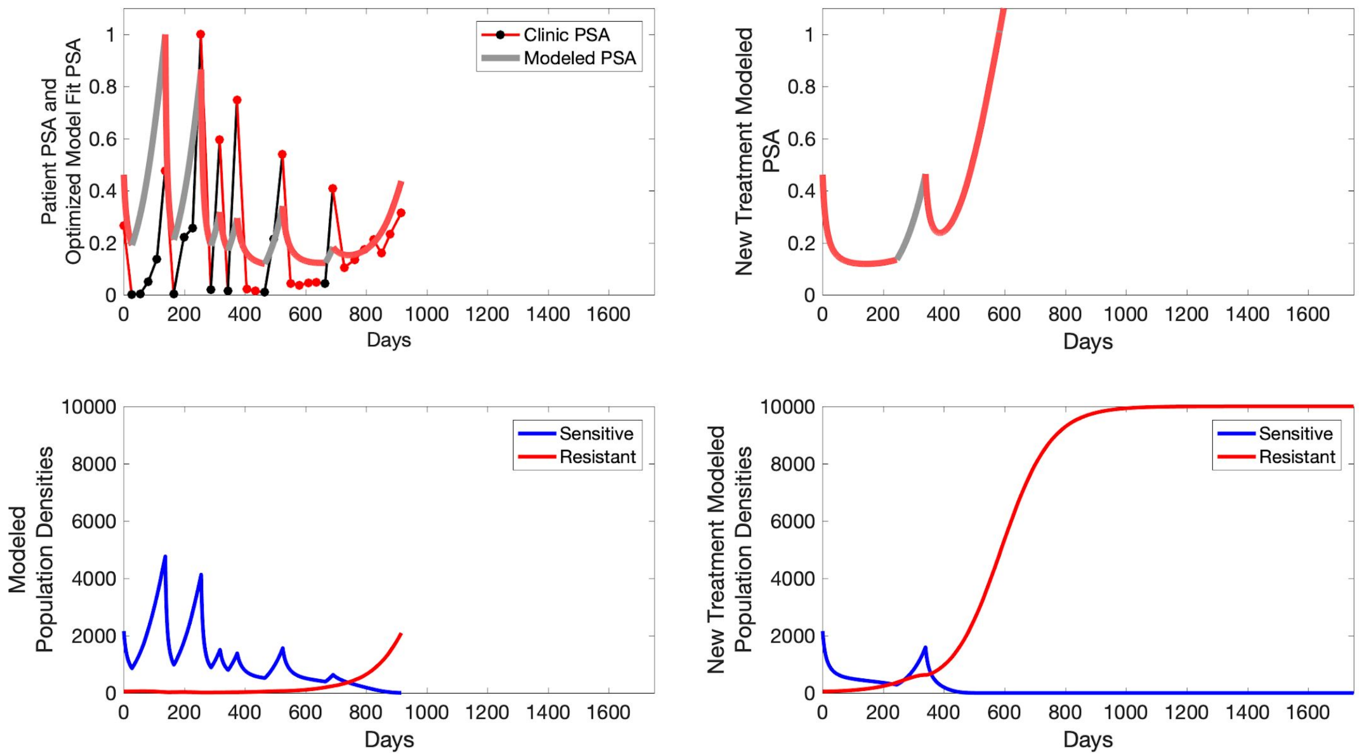

Clinic PSA
Modeled PSA
Patient PSA and Optimized Model Fit PSA
New Treatment Modeled PSA
Days
Sensitive
Resistant
Modeled Population Densities
New Treatment Modeled Population Densities

**P1002**

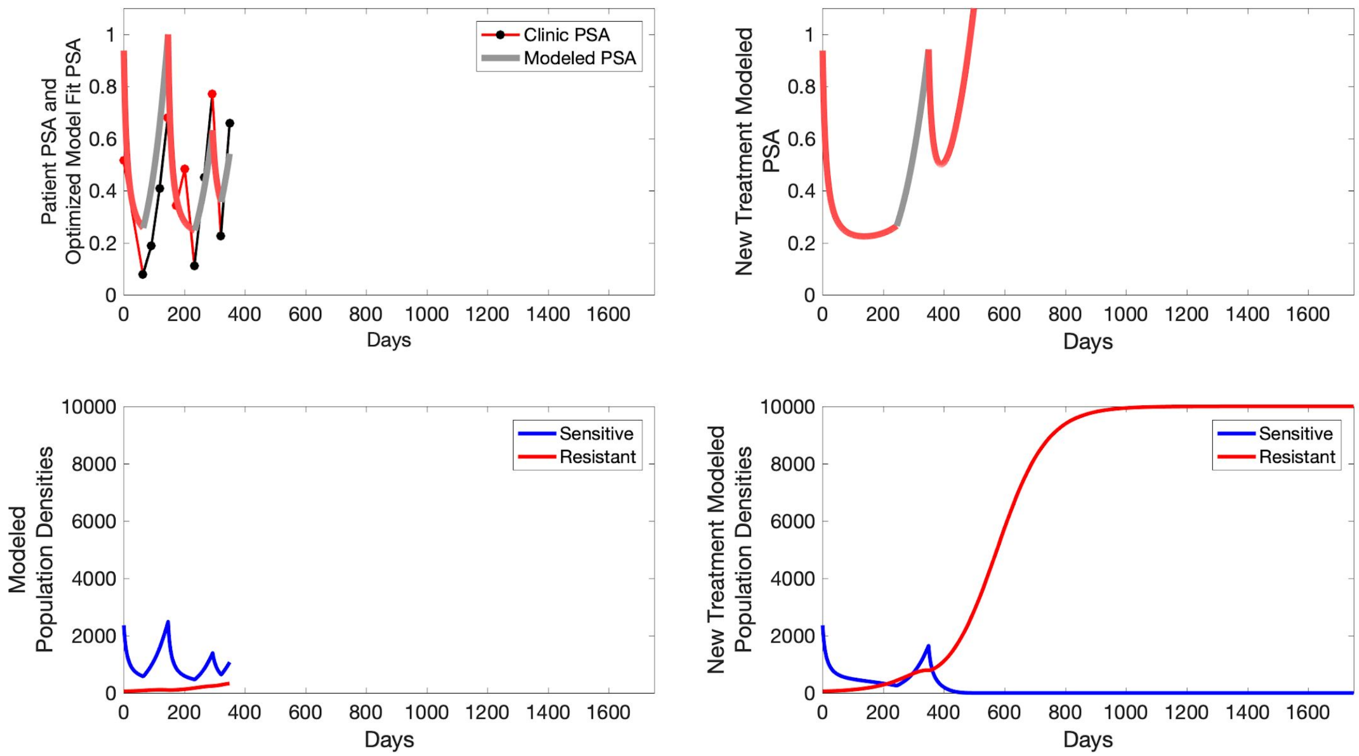
Patient PSA and
Optimized Model Fit PSA
Clinic PSA
Modeled PSA
Days
New Treatment Modeled
PSA
Days
Modeled
Population Densities
Sensitive
Resistant
Days
New Treatment Modeled
Population Densities
Sensitive
Resistant
Days

**P1003**

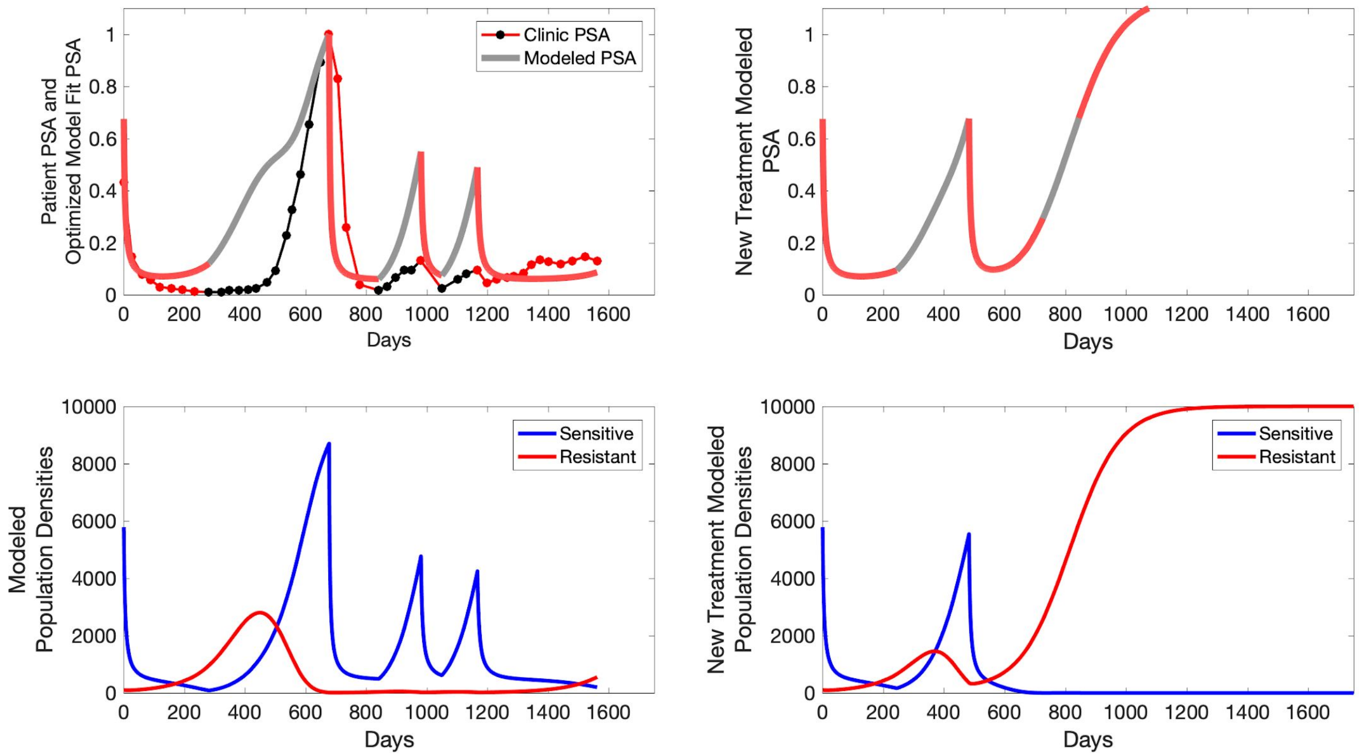

Clinic PSA
Modeled PSA
Patient PSA and Optimized Model Fit PSA
Days
New Treatment Modeled PSA
Days
Sensitive
Resistant
Modeled Population Densities
Days
Sensitive
Resistant
New Treatment Modeled Population Densities
Days

P1004

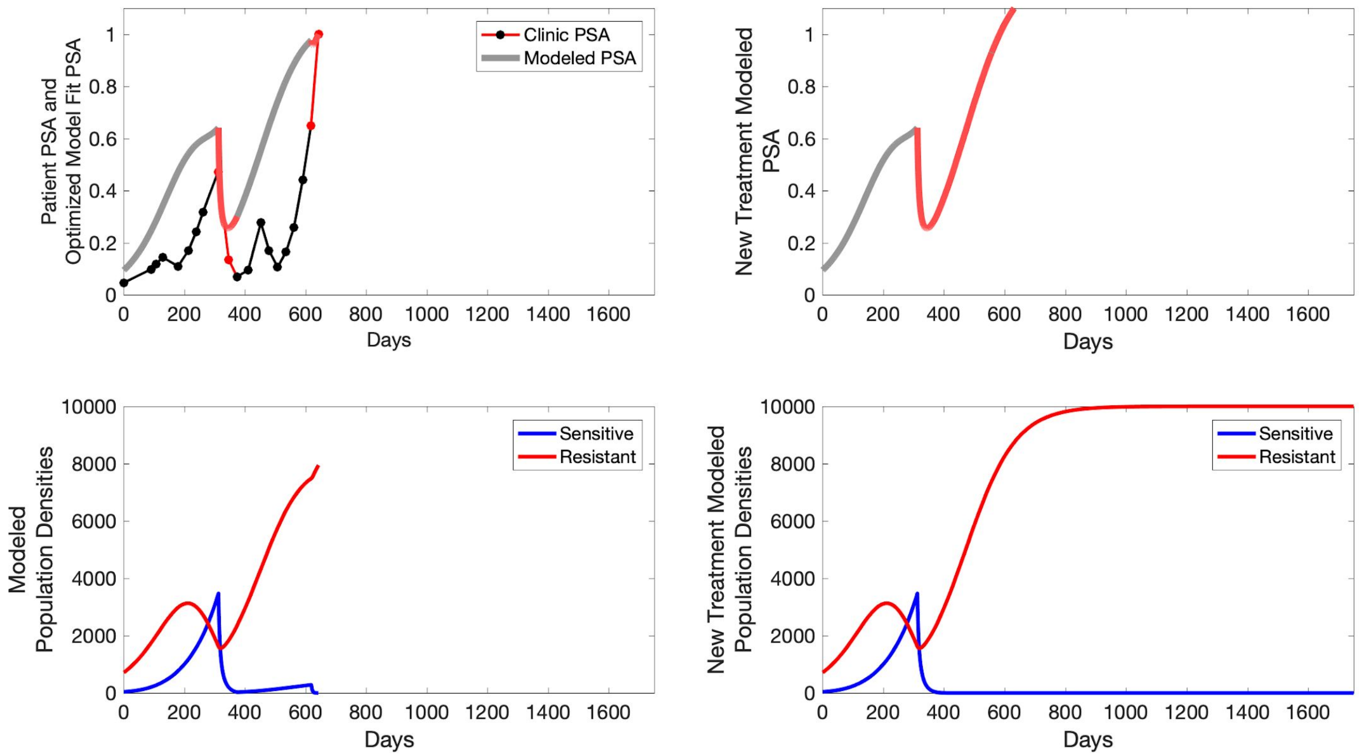
Patient PSA and Optimized Model Fit PSA
Clinic PSA
Modeled PSA
Days
New Treatment Modeled PSA
Days
Modeled Population Densities
Sensitive
Resistant
Days
New Treatment Modeled Population Densities
Sensitive
Resistant
Days

**P1005**

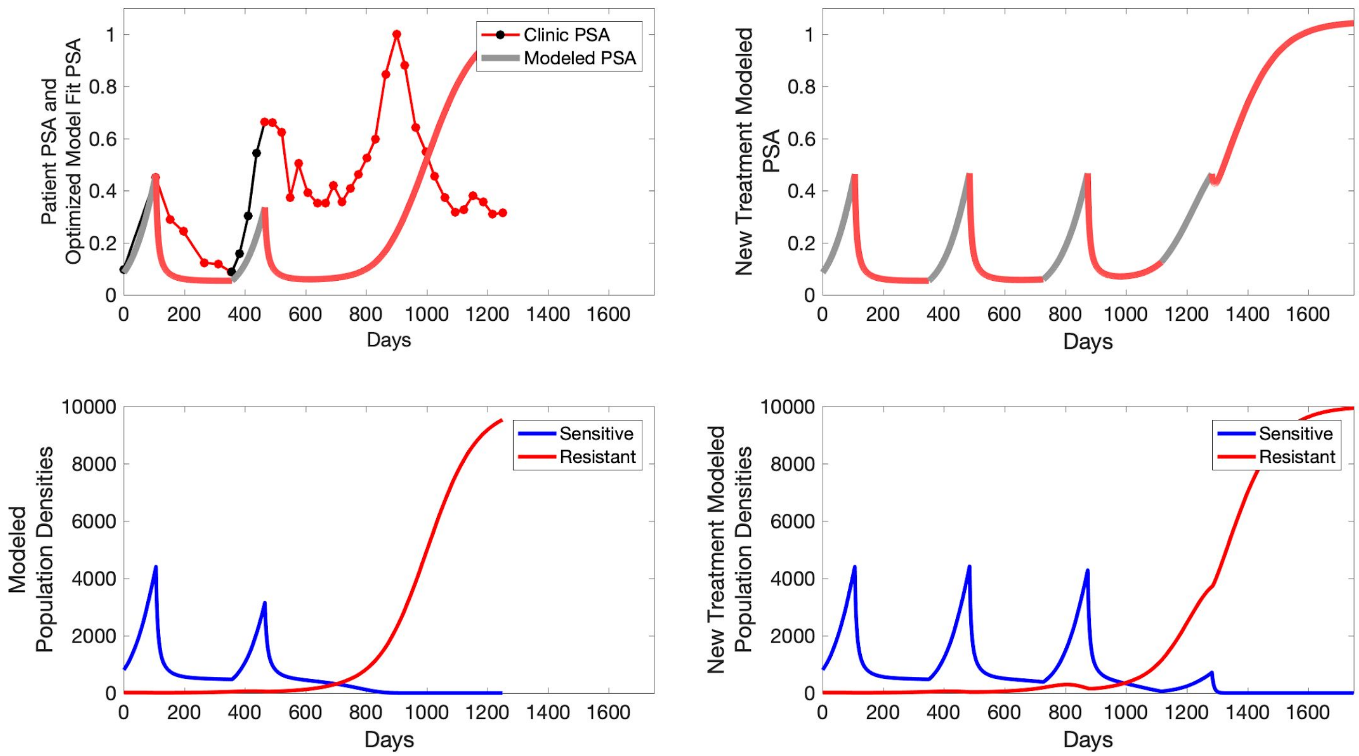

Patient PSA and Optimized Model Fit PSA
Clinic PSA
Modeled PSA
Days
New Treatment Modeled PSA
Modeled Population Densities
Sensitive
Resistant
New Treatment Modeled Population Densities

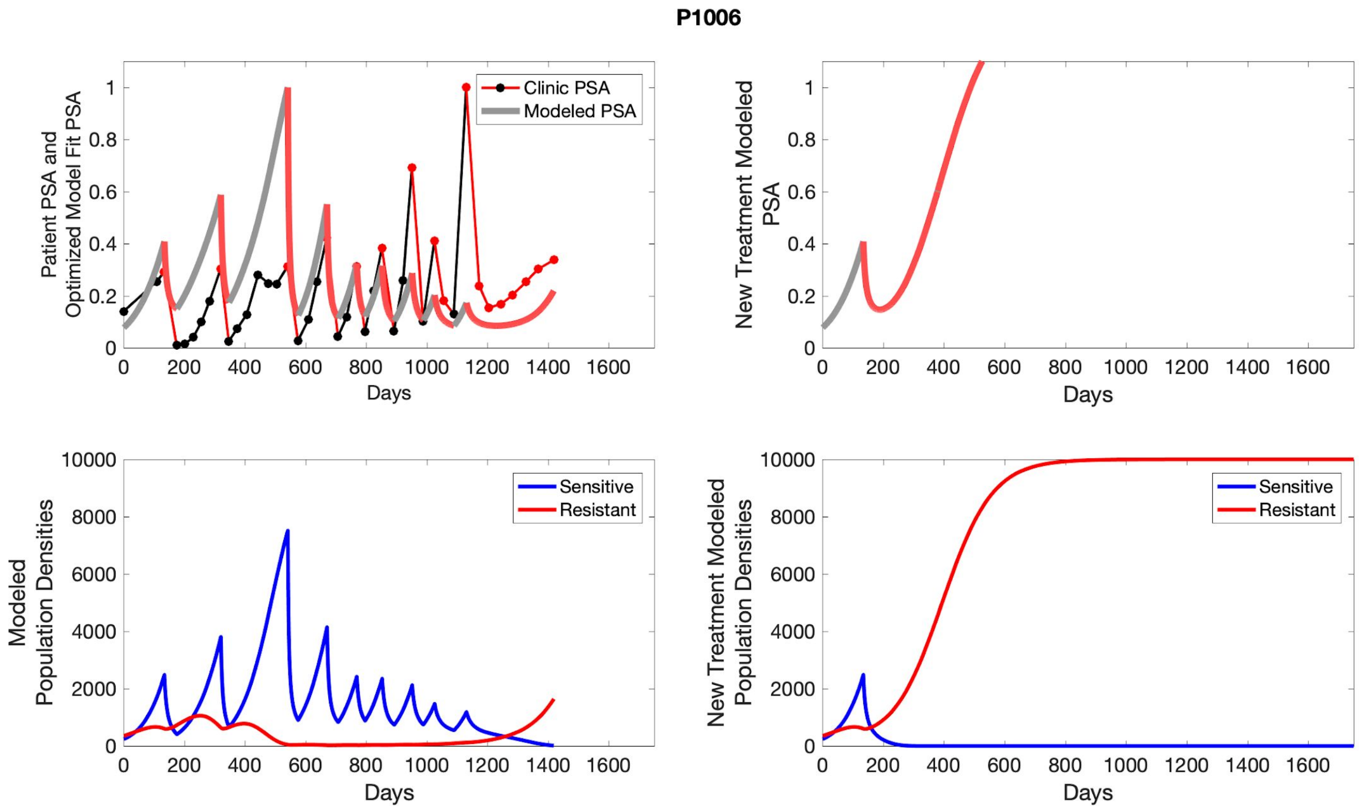

P1006
Patient PSA and Optimized Model Fit PSA
Clinic PSA
Modeled PSA
Days
New Treatment Modeled PSA
Days
Modeled Population Densities
Sensitive
Resistant
Days
New Treatment Modeled Population Densities
Sensitive
Resistant
Days

**P1007**

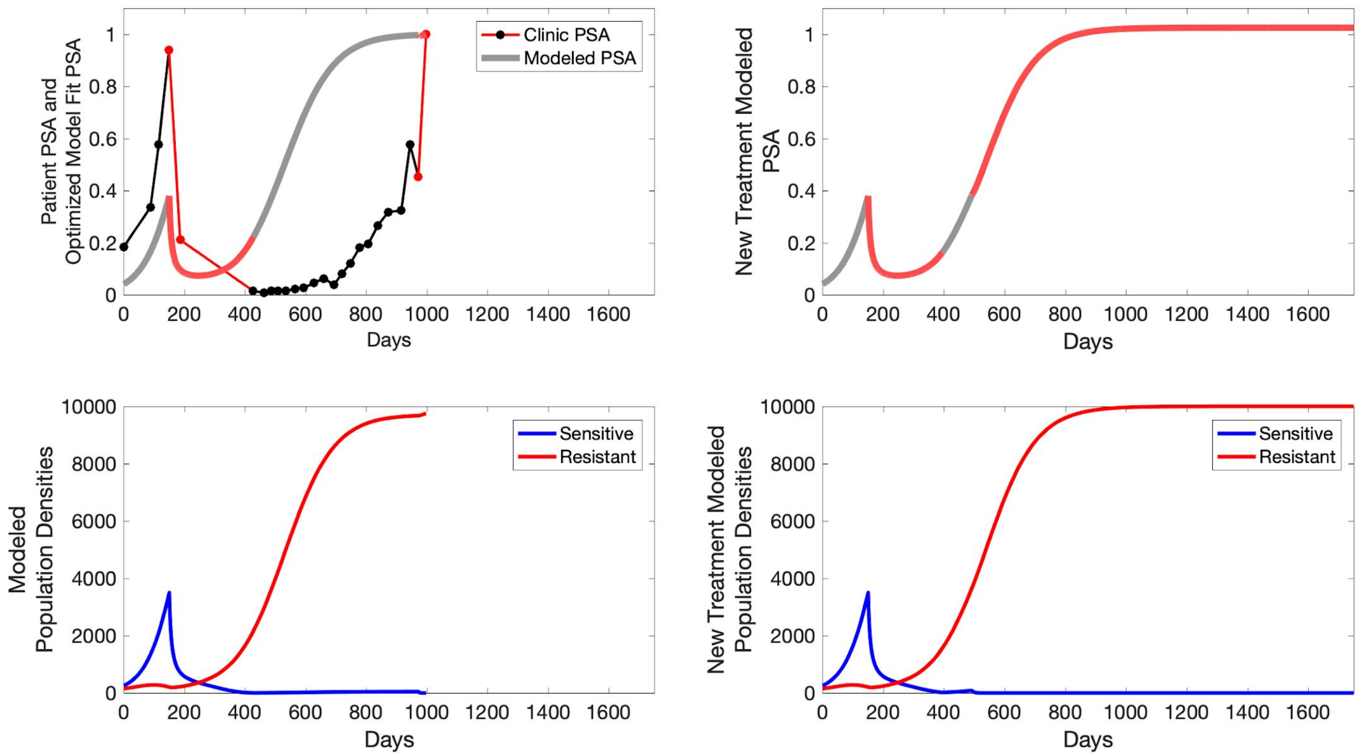

Clinic PSA
Modeled PSA
Patient PSA and Optimized Model Fit PSA
Days
New Treatment Modeled PSA
Days
Sensitive
Resistant
Modeled Population Densities
Days
Sensitive
Resistant
New Treatment Modeled Population Densities
Days

**P1009**

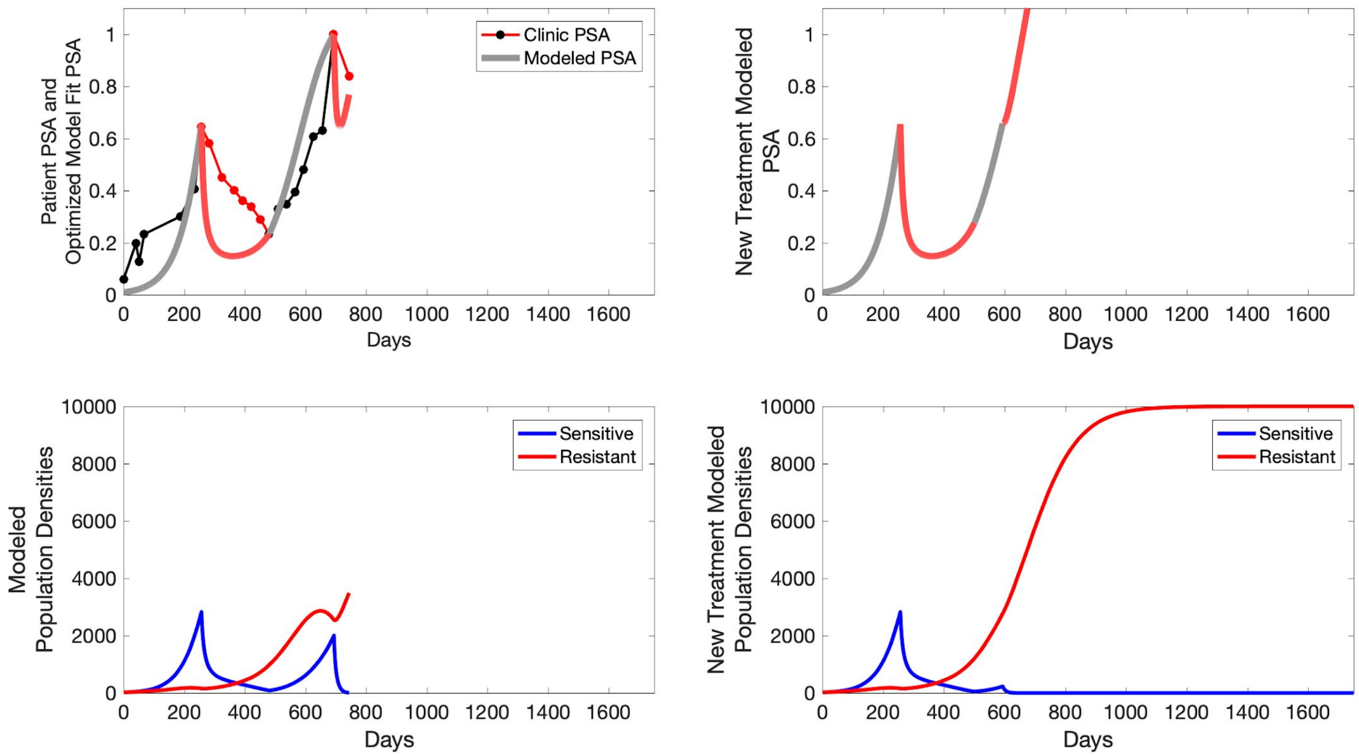

Patient PSA and Optimized Model Fit PSA
Clinic PSA
Modeled PSA
Days
New Treatment Modeled PSA
Days
Modeled Population Densities
Sensitive
Resistant
Days
New Treatment Modeled Population Densities
Sensitive
Resistant
Days

**P1010**

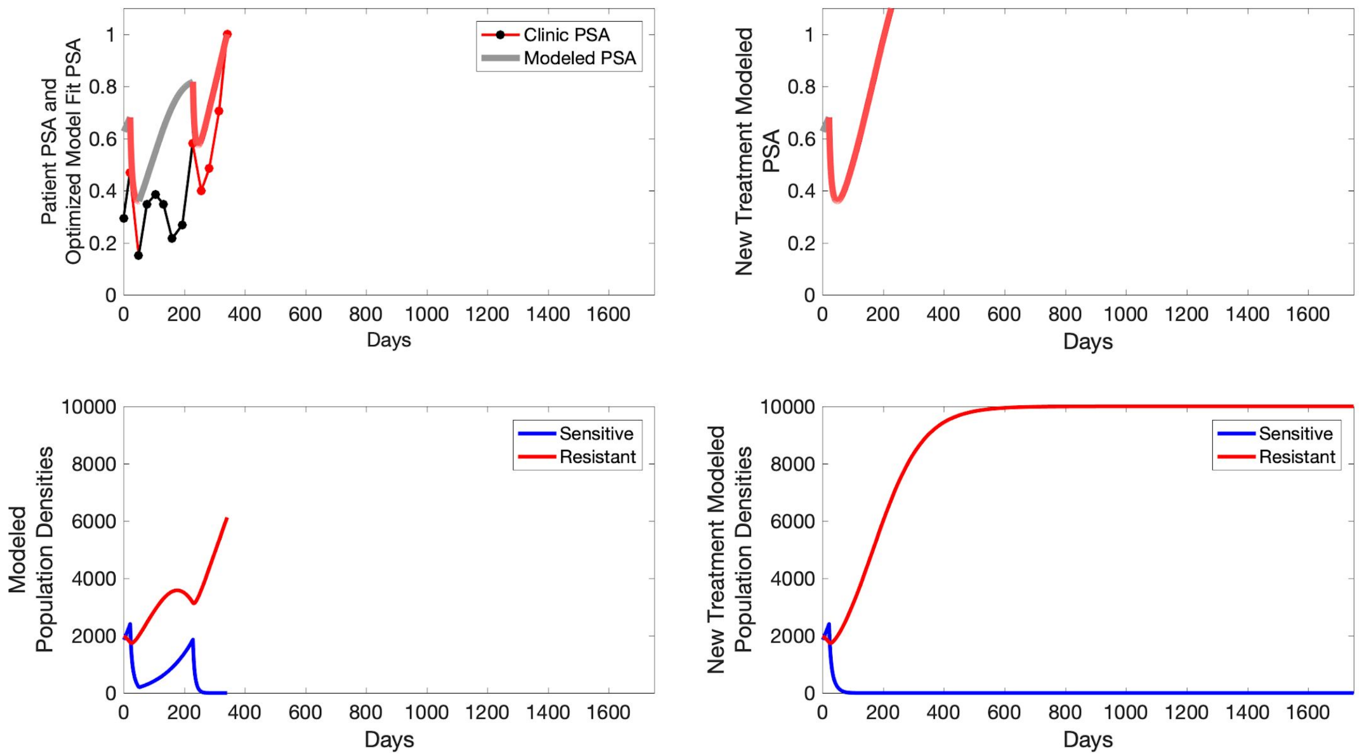
Patient PSA and Optimized Model Fit PSA
Clinic PSA
Modeled PSA
Days
New Treatment Modeled PSA
Modeled Population Densities
Sensitive
Resistant
New Treatment Modeled Population Densities

**P1011**

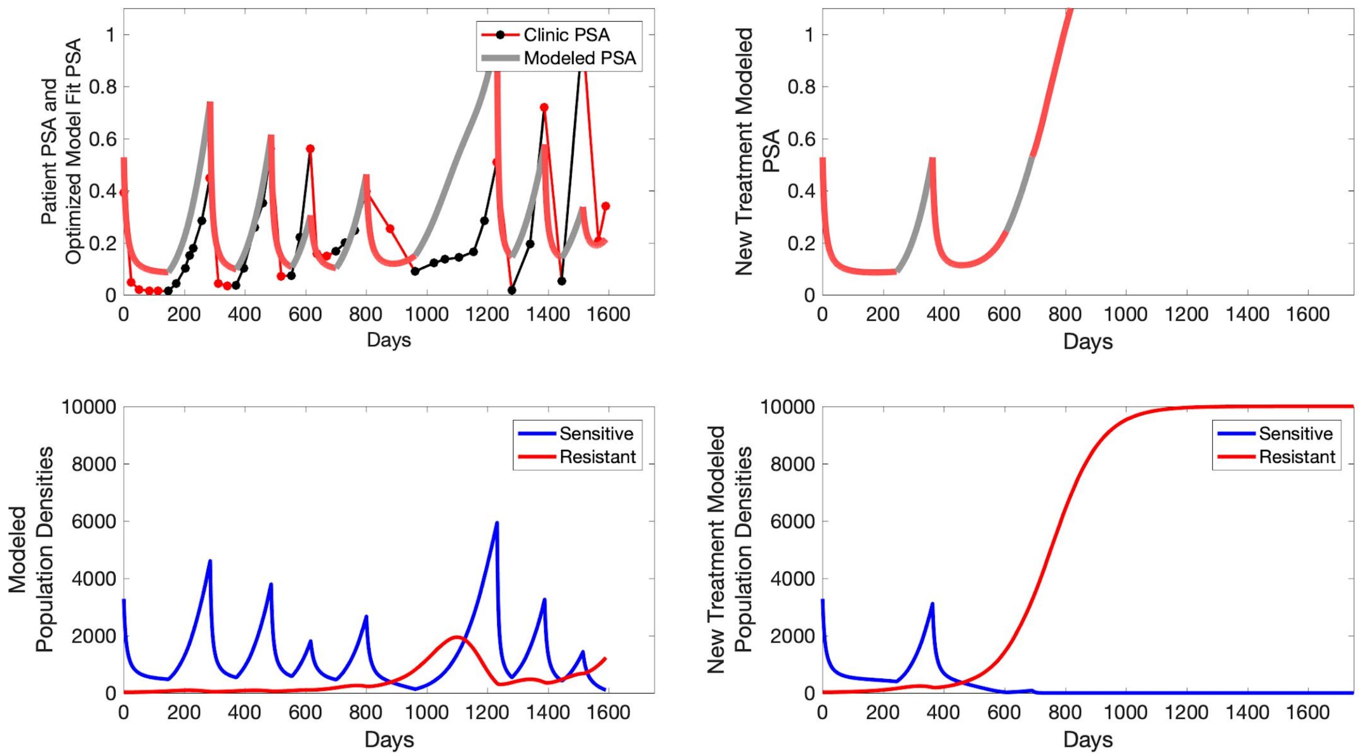

Patient PSA and Optimized Model Fit PSA
Clinic PSA
Modeled PSA
Days
New Treatment Modeled PSA
Days
Modeled Population Densities
Sensitive
Resistant
Days
New Treatment Modeled Population Densities
Sensitive
Resistant
Days

**P1012**

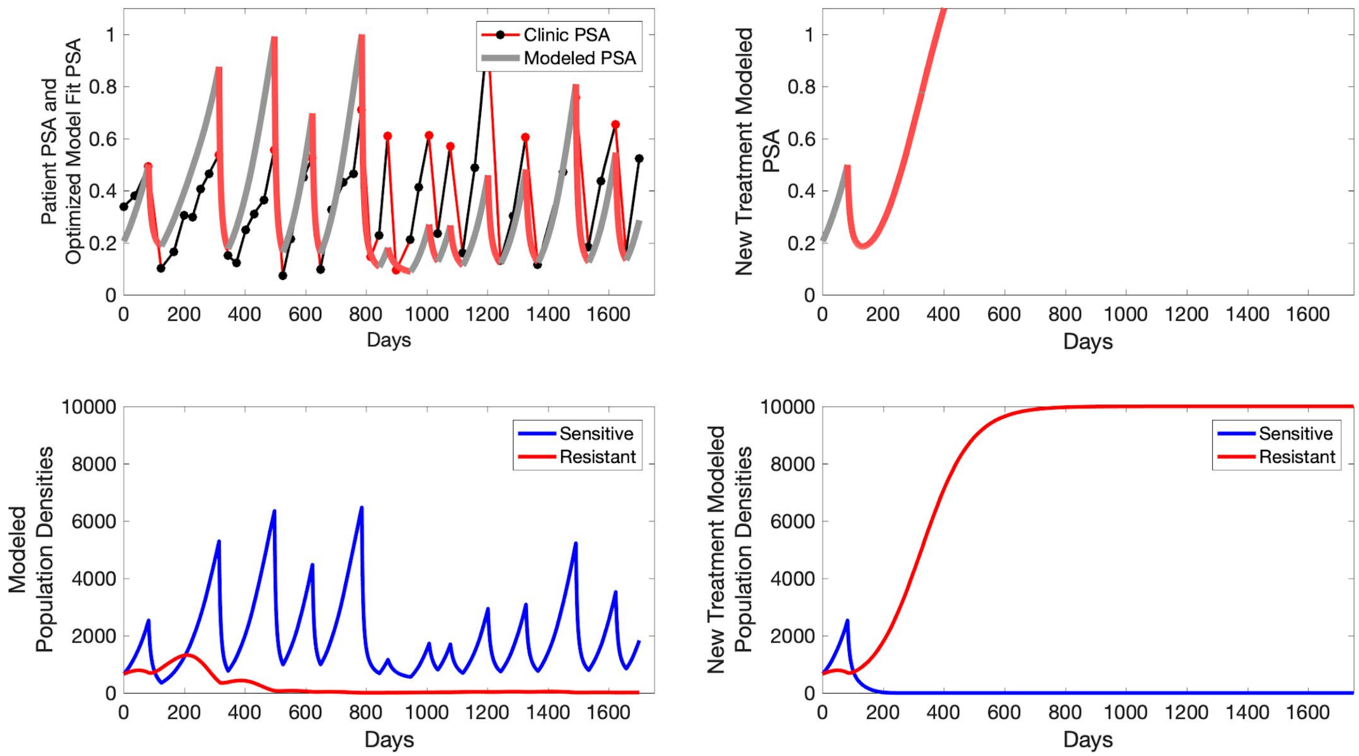

Clinic PSA
Modeled PSA
Patient PSA and
Optimized Model Fit PSA
Days
New Treatment Modeled
PSA
Days
Sensitive
Resistant
Modeled
Population Densities
Days
Sensitive
Resistant
New Treatment Modeled
Population Densities
Days

**P1014**

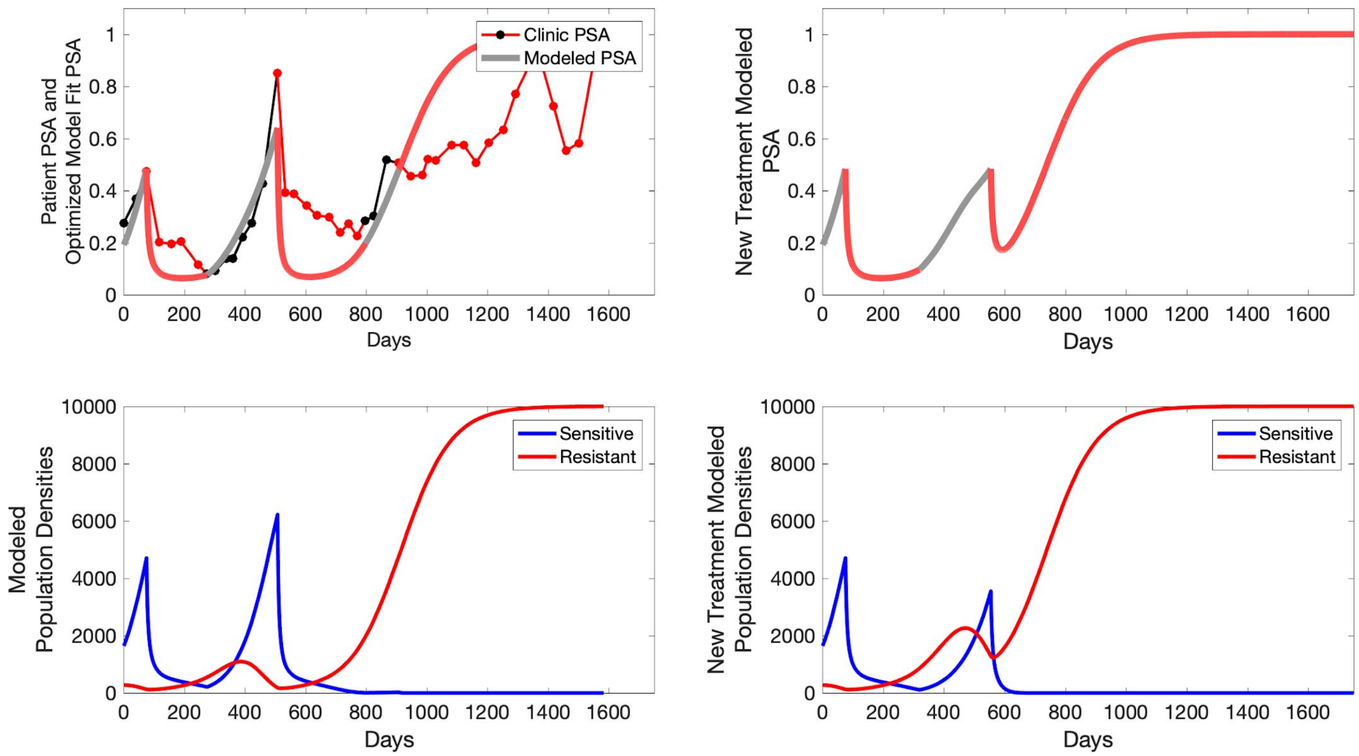
Clinic PSA
Modeled PSA
Patient PSA and Optimized Model Fit PSA
New Treatment Modeled PSA
Days
Sensitive
Resistant
Modeled Population Densities
New Treatment Modeled Population Densities
Days

**P1015**

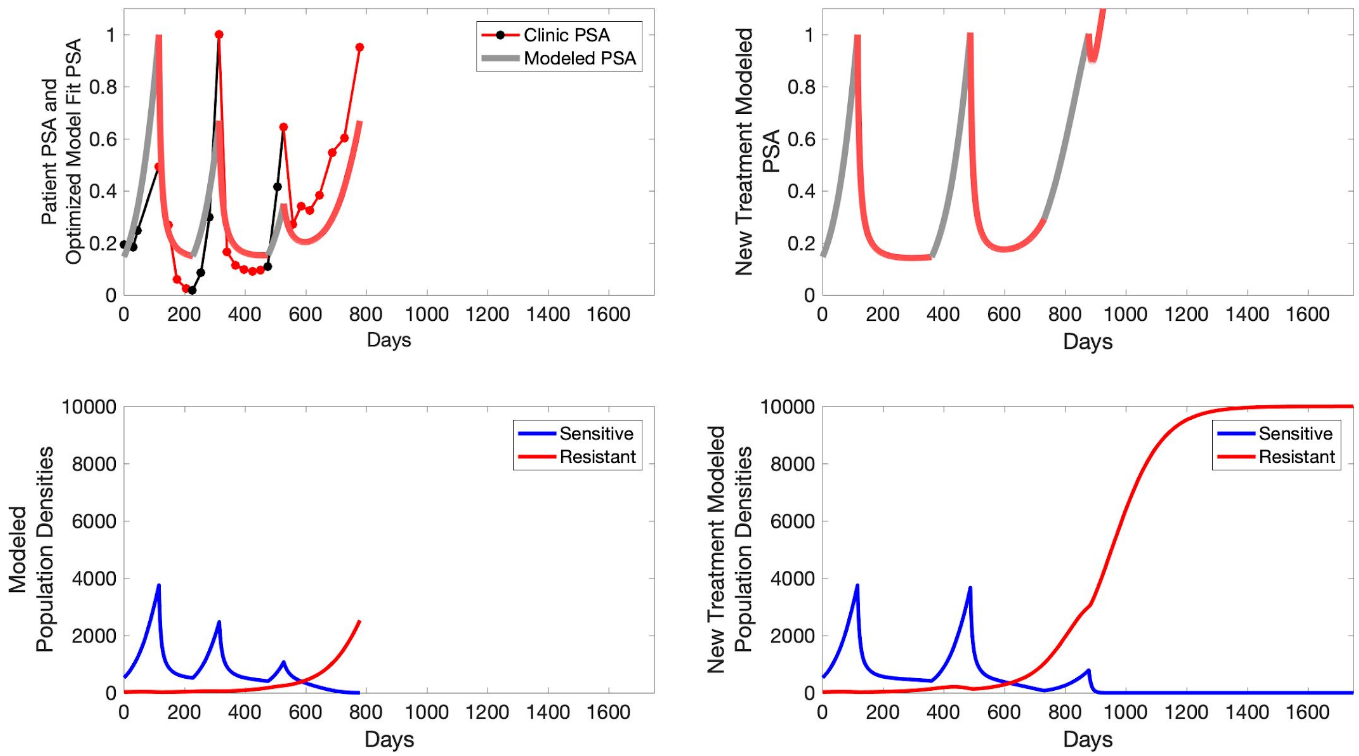

Patient PSA and Optimized Model Fit PSA
Clinic PSA
Modeled PSA
Days
New Treatment Modeled PSA
Days
Modeled Population Densities
Sensitive
Resistant
Days
New Treatment Modeled Population Densities
Sensitive
Resistant
Days

**P1016**

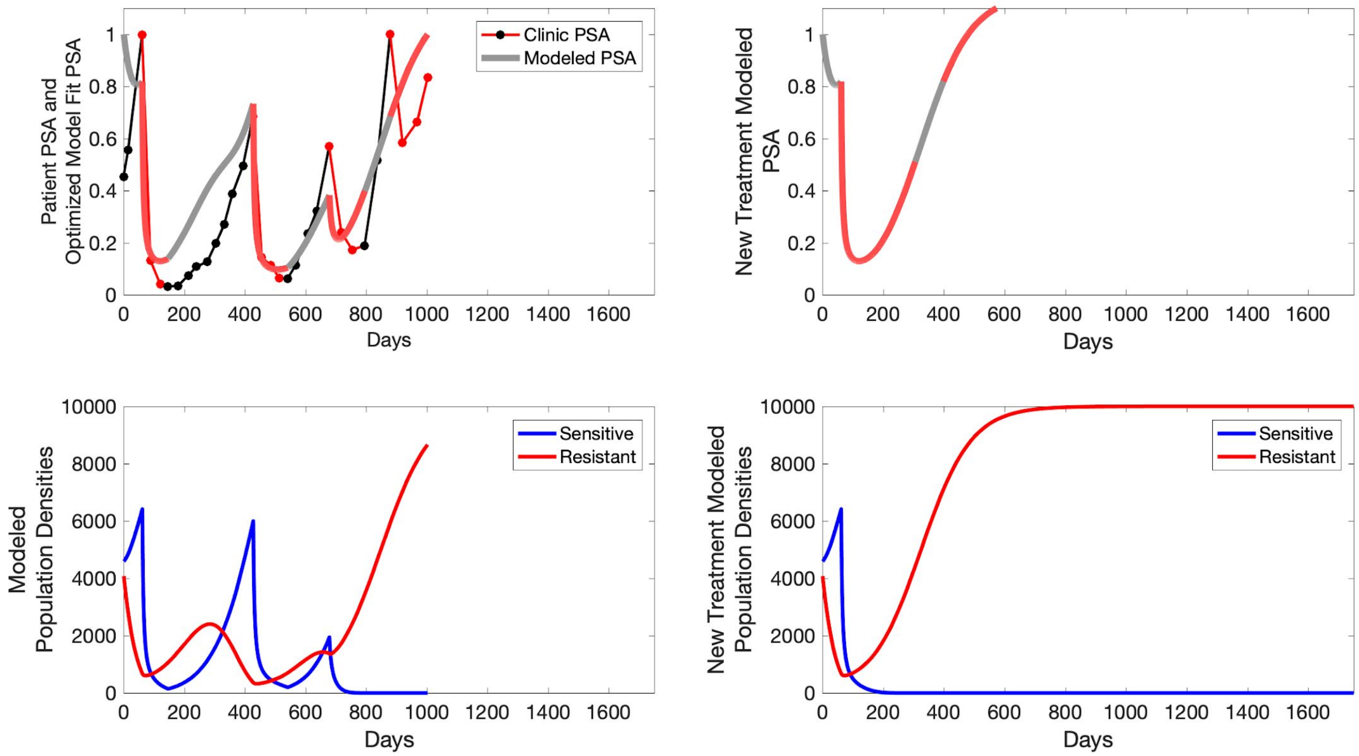

Patient PSA and Optimized Model Fit PSA
Clinic PSA
Modeled PSA
Days
New Treatment Modeled PSA
Days
Modeled Population Densities
Sensitive
Resistant
Days
New Treatment Modeled Population Densities
Sensitive
Resistant
Days

**P1017**

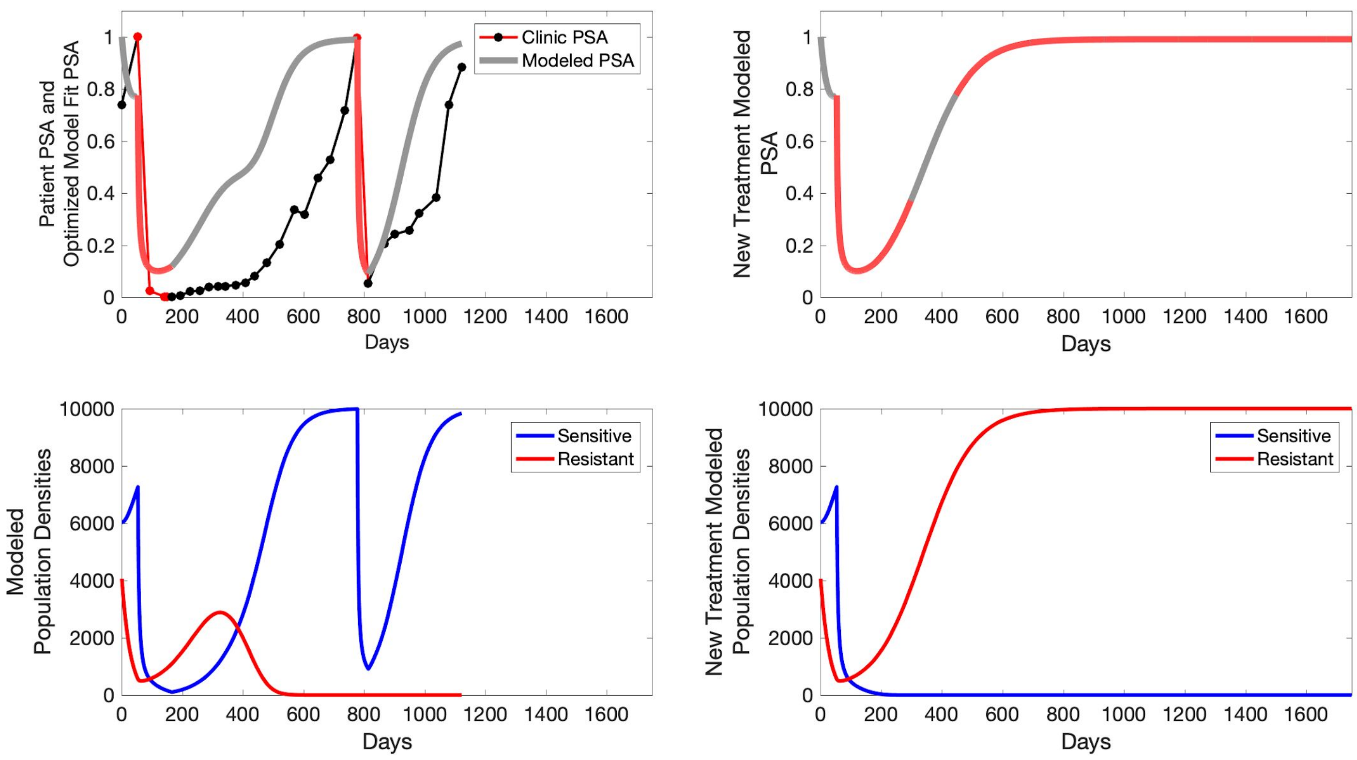

Patient PSA and
Optimized Model Fit PSA
Clinic PSA
Modeled PSA
Days
New Treatment Modeled
PSA
Days
Modeled
Population Densities
Sensitive
Resistant
Days
New Treatment Modeled
Population Densities
Sensitive
Resistant
Days

**P1018**

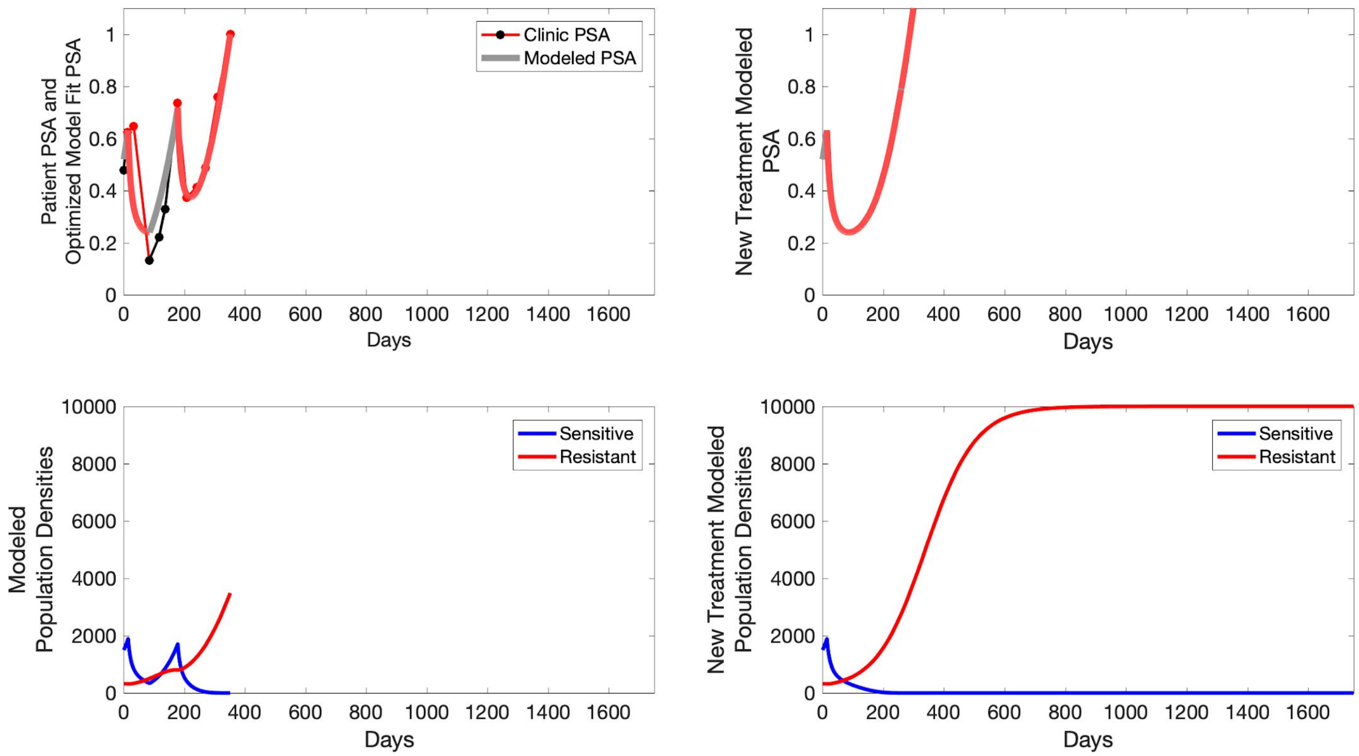

Patient PSA and Optimized Model Fit PSA
Clinic PSA
Modeled PSA
Days
New Treatment Modeled PSA
Days
Modeled Population Densities
Sensitive
Resistant
Days
New Treatment Modeled Population Densities
Sensitive
Resistant
Days

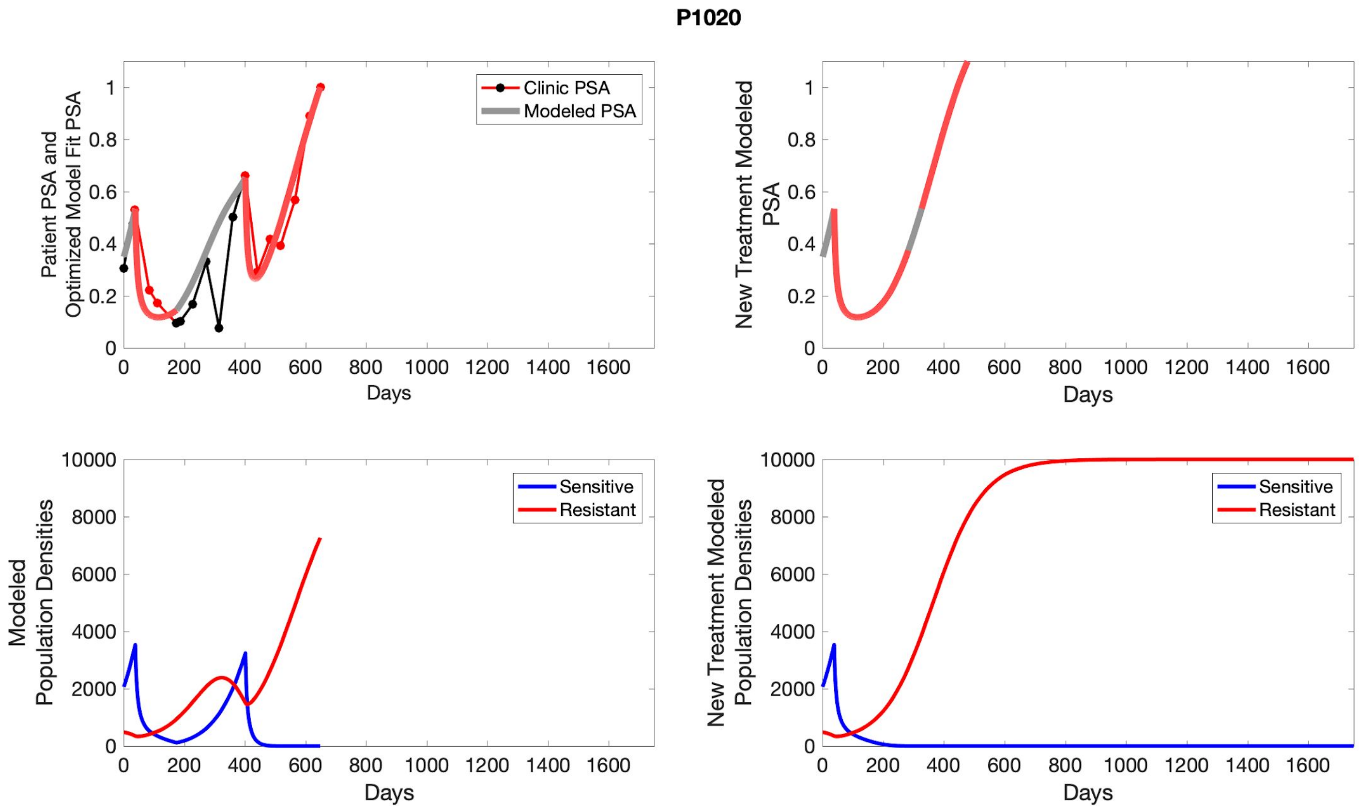
P1020
Patient PSA and Optimized Model Fit PSA
Clinic PSA
Modeled PSA
Days
New Treatment Modeled PSA
Modeled Population Densities
Sensitive
Resistant
New Treatment Modeled Population Densities

### S8 Comparison of TTP to estimated fraction of resistant cells in both cohorts

To evaluate model estimation of the fraction of cell that were resistant to treatment, we plotted the calculated fraction against observed survival.

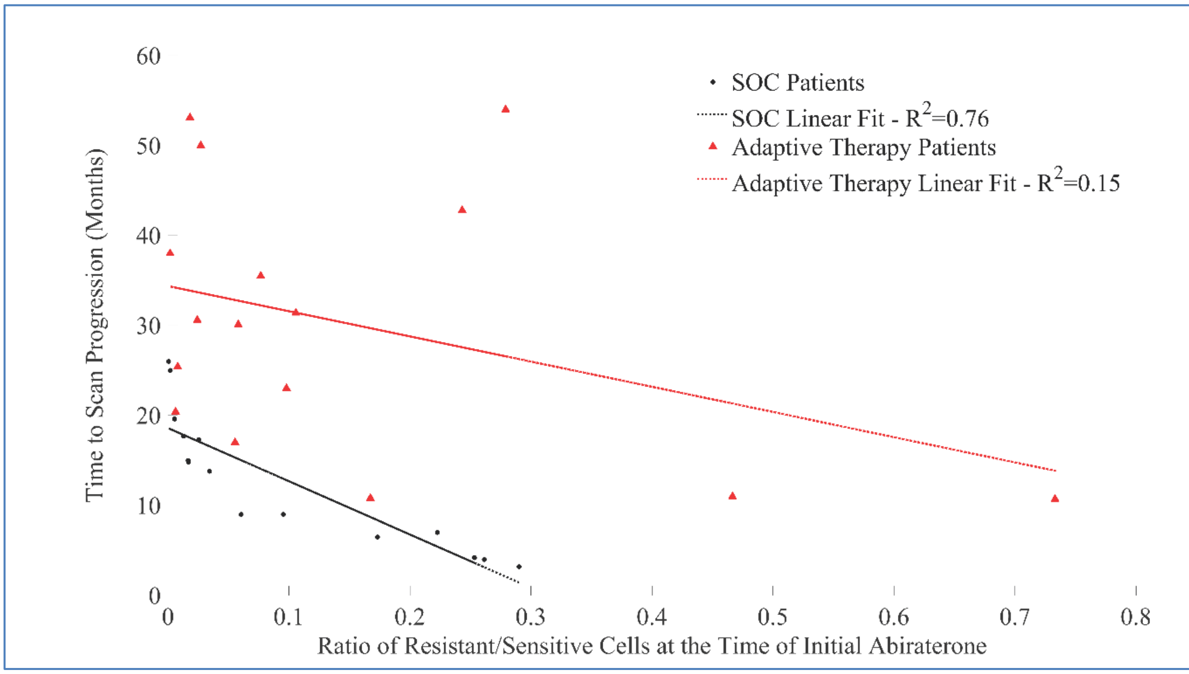


Figure S5: Comparing calculated fraction of resistant populations to TTP.

Analysis of PSA dynamics in both cohorts was used to estimate the pre-treatment fraction of resistant cells in each subject. We then compared that estimate to radiographic TTP in each patient. The correlations support the accuracy of the pre-treatment fraction estimates. The greater scatter in the adaptive therapy cohort probably reflects the more complex dynamics of treatment as described in the text. Note that, for every fraction of pre-treatment resistant cells, the adaptive therapy arm produced more favorable outcomes. (Only 15 standard of care patients are shown, as historical PSA measurements, and therefore mathematical analysis, is unavailable for one standard of care patient.)
